# Cost-optimal transmission reduction in early pandemic response: a multi-objective health-economic modelling study across 110 countries

**DOI:** 10.64898/2026.10.02.26363193

**Authors:** Jongmin Lee, Gerardo Chowell, Renier Mendoza, Victoria May P. Mendoza, Robin N Thompson, Eunok Jung

## Abstract

**Background:** Early pandemic responses must balance transmission control against socioeconomic costs but the strategy that minimises total costs and whether it varies by country remain unknown. We analysed country-specific cost-optimal intervention strategies and examined whether national indices could inform response targets.

**Methods:** We fitted mechanistic models to the first year of reported COVID-19 cases in 110 countries and assessed structural identifiability and parameter uncertainty. Multi-objective optimisation identified efficient trade-offs between transmission reduction and infections across 26-, 40-, and 52-week horizons without vaccination. We selected cost-minimising strategies using GDP-scaled intervention costs and infection costs incorporating quarantine and mortality. Excess-cost ratios, defined as the relative cost increase over the optimum, quantified deviations from the cost-minimising strategy. We also examined associations with national socioeconomic and health-system indices.

**Results:** Cost-optimal average transmission reduction ranged from 31.2% to 57.3% across countries. Across the time horizons, the mean excess-cost ratio for 110 countries was 55.2% (median 30.8%). Excess costs increased asymmetrically around the country-specific optimal transmission reduction: a five-percentage-point transmission reduction below the optimum was associated with excess-cost ratios exceeding 400%, compared with 10-80% for an equivalent increase above the optimum. The range of transmission reduction associated with excess-cost ratios below 40% widened 1.47-fold as the horizon increased from 26 to 52 weeks. Among national indices, the Socio-Demographic Index (SDI) and GDP per capita had the largest redundancy indices (0.40 and 0.37, respectively).

**Conclusions:** Under the modelled assumptions, transmission reduction below the country-specific optimum resulted in greater excess costs than reduction above the optimum, highlighting the potential value of precautionary early-response strategies when intervention effectiveness is uncertain. National socioeconomic and health-system indices may provide a broad indication of cost-optimal intervention intensity, although country-specific conditions remain important. These findings provide a quantitative framework for balancing intervention intensity, infection burden, and economic costs across diverse national settings.

**What is already known on this topic:**

❿ During the COVID-19 pandemic, both infections and control measures caused substantial economic losses, creating a trade-off between reducing infection-related costs and limiting intervention costs.
❿ Pandemic preparedness and health-system infrastructure have been assessed using national indices such as the GHS and UHC indices.

**What this study adds:**

❿ Across 110 countries, this study compares model-inferred responses with modelled cost-optimal strategies, quantifying excess costs under explicit epidemiological and economic assumptions.
❿ In four illustrative countries, modelled costs increased more after a five-percentage-point shortfall below the optimum than after an equivalent increase above it.
❿ National indices showed descriptive associations with modelled outcomes but did not establish validated country-specific transmission reduction targets.

**How this study might affect research, practice or policy:**

❿ Policymakers could evaluate precautionary early-response margins when intervention effectiveness is uncertain, while recognising that the appropriate margin depends on local conditions and cost assumptions.
❿ Preparedness plans could combine national indices with locally calibrated models and explicit assessment of feasibility, affordability and the distribution of costs.

## 1 Introduction

During large-scale disease outbreaks, policymakers confront a serious trade-off: they must implement measures to suppress transmission while simultaneously minimising socioeconomic damage.^1^ The coronavirus disease 2019 (COVID-19) pandemic sharply illuminated this challenge; facilitated by global air travel, a localised outbreak rapidly escalated into a simultaneous crisis affecting over 100 nations.^2-3^ The speed and scale of this spread often outpaced the capacity for rigorous scientific assessment, contributing to similar nonpharmaceutical intervention policies across heterogeneous countries.^4^ Although such interventions may reduce transmission, evidence-based analysis is essential to determine cost-optimal strategies that minimise overall societal damage and to build the public trust required to sustain these measures.^1,5-7^ However, critical gaps remain in understanding how national characteristics, such as population size, gross domestic product (GDP), Universal Health

Coverage (UHC), the Socio-demographic Index (SDI), and the Global Health Security (GHS) Index, can inform these policy decisions.^8-10,20-23^ Furthermore, it remains unclear which of these structural indices is most strongly associated with cost-optimal intervention intensity, limiting the ability to guide precise and context-specific policy. This question is particularly relevant to low- and middle-income countries (LMICs), where the feasibility of transmission control and the capacity to absorb its costs must be considered alongside health outcomes.

Previous studies have investigated these indices in relation to health financing, demographic stratification, and retrospective pandemic assessment. Research on health systems has focused on the financial aspects of care, analysing how national tax structures and funding mechanisms directly influence the scope of UHC.^5,11^ Global Burden of Disease studies have primarily used the SDI to stratify countries by development level, forecasting long-term shifts in population structure and projecting future trends for specific burdens such as stillbirths and falls.^12-15^ One study concludes that the GHS Index was not meaningfully associated with standardised COVID-19 infection or fatality rates, whereas structural factors related to the SDI were found to be significant correlates of cross-country variation in outcomes.^5^ In the domain of pandemic modelling, studies have integrated global air travel data to correct observational biases in early surveillance and quantify the risk of cross-border viral importation.^3,16^ Economic evaluations have relied on GDP metrics to assess the cost-effectiveness of response strategies, comparing the macroeconomic impact of interventions against health outcomes in both vaccination programs and containment measures.^17,18^ However, the extent to which these indices can inform country-specific cost-optimal transmission reduction remains uncertain.

Building on our previous optimisation framework, we analysed 110 countries with populations exceeding five million to compare model-inferred responses with country-specific cost-optimal strategies during the early COVID-19 pandemic. We integrated dynamic infectious-disease modelling, economic valuation and multi-objective optimisation to address three questions: how much modelled costs differed from the optimum; whether comparable departures below and above the optimum had similar cost consequences; and how national indices related to these results. The aim was to provide exploratory benchmarks for locally calibrated preparedness planning, not to infer causal policy effects or prescribe interventions from national indices alone.

## 2 Methods

### 2.1 Data sources

Confirmed COVID-19 case and death counts were obtained from Our World in Data as country-level daily time series.^19^ GDP per capita (2019, USD) and total population (2019) were extracted from the World Bank.^20^ Universal Health Coverage indices were obtained from Our World in Data and WHO.^8,21^ Socio-demographic Index (SDI; 2019) values were obtained from the Global Health Data Exchange, and Global Health Security index (GHS; 2019) scores from their database.^9,10,22,23^ We used ISO 3166-1 alpha-2 country codes in the figures.^24^ All variables were collated for 110 countries (population >5 million; Supplementary file 1, section A).

### 2.2 Categorisation criteria

We categorised the 110 countries by region, population size and quartiles of the national indices. Region was divided into seven GBD super-regions. Population was grouped into four bands (>100M, 30M-100M, 10M-30M, 5M-10M). The other indices were each divided into four quartile groups (Supplementary file 1, section A).

### 2.3 Modelling, parametrisation and assumptions

We used a deterministic SEIQRD compartmental model partitioning each country’s population into susceptible (S), exposed/latent (E), infectious (I), quarantined/isolated (Q), recovered (R), and deceased (D). The governing equations are

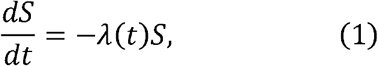

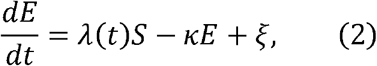

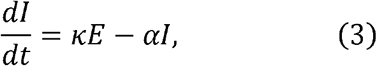

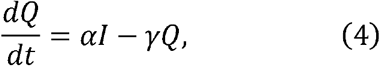

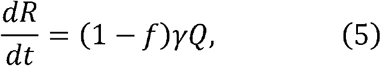

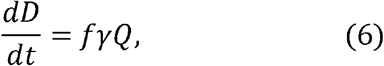

where *λ* (*t*) = (1 - *µ*(*t*))*R*_0_*αI*/*N* is the force of infection with *N* = *S* + *E*+ *I*+ *R*, defined as the non-isolated living population; *Q* is excluded because quarantined or isolated individuals are assumed not to participate in transmission, and *D* is excluded because deceased individuals are no longer part of the mixing population. *R*_0_ is the basic reproduction number, 1/*κ* the latent period, 1/*α* the infectious period, 1/*γ* the isolation-to-outcome period, *f* the case fatality ratio, *ξ* the daily imported latent individuals, and *µ*(*t*) the time-varying transmission reduction, interpreted as a proxy for the combined effects of non-pharmaceutical interventions. The model was adapted from our previously published framework^25^ and implemented in MATLAB. The fitted reduction is inferred from case data rather than directly measured policy stringency, and therefore does not identify the causal effect of individual interventions.

The model simulates infections (transitions from *S* to *E*), while parameter estimation is calibrated to cumulative confirmed cases (transitions from *I* to *Q*). The cost function (equation 8) is applied to simulated infections (*g*_2_). Because every infectious individual in the model is eventually isolated and reported, the model contains no separate undetected-infection compartment, so the simulated infections are on the same ascertainment scale as the confirmed cases used for calibration. The country-specific case fatality ratio is applied on this modelled ascertainment scale; neither simulated infections nor mortality-related costs should therefore be interpreted as estimates of the full population burden.

Disease parameters (*R*_0_=2.87, 1/*κ* =4 days, 1/*α* =5.2 days, 1/*γ* = 14 days) were fixed from literature (Supplementary file 1, section B). The case fatality ratio *f* was country-specific, calculated from reported deaths divided by confirmed cases with a one-year time horizon. Deaths were used only to calculate CFR and were not included in the parameter-fitting procedure, which used cumulative confirmed cases alone.

For each country, we estimated one importation parameter *ξ* (successfully imported latent cases), constrained to be non-negative so that it represents net importation of latent cases, and an transmission reduction rate over time *µ*(*t*), which is bounded between 0 (no reduction) and 1 (complete transmission block) and constructed using piecewise linear interpolation across the transmission reduction values *µ*_3_, *µ*_4_, …, *µ*_*K*_, where each *µ* represents transmission reduction during the i-th two-week interval. We define the primary case as the first imported latent individual that seeds the outbreak, and the index case as the first reported case, assumed two weeks after the primary case. Transmission reduction was fixed at zero over the first two two-week intervals (the first four weeks), representing the pre-response period before nonpharmaceutical interventions took effect; during these intervals, *µ*_1_= *µ*_2_= 0 was assumed; subsequent values were estimated, yielding 1+ (*K* − 2) estimated parameters per country, where *K* is the total number of two-week intervals in the simulation horizon (*K* = 13 for 26-week, *K* = 20 for 40-week, *K* = 26 for 52-week), giving 13, 20, and 26 estimated transmission reduction parameters, respectively. The fitted results represent the model-inferred transmission reduction and each country’s infected cases. The outbreak was seeded solely through importation.

Simulation lengths of 26, 40 and 52 weeks were considered as horizons without explicit vaccination (Supplementary file 1). Structural identifiability analysis indicated that the estimated parameters are globally identifiable under the assumed initial conditions and the additional assumption of a fixed reproduction number (Supplementary file 2). Structural identifiability under these assumptions does not establish practical identifiability from noisy surveillance data.

### 2.4 Multi-objective optimisation

For optimisation, the transmission reduction function is a decision variable, distinct from the function fitted to each country’s confirmed-case data. For each country, we optimised *µ*(*t*) to simultaneously minimise two objectives:

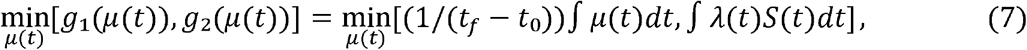

where *g*_1_ is the average transmission reduction (proxy for intervention intensity) and *g*_2_ is the total infections, subject to equations (1)-(6). A solution is Pareto optimal if no feasible alternative improves one objective without worsening the other. The set of all such solutions forms the Pareto front. Solutions were generated using NSGA-II via MATLAB’s *gamultiobj*, run 1,000 times independently; the final front was assembled from the combined non-dominated set (Supplementary file 1, section D).^26^

### 2.5 Economic evaluation

To select a single cost-optimal point from the Pareto front for each country, we converted objectives into monetary terms and minimised the sum of modelled intervention and infection costs:

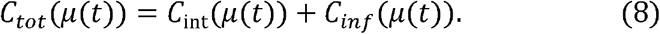

The intervention cost over the simulation horizon is *C*_int_ = *C*_1_ × *g*_1_, where *C*_1_ = (GDP_total_/ 365) × GDP_MaxRed_ is the maximum daily intervention cost. Here GDP_total_ is the country’s total GDP in USD (not per capita) and GDP_MaxRed_ = 0.10 (10% of daily GDP assumed lost under full intervention). Elsewhere, GDP_per_ indicates the GDP per capita.

The infection cost over the simulation horizon is *C*_*inf*_ = *C*_2_ × *g*_2_, where *C*_2_ = *C*_*H*_ + *f* × VSL is the cost per infected individual. *C*_*H*_ = GDP_per_ × 0.02 is the quarantine cost per patient, *f* is the country-specific fatality ratio, and VSL = GDP_per_ × 54.9 is the value of a statistical life.^27,28,32^ The multiplier 54.8966 is the ratio of the value of a statistical life to GDP per capita adopted from Sweis’s research on VSL estimates for the COVID-19 period.^32^ Multiplying it by each country’s GDP per capita gives an income-adjusted VSL, and the same multiplier is applied to all 110 countries, so that VSL differs across countries only through GDP per capita. The cost *C*_2_ is applied to every simulated infection (*g*_2_), rather than to the contemporaneous number of quarantined individuals. Mortality-related costs are monetary valuations of risk, not measured GDP losses; reporting total costs as a share of GDP provides a common scale for comparison.

The cost-optimal point minimises *C*_*tot*_ on the Pareto front, yielding a selected cost-optimal strategy *µ*^*^(*t*) per country (Figure 4, Supplementary file 1, section E). Throughout the paper, we distinguish three quantities: the model-inferred effective transmission reduction, the Pareto front (all efficient trade-offs), and the cost-optimal results (model-derived cost-minimising strategies).

### 2.6 Statistical analysis

Country-specific parameters were inferred using IMODE (run 25 times per country) followed by Bayesian inference via Delayed Rejection Adaptive Metropolis (DRAM) with IMODE-derived priors, burn-in, convergence assessment using 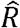 and ESS, and 300,000 simulations (Supplementary file 1, section C).^29,30^ Convergence diagnostics for the DRAM algorithm, including trace plots, posterior intervals, and posterior predictive checks, are provided in Supplementary file 2.

Pearson correlation coefficients were calculated across 110 countries between country-level indices (population, GDP per capita, UHC, SDI, GHS) and disease outcomes (confirmed cases, deaths, simulation results; Supplementary file 1, section F). Pearson correlation was chosen because the relationship between structural indices and epidemic outcomes is approximately linear on the scales used in this analysis. As a sensitivity check, we also computed Spearman rank correlations; the results were qualitatively similar (Supplementary file 1, section F). The correlation heatmap includes p-values; false-discovery-rate-adjusted q-values are reported in Supplementary file 1, section F. Four countries were selected for detailed illustration: Nepal, Republic of Korea, Ghana, and the United States of America.

## 3 Results

### 3.1 Model-inferred effective transmission reduction and epidemic outcomes

Across the 26-, 40-, and 52-week horizons, mean model-inferred average transmission reduction increased from 0.5111 to 0.5628, while the modelled proportion of infected individuals increased from 0.45% to 2.65% (Figure 1; Supplementary file 1, section G). The fitted points increasingly lay above the Pareto frontiers, indicating more infections than in efficient modelled strategies at comparable average transmission reduction. The standard deviation decreased for transmission reduction but increased for the proportion of infected individuals, showing that similar average reductions coexisted with increasingly heterogeneous modelled burdens (Supplementary file 1, section G). These comparisons describe fitted model outcomes, not independently measured policy effectiveness.

**Figure 1.**
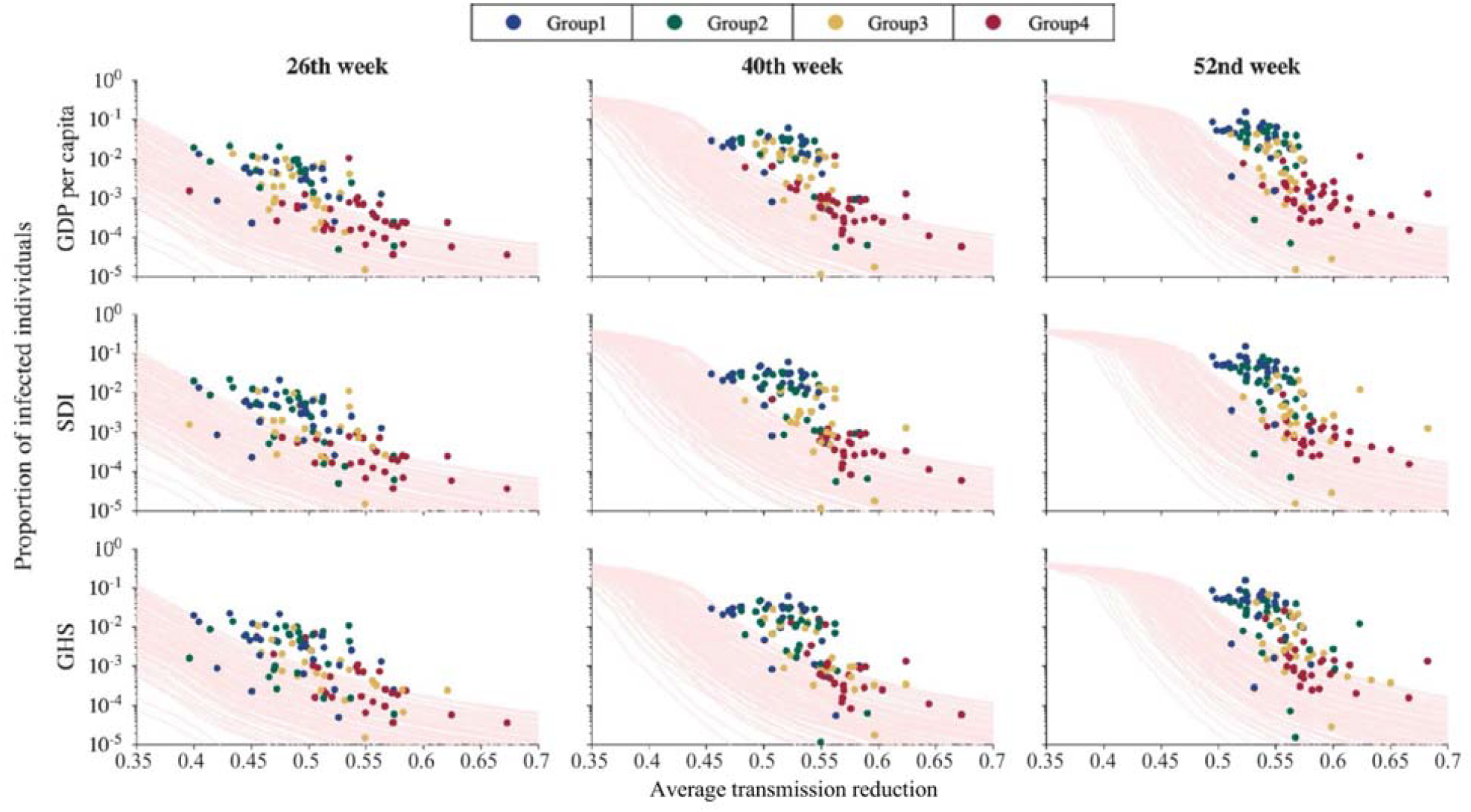
Pareto frontiers and data-fitted results categorised by indices for 110 countries under three simulation horizons. The figure is structured into three columns representing modelled simulation horizons (26, 40 and 52 weeks from the index case) and three rows corresponding to key country-level indices (GDP per capita, SDI, and GHS). Scattered circles represent the data-fitted results for each country, plotted against average transmission reduction (x-axis) and the proportion of infected individuals (y-axis). The pink curves represent the Pareto frontiers, indicating efficient trade-offs that simultaneously minimise intervention effort (average transmission reduction) and infection burden (total infections); monetary cost is applied subsequently to select a single cost-optimal point on each frontier. Coloured dots (blue, green, yellow, and red) indicate groups 1 to 4, respectively, with group definitions differing across the three categorising criteria as described in Methods 2.2 Categorisation criteria.

Modelled epidemic outcomes showed descriptive differences across country categories as the horizon extended. While the data points appear as a relatively homogeneous cluster at 26 weeks, they begin to form visible groups by 52 weeks across all rows (Figure 1). In particular, the SDI and income provided relatively better separation than other indices under the selected clusterevaluation metrics, although silhouette values near zero indicated weak cluster separation (Supplementary file 1, section H). Specifically, Group 1 (blue dots, representing high index values) tends to cluster toward lower transmission reduction and higher case counts, whereas Group 4 (red dots, representing lower SDI/Income) tends to cluster toward higher transmission reduction and lower case burdens (Figure 1). Nevertheless, differences among countries with similar inferred transmission reduction are not fully explained by these indices alone, and may also reflect differences in surveillance and other contextual factors.

### 3.2 Pareto frontiers and country-specific cost-optimal solutions

A wide range of proportions of infected individuals under similar model-inferred transmission reduction can be explored by visualising the trajectories of estimated points over time, which illustrate differences in the timing of transmission reduction and the associated modelled infection burdens. For instance, the United States and Nepal show different intensities of transmission reduction at the beginning, so that the relative position is above and below the cost-optimal solution, respectively (Figure 2). These data-fitted trajectories allow for a direct comparison of temporal strategies, showing that the Republic of Korea and Ghana followed trajectories closer to their cost-optimal paths under our model’s assumptions, with average transmission reduction exceeding 0.5, in contrast to the United States and Nepal (Figure 2, Supplementary file 1, section I, Supplementary file 3). These comparisons highlight the importance of examining both the timing and magnitude of transmission reduction within the model (Supplementary file 3).

**Figure 2.**
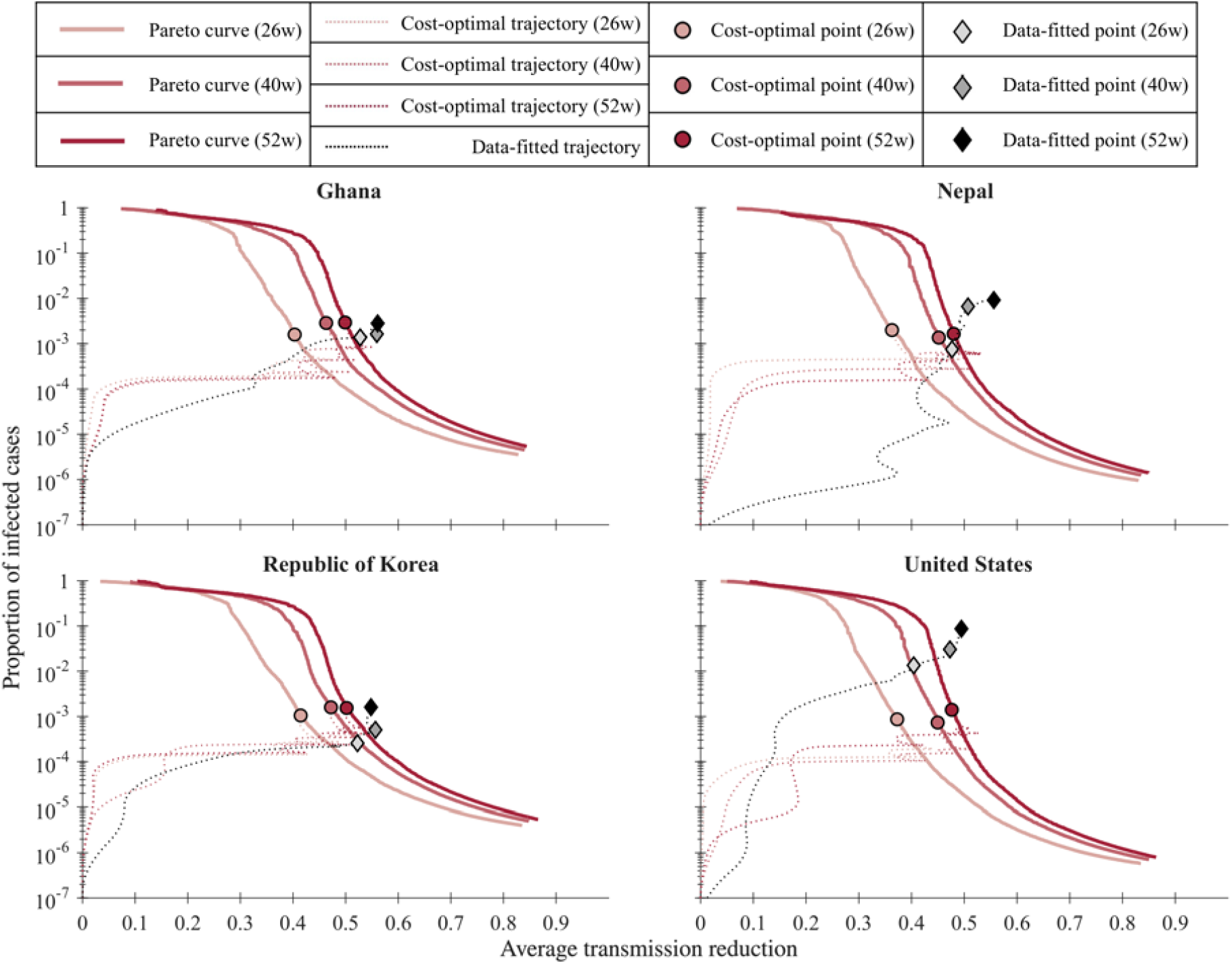
Pareto frontiers, cost-optimal solutions, and data-fitted results across three simulation horizons for four countries. The x-axis represents the average transmission reduction, and the y-axis (logarithmic scale) represents the proportion of infected individuals (the objective g2). The solid curves denote the Pareto-optimal frontiers, and circular dots indicate the cost-optimal points for each solid curve. Diamonds represent fitted results from each country’s confirmed-case trajectory. Dotted lines visualise the trajectories of cost-optimal and data-fitted results over time, showing how expected results for each strategy shift as the modelled horizon extends.

The gap in transmission reduction between cost-optimal solutions and data-fitted results narrowed on average across countries over time, although convergence in average reduction alone does not establish convergence in total costs, and substantial variation remained. For the 110 countries, the average transmission reduction for cost-optimal solutions shifted from 0.4097 to 0.5072, while data-fitted results moved from 0.5111 to 0.5628. However, the trends in these differences vary by country; 40 countries show a decreasing trend, 37 show an increasing trend, and 33 show a non-systematic pattern (Supplementary file 1, section I). No consistent relationship was observed between these trends and country groupings based on structural indices (Supplementary file 1, section I).

### 3.3 Excess-cost ratio

Under the default cost assumptions at 52 weeks, the excess-cost ratio was defined as 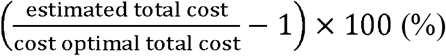. The mean excess-cost ratio was 65.5%. Across the 110 countries the excess-cost ratio had a median of 29.9%, an interquartile range of 15.5% to 96.7%, and a range of 8.0% to 400.2%. Twenty-seven countries had an excess-cost ratio above 100%. The countries with the largest excess-cost ratios at 52 weeks were Peru (400.2%), Czechia (337.5%), and the United Kingdom (235.2%) (Supplementary file 1, section J, Supplementary file 4, Supplementary file 5).

For each of the 26-, 40-, and 52-week horizons, we fitted a descriptive cross-country relationship between the excess-cost ratio and the difference in average transmission reduction, defined as model-inferred minus cost-optimal average reduction. The function is formulated as 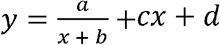, where *y* indicates the excess-cost ratio, *x* indicates this difference in average transmission reduction, with four fitted constants from *a* to *d. y*= 1 means 100% excess-cost ratio. In Figure 3, the shaded band around each curve is a pointwise 95% confidence interval for the fitted cross-country relationship. The confidence interval of excess-cost ratio is large near *x*= 0 and small for *x* > 0.1. To exclude points with a high excess-cost ratio, we identify a costeffective transmission reduction target based on the intersections between *y*= 0.4 and the bestfit curve by *x*_under_ and *x*_over_. This criterion categorises the difference in average transmission reduction into three area; an unstable under-intervention zone (*x*> *x*_under_; lower reduction and higher excess-cost ratio), the cost-effective transmission reduction targets (*x*_under_ ≤*x* ≤*x*_over_; lower excess-cost ratio), and an over-intervention zone (*x*> *x*_over_; higher-reduction and higher excess-cost ratio). The width of the lower excess-cost ratio range increased from 0.136 to 0.201 as the modelled horizon extended. For the 26-, 40-, and 52-week horizons, the differences in average transmission reduction within this range were 0.064-0.200, 0.061-0.246, and 0.052-0.253, respectively. These ranges correspond to 6.4-20.0, 6.1-24.6 and 5.2-25.3 percentage points above the country-specific optima. The 40% excess-cost ratio threshold is an exploratory classification criterion, not a validated cost-effectiveness threshold. The corresponding nonlinear fits had *R*^2^ values of 0.843, 0.547, and 0.701 (Supplementary file 4, Table S4.2).

**Figure 3.**
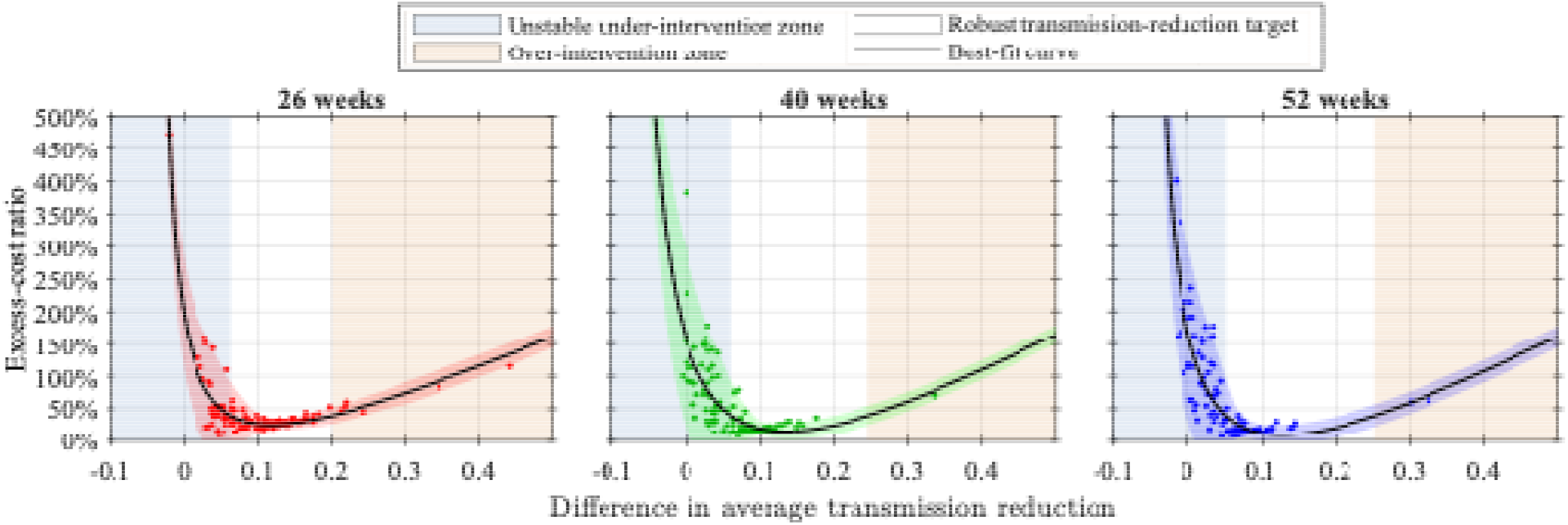
Excess-cost ratio versus difference in average transmission reduction for 110 countries. The x-axis indicates model-inferred average transmission reduction minus cost-optimal average transmission reduction and y-axis indicates percentage increase in model-estimated total cost relative to the cost-optimal total cost. The three panels show 26-, 40-, and 52-week modelled horizons. Circular points are individual countries, the black line is the best-fit rational curve, and the shaded band is the pointwise 95% confidence interval for the fitted relationship, not a country-specific prediction interval. The pale blue region shows the lower-reduction, higher excess-cost ratio range (***x*** < ***x***_**Under**_) and the orange region the higher-reduction, higher excess-cost ratio range (***x*** > ***x***_over_), where the fitted excess-cost ratio exceeds 40% of the modelled optimum. The unshaded region is an exploratory lower excess-cost ratio range, not a validated intervention target. The best-fit curve and confidence interval are drawn continuously across the plotted range: towards the left they rise steeply as the difference in transmission reduction approaches the vertical asymptote of the rational fit (***x*** = −***b***), and towards the right they are extrapolated to 0.5.

Separately from the cross-country fit in Figure 3, we examined costs along the country-specific Pareto frontiers around each optimum (Figure 4). On the lower-reduction side of the optimum, modelled costs were sensitive to small deviations. Starting from the cost-optimal solution (red circle), a five-percentage-point decrease in average transmission reduction leads to an increase in modelled total costs of 137% for Nepal, 72% for Ghana, 125% for the Republic of Korea, and 66% for the United States. In contrast, a five-percentage-point increase in reduction results in a much smaller cost escalation of approximately 6.5% to 8% (Figure 4). Under the modelled cost assumptions, this asymmetry supports evaluating a precautionary margin when achieved transmission reduction is uncertain. It does not establish the size of that margin or a universal prescription for stricter NPIs.

**Figure 4.**
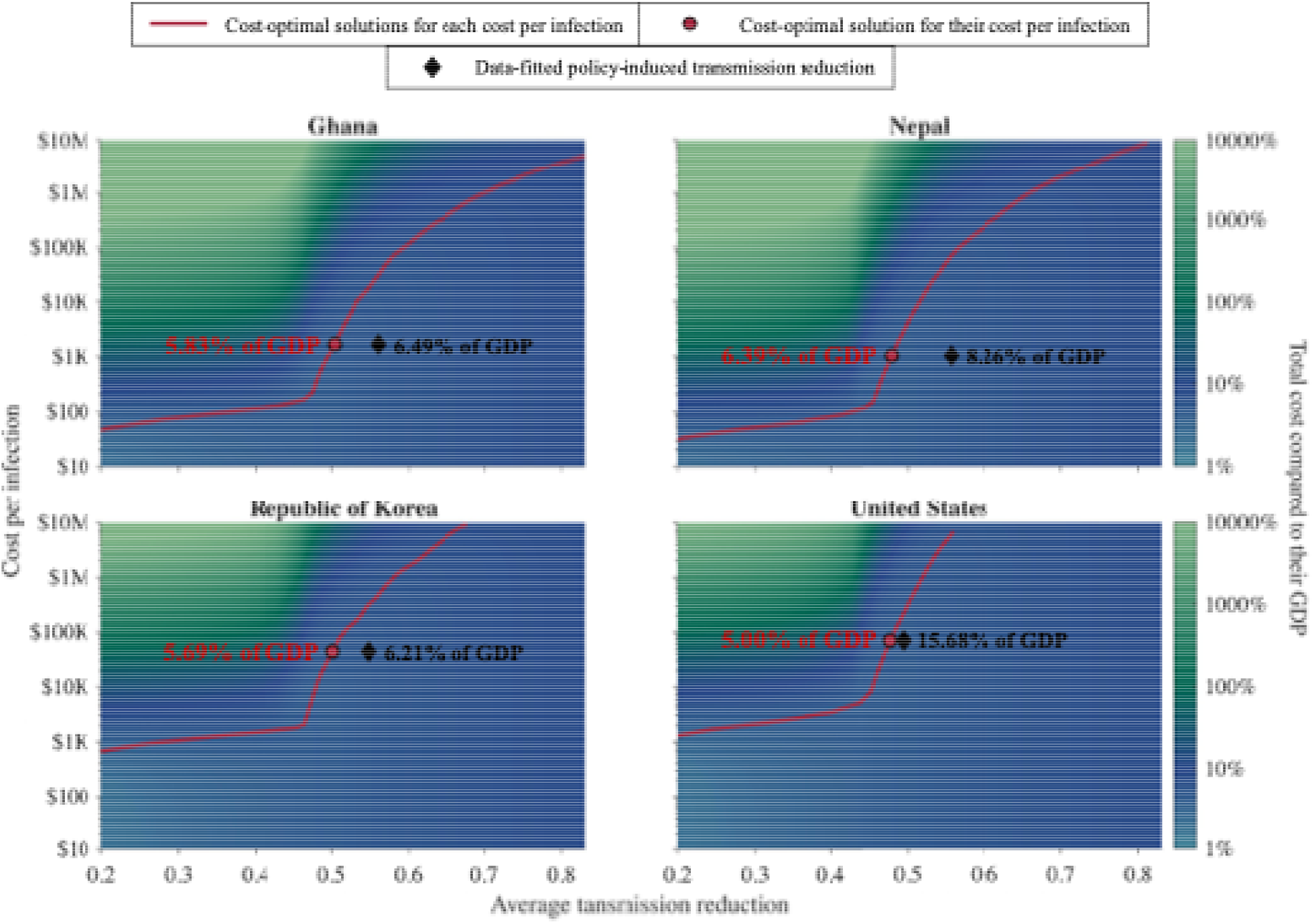
Cost-optimal transmission reduction and total costs across cost per infection in four countries. The x-axis shows average transmission reduction and the logarithmic y-axis shows assumed cost per infection. Background heatmaps show total cost (intervention plus infection costs) for Pareto-optimal strategies as a percentage of each country’s GDP; colours range from blue (1%) to green (>10,000%). Solid pink curves trace the cost-optimal transmission reduction, dark-red circles mark the optimum at each country’s default cost per infection, and black diamonds show model-inferred effective transmission reduction from the data-fitted trajectory. The background cost at a diamond’s coordinates represents a Pareto-efficient strategy, not the total cost of the fitted trajectory.

Comparing the total cost of the cost-optimal solution with the estimated point highlights this practical gap. Under the assumptions of our model, in the United States, although the black diamond indicates a model-inferred transmission reduction similar to the cost-optimal level (red circle), the model-inferred total cost was 15.68% of GDP_total_, which is 3.14 times the theoretical optimum of 5.00% (Supplementary file 1, section J). This suggests that, for the United States, the high volume of infections caused a substantial deviation from the Pareto-optimal infection scale assumed in the background heatmap. In contrast to the risks of under-intervention, Nepal, Ghana, and the Republic of Korea maintained relatively stable total costs as a share of total GDP with model-inferred average transmission reductions above the cost-optimal levels. Their estimated total costs are 6.39%, 5.83%, and 5.69% of GDP_total_, respectively, representing costs only 1.09 to 1.29 times higher than the corresponding optimal solutions (Supplementary file 1, section J, Supplementary file 4).

### 3.4 Association of structural indices with optimal intervention intensity

To provide category-level benchmarks beyond the four example countries, we present a distribution of cost-optimal average transmission reduction across six country-level characteristics. Figure 5 provides approximate category-level benchmarks for cost-optimal intervention intensity, stratified by structural indices, rather than exact country-specific prescriptions. These distributions can be interpreted as indicative ranges reflecting the structural characteristics of each country group. These within-sample distributions do not establish predictive performance for an individual country or a future pandemic (Figure 5). Higher cost per infection corresponds to higher average transmission reduction, and higher index values are generally associated with lower optimal transmission reduction levels (Figure 5, Supplementary file 1, section K). In particular, the range of average transmission reduction becomes narrower when infection costs exceed $10K in the Income and UHC panels. However, region and population size exhibit relatively weak associations with the distribution of average transmission reduction. When the assumed cost per infection is around $1K, the range of optimal transmission reduction is lower but more variable in countries with high index values, consistent with the model’s relative valuation of intervention and infection costs under these fixed-cost scenarios (Figure 5, Supplementary file 1, section K).

**Figure 5.**
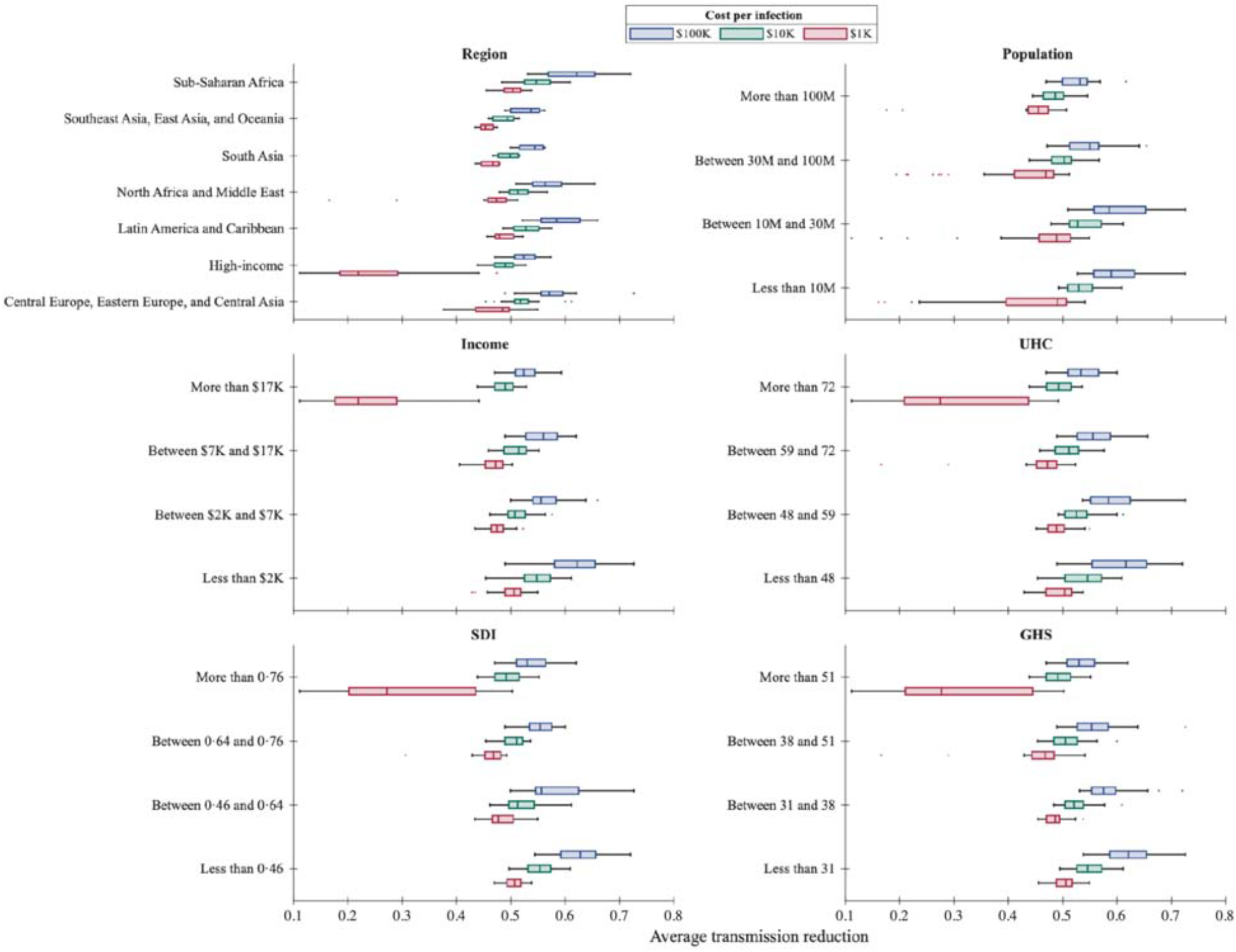
Distribution of cost-optimal average transmission reduction categorised by country-level factors under varying costs per infection. The figure displays box plots for six country-level characteristics: Region, Population, Income, Universal Health Coverage (UHC), Socio-demographic Index (SDI), and Global Health Security (GHS) index. The x-axis represents the cost-optimal average transmission reduction required to minimise the modelled total cost. Each panel illustrates how the distribution of optimal intervention intensity shifts as the assumed cost per infection increases from $1,000 (red) to $10,000 (green) and $100,000 (blue). The boxes represent the interquartile range (IQR), the central vertical lines indicate the median values, and the whiskers demonstrate the overall spread of optimal strategies within each country category.

## 4 Discussion

Three findings are relevant to pandemic preparedness. First, modelled costs increased more steeply below the cost-optimal average transmission reduction than for an equivalent increase above it in the illustrative country analyses (Figure 4). Second, cost-optimal strategies differed across countries and depended on the economic assumptions, arguing against a single universal target (Figure 2). Third, SDI and income showed descriptive associations with the distribution of optima, but these associations did not establish country-specific predictive accuracy (Figure 5; Supplementary file 1, section K). The contribution is an exploratory framework for comparing response options and the consequences of insufficient transmission reduction, rather than an index-based prescription for policy.

Cross-country distributions can provide a starting point for scenario comparisons, but their transferability to a future pandemic remains to be evaluated. Structural indices alone do not specify pathogen transmissibility, surveillance completeness, intervention effectiveness or infection-related costs. Category-level ranges should therefore complement local epidemiological evidence rather than substitute for it, particularly where surveillance capacity is limited. Prospective use would require recalibration as information accumulates and assessment of which feasible interventions could achieve the modelled reduction.

As the modelled horizon extends, inferred average transmission reductions become more similar across countries, while modelled epidemic outcomes show greater differences across SDI and income categories (Supplementary file 1, section G, section H). The narrowing variance in inferred transmission reduction does not establish convergence in implemented policies or in their effectiveness (Figure 1, Supplementary file 1, section G). The widening differences in modelled case burden may also reflect differences in timing, importation and ascertainment (Figure 1). These findings motivate monitoring the joint trajectories of intervention and epidemic outcomes when adapting response strategies (Figure 2, Supplementary file 1, section I).

Associations of SDI and income with modelled outcomes should not be interpreted as evidence that these indices identify the most effective preparedness investments (Supplementary file 1, section F, section H). The analyses are ecological and descriptive, and economic assumptions contribute to the observed patterns. In particular, the fixed dollar-valued infection-cost scenarios in Figure 5 differ from the default GDP-scaled valuation. These results cannot establish the causal benefits of economic development, rank preparedness investments, or determine how international assistance should be allocated.

Model-inferred responses and cost-optimal strategies describe different quantities. In the model-inferred results, countries with higher SDI or income values tended to have lower inferred transmission reduction and higher modelled infection proportions, a pattern that became more apparent as the modelled horizon extended (Figure 1). In cost-optimal solutions, however, higher-index countries generally require lower optimal transmission reduction (Figure 5). The former describes fitted effective transmission reduction, whereas the latter describes model-derived cost-minimising strategies. In the fixed infection-cost scenarios, GDP-scaled intervention costs affect the relative valuation of reducing transmission. However, the Pareto frontier alone does not guarantee that implemented policies were economically coherent, because Figure 1 plots only transmission reduction against the infected individual proportion and does not incorporate economic cost. To evaluate the alignment between model-inferred and cost-optimal strategies directly, we compared each country’s estimated trajectory with its cost-optimal point (Figure 2, Supplementary file 1, section I, Supplementary file 3) and quantified the total-cost difference between the two (Supplementary file 1, section J). These gaps showed no consistent relationship with any of the index-based categories, so these results do not demonstrate that structural development predicts proximity to the modelled optimum. This highlights the need to better understand the feasibility of implementing optimal strategies across different contexts, for both low- and high-income countries. In particular, even when a model-derived transmission reduction target is available, predicting which combination of non-pharmaceutical interventions will achieve that target is itself difficult, adding a further layer of uncertainty to translating these targets into specific policies.

The asymmetric costs around the modelled optimum support evaluating precautionary response margins when achieved transmission reduction is uncertain. In the four illustrative countries, a five-percentage-point decrease below the optimum increased modelled costs by 66-137%, whereas an equivalent increase raised costs by approximately 6.5-8% (Figure 4; Supplementary file 1, section J). This comparison is conditional on the model’s cost structure and Pareto frontier. For global health policy, a modelled cost optimum is also not an affordability or equity assessment. Income-scaled mortality valuations are economic modelling conventions, not a statement that lives have different intrinsic value, and national aggregates do not describe how burdens are distributed within countries. Any proposed response margin should therefore be assessed alongside feasibility, livelihood protection and continuity of essential services, rather than interpreted as a requirement for resource-constrained countries to sustain stricter restrictions.

Six limitations need to be acknowledged. First, under-reporting and testing heterogeneity represent a major limitation. Confirmed case and death counts are not equally comparable across countries: low reported incidence may reflect low transmission, limited testing capacity, under-reporting, delayed reporting, or different surveillance systems. Countries with weaker surveillance infrastructure may appear to have lower case burdens, which could bias the estimation of transmission reduction parameters. We partially address this through sensitivity analyses (Supplementary file 1, section B) and by using cumulative confirmed cases rather than daily counts, which are less sensitive to reporting delays; however, systematic cross-country differences in ascertainment rates remain a source of uncertainty. Furthermore, our model does not explicitly account for undetected infections, which limits our ability to analyse specific reporting dynamics. Applying the CFR on the modelled reported-case ascertainment scale does not recover the population infection fatality ratio (IFR). Unequal detection of cases and deaths can distort cross-country transmission estimates and mortality-related cost comparisons. Second, cost-optimal solutions depend on assumed economic parameters, including GDP-derived valuations and a per-infection cost combining quarantine and mortality components; alternative assumptions about the value of a statistical life, treatment cost, or isolation duration would shift individual country optima, and the robustness of qualitative patterns should be assessed across the full range of relevant assumptions. Third, structural indices such as SDI and GDP show associations with the outcomes studied but cannot fully represent political feasibility, public compliance, or health-system surge capacity. Fourth, the model does not account for behavioural feedback. It also assumes that isolated individuals do not transmit, whereas in practice isolation is imperfect. Fifth, the intervention cost function is deliberately simple. It models cost as increasing linearly alongside the average transmission reduction. In reality, different interventions achieving the same reduction can have very different, non-linear costs. Furthermore, intervention costs vary across countries beyond the GDP scaling applied here. The effect of alternative cost functions on both the numerical optima and the direction and magnitude of cost asymmetry requires evaluation. Therefore, constructing a more detailed cost function is an important direction for future work. Sixth, the model does not account for differences in population age structure. As a result, mortality and its related costs might be lower in countries with younger populations^33^. Incorporating age-structured mortality serves as a planned extension for future work.

## Conclusions

cross 110 countries, this study compared model-inferred transmission reduction with country-specific cost-optimal strategies to inform early pandemic response. Under the modelled assumptions, insufficient transmission reduction incurred greater cost penalties than comparable deviations above the optimum, supporting the evaluation of precautionary response margins when intervention effectiveness is uncertain. In the descriptive cross-country analyses, average transmission reductions of 6.4-20.0, 6.1-24.6, and 5.2-25.3 percentage points above country-specific optima corresponded to fitted excess-cost ratios below 40% over the 26-, 40-, and 52-week horizons, respectively. These ranges may serve as preliminary reference points for assessing country-specific transmission reduction strategies. Associations between national socioeconomic and health-system indices and cost-optimal strategies may also inform response planning, although country-specific targets require local epidemiological and economic calibration. Together, these findings provide a quantitative framework for developing transmission reduction strategies that balance intervention and infection costs while accounting for uncertainty and national circumstances.

## List of abbreviations

CFR: case fatality ratio
COVID-19: coronavirus disease 2019
DRAM: Delayed Rejection Adaptive Metropolis
ESS: Effective Sample Size
GBD: Global Burden of Disease
GDP: gross domestic product
GHS: Global Health Security Index
IFR: infection fatality rate
IMODE: improved multi-operator differential evolution
IQR: interquartile range
ISO: International Organization for Standardization
LMICs: low- and middle-income countries
MCMC: Markov Chain Monte Carlo
NPIs: non-pharmaceutical interventions
NSGA-II: non-dominated sorting genetic algorithm II
SDI: Socio-demographic Index
SEIQRD: susceptible-exposed-infectious-quarantined/isolated-recovered-deceased
UHC: universal health coverage
USD: United States dollars
VSL: value of a statistical life
WHO: World Health Organization

## Declarations

### Ethics approval and consent to participate

This study used publicly available aggregate country-level data and did not require ethics approval.

### Consent for publication

Not applicable.

### Data availability statement

All data sources used in this study are publicly available and cited in the Methods. Country-level epidemiological data were obtained from Our World in Data, economic and demographic indices from the World Bank, and health security indices from their respective databases. The MATLAB code for the SEIQRD model, multi-objective optimisation, and cost-optimisation, together with the processed datasets and Supplementary files, is available in a GitHub repository at https://github.com/ljm1729-scholar/110Countries. The data and code are also archived on Zenodo at https://doi.org/10.5281/zenodo.21982931.

## Authors’ contributions

JL and RM were responsible for the concept and design. JL, RM, VMPM, RNT, GC, and EJ were responsible for the drafting of the manuscript. JL, RM, VMPM, RNT, GC, and EJ were responsible for the acquisition, analysis, or interpretation of data. JL, RM, VMPM, and GC were responsible for the statistical analysis. GC and EJ were responsible for funding. JL, GC, and EJ were responsible for the administrative, technical, or material support. All authors directly accessed and verified the underlying data reported in the manuscript. All authors had access to all the data in this study. All authors read and approved the final version of the manuscript. EJ has final responsibility for the decision to submit the manuscript for publication.

## Acknowledgements

Not applicable.

## Supplementary files

### Supplementary file 1

**File name:** Supplementary file 1.pdf

**File format:** Portable Document Format (.pdf)

**Title:** Supplementary data, methods, and analyses

**Description:** Contains Data for the 110 countries; mathematical modelling; parameter-estimation; multi-objective optimization, cost-benefit analyses; correlation analyses; characteristics of Figure 1; categorization of the points; difference trends; cost-optimal solutions under various cost per infection, generalised cost-optimal intervention results (sections A-K).

### Supplementary file 2

**File name:** Supplementary file 2.pdf

**File format:** Portable Document Format (.pdf)

**Title:** Structural and practical identifiability

**Description:** Provides StructuralIdentifiability.jl code and structural-identifiability results for the SEIQRD model, together with MCMC/DRAM practical-identifiability diagnostics and trace plots. Joint and marginal posterior distributions are provided for Nepal, Ghana, the Republic of Korea, and the United States. The attached code in Zenodo can generate the joint and marginal posterior distribution for all countries.

### Supplementary file 3

**File name:** Supplementary file 3.pdf

**File format:** Portable Document Format (.pdf)

**Title:** Trajectories of transmission reduction and confirmed cases over time for 110 countries

**Description:** Summarises convergence, divergence, and non-monotonic patterns between data-fitted and cost-optimal results across 26-, 40-, and 52-week pre-vaccination horizons and presents country-level trajectory figures for all 110 countries.

### Supplementary file 4

**File name:** Supplementary file 4.pdf

**File format:** Portable Document Format (.pdf)

**Title:** Cost-optimal strategies across the range of cost per infection

**Description:** Reports estimated and cost-optimal transmission reductions, total costs, and excess-cost ratio across 26-, 40-, and 52-week pre-vaccination horizons for 110 countries; rational-curve excess cost analyses; intervention-zone thresholds; and country-level cost-optimal solutions across varying cost per infection assumptions.

### Supplementary file 5

**File name:** Supplementary file 5.xlsx

**File format:** Microsoft Excel workbook (.xlsx)

**Title:** 110-country cost-optimisation dataset with global country indices

**Description:** Contains country-level demographic, economic, health-system, institutional, and environmental indices for the 2019 reference year across 110 countries, combined with COVID-19 outbreak data and the study’s estimated results and cost-optimisation outputs. The data file includes README, Data, Metadata, ReferenceYears, and Checks sheets.

