## Supplementary file 1 for "Cost-optimal transmission reduction in early pandemic response: a multi-objective health-economic modelling study across 110 countries"

### [List of Supplementary 1]

**Supplementary 1A. Data for the 110 countries**

**Supplementary 1B. Mathematical modelling**

**Supplementary 1C. Parameter estimation**

**Supplementary 1D. Multi-objective optimisation**

**Supplementary 1E. Cost-benefit analysis**

**Supplementary 1F. Correlation between the country-specific indices and epidemic-related outcomes**

**Supplementary 1G. Characteristics of Figure 1**

**Supplementary 1H. Categorisation of the points**

**Supplementary 1I. Difference trends**

**Supplementary 1J. Cost-optimal solutions under various cost per infection**

**Supplementary 1K. Generalised cost-optimal intervention results for structural index category**

#### Supplementary 1A. Data for the 110 countries

Table S1 presents data for 110 countries with populations exceeding five million, grouped by six Global Burden of Disease (GBD) super-regions. Each entry includes ISO 3166-1 country codes and demographic, economic, and health indicators. Population (2019) and GDP per capita (2019, USD) were obtained from the World Bank. Universal Health Coverage (UHC; 2019) was obtained from the WHO. The Socio-demographic Index (SDI; 2019) was obtained from the Global Health Data Exchange. The Global Health Security Index (GHS; 2021) and its six categories were obtained from the GHS Index database: GHS1 (Prevention), GHS2 (Detection and Reporting), GHS3 (Rapid Response), GHS4 (Health System), GHS5 (Compliance with International Norms), and GHS6 (Risk Environment). Confirmed cases and deaths are cumulative totals for 52 weeks from each country's index case, obtained from Our World in Data. The case fatality ratio (CFR) was calculated as cumulative deaths divided by cumulative confirmed cases.

To assess the developmental and health security landscape of these nations, the table includes three critical indices: Universal Health Coverage (UHC), the Socio-demographic Index (SDI), and the Global Health Security (GHS) Index. These metrics provide context for the final columns of the table, which track epidemiological impact through 52-week totals of confirmed cases and deaths. The relationship between these figures is summarized as a Case Fatality Rate (CFR), offering a standardized measure of mortality relative to reported infections across regions ranging from high-income nations to Sub-Saharan Africa.

**Table S1.1: Data for 110 countries categorised by the Global Burden of Disease (GBD) super region.**

| Country name | 2-letter Country code[*] | 3-letter Country code[*] | Population (million) | GDP per capita (USD) | UHC[†] | SDI[‡] | GHS[§] | 52w confirmed[¤] | 52w deaths[¤] | CFR[#] |
|---|---|---|---|---|---|---|---|---|---|---|
| **Central Europe, Eastern Europe, and Central Asia** | | | | | | | | | | |
| Azerbaijan | AZ | AZE | 10.13 | 4806 | 48.1814 | 0.6898 | 34.2 | 233989 | 3213 | 1.37% |
| Belarus | BY | BLR | 9.34 | 6838 | 70.4622 | 0.7805 | 41.8 | 285959 | 1966 | 0.69% |
| Bulgaria | BG | BGR | 6.52 | 9839 | 62.5567 | 0.7624 | 61.4 | 259811 | 10593 | 4.08% |
| Czechia | CZ | CZE | 10.49 | 24063 | 81.944 | 0.8248 | 55 | 1334950 | 22257 | 1.67% |
| Hungary | HU | HUN | 9.60 | 16786 | 72.0281 | 0.7849 | 55 | 466017 | 15873 | 3.41% |
| Kazakhstan | KZ | KAZ | 19.12 | 9457 | 59.2368 | 0.7208 | 44.7 | 273521 | 3456 | 1.26% |
| Kyrgyzstan | KG | KGZ | 6.78 | 1422 | 52.9515 | 0.5965 | 43 | 82941 | 1024 | 1.23% |
| Poland | PL | POL | 37.63 | 15875 | 72.6564 | 0.8048 | 54.3 | 1797087 | 45311 | 2.52% |
| Romania | RO | ROU | 18.80 | 12992 | 69.5851 | 0.7603 | 45.5 | 799164 | 20287 | 2.54% |
| Russia | RU | RUS | 146.03 | 11448 | 68.9736 | 0.8037 | 47.1 | 3850439 | 73182 | 1.90% |
| Serbia | RS | SRB | 6.65 | 7756 | 63.3494 | 0.7858 | 45 | 482397 | 4542 | 0.94% |
| Slovakia | SK | SVK | 5.38 | 19406 | 77.9829 | 0.8064 | 52 | 323390 | 7836 | 2.42% |
| Tajikistan | TJ | TJK | 10.14 | 871 | 47.8775 | 0.5338 | 29.8 | 13714 | 91 | 0.66% |
| Ukraine | UA | UKR | 35.15 | 3620 | 56.7521 | 0.758 | 36.9 | 1425267 | 25863 | 1.81% |
| Uzbekistan | UZ | UZB | 36.72 | 2041 | 42.1848 | 0.6558 | 37.7 | 97217 | 649 | 0.67% |
| **High-income** | | | | | | | | | | |
| Australia | AU | AUS | 26.45 | 54973 | 89.4234 | 0.8406 | 73.2 | 28766 | 923 | 3.21% |
| Austria | AT | AUT | 8.98 | 49886 | 86.3701 | 0.8393 | 57.4 | 448598 | 10309 | 2.30% |
| Belgium | BE | BEL | 11.80 | 46717 | 87.3014 | 0.8495 | 61.9 | 721892 | 21595 | 2.99% |
| Canada | CA | CAN | 38.93 | 46353 | 90.3017 | 0.8706 | 67.6 | 737407 | 18697 | 2.54% |
| Denmark | DK | DNK | 5.95 | 59404 | 84.1397 | 0.8916 | 67.3 | 210732 | 2358 | 1.12% |
| Finland | FI | FIN | 5.55 | 48358 | 91.3486 | 0.8554 | 72 | 49713 | 724 | 1.46% |
| France | FR | FRA | 65.63 | 40408 | 90.7662 | 0.8338 | 62.6 | 2798460 | 76746 | 2.74% |
| Germany | DE | DEU | 83.22 | 47624 | 86.2487 | 0.8997 | 65.7 | 1783131 | 52753 | 2.96% |
| Greece | GR | GRC | 10.43 | 19335 | 80.1403 | 0.7881 | 50.6 | 197162 | 6552 | 3.32% |
| Israel | IL | ISR | 9.73 | 44141 | 81.3846 | 0.8032 | 50.7 | 722568 | 5361 | 0.74% |
| Italy | IT | ITA | 58.87 | 33813 | 88.895 | 0.8015 | 51.9 | 2541783 | 88279 | 3.47% |
| Japan | JP | JPN | 124.37 | 40416 | 96.3409 | 0.8671 | 58.8 | 328294 | 4446 | 1.35% |
| Netherlands | NL | NLD | 17.56 | 53555 | 89.5884 | 0.8846 | 67.7 | 1093519 | 15833 | 1.45% |
| Portugal | PT | PRT | 10.20 | 23343 | 83.533 | 0.7381 | 58.7 | 766862 | 16020 | 2.09% |

| | | | | | | | | | | |
|---|---|---|---|---|---|---|---|---|---|---|
| Republic of Korea | KR | KOR | 51.21 | 31902 | 89.1617 | 0.8807 | 65.9 | 72718 | 1249 | 1.72% |
| Spain | ES | ESP | 49.08 | 29776 | 90.0056 | 0.7639 | 60.4 | 2413174 | 59153 | 2.45% |
| United Kingdom | GB | GBR | 68.27 | 42663 | 87.9 | 0.8541 | 68.3 | 3880724 | 129420 | 3.33% |
| United States | US | USA | 340.11 | 65605 | 82.1377 | 0.8586 | 76.2 | 24794408 | 429874 | 1.73% |
| **Latin America and Caribbean** | | | | | | | | | | |
| Argentina | AR | ARG | 46.04 | 9956 | 61.1584 | 0.719 | 56.1 | 2226676 | 58111 | 2.61% |
| Bolivia | BO | BOL | 12.12 | 3504 | 52.3988 | 0.5907 | 32 | 258324 | 11903 | 4.61% |
| Brazil | BR | BRA | 215.35 | 9030 | 64.8276 | 0.6453 | 51 | 10455630 | 252835 | 2.42% |
| Chile | CL | CHL | 19.60 | 14496 | 74.3496 | 0.7651 | 53 | 821418 | 20476 | 2.49% |
| Colombia | CO | COL | 52.16 | 6473 | 74.3969 | 0.646 | 50 | 2269582 | 60300 | 2.66% |
| Costa Rica | CR | CRI | 5.27 | 12885 | 79.015 | 0.69 | 40.5 | 206640 | 2833 | 1.37% |
| Cuba | CU | CUB | 11.11 | 9232 | 72.5922 | 0.6617 | 32.3 | 60558 | 366 | 0.60% |
| Dominican Republic | DO | DOM | 11.33 | 8164 | 52.4998 | 0.6119 | 35.8 | 242660 | 3162 | 1.30% |
| Ecuador | EC | ECU | 18.12 | 6205 | 64.4544 | 0.6518 | 48.2 | 292943 | 16020 | 5.47% |
| El Salvador | SV | SLV | 6.52 | 4320 | 61.6782 | 0.554 | 42.9 | 62531 | 1975 | 3.16% |
| Guatemala | GT | GTM | 18.12 | 4512 | 52.0988 | 0.5286 | 31 | 182679 | 6563 | 3.59% |
| Haiti | HT | HTI | 11.74 | 1352 | 35.8119 | 0.4443 | 30.1 | 12758 | 251 | 1.97% |
| Honduras | HN | HND | 10.28 | 2502 | 54.2843 | 0.5042 | 26.3 | 177168 | 4325 | 2.44% |
| Mexico | MX | MEX | 130.29 | 10370 | 61.4368 | 0.6551 | 55.1 | 2094677 | 206535 | 9.86% |
| Paraguay | PY | PRY | 7.45 | 5821 | 63.3513 | 0.6261 | 39.8 | 177593 | 3436 | 1.93% |
| Peru | PE | PER | 34.05 | 7037 | 75.7594 | 0.6554 | 53.8 | 1358294 | 126092 | 9.28% |
| Venezuela | VE | VEN | 33.76 | 5158 | 60.9675 | 0.6023 | 21.4 | 144786 | 1422 | 0.98% |
| **North Africa and Middle East** | | | | | | | | | | |
| Afghanistan | AF | AFG | 41.13 | 497 | 39.2945 | 0.3225 | 27.9 | 55714 | 2443 | 4.38% |
| Algeria | DZ | DZA | 45.61 | 4469 | 64.8553 | 0.6482 | 26.8 | 112960 | 2979 | 2.64% |
| Egypt | EG | EGY | 109.26 | 2963 | 54.7965 | 0.5877 | 30.3 | 173202 | 9935 | 5.74% |
| Iran | IR | IRN | 89.52 | 3831 | 69.5146 | 0.6867 | 39.5 | 1566081 | 59409 | 3.79% |
| Iraq | IQ | IRQ | 45.51 | 5672 | 57.7248 | 0.6436 | 23.3 | 692241 | 13383 | 1.93% |
| Jordan | JO | JOR | 11.44 | 4170 | 69.9667 | 0.715 | 41.2 | 421415 | 4900 | 1.16% |
| Lebanon | LB | LBN | 5.73 | 8906 | 74.5332 | 0.7386 | 36.8 | 353371 | 4297 | 1.22% |
| Libya | LY | LBY | 7.02 | 9963 | 66.3277 | 0.7162 | 23.3 | 156116 | 2602 | 1.67% |
| Morocco | MA | MAR | 37.96 | 3508 | 58.0323 | 0.5478 | 35.6 | 485974 | 8676 | 1.79% |
| Saudi Arabia | SA | SAU | 36.95 | 27893 | 64.1955 | 0.8061 | 45 | 379474 | 6524 | 1.72% |
| Sudan | SD | SDN | 50.45 | 710 | 51.8347 | 0.525 | 30 | 30946 | 1953 | 6.31% |
| Syria | SY | SYR | 22.13 | 1110 | 57.5646 | 0.6133 | 18.7 | 18356 | 1227 | 6.68% |
| Tunisia | TN | TUN | 12.30 | 3529 | 68.1057 | 0.6732 | 32.1 | 237028 | 8167 | 3.45% |
| Turkey | TR | TUR | 85.66 | 9215 | 69.2104 | 0.6983 | 49.8 | 2866012 | 29421 | 1.03% |
| United Arab Emirates | AE | ARE | 10.26 | 44251 | 63.3574 | 0.843 | 40.1 | 300661 | 838 | 0.28% |
| Yemen | YE | YEM | 34.45 | 623 | 49.0458 | 0.444 | 19.9 | 5280 | 1032 | 19.55% |
| **Sub-Saharan Africa** | | | | | | | | | | |
| Angola | AO | AGO | 36.68 | 2190 | 39.1577 | 0.4405 | 25.2 | 22579 | 540 | 2.39% |
| Benin | BJ | BEN | 13.76 | 1131 | 44.6243 | 0.36 | 27 | 6818 | 90 | 1.32% |
| Burkina Faso | BF | BFA | 23.07 | 765 | 41.7968 | 0.2743 | 34.4 | 12372 | 144 | 1.16% |
| Burundi | BI | BDI | 13.76 | 210 | 49.94 | 0.2834 | 22.7 | 2964 | 6 | 0.20% |
| Cameroon | CM | CMR | 27.20 | 1555 | 42.29 | 0.4642 | 32.2 | 38988 | 588 | 1.51% |
| Chad | TD | TCD | 17.18 | 678 | 31.3724 | 0.2317 | 24.5 | 4410 | 157 | 3.56% |
| Cote d'Ivoire | CI | CIV | 28.71 | 2142 | 43.0401 | 0.4117 | 32.9 | 37304 | 211 | 0.57% |
| Democratic Republic of Congo | CD | COD | 105.79 | 557 | 45.1687 | 0.3693 | 26 | 27013 | 717 | 2.65% |
| Ethiopia | ET | ETH | 126.53 | 829 | 46.5216 | 0.3464 | 37.4 | 174054 | 2540 | 1.46% |
| Ghana | GH | GHA | 33.48 | 2186 | 49.1393 | 0.5516 | 31.6 | 88421 | 705 | 0.80% |
| Guinea | GN | GIN | 14.41 | 1031 | 32.3346 | 0.3228 | 28.5 | 17592 | 100 | 0.57% |
| Kenya | KE | KEN | 55.34 | 1960 | 51.6469 | 0.508 | 43.1 | 112805 | 1908 | 1.69% |
| Liberia | LR | LBR | 5.19 | 658 | 47.5997 | 0.3434 | 34.5 | 2054 | 91 | 4.43% |
| Madagascar | MG | MDG | 30.33 | 500 | 39.6908 | 0.3842 | 30.9 | 22484 | 351 | 1.56% |
| Malawi | MW | MWI | 22.98 | 581 | 55.5208 | 0.3701 | 27.8 | 33647 | 1120 | 3.33% |
| Mali | ML | MLI | 24.69 | 820 | 40.6607 | 0.2563 | 30.6 | 9773 | 376 | 3.85% |
| Mozambique | MZ | MOZ | 34.63 | 519 | 44.044 | 0.3132 | 29.6 | 67011 | 762 | 1.14% |
| Niger | NE | NER | 29.45 | 562 | 35.0256 | 0.1609 | 29.7 | 4918 | 185 | 3.76% |
| Nigeria | NG | NGA | 223.80 | 2265 | 38.339 | 0.4888 | 37 | 155417 | 1905 | 1.23% |
| Rwanda | RW | RWA | 14.09 | 810 | 59.3591 | 0.4231 | 31.1 | 20143 | 276 | 1.37% |
| Senegal | SN | SEN | 18.27 | 1431 | 49.6099 | 0.394 | 35.9 | 35632 | 919 | 2.58% |

| | | | | | | | | | | |
|---|---|---|---|---|---|---|---|---|---|---|
| Sierra Leone | SL | SLE | 8.79 | 844 | 42.1196 | 0.3458 | 34.1 | 3987 | 79 | 1.98% |
| Somalia | SO | SOM | 18.14 | 540 | 23.9397 | 0.075 | 17.9 | 9968 | 419 | 4.20% |
| South Africa | ZA | ZAF | 63.02 | 6534 | 59.7274 | 0.674 | 47.5 | 1520206 | 50647 | 3.33% |
| South Sudan | SS | SSD | 13.25 | 412 | 41.6941 | 0.2757 | 21.6 | 10372 | 114 | 1.10% |
| Tanzania | TZ | TZA | 61.74 | 1063 | 55.2493 | 0.4302 | 32.2 | 509 | 21 | 4.13% |
| Togo | TG | TGO | 9.60 | 826 | 42.8086 | 0.3957 | 26.1 | 7521 | 90 | 1.20% |
| Uganda | UG | UGA | 48.58 | 822 | 52.7479 | 0.4084 | 39 | 40688 | 332 | 0.82% |
| Zambia | ZM | ZMB | 20.57 | 1259 | 52.6937 | 0.4879 | 28 | 86273 | 1178 | 1.37% |
| Zimbabwe | ZW | ZWE | 17.32 | 1684 | 54.4611 | 0.4684 | 33.4 | 36662 | 1510 | 4.12% |
| **Southeast Asia, East Asia, and Oceania** | | | | | | | | | | |
| Bangladesh | BD | BGD | 169.38 | 2130 | 53.8827 | 0.4733 | 35.4 | 550330 | 8462 | 1.54% |
| Cambodia | KH | KHM | 16.59 | 2226 | 57.0794 | 0.4636 | 31 | 466 | 0 | 0.00% |
| China | CN | CHN | 1438.07 | 10144 | 69.7123 | 0.7037 | 49 | 96972 | 4791 | 4.94% |
| India | IN | IND | 1425.42 | 2041 | 46.8261 | 0.5608 | 43.6 | 10746183 | 154274 | 1.44% |
| Indonesia | ID | IDN | 281.19 | 4107 | 48.7277 | 0.6469 | 49.2 | 1379662 | 37266 | 2.70% |
| Laos | LA | LAO | 7.31 | 2589 | 43.8545 | 0.4788 | 32.8 | 49 | 0 | 0.00% |
| Malaysia | MY | MYS | 34.05 | 10920 | 66.5736 | 0.7346 | 55.1 | 183801 | 667 | 0.36% |
| Myanmar | MM | MMR | 51.32 | 1426 | 46.9541 | 0.5241 | 37.8 | 142385 | 3206 | 2.25% |
| Nepal | NP | NPL | 30.55 | 1203 | 47.2804 | 0.422 | 35.6 | 269450 | 2934 | 1.09% |
| Pakistan | PK | PAK | 241.50 | 1390 | 39.1679 | 0.491 | 31.3 | 578797 | 12837 | 2.22% |
| Philippines | PH | PHL | 114.12 | 3401 | 54.7117 | 0.6385 | 43.5 | 525614 | 10669 | 2.03% |
| Sri Lanka | LK | LKA | 21.41 | 4083 | 65.5627 | 0.6935 | 33.1 | 64157 | 316 | 0.49% |
| Thailand | TH | THA | 65.93 | 7606 | 71.6003 | 0.6753 | 68.9 | 12054 | 70 | 0.58% |
| Vietnam | VN | VNM | 101.34 | 3441 | 59.7068 | 0.6177 | 42.2 | 1548 | 35 | 2.26% |

*International Organization for Standardization (ISO) 3166-1 country code
†Universal health coverage (UHC)
‡Socio-demographic index (SDI)
§Global Health Security Index (GHS; 2019). GHS_1: Prevention; GHS_2: Detection and Reporting; GHS_3: Rapid Response; GHS_4: Health System; GHS_5: Compliance with International Norms; GHS_6: Risk Environment
¤Confirmed and death cases for 52 weeks from the index case
#Case fatality rate (CFR)

The first set of charts presents basic national characteristics, showing that population and GDP per capita vary widely across the dataset. The population distribution is skewed, with most countries clustered below 100 million and a few outliers reaching 1.5 billion. GDP per capita spans a broad range from roughly $200 to $70K. Additionally, the Cost per infection histogram shows that the economic burden of the disease varies significantly, with a concentration of countries between $1,000 and $100K on a log scale.

The remaining charts highlight health infrastructure and epidemiological results. The histograms for Universal Health Coverage (UHC) and the Socio-demographic Index (SDI) show broad distributions, with UHC values mostly between 40 and 60, and SDI heavily clustered between 0.6 and 0.9. Conversely, the Global Health Security (GHS) index is mainly concentrated in the 30-40 range, indicating similar levels of health security preparedness across many countries. Lastly, mortality and infection rates are represented by the Case Fatality Rate (CFR) and the proportion of confirmed cases. Most countries have a CFR below 0.04, but the proportion of confirmed cases varies widely, covering multiple orders of magnitude on a logarithmic scale.

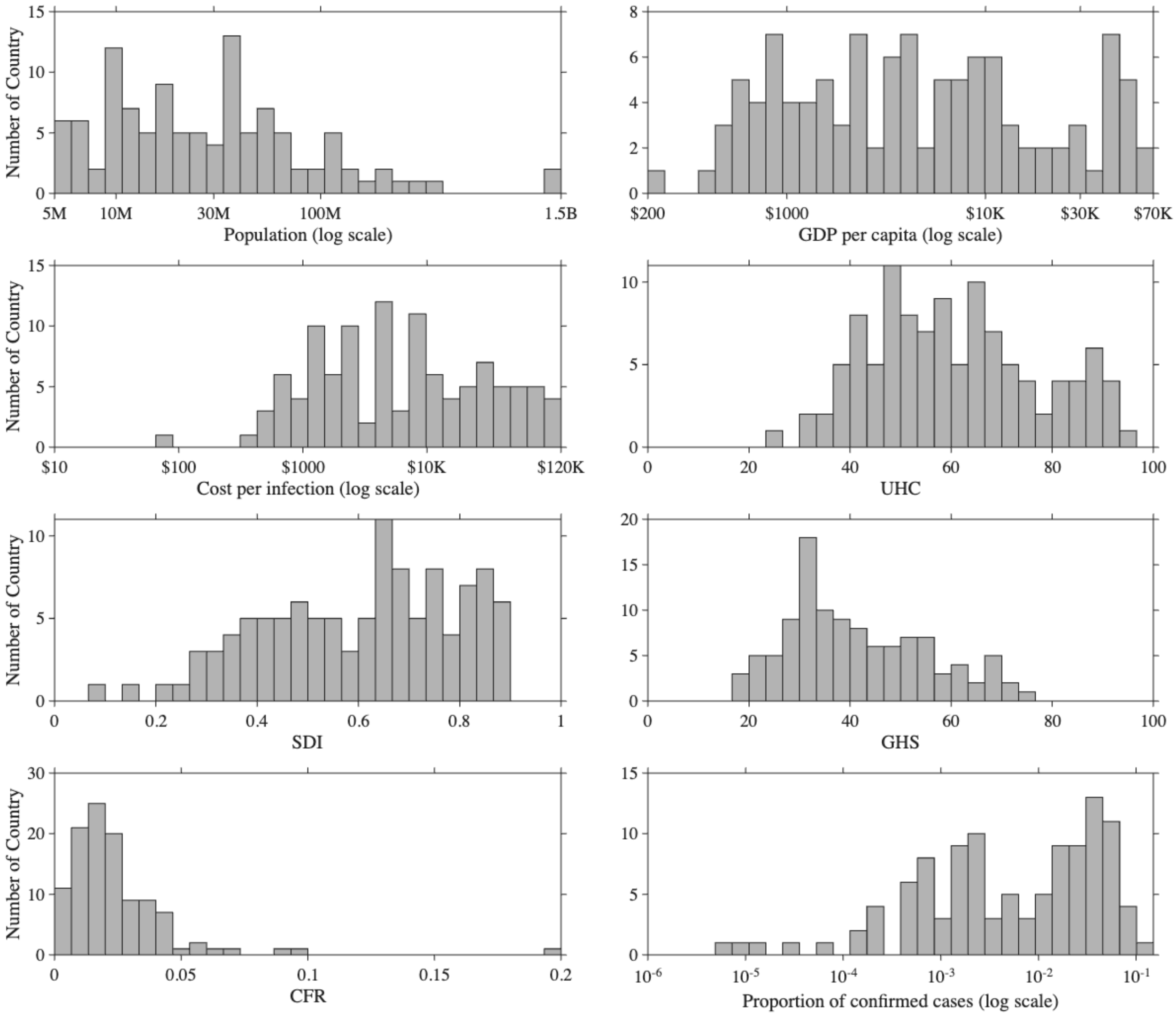


**Figure S1.1. Distribution of Demographic, Socioeconomic, and Health Indicators Across 110 Countries.**

#### Supplementary 1B. Mathematical modelling

We assumed the beginning of a pandemic that doesn't have a developed vaccine yet.[1,2] When we think of the origin of the disease, it could be a spillover from a wild animal or a widespread outbreak from an isolated community.[3-5] Also, it is not always true that the primary case, the first imported case, is identical to the index case, the first observed case. We assumed that most primary cases are originated from the outside of the country. The unknown epidemic occurs once the primary cases spread the disease, and after a period proportional to the disease's latent period, the country identifies an index case. We assume the period from the primary case to the index case is two weeks. Since there is no vaccine against the disease, the government has no choice but to implement non-pharmaceutical interventions, like it did during the COVID-19 pandemic. We assume these interventions are changed every two weeks, but the public continuously changes their behavior to reduce transmission. Regarding vaccine development, the government can adopt different strategies, but it took about a year during COVID-19. We assume the period from the index case to vaccine deployment can be 26, 40, or 52 weeks in the future, which is the same or shorter than in previous pandemics.

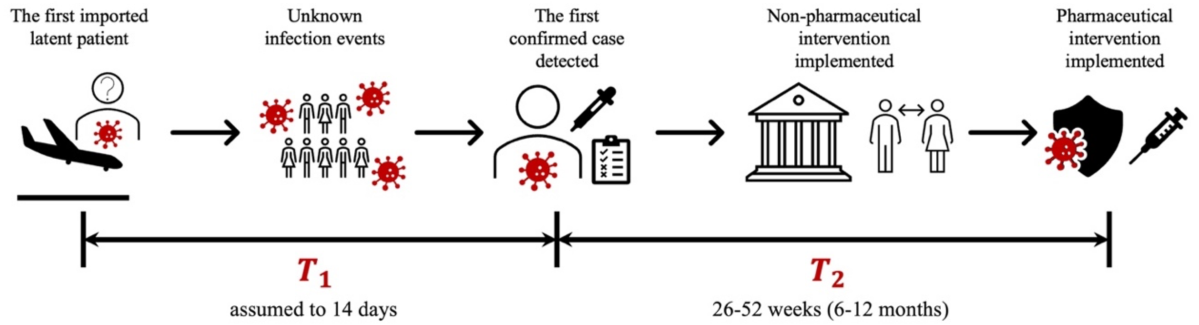


**Figure S1.2. Timeline of pandemic progression and intervention, divided into two distinct intervals labeled $T_1$ and $T_2$.** The process begins on the left with the arrival of the first imported latent patient, followed by a period of unknown infection events where the virus spreads undetected within the community. This initial phase, $T_1$, is assumed to last 14 days and concludes with the detection of the first confirmed case. The timeline then transitions to phase $T_2$, spanning 26 to 52 weeks (6-12 months), which details the public health response; this period starts with the implementation of non-pharmaceutical interventions (such as social distancing and institutional controls) and culminates in the eventual deployment of pharmaceutical interventions, represented by a shield and syringe.

We adopt the deterministic SEIQRD model to simulate an epidemic in a country on an emerging respiratory infectious disease pandemic. The population is partitioned into Susceptible ($S$, green), Exposed ($E$), Infectious ($I$, red), Quarantined/Isolated ($Q$), Recovered ($R$), and Deceased ($D$). Susceptibles become exposed at rate $\lambda(t)S$, where $\lambda(t)$ is the time-varying force of infection (red dashed arrow) defined in the figure as $\lambda(t) = (1 - \mu(t))\, R_\alpha I/N$ with $N = S + E + I + R$; here $\mu(t)$ denotes the intervention (mitigation) intensity and the term $R_\alpha I/N$ captures transmission potential proportional to infectious prevalence. Imported infections enter the system at value $\xi$ (black dashed arrow to E; import from abroad). Exposed individuals progress to infectious at rate κE; infectious individuals are isolated at rate αI. Isolated cases leave $Q$ at rate $\gamma Q$, recovering with probability $1 - f$ to $R$ at $(1 - f)\gamma Q$ or dying with probability $f$ to $D$ at $f\gamma Q$. Unless otherwise specified, parameters $\beta, \kappa, \alpha, f, \gamma$ are fixed from prior literature, while $\xi$ and $\mu(t)$ are estimated from data. Arrows indicate flow terms; solid lines represent within-population transitions, black dashed lines indicate importation, and red dashed lines denote infection pressure.

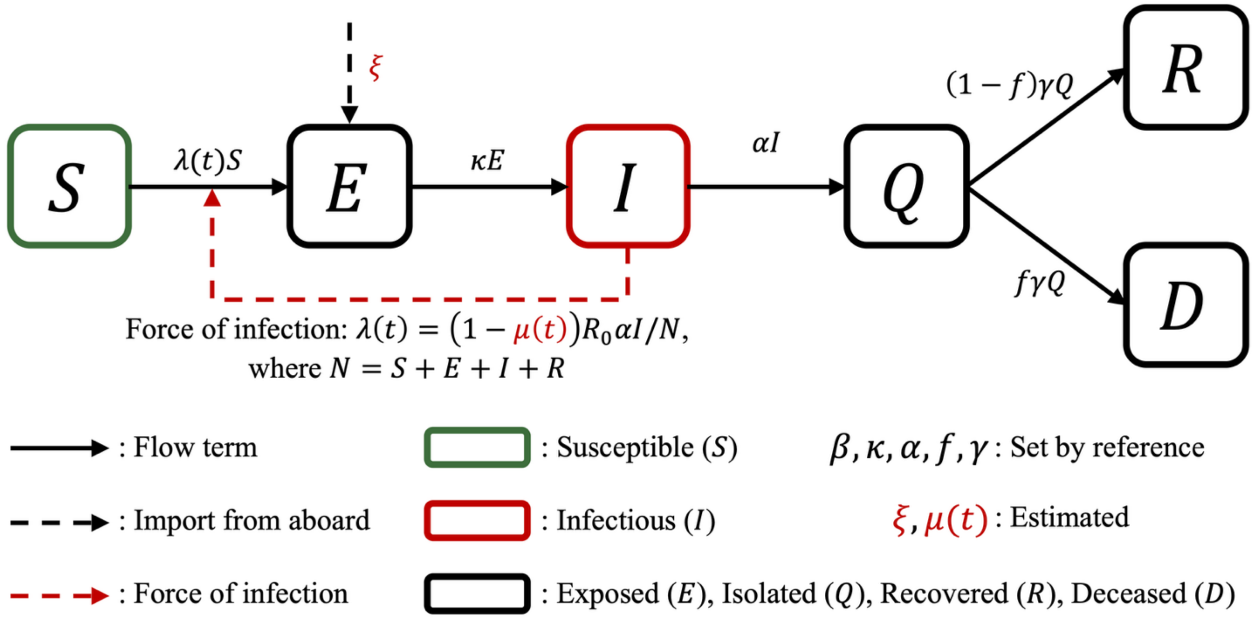


**Figure S1.3. Flowchart for the mathematical model.**

The parameters

$$\frac{dS}{dt} = -\lambda(t)S,$$
$$\frac{dE}{dt} = \lambda(t)S - \kappa E + \xi,$$
$$\frac{dI}{dt} = \kappa E - \alpha I,$$
$$\frac{dQ}{dt} = \alpha I - \gamma Q,$$
$$\frac{dR}{dt} = (1 - f)\gamma Q,$$
$$\frac{dD}{dt} = f\gamma Q.$$

**Table S1.2. Parameters for the mathematical model.** The four characteristics of the disease are obtained from the references.

| Parameters | Description | Value | references |
|---|---|---|---|
| $R_0$ | Basic reproduction number | 2.87 | [6] |
| $1/\kappa$ | Latent period | 4 days | [7] |
| $1/\alpha$ | Infectious period | 5.2 days | [8] |
| $1/\gamma$ | Recovery period | 14 days | [9,10] |
| $\xi$ | Imported latent people per day | Different by country | estimated |
| $f$ | Case fatality ratio | Different by country | calculated |
| $\mu$ | Transmission reduction over time | Different by country | estimated |

Identifiability refers to whether a parameter can be determined from a mathematical model and the associated data.[11] We conducted a structural identifiability analysis to determine whether the mathematical model can estimate states and parameters from given observable data and other information.[12,13] The results can be globally identifiable, locally identifiable, and nonidentifiable, meaning the parameter can be uniquely determined within the entire possible range, can be determined only within a neighborhood, or cannot be determined to a single value within a range, respectively. The states $E$, $I$, $Q$, and parameters $\kappa$, $\alpha$ are locally identifiable, and $R_0$, $\mu$ are nonidentifiable with the observable such as daily confirmed cases. Since we introduce the $\xi$ and assume there are no infected people at the beginning of the pandemic, $S$ is the population and the other states are zero i.e. we know all initial conditions. With known initial condition and observable, all the states and parameters are globally identifiable except for $R_0$ and $\mu$. Since we assumed there is a given basic reproduction number from an epidemiological reference, all the states and parameters can be uniquely determined from the observable.

**Table S1.3. Structural identifiability result with different conditions.**

| | Identifiability with observable | Identifiability with observable and initial condition | Identifiability with observable, initial condition, and known basic reproduction number |
|---|---|---|---|
| $S(t)$ | Globally | Globally | Globally |
| $E(t)$ | Locally | Globally | Globally |
| $I(t)$ | Locally | Globally | Globally |
| $Q(t)$ | Locally | Globally | Globally |
| $R(t)$ | Globally | Globally | Globally |
| $D(t)$ | Globally | Globally | Globally |
| $R_0$ | Nonidentifiable | Nonidentifiable | Globally |
| $\kappa$ | Locally | Globally | Globally |
| $\alpha$ | Locally | Globally | Globally |
| $\gamma$ | Globally | Globally | Globally |
| $\xi$ | Globally | Globally | Globally |
| $f$ | Globally | Globally | Globally |
| $\mu$ | Nonidentifiable | Nonidentifiable | Globally |

#### Supplementary 1C. Parameter estimation

To reduce computational time and improve accuracy, we used two algorithms - the global optimisation algorithm named improved multi-operator differential evolution (IMODE) and the Markov chain Monte Carlo (MCMC) method.[14-16] In our simulations, we used the cumulative confirmed case data and minimised the $L_2$-norm between the data and the corresponding output from the simulation results. We estimated the policy-related parameters $\xi$, $\mu_3$, $\mu_4$, $\cdots$, $\mu_K$ as described in the main text. IMODE identifies the optimal solution faster, whereas MCMC provides the solution with posterior information.

When using a local optimiser, it is often necessary to consider user-specified initial values because the solution can be sensitive to the starting point. However, employing a global optimiser removes this issue since it does not require an initial value for optimisation. To reduce the randomness of the global optimiser, IMODE was run 25 times, and the best solution was selected. Each simulation was terminated after 100,000 functional evaluations. Since the IMODE algorithm effectively searches the loss landscape for the most suitable solution, its results were used as prior information in the MCMC process.

The Markov chain Monte Carlo (MCMC) algorithm was implemented using MATLAB.[15,16] This package requires a parameter list, prior distribution of the parameters, system equations, and the objective function for the MCMC process. The delayed rejection adaptive metropolis (DRAM) algorithm was employed to explore the posterior distribution.[15] We set the prior distribution as a normal distribution with a mean value obtained from the IMODE results and a standard deviation of 0.05. We propose the joint posterior distribution of the estimated parameters

#### Supplementary 1D. Multi-objective optimisation

In multi-objective optimisation, Pareto solutions are defined strictly with respect to the feasible set, the subset of the decision space that satisfies all constraints (e.g., physical limits, budgets, policy rules), points that violate any constraint lie in the infeasible set and are excluded from consideration regardless of how attractive their objective values might appear. Mapping feasible decisions into objective space yields a cloud of feasible objective vectors; within this cloud, a decision is Pareto-efficient (nondominated) if there exists no other feasible decision that is at least as good in every objective and strictly better in at least one. The collection of all such nondominated objective vectors forms the Pareto frontier, which acts as the efficiency boundary of what is achievable under the given constraints. Conceptually, infeasible points are irrelevant to Pareto analysis because they are not implementable, whereas dominated feasible points are relevant but suboptimal, since a feasible alternative improves one or more objectives without worsening others. Tightening constraints contracts the feasible set and typically pushes the frontier inward (reducing attainable performance), while relaxing constraints expands the feasible set and can reveal new Pareto improvements.

Multi-objective optimisation identifies solutions near the Pareto curve, that is, a set of Pareto solutions in the objective plane composed of the co-domain of $g_1$ and $g_2$. A solution is considered Pareto optimal if it is not dominated by any other solution; that is, no other solution performs better in at least one objective without performing worse in another. We obtained the Pareto curve using the built-in function *multiobjga*, which employs the non-dominated sorting genetic algorithm II (NSGA-II) in MATLAB.[17] To ensure accuracy, we independently ran *multiobjga* one thousand times and assembled the Pareto solution by locating the Pareto front.

$$\min_{\mu(t)} \begin{bmatrix} g_1(\mu(t)) \\ g_2(\mu(t)) \end{bmatrix} = \min_{\mu(t)} \begin{bmatrix} \text{(Average transmission reduction)} \\ \text{(Number of infection)} \end{bmatrix}.$$

#### Supplementary 1E. Cost-benefit analysis

As the Pareto solutions represent outputs rather than costs, we introduce health-economic parameters to convert each objective into monetary cost. The total societal cost is $C_{tot} = C_{int} + C_{inf}$ , where the two components are defined below. Intervention cost (USD, over the simulation horizon): $C_{int}(\mu(t)) = C_1 \times g_1(\mu(t))$, where $C_1 = GDP_{total} \times GDP_{MaxRed}$ (USD/day) is the maximum daily intervention cost. $GDP_{total}$ is the country's total GDP (USD, not per capita) and $GDP_{MaxRed} = 0.10$ (10% of daily GDP assumed lost under full intervention). Infection cost (USD, over the simulation horizon): $C_{inf}(\mu(t)) = C_2 \times g_2(\mu(t))$, where $C_2 = C_H + f \times VSL$ (USD/infection) is the cost per infected individual. $C_H = 0.02 \times \text{GDP}_{\text{per}}$ is the average quarantine or hospitalisation cost, $f$ is the country-specific case fatality ratio (dimensionless), and VSL = $\text{GDP}_{\text{per}}$ ×54.8966 is the value of a statistical life[18-22]. The cost $C_2$ is applied to every simulated infection ($g_2$), not to confirmed cases or quarantined individuals.

**Supplementary 1F. Correlation between the country-specific indices and epidemic-related outcomes**

This section reports how the 100 country-level indices used in the study relate to one another and to the 60 epidemic and cost outcomes of the cross-country simulations, across all 110 countries. It is organised around the three quantities that the figures display, the correlation value, its Benjamini-Hochberg false-discovery-rate q-value, and the redundancy index, each defined in turn below. All correlations are computed pairwise-complete: a country contributes to a pair only where both quantities are observed (blank cells are treated as missing), and any pair supported by fewer than 30 common countries is excluded.

**Correlation value**

For two country-level variables *x* and *y* measured on the *n* countries where both are observed, the Pearson correlation coefficient is

$$r_{xy} = \frac{\sum_i (x_i - \bar{x})(y_i - \bar{y})}{\sqrt{\sum_i (x_i - \bar{x})^2 \cdot \sum_i (y_i - \bar{y})^2}}$$

where $\bar{x}$ and $\bar{y}$ are the means over those *n* countries. It ranges from $r = -1$ (the two move in exactly opposite directions) through 0 (no linear relationship) to +1 (they move together), and is mapped onto a red-white-blue colour scale in Figures S1.4 and S1.5. The redundancy ranking (third subsection) instead uses the Spearman rank correlation $r^s$, the same formula applied after replacing each value by its within-sample rank, which captures any monotone (not only linear) association and resists the skew of national indices. Any pair supported by fewer than 30 common countries ($n < 30$) is treated as missing and excluded. Each coefficient's significance is obtained from the statistic

$$t = r\sqrt{\frac{n - 2}{1 - r^2}}$$

which follows a Student *t* distribution with $n - 2$ degrees of freedom and gives a two-sided *p*-value; here *n* is the number of countries with both variables observed, and this *p*-value feeds the *q*-value defined in the next subsection.

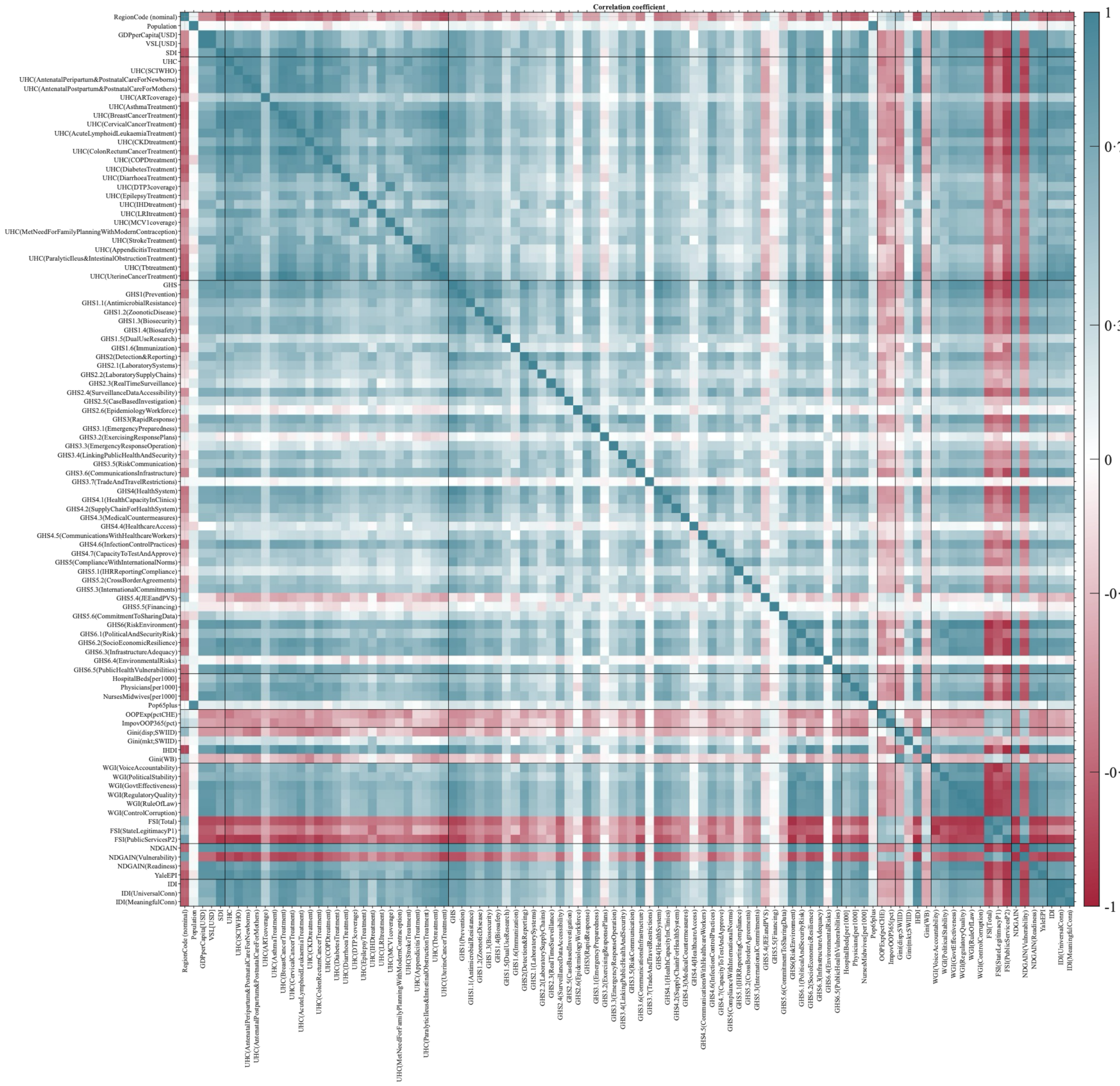


**Figure S1.4: Correlation coefficient between every pair of the 100 country indices, across the 110 countries.** Each cell is the Pearson correlation r for one pair of indices, coloured from red (r = −1, the indices move in opposite directions) through white (r = 0, unrelated) to blue (r = +1, they move together); the matrix is symmetric, with a blue diagonal where each index matches itself. Black lines separate the nine thematic index groups. The strong blue blocks along the diagonal show that indices within a theme, especially the universal-health-coverage service items and the Global Health Security sub-scores, largely repeat one another (82% of the 4950 pairs are significant at q < 0.05; Figure S1.6). This heavy overlap is what the redundancy index of Figure S1.8 is designed to accommodate.

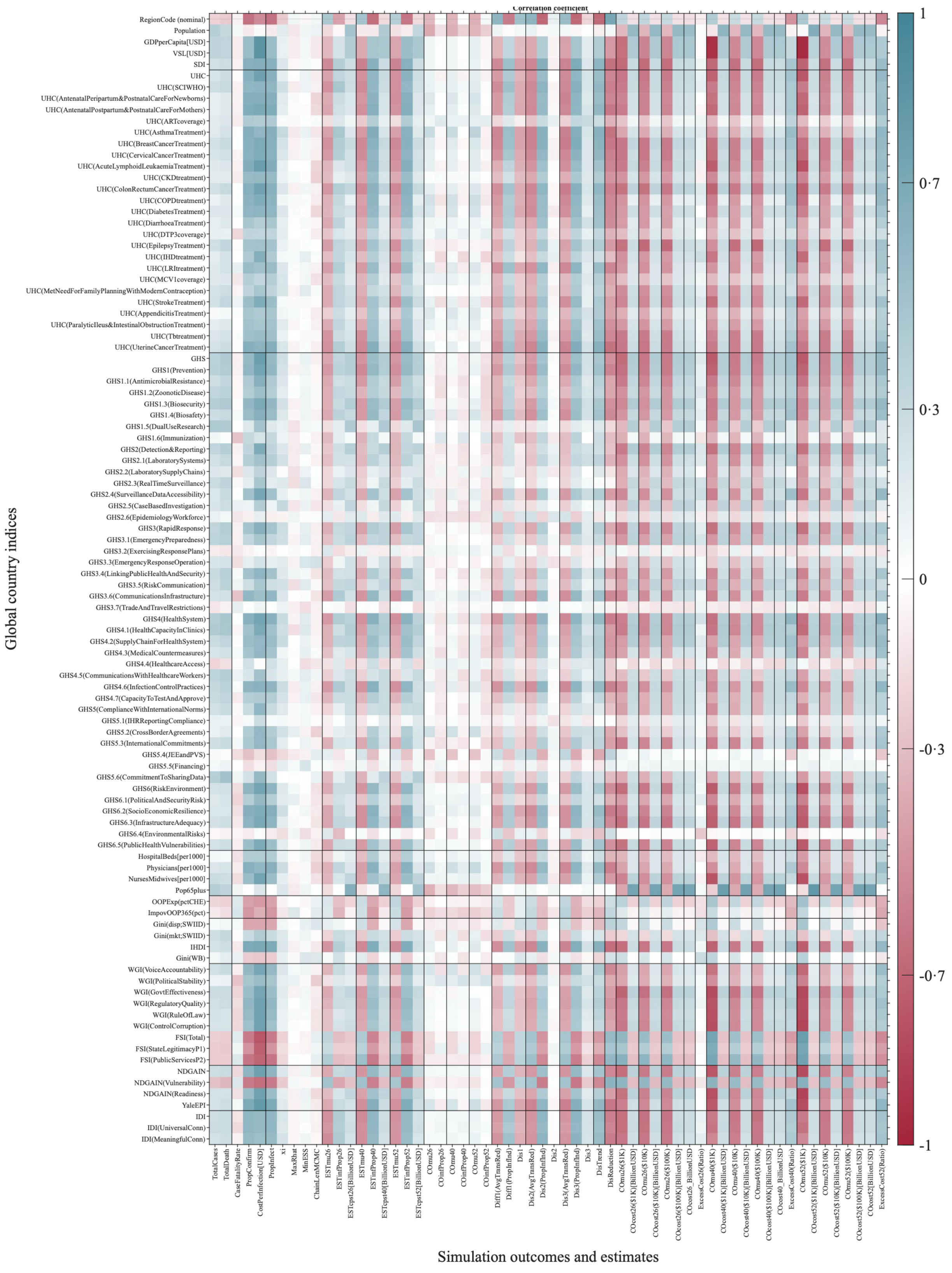


**Figure S1.5: Correlation coefficient between the 100 country indices (rows) and the 60 simulation outcomes (columns), across the 110 countries.** Colour encodes the Pearson correlation r on the same red-white-blue scale as

Figure S1.4 (red = negative, white = none, blue = positive). Rows are grouped by index theme and columns by outcome type (fitted epidemic quantities, and the cost-optimal transmission reductions and their costs at several cost-per-infection levels and time horizons). These associations are weaker and more selective than those between indices: 63% of the 6000 index-outcome pairs are significant ($q < 0.05$; Figure S1.7) but the median absolute correlation is only 0.27. The strongest single links involve general development and health-capacity indices; for example the socio-demographic index with total cases ($r \approx 0.69$), the population aged 65 and over with total deaths (0.66), physicians per 1000 people with the infected proportion (0.78), and GDP per capita with the cost per infection (0.88).

#### Benjamini-Hochberg FDR q-value

Each heat map evaluates many correlations at once, $m = N(N-1)/2 = 4950$ unique pairs among the $N = 100$ indices (Figure S1.6), and $m = N \times M = 100 \times 60 = 6000$ index-outcome pairs (Figure S1.7), so some would appear significant by chance alone. The Benjamini-Hochberg false-discovery-rate procedure controls this. Ordering the $m$ two-sided $p$-values as $p_{(1)} \le p_{(2)} \le \cdots \le p_{(m)}$, the $q$-value of the $i$-th is

$$q_{(i)} = \min_{k \ge i} \left\{ \min\left(1, \frac{m}{k}\, p_{(k)}\right) \right\}$$

and a correlation is treated as reliable when $q < 0.05$. Equivalently, the $q$-value is the smallest false-discovery rate at which that correlation would be declared significant. For the symmetric index-index matrix the correction is taken over the $N(N-1)/2$ unique off-diagonal pairs; for the rectangular index-outcome matrix, over all $N \times M$ evaluated cells.

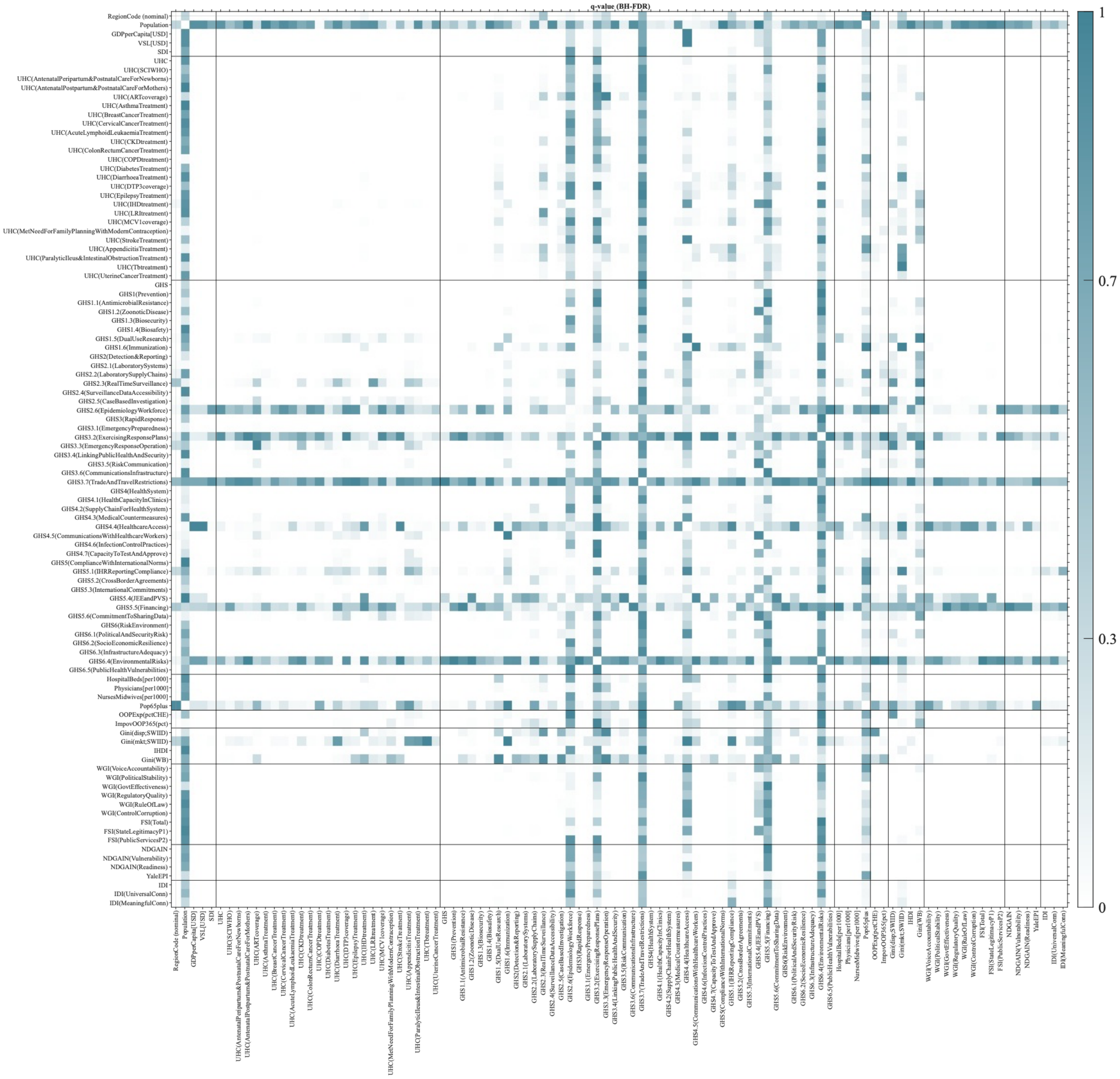


**Figure S1.6: Statistical significance of the index-index correlations of Figure S1.4.** Each cell is the Benjamini-Hochberg false-discovery-rate q-value for one pair of indices, on a scale from white (q = 0, strongly significant) to blue (q = 1, not significant); a pair is treated as reliable when q < 0.05 (the pale cells). Most cells are pale, confirming that the great majority of index pairs are significantly correlated; the blue cells mark the minority that are not, mainly pairs involving the region code and total population.

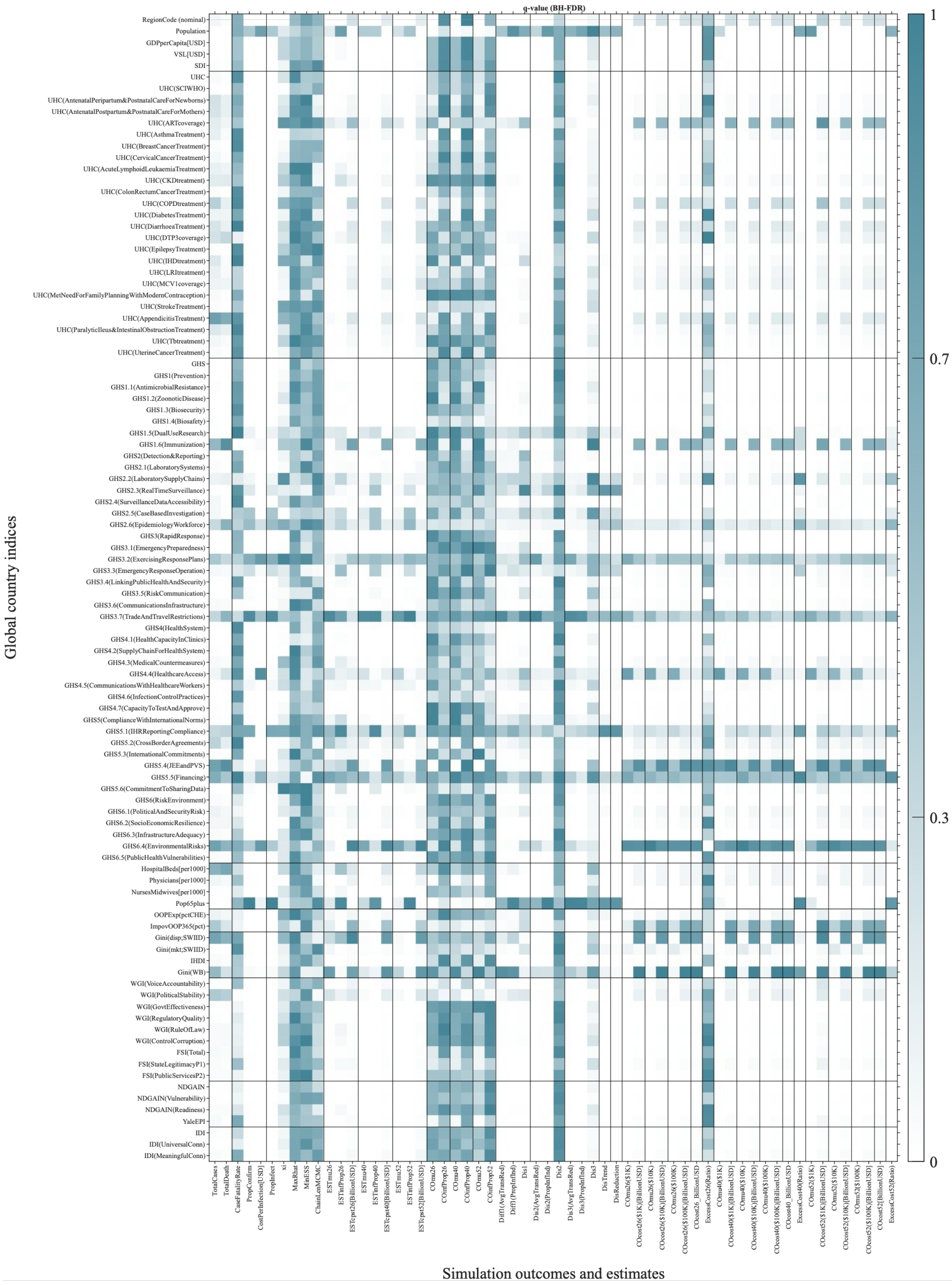
q-value (BH-FDR)
Global country indices
Simulation outcomes and estimates
1
0.7
0.3
0
RegionCode (nominal)
Population
GDPperCapita[USD]
VSL[USD]
SDI
UHC
UHC(SCIWHO)
UHC(AntenatalPeripartum&PostnatalCareForNewborns)
UHC(AntenatalPostpartum&PostnatalCareForMothers)
UHC(ARTcoverage)
UHC(AsthmaTreatment)
UHC(BreastCancerTreatment)
UHC(CervicalCancerTreatment)
UHC(AcuteLymphoidLeukaemiaTreatment)
UHC(CKDtreatment)
UHC(ColonRectumCancerTreatment)
UHC(COPDtreatment)
UHC(DiabetesTreatment)
UHC(DiarrhoeaTreatment)
UHC(DTP3coverage)
UHC(EpilepsyTreatment)
UHC(IHDtreatment)
UHC(LRItreatment)
UHC(MCV1coverage)
UHC(MetNeedForFamilyPlanningWithModernContraception)
UHC(StrokeTreatment)
UHC(AppendicitisTreatment)
UHC(ParalyticIleus&IntestinalObstructionTreatment)
UHC(Tbtreatment)
UHC(UterineCancerTreatment)
GHS
GHS1(Prevention)
GHS1.1(AntimicrobialResistance)
GHS1.2(ZoonoticDisease)
GHS1.3(Biosecurity)
GHS1.4(Biosafety)
GHS1.5(DualUseResearch)
GHS1.6(Immunization)
GHS2(Detection&Reporting)
GHS2.1(LaboratorySystems)
GHS2.2(LaboratorySupplyChains)
GHS2.3(RealTimeSurveillance)
GHS2.4(SurveillanceDataAccessibility)
GHS2.5(CaseBasedInvestigation)
GHS2.6(EpidemiologyWorkforce)
GHS3(RapidResponse)
GHS3.1(EmergencyPreparedness)
GHS3.2(ExercisingResponsePlans)
GHS3.3(EmergencyResponseOperation)
GHS3.4(LinkingPublicHealthAndSecurity)
GHS3.5(RiskCommunication)
GHS3.6(CommunicationsInfrastructure)
GHS3.7(TradeAndTravelRestrictions)
GHS4(HealthSystem)
GHS4.1(HealthCapacityInClinics)
GHS4.2(SupplyChainForHealthSystem)
GHS4.3(MedicalCountermeasures)
GHS4.4(HealthcareAccess)
GHS4.5(CommunicationsWithHealthcareWorkers)
GHS4.6(InfectionControlPractices)
GHS4.7(CapacityToTestAndApprove)
GHS5(ComplianceWithInternationalNorms)
GHS5.1(IHRReportingCompliance)
GHS5.2(CrossBorderAgreements)
GHS5.3(InternationalCommitments)
GHS5.4(JEEandPVS)
GHS5.5(Financing)
GHS5.6(CommitmentToSharingData)
GHS6(RiskEnvironment)
GHS6.1(PoliticalAndSecurityRisk)
GHS6.2(SocioEconomicResilience)
GHS6.3(InfrastructureAdequacy)
GHS6.4(EnvironmentalRisks)
GHS6.5(PublicHealthVulnerabilities)
HospitalBeds[per1000]
Physicians[per1000]
NursesMidwives[per1000]
Pop65plus
OOPExp(pctCHE)
ImpovOOP365(pct)
Gini(disp;SWIID)
Gini(mkt;SWIID)
IHDI
Gini(WB)
WGI(VoiceAccountability)
WGI(PoliticalStability)
WGI(GovtEffectiveness)
WGI(RegulatoryQuality)
WGI(RuleOfLaw)
WGI(ControlCorruption)
FSI(Total)
FSI(StateLegitimacyP1)
FSI(PublicServicesP2)
NDGAIN
NDGAIN(Vulnerability)
NDGAIN(Readiness)
YaleEPI
IDI
IDI(UniversalConn)
IDI(MeaningfulConn)
TotalCases
TotalDeath
CaseFatalityRate
PropConfirm
CostPerInfection[USD]
PropInfect
xi
MaxRhat
MinESS
ChainLenMCMC
ESTmu26
ESTinfProp26
ESTcpst26[BillionUSD]
ESTmu40
ESTinfProp40
ESTcpst40[BillionUSD]
ESTmu52
ESTinfProp52
ESTcpst52[BillionUSD]
COmu26
COinfProp26
COmu40
COinfProp40
COmu52
COinfProp52
Diff1(AvgTransRed)
Diff1(PropInfInd)
Dis1
Dis2(AvgTransRed)
Dis2(PropInfInd)
Dis2
Dis3(AvgTransRed)
Dis3(PropInfInd)
Dis3
DisTrend
DisReduction
COmu26($1K)
COcost26($1K)[BillionUSD]
COmu26($10K)
COcost26($10K)[BillionUSD]
COmu26($100K)
COcost26($100K)[BillionUSD]
COcost26_BillionUSD
ExcessCost26(Ratio)
COmu40($1K)
COcost40($1K)[BillionUSD]
COmu40($10K)
COcost40($10K)[BillionUSD]
COmu40($100K)
COcost40($100K)[BillionUSD]
COcost40_BillionUSD
ExcessCost40(Ratio)
COmu52($1K)
COcost52($1K)[BillionUSD]
COmu52($10K)
COcost52($10K)[BillionUSD]
COmu52($100K)
COcost52($100K)[BillionUSD]
COcost52[BillionUSD]
ExcessCost52(Ratio)

**Figure S1.7: Statistical significance of the index-outcome correlations of Figure S1.5.** Each cell is the Benjamini-Hochberg false-discovery-rate q-value for one index-outcome pair, on the same white (q = 0) to blue (q = 1) scale, with q < 0.05 (pale cells) treated as reliable. The significant (pale) cells cluster in the development, demographic and healthcare-capacity index rows rather than spreading evenly, indicating that the predictability of the epidemic outcomes is concentrated in broad markers of national development and health-system capacity rather than in the purpose-built preparedness scores.

#### Redundancy index

Because the indices overlap heavily (Figure S1.4), counting significant index-outcome associations would overstate how much independent information they carry. To summarise how strongly a single index $x$ is associated with the whole block of $p = 60$ outcomes $y_1, \cdots, y_p$, the redundancy index is the mean squared Spearman correlation

$$R_d(x) = \frac{1}{p} \sum_{j=1}^{p} \left( r^s_{x,y_j} \right)^2$$

so the redundancy index ($R_d(x)$) lies between 0 (no association with any outcome) and 1 (association with all of them), the rank-based, multi-outcome analogue of a coefficient of determination; squaring makes positive and negative associations count alike. The thematic-group score in the lower panel of Figure S1.8 is the unweighted mean over a group's member indices,

$$\bar{R}_d(g) = \frac{1}{|g|} \sum_{x \in g} R_d(x)$$

Uncertainty is quantified by a country-level bootstrap: for each of $B = 2000$ resamples, the $n$ countries are drawn with replacement and $R_d$ is recomputed, giving the percentile 95% interval $[Q_{2.5}, Q_{97.5}]$ drawn on each index in Figure S1.8. An index is counted as tied with the top-ranked index when

$$Q_{97.5}(x) \geq Q_{2.5}(x_{(1)})$$

that is, when its bootstrap interval reaches the lower confidence bound of the rank-1 index; 42 of the 100 indices meet this criterion (marked by the dashed line in Figure S1.8).

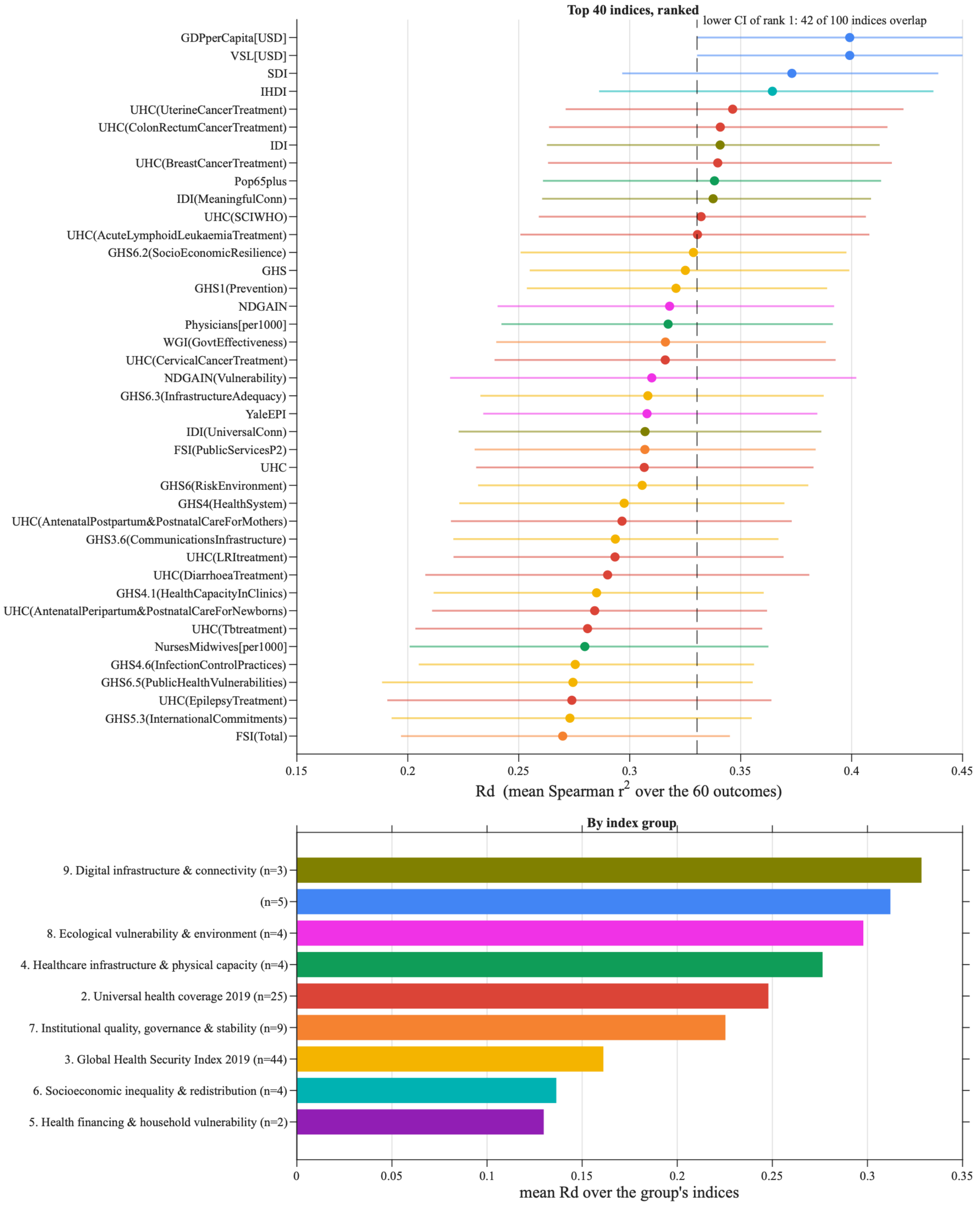


**Figure S1.8: Ranking of the 100 country indices by their overall association with the epidemic outcomes, measured by the redundancy index Rd.** Upper panel: the 40 highest-ranked indices, each a point at its Rd and ordered from strongest (top) to weakest, with a horizontal bar giving the 95% bootstrap confidence interval (2000 country-level resamples); colour marks the thematic group. The dashed vertical line marks the lower confidence bound of the top-ranked index (GDP per capita); the 42 indices whose bars reach past it are statistically indistinguishable from the top, so no single index is the clear best. Lower panel: the nine thematic groups ranked by their average Rd, with the number of member indices in parentheses, digital-infrastructure and broad development indices rank highest, the health-financing and Global Health Security preparedness groups lowest. Decimals use the middle dot.

Taken together, the indices are highly redundant (median absolute index-index correlation 0.48, with some pairs almost identical) while their individual links to the epidemic outcomes are weaker (median 0.27). Broad measures of national development and wealth, GDP per capita, the value of a statistical life, the socio-demographic index and the inequality-adjusted human development index, hold the highest redundancy ranks, whereas the Global Health Security Index preparedness sub-scores and health-financing indicators rank lowest. Because so many indices overlap, no single index is the best predictor: 42 of the 100 are statistically tied with the top-ranked one.

#### Supplementary 1G. Characteristics of Figure 1

We analyse the characteristics of the estimated points and the circumscribed ellipse about them. As the simulation period is longer, the transmission reduction and confirmed cases are larger. The variance of confirmed cases is also larger, but the variance of transmission reduction is getting smaller. In the normalized objective space with a logarithmic y-axis, we make a circumscribed ellipse about the estimated points. The ellipse is getting flatter as the simulation period is increased.

**Table S1.4. The characteristic of estimated points for 110 countries and a circumscribed ellipse about them.**

| | **Simulation period** | **26w** | **40w** | **52w** |
|---|---|---|---|---|
| Estimated points for 110 countries | Median of x-value | 0.5111 | 0.5441 | 0.5628 |
| | Median of y-value | 0.0033 | 0.0111 | 0.0203 |
| | Std of x-value | 0.0588 | 0.0489 | 0.0452 |
| | Std of y-value | 0.0045 | 0.0134 | 0.0265 |
| Ellipse | Semi-major axis | 3.06 | 1.99 | 1.59 |
| | Semi-minor axis | 1 | 1 | 1 |
| | Rotated angle | 12 | 15 | 20 |
| | Eccentricity | 0.945 | 0.865 | 0.777 |
| | Flattening | 0.673 | 0.497 | 0.371 |
| | x-value of center | 0.53 | 0.55 | 0.58 |
| | y-value of center | 5.1e-4 | 8.2e-4 | 1.5e-3 |

#### Supplementary 1H. Categorisation of the points

We use six criteria to categorise the 110 countries. Region is divided into six groups based on Global Burden of Disease (GBD) super-regions: Central Europe, Eastern Europe, and Central Asia; High income; Latin America and Caribbean; North Africa and Middle East; Southeast Asia, East Asia, and Oceania; and Sub-Saharan Africa. Population is divided into four groups: more than 100M, 30M-100M, 10M-30M, and 5M-10M. Income divides the countries into four groups: more than $17K, $7K-$17K, $2K-$7K, and less than $2K. UHC, SDI, and GHS each divide the countries into four quartiles: more than 72, 59-72, 48-59, and less than 48; more than 0.76, 0.64-0.76, 0.46-0.64, and less than 0.46; and more than 50.7, 37.7-50.7, 31-37.7, and less than 31, respectively.

A higher silhouette score is better, but here it is uninformative because all scores are close to zero. SDI performs best on the Calinski-Harabasz score, which is the highest among the six criteria, and Income performs best on the Davies-Bouldin score, which is the lowest among the six criteria. The variance is computed as the distance from the estimated points to their centre. A higher variance within a group defined by a given criterion indicates that countries in that group had more heterogeneous data-fitted responses and confirmed-case outcomes. North Africa and Middle East have lowest variance, and Southeast Asia, East Asia, and Oceania have highest variance with respect to the region criteria. Population between 10M and 30M have the lowest variance, and population between 5M and 10M have the highest variance. For the Income, UHC, SDI, and GHS, G4 have the lowest variance

**Table S1.5. Three evaluation scores and the variance for six criteria.**

| Criteria | Evaluation scores | | | Variance | | | | | |
|---|---|---|---|---|---|---|---|---|---|
| | Silhouette (↑) | Calinski-Harabasz (↑) | Davies-Bouldin (↓) | G1* | G2* | G3* | G4* | G5* | G6* |
| Region | -0.0695 | 10.9863 | 5.5254 | 0.116 | 0.188 | 0.127 | 0.080 | 0.371 | 0.287 |
| Population | -0.0425 | 1.7782 | 5.3592 | 0.372 | 0.384 | 0.185 | 0.644 | | |
| Income | 0.0042 | 24.4024 | 2.4783 | 0.262 | 0.531 | 0.221 | 0.108 | | |
| UHC§ | -0.0277 | 18.1906 | 2.8652 | 0.263 | 0.436 | 0.339 | 0.108 | | |
| SDI† | -0.0016 | 24.5844 | 2.6443 | 0.269 | 0.453 | 0.205 | 0.109 | | |
| GHS‡ | -0.0287 | 14.3744 | 4.2772 | 0.171 | 0.645 | 0.343 | 0.164 | | |

***Region**: G1:Central Europe, Eastern Europe, and Central Asia; G2:High income; G3:Latin America and Caribbean; G4:North Africa and Middle East; G5: Southeast Asia, East Asia, and Oceania; G6:Sub-Saharan Africa.
**Population:**G1:more than 100M, G2:30M-100M, G3:10M-30M, G4:5M-10M.
**Income**: G1:more than $17K, G2:$7K-$17K, G3:$2K-$7K, G4:less than $2K.
**UHC**: G1:more than 72, G2:59.3-72, G3:47.9-59.3, G4:less than 47.9.
**SDI**: G1:more than 0.76, G2:0.64-0.76, G3:0.46-0.64, G4:less than 0.46.
**GHS**: G1:more than 50.7, G2:37.7-50.7, G3:31-37.7, G4:less than 31.
§Universal Health Coverage
†Socio-demographic index
‡Global health security index

#### Supplementary 1I. Difference trends

The table evaluates the difference between optimal and estimated trajectories across 110 countries over three distinct time periods (26, 40, and 52 weeks). Globally, the data reveals a balanced distribution of policy alignment and divergence. Specifically, 40 countries exhibited a decreasing trend (D) in the distance between their actual data-fitted outcomes and cost-optimal solutions, indicating that their pandemic interventions became increasingly optimal over time. Conversely, 37 countries showed an increasing trend (I), suggesting a growing divergence from optimal strategies as the pandemic progressed. The remaining 33 countries demonstrated a random (R) or fluctuating pattern, lacking a clear directional trajectory.

**Table S1.6. Difference trends between cost-optimal and estimated trajectories across 110 countries**. This table details the numerical difference between theoretical cost-optimal interventions and actual data-fitted (estimated) outcomes for each country across three vaccine development timelines: 26, 40, and 52 weeks. Based on the progression of these differences over time, each country's trajectory is classified into one of three overall trends: Decrease (D, indicating a narrowing gap and alignment with optimal strategies), Increase (I, indicating growing divergence from the optimal path), or Random (R, indicating fluctuating alignment).

| | Difference between optimal and estimated | | | | | Difference between optimal and estimated | | | |
|---|---|---|---|---|---|---|---|---|---|
| Country | Trend† | 26w‡ | 40w‡ | 52w‡ | Country | Trend† | 26w‡ | 40w‡ | 52w‡ |
| Afghanistan | Random | 1.139 | 0.675 | 0.878 | Laos | Decreasing | 5.457 | 4.508 | 4.274 |
| Algeria | Decreasing | 0.856 | 0.688 | 0.686 | Lebanon | Increasing | 0.817 | 1.097 | 1.398 |
| Angola | Decreasing | 1.370 | 0.980 | 0.944 | Liberia | Random | 1.475 | 1.052 | 1.087 |
| Argentina | Increasing | 1.162 | 1.484 | 1.723 | Libya | Increasing | 0.819 | 1.041 | 1.098 |
| Australia | Random | 0.704 | 0.956 | 0.579 | Madagascar | Random | 1.088 | 1.184 | 0.832 |
| Austria | Increasing | 0.822 | 1.519 | 1.548 | Malawi | Decreasing | 1.666 | 0.783 | 0.709 |
| Azerbaijan | Increasing | 0.909 | 1.055 | 1.231 | Malaysia | Decreasing | 1.921 | 0.973 | 0.598 |
| Bangladesh | Decreasing | 1.026 | 0.874 | 0.823 | Mali | Decreasing | 1.462 | 0.842 | 0.773 |
| Belarus | Random | 1.022 | 0.790 | 1.010 | Mexico | Increasing | 1.331 | 1.442 | 1.876 |
| Belgium | Increasing | 0.973 | 1.579 | 1.633 | Morocco | Random | 0.823 | 1.138 | 1.110 |
| Benin | Decreasing | 1.976 | 1.449 | 0.996 | Mozambique | Decreasing | 1.340 | 0.957 | 0.697 |
| Bolivia | Random | 1.401 | 1.400 | 1.470 | Myanmar | Random | 1.075 | 0.963 | 1.299 |
| Brazil | Increasing | 1.297 | 1.679 | 1.750 | Nepal | Random | 1.151 | 0.888 | 1.069 |
| Bulgaria | Increasing | 0.580 | 1.294 | 1.459 | Netherlands | Increasing | 0.573 | 1.274 | 1.455 |
| Burkina Faso | Decreasing | 2.088 | 1.345 | 1.260 | Niger | Decreasing | 2.785 | 1.792 | 1.633 |
| Burundi | Decreasing | 2.932 | 2.070 | 1.669 | Nigeria | Decreasing | 1.645 | 1.323 | 0.802 |
| Cambodia | Decreasing | 3.255 | 2.894 | 2.647 | Pakistan | Random | 1.240 | 0.787 | 0.805 |
| Cameroon | Decreasing | 1.162 | 0.969 | 0.477 | Paraguay | Increasing | 0.617 | 0.945 | 1.246 |
| Canada | Random | 1.061 | 0.902 | 1.227 | Peru | Increasing | 1.820 | 1.966 | 2.004 |
| Chad | Decreasing | 1.638 | 1.248 | 0.790 | Philippines | Random | 0.807 | 0.913 | 0.859 |
| Chile | Random | 1.364 | 1.324 | 1.484 | Poland | Random | 0.953 | 1.690 | 1.674 |
| China | Decreasing | 2.326 | 1.772 | 1.187 | Portugal | Increasing | 0.783 | 1.307 | 1.572 |
| Colombia | Random | 1.358 | 1.331 | 1.764 | Romania | Increasing | 0.820 | 1.274 | 1.397 |
| Costa Rica | Increasing | 0.783 | 1.180 | 1.195 | Russia | Increasing | 0.943 | 1.283 | 1.380 |
| Cote d'Ivoire | Decreasing | 1.551 | 1.285 | 0.735 | Rwanda | Decreasing | 1.459 | 1.073 | 0.717 |
| Cuba | Decreasing | 1.544 | 1.173 | 0.503 | Saudi Arabia | Random | 1.141 | 1.218 | 1.023 |
| Czechia | Increasing | 0.712 | 1.463 | 1.865 | Senegal | Decreasing | 1.031 | 0.890 | 0.581 |
| Democratic Republic of Congo | Decreasing | 1.854 | 1.318 | 1.157 | Serbia | Increasing | 0.802 | 1.226 | 1.455 |
| Denmark | Increasing | 0.773 | 0.891 | 1.245 | Sierra Leone | Random | 1.555 | 1.180 | 1.226 |
| Dominican Republic | Random | 0.861 | 0.811 | 0.943 | Slovakia | Increasing | 0.496 | 1.206 | 1.547 |
| Ecuador | Increasing | 1.110 | 1.127 | 1.339 | Somalia | Decreasing | 1.581 | 1.320 | 0.664 |
| Egypt | Random | 1.113 | 0.873 | 0.878 | South Africa | Random | 1.353 | 1.333 | 1.423 |
| El Salvador | Random | 0.808 | 0.792 | 0.812 | Republic of Korea | Decreasing | 1.211 | 0.949 | 0.471 |
| Ethiopia | Decreasing | 1.319 | 1.126 | 0.674 | South Sudan | Decreasing | 1.778 | 1.398 | 0.964 |
| Finland | Random | 1.191 | 0.552 | 0.706 | Spain | Increasing | 1.040 | 1.458 | 1.681 |
| France | Increasing | 0.796 | 1.498 | 1.759 | Sri Lanka | Decreasing | 2.217 | 0.925 | 0.768 |
| Germany | Random | 1.142 | 0.809 | 1.379 | Sudan | Random | 1.362 | 0.694 | 0.802 |
| Ghana | Decreasing | 1.248 | 0.994 | 0.620 | Syria | Decreasing | 1.015 | 0.733 | 0.625 |
| Greece | Increasing | 0.544 | 1.078 | 1.191 | Tajikistan | Increasing | 1.139 | 1.291 | 1.496 |
| Guatemala | Random | 0.983 | 1.184 | 1.079 | Tanzania | Decreasing | 3.891 | 3.876 | 3.747 |
| Guinea | Decreasing | 1.379 | 1.220 | 0.872 | Thailand | Decreasing | 2.494 | 2.014 | 1.169 |

| | | | | | | | | | |
|---|---|---|---|---|---|---|---|---|---|
| Haiti | Random | 1.196 | 0.826 | 1.015 | Togo | Decreasing | 1.689 | 1.244 | 0.772 |
| Honduras | Increasing | 0.831 | 0.915 | 1.060 | Tunisia | Increasing | 0.711 | 0.967 | 1.241 |
| Hungary | Increasing | 0.514 | 1.574 | 1.693 | Turkey | Increasing | 0.740 | 1.321 | 1.379 |
| India | Random | 0.853 | 1.143 | 1.113 | Uganda | Random | 1.680 | 1.134 | 1.269 |
| Indonesia | Random | 0.876 | 0.842 | 0.955 | Ukraine | Increasing | 0.889 | 1.339 | 1.473 |
| Iran | Increasing | 1.011 | 1.217 | 1.293 | United Arab Emirates | Random | 0.971 | 0.858 | 0.950 |
| Iraq | Increasing | 0.976 | 1.159 | 1.207 | United Kingdom | Increasing | 1.002 | 1.374 | 1.820 |
| Israel | Increasing | 0.955 | 1.341 | 1.662 | United States | Increasing | 1.131 | 1.386 | 1.981 |
| Italy | Increasing | 1.014 | 1.545 | 1.780 | Uzbekistan | Decreasing | 1.911 | 1.057 | 0.964 |
| Japan | Decreasing | 1.229 | 0.780 | 0.690 | Venezuela | Random | 0.854 | 0.954 | 0.761 |
| Jordan | Increasing | 1.213 | 1.387 | 1.548 | Vietnam | Decreasing | 2.693 | 2.200 | 2.050 |
| Kazakhstan | Decreasing | 1.177 | 0.971 | 0.969 | Yemen | Decreasing | 1.830 | 1.672 | 0.765 |
| Kenya | Decreasing | 1.318 | 0.930 | 0.705 | Zambia | Decreasing | 1.231 | 0.788 | 0.674 |
| Kyrgyzstan | Random | 1.003 | 0.795 | 0.944 | Zimbabwe | Random | 1.238 | 0.590 | 0.671 |

†Decreasing: distance between cost-optimal and estimated decreases over time. Random: no clear monotonic trend. Increasing: distance between cost-optimal and estimated increases over time.
‡Euclidean distance between the cost-optimal and data-fitted points in the normalised objective space

No significant relationship was observed between the categorisation criteria and the average trend in difference over time. The average trends are near zero in most categories. The G4 group in the Region category (North Africa and Middle East) has the highest average trend value, indicating that the distance between the cost-optimal solution and the implemented policy increased over time in this group.

**Table S1.7. The average trends in difference over time for 110 countries.** The average calculated by changing the decreasing trend to -1, random trend to 0, and increasing trend to 1.

| Criteria | Average trend (-1: decreasing, 0: random, 1: increasing) | | | | | |
|---|---|---|---|---|---|---|
| | **G1*** | **G2*** | **G3*** | **G4*** | **G5*** | **G6*** |
| Region | -0.0667 | -0.1111 | -0.1765 | 0.25 | 0.1333 | -0.1333 |
| Population | -0.125 | -0.0571 | 0.075 | -0.1053 | | |
| Income | -0.1364 | 0.0952 | 0.0625 | -0.1143 | | |
| UHC§ | -0.1071 | 0 | 0.1852 | -0.1786 | | |
| SDI† | -0.1071 | 0 | 0.1852 | -0.1786 | | |
| GHS‡ | -0.1071 | 0 | 0.1852 | -0.1786 | | |

***Region**:G1:Central Europe, Eastern Europe, and Central Asia; G2:High income; G3:Latin America and Caribbean; G4:North Africa and Middle East; G5: Southeast Asia, East Asia, and Oceania; G6:Sub-Saharan Africa.
**Population**: G1:more than 100M, G2:30M-100M, G3:10M-30M, G4:5M-10M.
**Income**: G1:more than $17K, G2:$7K-$17K, G3:$2K-$7K, G4:less than $2K.
**UHC**: G1:more than 72, G2:59.3-72, G3:47.9-59.3, G4:less than 47.9.
**SDI**: G1:more than 0.76, G2:0.64-0.76, G3:0.46-0.64, G4:less than 0.46.
**GHS**: G1:more than 50.7, G2:37.7-50.7, G3:31-37.7, G4:less than 31.
§Universal Health Coverage
†Socio-demographic index
‡Global health security index

**Supplementary 1J. Cost-optimal solutions under various cost per infection**

The data show a clear relationship between the assumed cost per infection and the intensity of the cost-optimal intervention. As the cost per infection increases, the model favours more stringent transmission reduction to minimise the total societal burden. Moving from $1K to $100K per infection raises the cost-optimal transmission reduction across countries. For example, high-income countries such as Australia have a cost-optimal transmission reduction of 0.1667 at $1K, rising to 0.4906 at $10K and 0.5196 at $100K. Afghanistan's cost-optimal reduction rises from 0.5153 at $1K to 0.6475 at $100K. This indicates that a higher valuation of life or greater disease severity shifts the cost-optimal strategy toward more stringent containment.

A direct comparison between the Estimated result and the Default cost-optimal solution highlights an optimality gap in real-world pandemic management, revealing that actual implementation often results in total costs that exceed theoretical minimums.

$$Excess\ cost\ =\ (estimated\ total\ cost\ /\ cost\ optimal\ total\ cost)\ -1.$$

The average gap is 65.5%; specifically, 67 countries exhibited a gap of less than 50%, while 27 countries showed a gap exceeding 100%. Even when countries implement transmission reductions close to their default optimal levels, the actual economic burden is frequently disproportionately higher. For instance, Brazil's estimated total cost reached 254.222 billion USD, which is more than 2.5 times its default theoretical optimal cost of 98.539 billion USD. Likewise, Argentina incurred an estimated total cost of 63.014 billion USD compared to an optimal target of 24.375 billion USD, underscoring how slight deviations in policy timing or efficacy can trigger compounding economic losses in reality.

**Table S1.8. Cost-optimal solutions and total economic burden under varying costs per infection.** This supplementary table compares the estimated actual outcomes with theoretical cost-optimal strategies for a list of countries. It details the average policy-induced transmission reduction and the corresponding total societal cost (in billions of USD). The cost-optimal solutions are calculated under four distinct scenarios: a country-specific Default cost per infection, and three fixed hypothetical valuations of $1,000, $10,000, and $100,000 per infection, illustrating how the optimal policy intensity shifts as the economic weight of an infection increases.

| Country | Estimated result (Default cost per infection) | | Cost-optimal solution (under different cost per infection) | | | | | | | |
|---|---|---|---|---|---|---|---|---|---|---|
| | | | Default | | $1K | | $10K | | $100K | |
| | Average transmission reduction | Total cost (Billion USD) | Cost-optimal transmission reduction | Total cost (Billion USD) | Cost-optimal transmission reduction | Total cost (Billion USD) | Cost-optimal transmission reduction | Total cost (Billion USD) | Cost-optimal transmission reduction | Total cost (Billion USD) |
| Afghanistan | 0.6017 | 1.301 | 0.5146 | 1.115 | 0.5153 | 1.080 | 0.5691 | 1.227 | 0.6475 | 1.450 |
| Algeria | 0.5664 | 12.382 | 0.5129 | 10.724 | 0.4826 | 10.060 | 0.5115 | 10.728 | 0.5671 | 12.128 |
| Angola | 0.5740 | 4.681 | 0.4821 | 3.989 | 0.4693 | 3.858 | 0.4968 | 4.100 | 0.5440 | 4.616 |
| Argentina | 0.5394 | 63.014 | 0.5193 | 24.375 | 0.4741 | 22.281 | 0.5142 | 24.008 | 0.5494 | 26.985 |
| Australia | 0.5807 | 87.447 | 0.5230 | 78.732 | 0.1667 | 19.300 | 0.4906 | 72.634 | 0.5196 | 78.599 |
| Austria | 0.5411 | 60.755 | 0.5203 | 24.145 | 0.1611 | 18.384 | 0.4930 | 22.707 | 0.5273 | 24.559 |
| Azerbaijan | 0.5620 | 3.902 | 0.5091 | 2.582 | 0.4859 | 2.439 | 0.5220 | 2.625 | 0.5755 | 2.943 |
| Bangladesh | 0.5626 | 21.585 | 0.4873 | 18.206 | 0.4767 | 17.659 | 0.5105 | 18.890 | 0.5556 | 20.934 |
| Belarus | 0.5495 | 4.904 | 0.5178 | 3.462 | 0.4967 | 3.267 | 0.5333 | 3.548 | 0.5961 | 4.071 |
| Belgium | 0.5251 | 96.666 | 0.5276 | 30.715 | 0.2143 | 16.432 | 0.4909 | 28.048 | 0.5351 | 30.715 |
| Benin | 0.5684 | 0.890 | 0.4802 | 0.761 | 0.4800 | 0.761 | 0.5109 | 0.820 | 0.5528 | 0.909 |
| Bolivia | 0.5435 | 4.877 | 0.5078 | 2.252 | 0.4755 | 2.075 | 0.5122 | 2.252 | 0.5563 | 2.485 |
| Brazil | 0.5207 | 254.222 | 0.4905 | 98.539 | 0.4582 | 90.545 | 0.4856 | 96.157 | 0.5213 | 105.159 |
| Bulgaria | 0.5586 | 10.086 | 0.5727 | 3.859 | 0.5022 | 3.335 | 0.5517 | 3.694 | 0.6200 | 4.316 |
| Burkina Faso | 0.6123 | 1.086 | 0.5079 | 0.940 | 0.5194 | 0.941 | 0.5716 | 1.068 | 0.6542 | 1.272 |
| Burundi | 0.5818 | 0.168 | 0.4732 | 0.142 | 0.5177 | 0.156 | 0.5731 | 0.173 | 0.6536 | 0.208 |
| Cambodia | 0.5982 | 2.203 | 0.4525 | 1.705 | 0.4595 | 1.737 | 0.4948 | 1.885 | 0.5374 | 2.072 |
| Cameroon | 0.5680 | 2.480 | 0.5204 | 2.263 | 0.5060 | 2.236 | 0.5484 | 2.443 | 0.6263 | 2.892 |
| Canada | 0.5354 | 158.350 | 0.4989 | 93.333 | 0.2154 | 47.023 | 0.4708 | 87.179 | 0.5109 | 94.965 |
| Chad | 0.5711 | 0.671 | 0.5064 | 0.608 | 0.4998 | 0.601 | 0.5373 | 0.651 | 0.6108 | 0.754 |
| Chile | 0.5489 | 35.557 | 0.5279 | 15.618 | 0.4334 | 13.698 | 0.5153 | 15.045 | 0.5580 | 16.744 |
| China | 0.5626 | 821.592 | 0.4740 | 708.414 | 0.4395 | 650.009 | 0.4582 | 686.591 | 0.4928 | 737.813 |
| Colombia | 0.5346 | 45.084 | 0.4995 | 17.196 | 0.4672 | 16.068 | 0.4986 | 17.196 | 0.5336 | 18.805 |
| Costa Rica | 0.5676 | 6.608 | 0.5396 | 3.829 | 0.4557 | 3.437 | 0.5347 | 3.754 | 0.6002 | 4.327 |
| Cote d'Ivoire | 0.5421 | 3.378 | 0.4735 | 2.984 | 0.4697 | 2.972 | 0.5039 | 3.221 | 0.5568 | 3.582 |
| Cuba | 0.5675 | 6.243 | 0.5222 | 5.637 | 0.4866 | 5.207 | 0.5360 | 5.751 | 0.5964 | 6.556 |
| Czechia | 0.5233 | 60.323 | 0.5325 | 13.787 | 0.3869 | 12.534 | 0.5188 | 13.559 | 0.5619 | 14.968 |
| Democratic Republic of Congo | 0.5879 | 3.483 | 0.4981 | 3.094 | 0.5071 | 3.094 | 0.5455 | 3.388 | 0.6161 | 3.933 |
| Denmark | 0.5495 | 30.733 | 0.5373 | 19.547 | 0.1726 | 9.825 | 0.5041 | 18.393 | 0.5442 | 20.133 |
| Dominican Republic | 0.5695 | 7.225 | 0.5303 | 5.147 | 0.4874 | 4.717 | 0.5389 | 5.229 | 0.5975 | 5.950 |
| Ecuador | 0.5348 | 12.90048 | 0.5394 | 6.312 | 0.4747 | 5.470 | 0.5260 | 6.181 | 0.5828 | 6.908 |
| Egypt | 0.5620 | 19.979 | 0.4988 | 16.432 | 0.4653 | 15.410 | 0.4950 | 16.432 | 0.5378 | 18.515 |
| El Salvador | 0.5717 | 2.195 | 0.5574 | 1.648 | 0.5105 | 1.482 | 0.5636 | 1.683 | 0.6382 | 1.962 |
| Ethiopia | 0.5494 | 5.932 | 0.4824 | 5.208 | 0.4860 | 5.224 | 0.5194 | 5.606 | 0.5688 | 6.344 |
| Finland | 0.5744 | 18.054 | 0.5440 | 15.152 | 0.2124 | 2.750 | 0.5122 | 14.190 | 0.5589 | 15.692 |
| France | 0.4996 | 354.233 | 0.4644 | 127.917 | 0.1767 | 81.594 | 0.4481 | 120.322 | 0.4722 | 128.821 |
| Germany | 0.5248 | 377.048 | 0.5014 | 205.021 | 0.1986 | 90.218 | 0.4739 | 192.430 | 0.5055 | 206.745 |
| Ghana | 0.5609 | 4.241 | 0.4989 | 3.806 | 0.4935 | 3.716 | 0.5333 | 4.093 | 0.5952 | 4.616 |
| Greece | 0.5620 | 19.305 | 0.5555 | 11.601 | 0.4413 | 9.995 | 0.5271 | 11.022 | 0.5724 | 12.212 |
| Guatemala | 0.5734 | 6.549 | 0.5273 | 4.464 | 0.4948 | 4.090 | 0.5271 | 4.464 | 0.5839 | 5.077 |
| Guinea | 0.5891 | 0.884 | 0.5125 | 0.788 | 0.5201 | 0.804 | 0.5716 | 0.890 | 0.6637 | 1.093 |
| Haiti | 0.6145 | 0.996 | 0.5182 | 0.874 | 0.5114 | 0.836 | 0.5703 | 0.953 | 0.6533 | 1.121 |
| Honduras | 0.5651 | 2.202 | 0.5547 | 1.486 | 0.5223 | 1.396 | 0.5753 | 1.561 | 0.6596 | 1.853 |
| Hungary | 0.5379 | 27.530 | 0.5450 | 9.095 | 0.4832 | 8.069 | 0.5164 | 8.553 | 0.5703 | 9.648 |
| India | 0.5217 | 176.449 | 0.4517 | 134.045 | 0.4446 | 131.600 | 0.4692 | 139.407 | 0.4974 | 148.236 |
| Indonesia | 0.5352 | 73.596 | 0.4876 | 57.732 | 0.4513 | 53.498 | 0.4920 | 58.460 | 0.5362 | 64.650 |
| Iran | 0.5240 | 34.328 | 0.5057 | 18.188 | 0.4824 | 16.906 | 0.5113 | 18.235 | 0.5715 | 20.477 |
| Iraq | 0.5411 | 19.527 | 0.5055 | 13.301 | 0.4637 | 12.238 | 0.5034 | 13.385 | 0.5501 | 14.978 |
| Israel | 0.5183 | 48.364 | 0.5166 | 22.587 | 0.2363 | 16.679 | 0.4959 | 21.919 | 0.5314 | 23.816 |

| | | | | | | | | | | |
|---|---|---|---|---|---|---|---|---|---|---|
| Italy | 0.5141 | 302.100 | 0.5065 | 103.726 | 0.2722 | 88.651 | 0.4687 | 95.560 | 0.5083 | 104.553 |
| Japan | 0.5113 | 275.165 | 0.4564 | 236.117 | 0.2051 | 193.098 | 0.4452 | 226.804 | 0.4698 | 242.516 |
| Jordan | 0.5105 | 4.346 | 0.4655 | 2.255 | 0.4507 | 2.201 | 0.4783 | 2.338 | 0.5095 | 2.512 |
| Kazakhstan | 0.5522 | 12.463 | 0.5005 | 9.328 | 0.4740 | 8.715 | 0.5057 | 9.490 | 0.5516 | 10.524 |
| Kenya | 0.5381 | 6.103 | 0.4745 | 5.241 | 0.4665 | 5.165 | 0.4950 | 5.557 | 0.5435 | 6.125 |
| Kyrgyzstan | 0.6230 | 0.711 | 0.5482 | 0.553 | 0.5412 | 0.545 | 0.6001 | 0.623 | 0.7256 | 0.786 |
| Laos | 0.7641 | 1.441 | 0.4626 | 0.893 | 0.4674 | 0.911 | 0.5023 | 0.988 | 0.5526 | 1.091 |
| Lebanon | 0.5440 | 6.053 | 0.5120 | 2.751 | 0.4924 | 2.563 | 0.5209 | 2.763 | 0.5799 | 3.145 |
| Liberia | 0.6501 | 0.225 | 0.5521 | 0.198 | 0.5376 | 0.191 | 0.6087 | 0.222 | 0.7180 | 0.276 |
| Libya | 0.5574 | 6.057 | 0.5316 | 3.897 | 0.4556 | 3.530 | 0.5327 | 3.849 | 0.5894 | 4.412 |
| Madagascar | 0.5660 | 0.869 | 0.4934 | 0.781 | 0.5084 | 0.799 | 0.5593 | 0.906 | 0.6408 | 1.054 |
| Malawi | 0.5909 | 0.827 | 0.5213 | 0.722 | 0.5172 | 0.720 | 0.5720 | 0.801 | 0.6515 | 0.955 |
| Malaysia | 0.5588 | 21.715 | 0.4995 | 19.355 | 0.4480 | 18.393 | 0.5155 | 19.792 | 0.5618 | 21.896 |
| Mali | 0.5745 | 1.183 | 0.5094 | 1.095 | 0.5061 | 1.053 | 0.5520 | 1.172 | 0.6211 | 1.355 |
| Mexico | 0.5339 | 210.086 | 0.5313 | 72.250 | 0.4720 | 64.878 | 0.4977 | 69.227 | 0.5342 | 73.334 |
| Morocco | 0.5505 | 9.479 | 0.4827 | 6.761 | 0.4614 | 6.274 | 0.5028 | 6.915 | 0.5515 | 7.697 |
| Mozambique | 0.5592 | 1.036 | 0.4895 | 0.906 | 0.5038 | 0.933 | 0.5474 | 1.027 | 0.6219 | 1.201 |
| Myanmar | 0.6001 | 4.684 | 0.4790 | 3.589 | 0.4694 | 3.509 | 0.5007 | 3.817 | 0.5533 | 4.262 |
| Nepal | 0.5560 | 2.339 | 0.4799 | 1.808 | 0.4790 | 1.808 | 0.5163 | 1.961 | 0.5628 | 2.193 |
| Netherlands | 0.5357 | 113.655 | 0.5334 | 52.154 | 0.2326 | 31.437 | 0.4994 | 48.043 | 0.5477 | 53.394 |
| Niger | 0.6670 | 1.106 | 0.5273 | 0.911 | 0.5254 | 0.906 | 0.5789 | 0.999 | 0.6641 | 1.226 |
| Nigeria | 0.5578 | 28.546 | 0.4808 | 24.791 | 0.4697 | 24.345 | 0.4982 | 26.053 | 0.5397 | 28.583 |
| Pakistan | 0.5570 | 19.927 | 0.4836 | 16.721 | 0.4736 | 16.352 | 0.5127 | 17.913 | 0.5643 | 19.934 |
| Paraguay | 0.5508 | 4.014 | 0.5461 | 2.445 | 0.5048 | 2.239 | 0.5436 | 2.445 | 0.6198 | 2.855 |
| Peru | 0.5257 | 67.235 | 0.5422 | 13.439 | 0.4784 | 11.724 | 0.5107 | 12.607 | 0.5596 | 14.011 |
| Philippines | 0.5575 | 24.017 | 0.4879 | 19.503 | 0.4720 | 18.625 | 0.5014 | 20.062 | 0.5514 | 22.217 |
| Poland | 0.5054 | 82.564 | 0.4807 | 29.855 | 0.4055 | 27.036 | 0.4689 | 28.914 | 0.5068 | 31.510 |
| Portugal | 0.5387 | 40.802 | 0.5436 | 13.568 | 0.3062 | 10.779 | 0.5231 | 12.772 | 0.5732 | 14.432 |
| Romania | 0.5419 | 31.759 | 0.5258 | 13.508 | 0.4848 | 12.050 | 0.5123 | 13.016 | 0.5700 | 14.808 |
| Russia | 0.5245 | 150.955 | 0.4858 | 83.632 | 0.4543 | 76.822 | 0.4825 | 83.468 | 0.5267 | 90.943 |
| Rwanda | 0.5779 | 0.676 | 0.5109 | 0.615 | 0.5232 | 0.619 | 0.5765 | 0.702 | 0.6567 | 0.822 |
| Saudi Arabia | 0.5453 | 69.847 | 0.4956 | 52.200 | 0.2897 | 43.339 | 0.4802 | 50.643 | 0.5133 | 55.359 |
| Senegal | 0.5824 | 1.608 | 0.5261 | 1.438 | 0.5029 | 1.359 | 0.5535 | 1.503 | 0.6277 | 1.801 |
| Serbia | 0.5235 | 6.001 | 0.5167 | 2.789 | 0.4972 | 2.618 | 0.5264 | 2.805 | 0.5836 | 3.202 |
| Sierra Leone | 0.6330 | 0.473 | 0.5250 | 0.405 | 0.5183 | 0.395 | 0.5772 | 0.453 | 0.6767 | 0.554 |
| Slovakia | 0.5504 | 16.382 | 0.5594 | 6.113 | 0.3761 | 5.303 | 0.5287 | 5.790 | 0.5931 | 6.600 |
| Somalia | 0.5811 | 0.582 | 0.5198 | 0.537 | 0.5187 | 0.527 | 0.5744 | 0.594 | 0.6562 | 0.706 |
| South Africa | 0.5420 | 43.834 | 0.5193 | 22.137 | 0.4741 | 19.897 | 0.5084 | 21.832 | 0.5576 | 24.106 |
| Republic of Korea | 0.5482 | 92.520 | 0.5013 | 84.733 | 0.1646 | 53.388 | 0.4829 | 80.835 | 0.5184 | 88.881 |
| South Sudan | 0.6011 | 0.331 | 0.5117 | 0.284 | 0.5268 | 0.299 | 0.5879 | 0.342 | 0.6822 | 0.414 |
| Spain | 0.5032 | 193.732 | 0.4933 | 73.866 | 0.3553 | 67.878 | 0.4813 | 71.749 | 0.5060 | 75.976 |
| Sri Lanka | 0.5387 | 4.872 | 0.4619 | 4.214 | 0.4546 | 4.075 | 0.4878 | 4.413 | 0.5341 | 4.811 |
| Sudan | 0.5974 | 2.215 | 0.5172 | 1.902 | 0.5085 | 1.856 | 0.5433 | 2.068 | 0.6342 | 2.460 |
| Syria | 0.5658 | 1.472 | 0.5075 | 1.298 | 0.4895 | 1.224 | 0.5236 | 1.346 | 0.5866 | 1.515 |
| Tajikistan | 0.6823 | 0.609 | 0.5383 | 0.491 | 0.5491 | 0.504 | 0.6115 | 0.584 | 0.7260 | 0.726 |
| Tanzania | 0.8071 | 5.286 | 0.4819 | 3.243 | 0.4720 | 3.149 | 0.4994 | 3.406 | 0.5495 | 3.783 |
| Thailand | 0.5312 | 26.654 | 0.4553 | 23.463 | 0.4443 | 22.726 | 0.4664 | 23.785 | 0.4889 | 25.077 |
| Togo | 0.5598 | 0.450 | 0.4862 | 0.392 | 0.4904 | 0.401 | 0.5297 | 0.438 | 0.5873 | 0.497 |
| Tunisia | 0.5648 | 4.196 | 0.5243 | 2.401 | 0.4899 | 2.183 | 0.5302 | 2.415 | 0.5972 | 2.734 |
| Turkey | 0.5187 | 64.499 | 0.4840 | 38.942 | 0.4596 | 36.875 | 0.4877 | 39.435 | 0.5273 | 43.437 |
| Uganda | 0.6016 | 2.420 | 0.4865 | 2.039 | 0.5014 | 2.065 | 0.5435 | 2.251 | 0.6059 | 2.619 |
| Ukraine | 0.5331 | 13.660 | 0.5045 | 6.656 | 0.4861 | 6.333 | 0.5177 | 6.804 | 0.5675 | 7.609 |
| United Arab Emirates | 0.5427 | 31.206 | 0.5065 | 23.772 | 0.1662 | 14.628 | 0.4995 | 23.262 | 0.5416 | 25.590 |
| United Kingdom | 0.5089 | 507.486 | 0.5038 | 151.391 | 0.1671 | 103.161 | 0.4678 | 140.293 | 0.5013 | 151.391 |
| United States | 0.4945 | 3482.136 | 0.4898 | 1109.648 | 0.1950 | 686.066 | 0.4613 | 1044.879 | 0.4904 | 1109.648 |
| Uzbekistan | 0.5797 | 4.460 | 0.4845 | 3.700 | 0.4809 | 3.690 | 0.5101 | 3.993 | 0.5617 | 4.457 |
| Venezuela | 0.5543 | 10.278 | 0.4865 | 8.737 | 0.4640 | 8.325 | 0.5007 | 8.973 | 0.5496 | 9.894 |
| Vietnam | 0.5673 | 19.736 | 0.4518 | 16.166 | 0.4365 | 15.383 | 0.4585 | 16.307 | 0.4921 | 17.553 |
| Yemen | 0.6199 | 1.376 | 0.5436 | 1.212 | 0.5016 | 1.115 | 0.5496 | 1.242 | 0.6208 | 1.440 |
| Zambia | 0.5745 | 1.596 | 0.5112 | 1.394 | 0.5080 | 1.373 | 0.5587 | 1.525 | 0.6391 | 1.799 |
| Zimbabwe | 0.5915 | 1.878 | 0.5317 | 1.626 | 0.5112 | 1.539 | 0.5527 | 1.686 | 0.6262 | 2.001 |

#### Supplementary 1K. Generalised cost-optimal intervention results for structural index category.

Table S1.9-11 represent raw data for the Figure 4 in the main manuscript with different vaccine development timing scenario.

**Table S1.9. Generalised cost-optimal intervention results for 26 weeks vaccine development timing.**

**Region**

| Category | Cost per infection | Cost-optimal average transmission reduction | | | | |
|---|---|---|---|---|---|---|
| | | Min | 1Q | Median | 3Q | Max |
| High-income | $100K | 0.3485 | 0.4111 | 0.4270 | 0.4529 | 0.5118 |
| | $10K | 0.3017 | 0.3535 | 0.3759 | 0.4063 | 0.4284 |
| | $1K | 0.2686 | 0.2960 | 0.3112 | 0.3274 | 0.3728 |
| Central Europe, Eastern Europe, and Central Asia | $100K | 0.4324 | 0.4905 | 0.4998 | 0.5347 | 0.5590 |
| | $10K | 0.4124 | 0.4194 | 0.4268 | 0.4452 | 0.4617 |
| | $1K | 0.3052 | 0.3594 | 0.3737 | 0.3971 | 0.4110 |
| North Africa and Middle East | $100K | 0.4231 | 0.4569 | 0.4936 | 0.5290 | 0.6039 |
| | $10K | 0.3655 | 0.4005 | 0.4235 | 0.4430 | 0.4946 |
| | $1K | 0.3093 | 0.3447 | 0.3669 | 0.3961 | 0.4253 |
| Latin America and Caribbean | $100K | 0.4248 | 0.4762 | 0.5117 | 0.5461 | 0.6003 |
| | $10K | 0.3663 | 0.4159 | 0.4300 | 0.4544 | 0.4971 |
| | $1K | 0.3207 | 0.3596 | 0.3780 | 0.4044 | 0.4273 |
| Southeast Asia, East Asia, and Oceania | $100K | 0.3911 | 0.4021 | 0.4489 | 0.4768 | 0.4865 |
| | $10K | 0.3382 | 0.3440 | 0.3934 | 0.4087 | 0.4218 |
| | $1K | 0.2925 | 0.2974 | 0.3410 | 0.3549 | 0.3573 |
| Sub-Saharan Africa | $100K | 0.4045 | 0.4866 | 0.5608 | 0.6029 | 0.6715 |
| | $10K | 0.3521 | 0.4167 | 0.4642 | 0.4994 | 0.5406 |
| | $1K | 0.3014 | 0.3643 | 0.4070 | 0.4257 | 0.4520 |

**Population**

| Category | Cost per infection | Cost-optimal average transmission reduction | | | | |
|---|---|---|---|---|---|---|
| | | Min | 1Q | Median | 3Q | Max |
| More than 100M | $100K | 0.3579 | 0.4018 | 0.4436 | 0.4714 | 0.5599 |
| | $10K | 0.3113 | 0.3460 | 0.3939 | 0.4106 | 0.4611 |
| | $1K | 0.2686 | 0.2987 | 0.3384 | 0.3566 | 0.4067 |
| Between 30M and 100M | $100K | 0.3485 | 0.4298 | 0.4768 | 0.4996 | 0.6039 |
| | $10K | 0.3017 | 0.3728 | 0.4150 | 0.4280 | 0.4946 |
| | $1K | 0.2706 | 0.3207 | 0.3573 | 0.3695 | 0.4257 |
| Between 10M and 30M | $100K | 0.4231 | 0.4861 | 0.5249 | 0.5981 | 0.6875 |
| | $10K | 0.3655 | 0.4170 | 0.4356 | 0.4970 | 0.5515 |
| | $1K | 0.3093 | 0.3625 | 0.3891 | 0.4231 | 0.4550 |
| Lesser than 10M | $100K | 0.4366 | 0.4833 | 0.5232 | 0.5624 | 0.6715 |
| | $10K | 0.3838 | 0.4161 | 0.4413 | 0.4629 | 0.5080 |
| | $1K | 0.2964 | 0.3594 | 0.3962 | 0.4111 | 0.4575 |

**Income**

| Category | Cost per infection | Cost-optimal average transmission reduction | | | | |
|---|---|---|---|---|---|---|
| | | Min | 1Q | Median | 3Q | Max |
| More than $17K | $100K | 0.3485 | 0.4120 | 0.4356 | 0.4609 | 0.5232 |
| | $10K | 0.3017 | 0.3549 | 0.3813 | 0.4067 | 0.4413 |
| | $1K | 0.2686 | 0.2964 | 0.3177 | 0.3348 | 0.3772 |
| Between $7K and $17K | $100K | 0.3911 | 0.4385 | 0.4879 | 0.5158 | 0.5590 |
| | $10K | 0.3382 | 0.3846 | 0.4201 | 0.4350 | 0.4617 |
| | $1K | 0.2960 | 0.3338 | 0.3663 | 0.3839 | 0.4110 |
| Between $2K and $7K | $100K | 0.3990 | 0.4583 | 0.4797 | 0.5117 | 0.5835 |
| | $10K | 0.3895 | 0.4060 | 0.4153 | 0.4300 | 0.4633 |
| | $1K | 0.3162 | 0.3517 | 0.3597 | 0.3780 | 0.4111 |
| Lesser than $2K | $100K | 0.4045 | 0.5123 | 0.5747 | 0.6044 | 0.6875 |
| | $10K | 0.3521 | 0.4344 | 0.4740 | 0.5021 | 0.5515 |
| | $1K | 0.3436 | 0.3783 | 0.4169 | 0.4263 | 0.4575 |

**UHC**

| Category | Cost per infection | Cost-optimal average transmission reduction | | | | |
|---|---|---|---|---|---|---|
| | | Min | 1Q | Median | 3Q | Max |
| More than 72 | $100K | 0.3485 | 0.4148 | 0.4478 | 0.4947 | 0.5403 |
| | $10K | 0.3017 | 0.3553 | 0.4005 | 0.4212 | 0.4437 |
| | $1K | 0.2686 | 0.3011 | 0.3270 | 0.3692 | 0.4009 |
| Between 59.3 and 72 | $100K | 0.3911 | 0.4361 | 0.4799 | 0.5165 | 0.6080 |
| | $10K | 0.3382 | 0.3773 | 0.4196 | 0.4371 | 0.5046 |
| | $1K | 0.2925 | 0.3211 | 0.3629 | 0.3848 | 0.4304 |
| Between 47.9 and 59.3 | $100K | 0.4405 | 0.4658 | 0.5184 | 0.5774 | 0.6875 |
| | $10K | 0.3895 | 0.4106 | 0.4352 | 0.4727 | 0.5515 |
| | $1K | 0.3390 | 0.3564 | 0.3841 | 0.4178 | 0.4575 |
| Lesser than 47.9 | $100K | 0.4045 | 0.4871 | 0.5599 | 0.6026 | 0.6715 |
| | $10K | 0.3521 | 0.4173 | 0.4611 | 0.5008 | 0.5406 |
| | $1K | 0.3014 | 0.3583 | 0.4067 | 0.4256 | 0.4520 |

**SDI**

| Category | Cost per infection | Cost-optimal average transmission reduction | | | | |
|---|---|---|---|---|---|---|
| | | Min | 1Q | Median | 3Q | Max |
| More than 0.76 | $100K | 0.3485 | 0.4148 | 0.4396 | 0.4943 | 0.5590 |
| | $10K | 0.3017 | 0.3555 | 0.3899 | 0.4207 | 0.4617 |
| | $1K | 0.2686 | 0.3019 | 0.3221 | 0.3671 | 0.4110 |
| Between 0.64 and 0.76 | $100K | 0.3911 | 0.4455 | 0.4865 | 0.5076 | 0.5411 |
| | $10K | 0.3655 | 0.3948 | 0.4196 | 0.4281 | 0.4503 |
| | $1K | 0.2960 | 0.3393 | 0.3629 | 0.3735 | 0.4069 |
| Between 0.46 and 0.64 | $100K | 0.3990 | 0.4624 | 0.4886 | 0.5730 | 0.6875 |
| | $10K | 0.3398 | 0.4060 | 0.4202 | 0.4713 | 0.5515 |
| | $1K | 0.2925 | 0.3535 | 0.3607 | 0.4155 | 0.4575 |
| Lesser than 0.46 | $100K | 0.4624 | 0.5320 | 0.5800 | 0.6056 | 0.6715 |
| | $10K | 0.4089 | 0.4480 | 0.4768 | 0.5022 | 0.5406 |
| | $1K | 0.3576 | 0.3984 | 0.4181 | 0.4271 | 0.4520 |

**GHS**

| Category | Cost per infection | Cost-optimal average transmission reduction | | | | |
|---|---|---|---|---|---|---|
| | | Min | 1Q | Median | 3Q | Max |
| More than 50.7 | $100K | 0.3485 | 0.4140 | 0.4396 | 0.4869 | 0.5590 |
| | $10K | 0.3017 | 0.3546 | 0.3899 | 0.4197 | 0.4617 |
| | $1K | 0.2686 | 0.2990 | 0.3241 | 0.3637 | 0.4110 |
| Between 37.7 and 50.7 | $100K | 0.3911 | 0.4405 | 0.4735 | 0.5145 | 0.5835 |
| | $10K | 0.3382 | 0.3886 | 0.4124 | 0.4359 | 0.4821 |
| | $1K | 0.2925 | 0.3260 | 0.3562 | 0.3821 | 0.4575 |
| Between 31 and 37.7 | $100K | 0.4450 | 0.4796 | 0.5025 | 0.5610 | 0.6715 |
| | $10K | 0.3931 | 0.4128 | 0.4292 | 0.4660 | 0.5406 |
| | $1K | 0.3404 | 0.3574 | 0.3737 | 0.4098 | 0.4520 |
| Lesser than 31 | $100K | 0.4620 | 0.5159 | 0.5718 | 0.6030 | 0.6875 |
| | $10K | 0.4042 | 0.4351 | 0.4714 | 0.4980 | 0.5515 |
| | $1K | 0.3517 | 0.3823 | 0.4146 | 0.4259 | 0.4550 |

*__Region__: G1:Central Europe, Eastern Europe, and Central Asia; G2:High income; G3:Latin America and Caribbean; G4:North Africa and Middle East; G5: Southeast Asia, East Asia, and Oceania; G6:Sub-Saharan Africa.
**Population**: G1:more than 100M, G2:30M-100M, G3:10M-30M, G4:5M-10M.
**Income**: G1:more than $17K, G2:$7K-$17K, G3:$2K-$7K, G4:less than $2K.
**UHC**: G1:more than 72, G2:59.3-72, G3:47.9-59.3, G4:less than 47.9.
**SDI**: G1:more than 0.76, G2:0.64-0.76, G3:0.46-0.64, G4:less than 0.46.
**GHS**: G1:more than 50.7, G2:37.7-50.7, G3:31-37.7, G4:less than 31.

#### Table S1.10. Generalised cost-optimal intervention results for 40 weeks vaccine development timing.

**Region**

| Category | Cost per infection | Cost-optimal average transmission reduction | | | | |
|---|---|---|---|---|---|---|
| | | **Min** | **1Q** | **Median** | **3Q** | **Max** |
| High-income | $100K | 0.4312 | 0.4721 | 0.4894 | 0.5107 | 0.5539 |
| | $10K | 0.3978 | 0.4352 | 0.4487 | 0.4623 | 0.4900 |
| | $1K | 0.1871 | 0.2271 | 0.2885 | 0.3369 | 0.4322 |
| Central Europe, Eastern Europe, and Central Asia | $100K | 0.4757 | 0.5336 | 0.5488 | 0.5790 | 0.6027 |
| | $10K | 0.4306 | 0.4773 | 0.4863 | 0.5116 | 0.5150 |
| | $1K | 0.3919 | 0.4317 | 0.4466 | 0.4650 | 0.5139 |
| North Africa and Middle East | $100K | 0.4890 | 0.5114 | 0.5370 | 0.5738 | 0.6381 |
| | $10K | 0.4407 | 0.4638 | 0.4854 | 0.5059 | 0.5372 |
| | $1K | 0.4046 | 0.4261 | 0.4429 | 0.4596 | 0.4834 |
| Latin America and Caribbean | $100K | 0.4887 | 0.5269 | 0.5555 | 0.5831 | 0.6324 |
| | $10K | 0.4504 | 0.4743 | 0.4877 | 0.5146 | 0.5501 |
| | $1K | 0.4169 | 0.4330 | 0.4459 | 0.4653 | 0.4880 |
| Southeast Asia, East Asia, and Oceania | $100K | 0.4527 | 0.4688 | 0.5049 | 0.5133 | 0.5311 |
| | $10K | 0.4203 | 0.4317 | 0.4557 | 0.4698 | 0.4782 |
| | $1K | 0.3893 | 0.4040 | 0.4254 | 0.4362 | 0.4438 |
| Sub-Saharan Africa | $100K | 0.4627 | 0.5378 | 0.6017 | 0.6373 | 0.6996 |
| | $10K | 0.4286 | 0.4801 | 0.5156 | 0.5467 | 0.5851 |
| | $1K | 0.3999 | 0.4410 | 0.4700 | 0.4891 | 0.5128 |

**Population**

| Category | Cost per infection | Cost-optimal average transmission reduction | | | | |
|---|---|---|---|---|---|---|
| | | **Min** | **1Q** | **Median** | **3Q** | **Max** |
| More than 100M | $100K | 0.4396 | 0.4598 | 0.4988 | 0.5218 | 0.6033 |
| | $10K | 0.4079 | 0.4262 | 0.4550 | 0.4750 | 0.5206 |
| | $1K | 0.3893 | 0.3955 | 0.4211 | 0.4342 | 0.4714 |
| Between 30M and 100M | $100K | 0.4312 | 0.4894 | 0.5267 | 0.5474 | 0.6246 |
| | $10K | 0.3978 | 0.4451 | 0.4721 | 0.4877 | 0.5372 |
| | $1K | 0.3418 | 0.4040 | 0.4371 | 0.4459 | 0.4834 |
| Between 10M and 30M | $100K | 0.4889 | 0.5315 | 0.5696 | 0.6313 | 0.7194 |
| | $10K | 0.4407 | 0.4785 | 0.5010 | 0.5418 | 0.5935 |
| | $1K | 0.3919 | 0.4310 | 0.4542 | 0.4816 | 0.5170 |
| Lesser than 10M | $100K | 0.4900 | 0.5288 | 0.5657 | 0.6007 | 0.7081 |
| | $10K | 0.4525 | 0.4765 | 0.5028 | 0.5190 | 0.5536 |
| | $1K | 0.4373 | 0.4383 | 0.4562 | 0.4729 | 0.5139 |

**Income**

| Category | Cost per infection | Cost-optimal average transmission reduction | | | | |
|---|---|---|---|---|---|---|
| | | **Min** | **1Q** | **Median** | **3Q** | **Max** |
| More than $17K | $100K | 0.4312 | 0.4761 | 0.4904 | 0.5200 | 0.5708 |
| | $10K | 0.3978 | 0.4365 | 0.4533 | 0.4750 | 0.5095 |
| | $1K | 0.1871 | 0.2378 | 0.3008 | 0.3418 | 0.4617 |
| Between $7K and $17K | $100K | 0.4569 | 0.4935 | 0.5336 | 0.5572 | 0.6027 |
| | $10K | 0.4238 | 0.4499 | 0.4782 | 0.4992 | 0.5127 |
| | $1K | 0.3893 | 0.4193 | 0.4404 | 0.4528 | 0.4754 |
| Between $2K and $7K | $100K | 0.4527 | 0.5133 | 0.5281 | 0.5555 | 0.5947 |
| | $10K | 0.4407 | 0.4686 | 0.4785 | 0.4876 | 0.5150 |
| | $1K | 0.4046 | 0.4291 | 0.4389 | 0.4466 | 0.4701 |
| Lesser than $2K | $100K | 0.4627 | 0.5617 | 0.6132 | 0.6403 | 0.7194 |
| | $10K | 0.4286 | 0.4911 | 0.5269 | 0.5474 | 0.5935 |
| | $1K | 0.3999 | 0.4505 | 0.4773 | 0.4896 | 0.5170 |

**UHC**

| Category | Cost per infection | Cost-optimal average transmission reduction | | | | |
|---|---|---|---|---|---|---|
| | | **Min** | **1Q** | **Median** | **3Q** | **Max** |
| More than 72 | $100K | 0.4312 | 0.4767 | 0.5052 | 0.5422 | 0.5792 |
| | $10K | 0.3978 | 0.4371 | 0.4598 | 0.4865 | 0.5127 |
| | $1K | 0.1871 | 0.2498 | 0.3393 | 0.4333 | 0.4637 |
| Between 59.3 and 72 | $100K | 0.4527 | 0.4922 | 0.5293 | 0.5631 | 0.6454 |
| | $10K | 0.4203 | 0.4504 | 0.4778 | 0.5016 | 0.5406 |
| | $1K | 0.3893 | 0.4147 | 0.4410 | 0.4512 | 0.4891 |
| Between 47.9 and 59.3 | $100K | 0.5001 | 0.5215 | 0.5634 | 0.6140 | 0.7194 |
| | $10K | 0.4547 | 0.4702 | 0.4982 | 0.5294 | 0.5935 |
| | $1K | 0.4181 | 0.4329 | 0.4522 | 0.4761 | 0.5170 |
| Lesser than 47.9 | $100K | 0.4627 | 0.5381 | 0.6001 | 0.6379 | 0.6996 |
| | $10K | 0.4286 | 0.4793 | 0.5113 | 0.5451 | 0.5851 |
| | $1K | 0.3999 | 0.4413 | 0.4686 | 0.4868 | 0.5128 |

**SDI**

| Category | Cost per infection | Cost-optimal average transmission reduction | | | | |
|---|---|---|---|---|---|---|
| | | **Min** | **1Q** | **Median** | **3Q** | **Max** |
| More than 0.76 | $100K | 0.4312 | 0.4764 | 0.4962 | 0.5409 | 0.6027 |
| | $10K | 0.3978 | 0.4376 | 0.4546 | 0.4817 | 0.5150 |
| | $1K | 0.1871 | 0.2510 | 0.3317 | 0.4327 | 0.4754 |
| Between 0.64 and 0.76 | $100K | 0.4569 | 0.5008 | 0.5311 | 0.5492 | 0.5818 |
| | $10K | 0.4238 | 0.4542 | 0.4782 | 0.4877 | 0.5138 |
| | $1K | 0.3893 | 0.4208 | 0.4404 | 0.4456 | 0.4637 |
| Between 0.46 and 0.64 | $100K | 0.4527 | 0.5140 | 0.5389 | 0.6071 | 0.7194 |
| | $10K | 0.4203 | 0.4689 | 0.4822 | 0.5273 | 0.5935 |
| | $1K | 0.3911 | 0.4323 | 0.4412 | 0.4734 | 0.5170 |
| Lesser than 0.46 | $100K | 0.5182 | 0.5707 | 0.6176 | 0.6428 | 0.6996 |
| | $10K | 0.4696 | 0.4952 | 0.5269 | 0.5479 | 0.5851 |
| | $1K | 0.4310 | 0.4548 | 0.4783 | 0.4900 | 0.5128 |

**GHS**

| Category | Cost per infection | Cost-optimal average transmission reduction | | | | |
|---|---|---|---|---|---|---|
| | | **Min** | **1Q** | **Median** | **3Q** | **Max** |
| More than 50.7 | $100K | 0.4312 | 0.4760 | 0.4962 | 0.5306 | 0.6027 |
| | $10K | 0.3978 | 0.4361 | 0.4541 | 0.4776 | 0.5123 |
| | $1K | 0.1871 | 0.2510 | 0.3418 | 0.4266 | 0.4754 |
| Between 37.7 and 50.7 | $100K | 0.4527 | 0.4963 | 0.5258 | 0.5552 | 0.6275 |
| | $10K | 0.4203 | 0.4524 | 0.4721 | 0.4996 | 0.5387 |
| | $1K | 0.3893 | 0.4150 | 0.4327 | 0.4512 | 0.4738 |
| Between 31 and 37.7 | $100K | 0.5028 | 0.5245 | 0.5493 | 0.6005 | 0.6996 |
| | $10K | 0.4504 | 0.4782 | 0.4907 | 0.5184 | 0.5536 |
| | $1K | 0.4228 | 0.4374 | 0.4478 | 0.4719 | 0.5128 |
| Lesser than 31 | $100K | 0.5163 | 0.5637 | 0.6148 | 0.6372 | 0.7194 |
| | $10K | 0.4673 | 0.4929 | 0.5269 | 0.5479 | 0.5935 |
| | $1K | 0.4291 | 0.4478 | 0.4728 | 0.4868 | 0.5170 |

***Region**: G1:Central Europe, Eastern Europe, and Central Asia; G2:High income; G3:Latin America and Caribbean; G4:North Africa and Middle East; G5: Southeast Asia, East Asia, and Oceania; G6:Sub-Saharan Africa.
**Population**: G1:more than 100M, G2:30M-100M, G3:10M-30M, G4:5M-10M.
**Income**: G1:more than $17K, G2:$7K-$17K, G3:$2K-$7K, G4:less than $2K.
**UHC**: G1:more than 72, G2:59.3-72, G3:47.9-59.3, G4:less than 47.9.
**SDI**: G1:more than 0.76, G2:0.64-0.76, G3:0.46-0.64, G4:less than 0.46.
**GHS**: G1:more than 50.7, G2:37.7-50.7, G3:31-37.7, G4:less than 31.

**Table S1.11. Generalised cost-optimal intervention results for 52 weeks vaccine development timing (corresponds to Figure 4 in main manuscript).**

**Region**

| Category | Cost per infection | Cost-optimal average transmission reduction Min | 1Q | Median | 3Q | Max |
|---|---|---|---|---|---|---|
| High-income | $100K | 0.4698 | 0.5055 | 0.519 | 0.5442 | 0.5732 |
| | $10K | 0.4452 | 0.4708 | 0.4868 | 0.4994 | 0.5271 |
| | $1K | 0.1611 | 0.1726 | 0.2088 | 0.2363 | 0.3062 |
| Central Europe, Eastern Europe, and Central Asia | $100K | 0.5267 | 0.5618 | 0.5703 | 0.5954 | 0.62 |
| | $10K | 0.4825 | 0.5107 | 0.5177 | 0.5321 | 0.5517 |
| | $1K | 0.4055 | 0.4592 | 0.4848 | 0.497 | 0.5491 |
| North Africa and Middle East | $100K | 0.5095 | 0.5397 | 0.5693 | 0.5933 | 0.6475 |
| | $10K | 0.4783 | 0.4973 | 0.5114 | 0.5314 | 0.5691 |
| | $1K | 0.4507 | 0.4576 | 0.4738 | 0.4911 | 0.5153 |
| Latin America and Caribbean | $100K | 0.5213 | 0.5495 | 0.5828 | 0.6051 | 0.6596 |
| | $10K | 0.4856 | 0.5082 | 0.526 | 0.5401 | 0.5753 |
| | $1K | 0.4334 | 0.4664 | 0.4755 | 0.4973 | 0.5223 |
| Southeast Asia, East Asia, and Oceania | $100K | 0.4889 | 0.4928 | 0.5368 | 0.5526 | 0.5618 |
| | $10K | 0.4582 | 0.4664 | 0.4934 | 0.5014 | 0.5155 |
| | $1K | 0.4365 | 0.4443 | 0.453 | 0.4674 | 0.472 |
| Sub-Saharan Africa | $100K | 0.4974 | 0.5576 | 0.6186 | 0.6536 | 0.7202 |
| | $10K | 0.4692 | 0.5109 | 0.5465 | 0.5716 | 0.6087 |
| | $1K | 0.4446 | 0.4767 | 0.5034 | 0.5177 | 0.5376 |

**Population**

| Category | Cost per infection | Cost-optimal average transmission reduction Min | 1Q | Median | 3Q | Max |
|---|---|---|---|---|---|---|
| More than 100M | $100K | 0.4698 | 0.4951 | 0.5352 | 0.5535 | 0.6161 |
| | $10K | 0.4452 | 0.4653 | 0.4935 | 0.5059 | 0.5455 |
| | $1K | 0.4365 | 0.4421 | 0.4618 | 0.4728 | 0.5071 |
| Between 30M and 100M | $100K | 0.4722 | 0.5146 | 0.5501 | 0.5674 | 0.6408 |
| | $10K | 0.4481 | 0.4817 | 0.5028 | 0.5161 | 0.5593 |
| | $1K | 0.3553 | 0.4152 | 0.4693 | 0.4826 | 0.5153 |
| Between 10M and 30M | $100K | 0.5095 | 0.5565 | 0.5915 | 0.6524 | 0.726 |
| | $10K | 0.4783 | 0.5116 | 0.5286 | 0.5709 | 0.6115 |
| | $1K | 0.3869 | 0.457 | 0.4884 | 0.5143 | 0.5491 |
| Lesser than 10M | $100K | 0.5273 | 0.5618 | 0.5894 | 0.62 | 0.6767 |
| | $10K | 0.493 | 0.5133 | 0.5297 | 0.5497 | 0.6001 |
| | $1K | 0.2363 | 0.396 | 0.4904 | 0.5042 | 0.5412 |

**Income**

| Category | Cost per infection | Cost-optimal average transmission reduction Min | 1Q | Median | 3Q | Max |
|---|---|---|---|---|---|---|
| More than $17K | $100K | 0.4698 | 0.506 | 0.5235 | 0.5479 | 0.5931 |
| | $10K | 0.4452 | 0.4714 | 0.4908 | 0.5041 | 0.5287 |
| | $1K | 0.1611 | 0.1726 | 0.2134 | 0.2897 | 0.4412 |
| Between $7K and $17K | $100K | 0.4889 | 0.5272 | 0.5596 | 0.5851 | 0.62 |
| | $10K | 0.4582 | 0.4872 | 0.5142 | 0.5279 | 0.5517 |
| | $1K | 0.4055 | 0.4527 | 0.472 | 0.4853 | 0.5022 |
| Between $2K and $7K | $100K | 0.4921 | 0.5397 | 0.556 | 0.5828 | 0.6382 |
| | $10K | 0.4585 | 0.4982 | 0.5076 | 0.526 | 0.5636 |
| | $1K | 0.4365 | 0.464 | 0.4731 | 0.4861 | 0.5105 |
| Lesser than $2K | $100K | 0.4974 | 0.5822 | 0.6263 | 0.6547 | 0.726 |
| | $10K | 0.4692 | 0.5226 | 0.552 | 0.5722 | 0.6115 |
| | $1K | 0.4446 | 0.4886 | 0.5071 | 0.5184 | 0.5491 |

**UHC**

| Category | Cost per infection | Cost-optimal average transmission reduction Min | 1Q | Median | 3Q | Max |
|---|---|---|---|---|---|---|
| More than 72 | $100K | 0.4698 | 0.5076 | 0.5344 | 0.5661 | 0.6002 |
| | $10K | 0.4452 | 0.4726 | 0.4972 | 0.5159 | 0.536 |
| | $1K | 0.1611 | 0.1968 | 0.2542 | 0.4373 | 0.4924 |
| Between 59.3 and 72 | $100K | 0.4889 | 0.5269 | 0.5576 | 0.588 | 0.6567 |
| | $10K | 0.4582 | 0.4861 | 0.5113 | 0.5292 | 0.5765 |
| | $1K | 0.4365 | 0.4516 | 0.472 | 0.4887 | 0.5232 |
| Between 47.9 and 59.3 | $100K | 0.5362 | 0.5514 | 0.5853 | 0.631 | 0.726 |
| | $10K | 0.492 | 0.5031 | 0.5254 | 0.5531 | 0.6115 |
| | $1K | 0.4513 | 0.472 | 0.4884 | 0.5082 | 0.5491 |
| Lesser than 47.9 | $100K | 0.4974 | 0.558 | 0.6161 | 0.6539 | 0.7202 |
| | $10K | 0.4692 | 0.5103 | 0.5455 | 0.5713 | 0.6087 |
| | $1K | 0.4446 | 0.475 | 0.5038 | 0.5176 | 0.5376 |

**SDI**

| Category | Cost per infection | Cost-optimal average transmission reduction Min | 1Q | Median | 3Q | Max |
|---|---|---|---|---|---|---|
| More than 0.76 | $100K | 0.4698 | 0.5079 | 0.5314 | 0.564 | 0.62 |
| | $10K | 0.4452 | 0.4733 | 0.493 | 0.5152 | 0.5517 |
| | $1K | 0.1611 | 0.1904 | 0.2363 | 0.4353 | 0.5022 |
| Between 0.64 and 0.76 | $100K | 0.4889 | 0.5342 | 0.5596 | 0.5749 | 0.6002 |
| | $10K | 0.4582 | 0.4935 | 0.5107 | 0.5217 | 0.536 |
| | $1K | 0.4395 | 0.4548 | 0.472 | 0.482 | 0.4924 |
| Between 0.46 and 0.64 | $100K | 0.4921 | 0.55 | 0.5643 | 0.6263 | 0.726 |
| | $10K | 0.4585 | 0.5007 | 0.5127 | 0.5472 | 0.6115 |
| | $1K | 0.4365 | 0.4667 | 0.4767 | 0.5075 | 0.5491 |
| Lesser than 0.46 | $100K | 0.544 | 0.5919 | 0.6277 | 0.6557 | 0.7202 |
| | $10K | 0.4968 | 0.5316 | 0.5535 | 0.5728 | 0.6087 |
| | $1K | 0.4693 | 0.4927 | 0.5071 | 0.5186 | 0.5376 |

**GHS**

| Category | Cost per infection | Cost-optimal average transmission reduction Min | 1Q | Median | 3Q | Max |
|---|---|---|---|---|---|---|
| More than 50.7 | $100K | 0.4698 | 0.5066 | 0.5314 | 0.5591 | 0.62 |
| | $10K | 0.4452 | 0.4713 | 0.493 | 0.5144 | 0.5517 |
| | $1K | 0.1611 | 0.1977 | 0.2722 | 0.4452 | 0.5022 |
| Between 37.7 and 50.7 | $100K | 0.4921 | 0.5289 | 0.5536 | 0.5834 | 0.6382 |
| | $10K | 0.4582 | 0.4888 | 0.5057 | 0.5269 | 0.5636 |
| | $1K | 0.4365 | 0.4509 | 0.4694 | 0.4842 | 0.5105 |
| Between 31 and 37.7 | $100K | 0.5341 | 0.5558 | 0.5755 | 0.6191 | 0.6767 |
| | $10K | 0.4876 | 0.5077 | 0.5209 | 0.546 | 0.5772 |
| | $1K | 0.4546 | 0.4724 | 0.4861 | 0.5009 | 0.5376 |
| Lesser than 31 | $100K | 0.5378 | 0.5868 | 0.6219 | 0.6535 | 0.726 |
| | $10K | 0.495 | 0.5251 | 0.5496 | 0.5719 | 0.6115 |
| | $1K | 0.4556 | 0.4843 | 0.5071 | 0.5176 | 0.5491 |

***Region**: G1:Central Europe, Eastern Europe, and Central Asia; G2:High income; G3:Latin America and Caribbean; G4:North Africa and Middle East; G5: Southeast Asia, East Asia, and Oceania; G6:Sub-Saharan Africa.
**Population**: G1:more than 100M, G2:30M-100M, G3:10M-30M, G4:5M-10M.
**Income**: G1:more than $17K, G2:$7K-$17K, G3:$2K-$7K, G4:less than $2K.
**UHC**: G1:more than 72, G2:59.3-72, G3:47.9-59.3, G4:less than 47.9.
**SDI**: G1:more than 0.76, G2:0.64-0.76, G3:0.46-0.64, G4:less than 0.46.
**GHS**: G1:more than 50.7, G2:37.7-50.7, G3:31-37.7, G4:less than 31.
