## Supplementary file 2 for "Cost-optimal transmission reduction in early pandemic response: a multi-objective health-economic modelling study across 110 countries"

### Supplementary 2. Structural and practical identifiability

We analyse the structural identifiability of the mathematical model in the main manuscript using the StructuralIdentifiability.jl package, and the practical identifiability of the mathematical model in the main manuscript using the joint posterior distribution obtained by MCMC sampling.

#### 2.1. Structural identifiability analysis

This appendix provides the Julia code and results for the structural identifiability analysis of the SEIQRD model described in the main text. The analysis was performed using the StructuralIdentifiability.jl package [1,2]. The code and output are presented for reproducibility. The structural identifiability analysis applies to a simplified version of the model in which the transmission reduction is treated as a single constant parameter $\mu$ rather than the piecewise time-varying sequence $\mu_3, \mu_4, \cdots, \mu_k$ used for estimation in the main manuscript. Treating $\mu$ as constant gives a conservative test of identifiability, because the time-varying formulation only adds further constraints from the observed trajectory. Identifiability of the full time-varying sequence is assessed empirically through the practical identifiability analysis below, using the joint posterior distributions obtained by MCMC sampling, which show well-constrained, unimodal marginals for the estimated transmission-reduction parameters.

The SEIQRD model with importation is specified as an ODE system with seven parameters $(R0, \kappa, \alpha, f, \gamma, \mu, \xi)$ and six state variables $(S, E, I, Q, R, D)$. One output, daily confirmed cases $(\alpha I)$, is assumed observable.

*Scenario 1: Identifiability with observable only*. This scenario tests whether all parameters and states can be uniquely determined from observed outputs alone, without knowledge of initial conditions.

*Scenario 2: Identifiability with observable and known initial conditions*. This scenario additionally assumes that all initial conditions $(S(0), E(0), I(0), Q(0), R(0), D(0))$ are known. In our model, at the start of the pandemic, $S(0)$ equals the total population, and all other compartments are zero. Only the imported cases $\xi$ from abroad initiate the pandemic.

*Scenario 3: Identifiability with observable, known initial conditions, and reproduction number*. This scenario further assumes that the reproduction number is known from the epidemiological paper for the disease.

**Table S2.1. Structural identifiability results for three scenarios.**

| Compartment / Parameter | Scenario 1 (observable only) | Scenario 2 (observable + known initial conditions) | Scenario 3 (observable + known initial conditions + $R_0$) |
|---|---|---|---|
| $S(t)$ | Globally identifiable | Globally identifiable | Globally identifiable |
| $E(t)$ | Locally identifiable | Globally identifiable | Globally identifiable |
| $I(t)$ | Locally identifiable | Globally identifiable | Globally identifiable |
| $Q(t)$ | Globally identifiable | Globally identifiable | Globally identifiable |

| | | | |
|---|---|---|---|
| $R(t)$ | Globally identifiable | Globally identifiable | Globally identifiable |
| $D(t)$ | Non-identifiable | Globally identifiable | Globally identifiable |
| $R_0$ | Non-identifiable | Non-identifiable | Globally identifiable |
| $\kappa$ | Locally identifiable | Globally identifiable | Globally identifiable |
| $\alpha$ | Locally identifiable | Globally identifiable | Globally identifiable |
| $\gamma$ | Globally identifiable | Globally identifiable | Globally identifiable |
| $\xi$ | Globally identifiable | Globally identifiable | Globally identifiable |
| $f$ | Globally identifiable | Globally identifiable | Globally identifiable |
| $\mu$ | Non-identifiable | Non-identifiable | Globally identifiable |

With observable alone (Scenario 1), $R_0$ and $\mu$ are non-identifiable because they enter the force of infection as a product $(1-\mu)R_0$, making them individually indistinguishable. When initial conditions are known (Scenario 2), the latent states become globally identifiable, but $R_0$ and $\mu$ remain non-identifiable. Only when $R_0$ is additionally fixed to a value (Scenario 3) do all parameters become globally identifiable. This justifies our assumption that researchers can estimate the disease's reproduction number using case data from the first-outbreak country, thereby resolving the identifiability of the full parameter set.

**Julia code (SEIQRD_imported.jl)**

```
# Jongmin Lee, Gerardo Chowell, Renier Mendoza, Victoria May P. Mendoza, and Eunok Jung
# Structural identifiability analysis of the SEIQRD model
# with importation for infectious disease transmission

using StructuralIdentifiability

ode = @ODEmodel(
   s'(t) = -(1-mu)*R0*alpha*s(t)*i(t)/(s(t)+e(t)+i(t)+r(t)),
   e'(t) = (1-mu)*R0*alpha*s(t)*i(t)/(s(t)+e(t)+i(t)+r(t))
         - kappa*e(t) + xi,
   i'(t) = kappa*e(t) - alpha*i(t),
   q'(t) = alpha*i(t) - gamma*q(t),
   r'(t) = (1-f)*gamma*q(t),
   d'(t) = f*gamma*q(t),
   y1(t) = alpha*i(t)    # daily confirmed cases
)

# Scenario 1: observable only
println("=== Scenario 1: Observable only ===")
@time println(assess_identifiability(ode))

# Scenario 2: observable + known initial conditions
println("=== Scenario 2: Observable + known initial conditions ===")
@time println(assess_identifiability(ode, known_ic=[s, e, i, q, r, d]))
```

```
# Scenario 3: observable + known initial conditions + known R0
println("=== Scenario 3: Observable + known ICs + mu known ===")
@time println(assess_identifiability(ode, known_ic=[s, e, i, q, r, d, R0]))

# Identifiable functions (for diagnostic purposes)
println("=== Identifiable functions ===")
println(find_identifiable_functions(ode, with_states = true))
```

#### 2.2. Practical identifiability

##### 2.2.1. Trace plot and posterior distribution

We conducted Markov Chain Monte Carlo (MCMC) parameter estimation to evaluate parameter uncertainty and obtain posterior distributions using data from the Republic of Korea. The estimation covered a 27-dimensional parameter space, which includes transmission reduction parameters $\mu_j$ ($j \in [3, \cdots, 28]$) and the average imported cases per day $\zeta$. We estimated these parameters using the Delayed Rejection Adaptive Metropolis (DRAM) algorithm.

Most parameters show unimodal, symmetric marginal posteriors on the diagonal, indicating stable convergence. The sharp peaks on the diagonal indicate that the data narrows the prior distribution down to a well-defined posterior. Because these uncertainties are isolated to specific parameters, policy simulations derived from this model for each country can be used with a measurable degree of confidence.

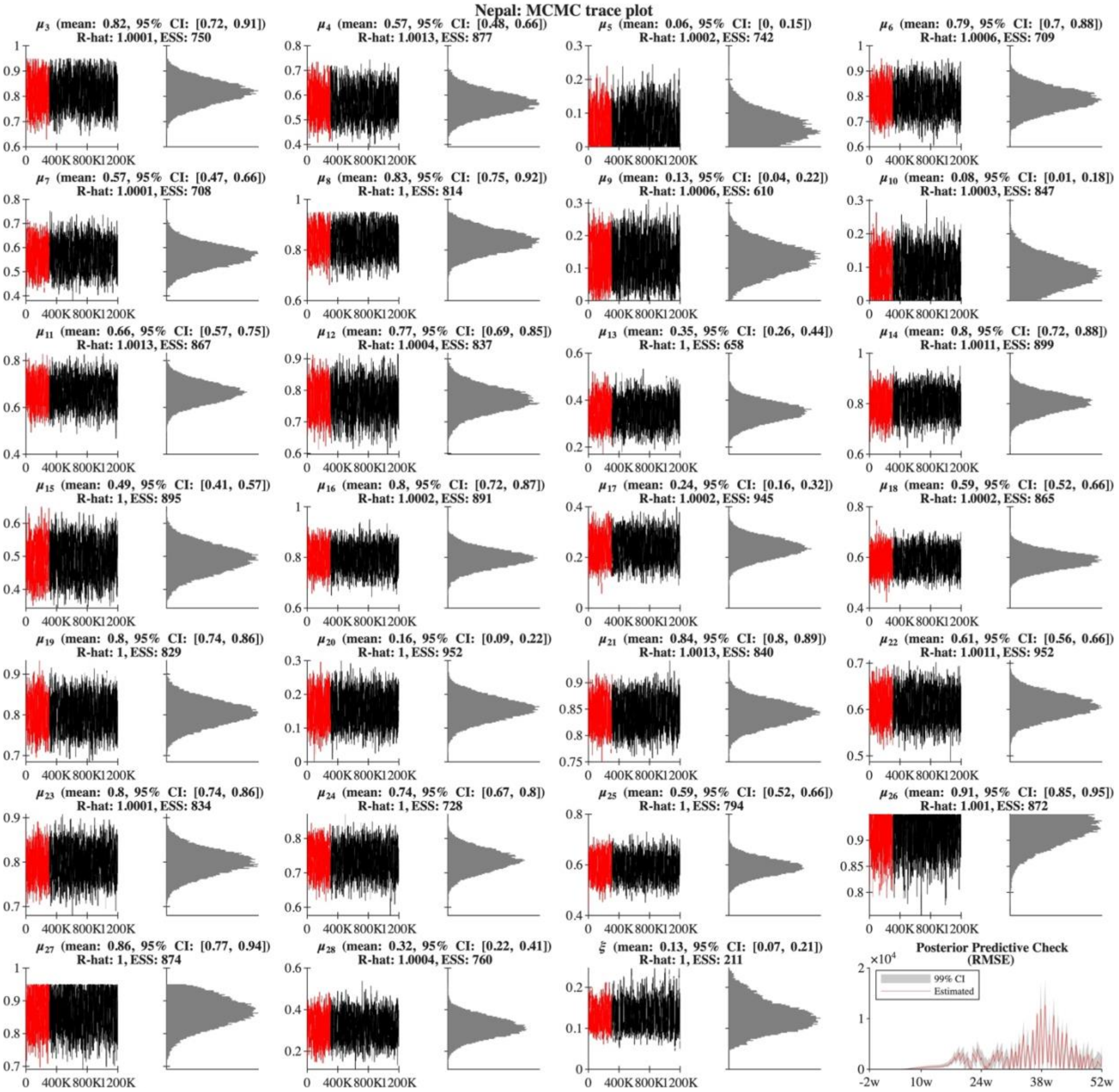


Figure S2.2.1. MCMC parameter estimation and posterior distributions for Nepal.

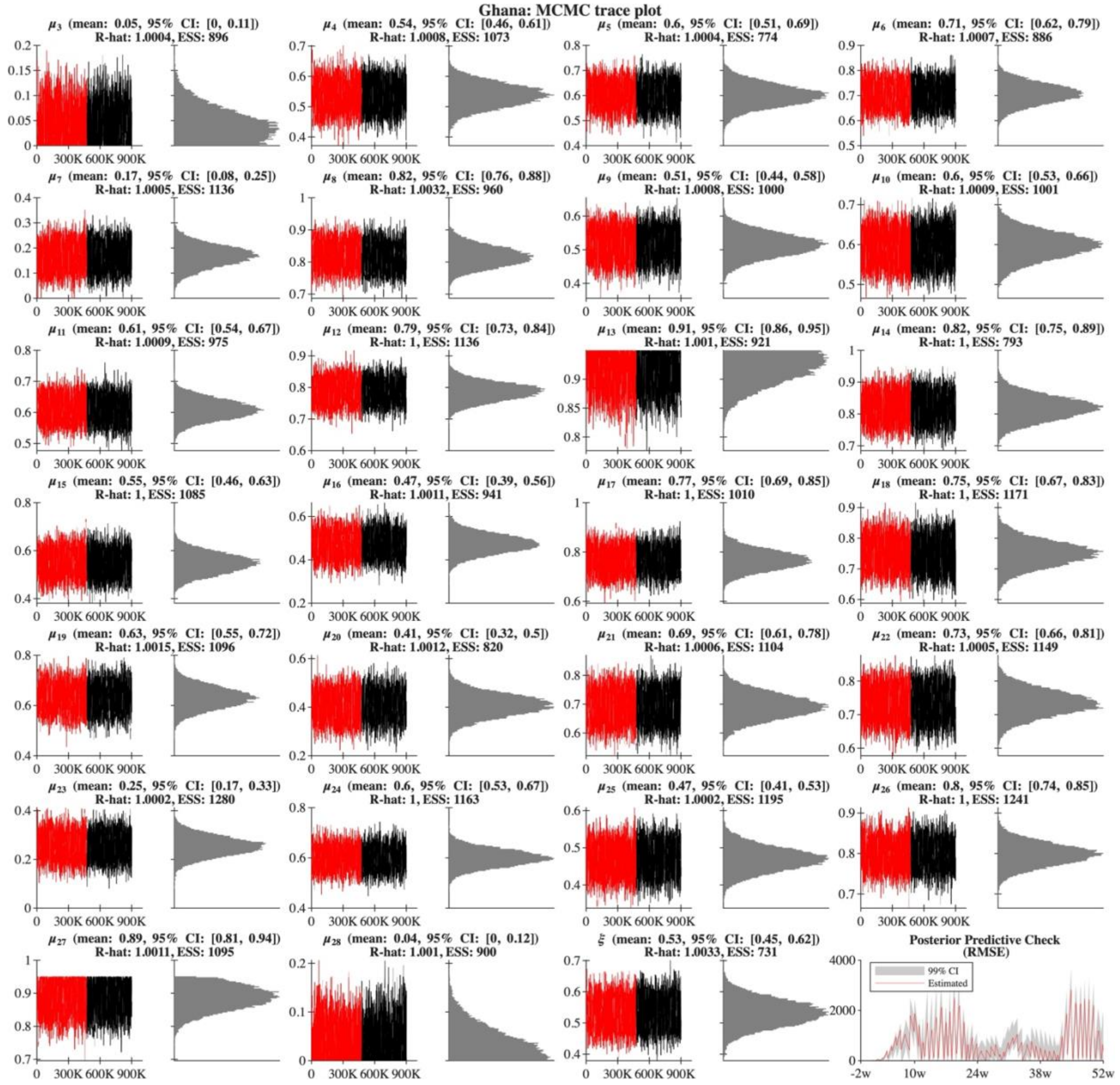


Figure S2.2.2. MCMC parameter estimation and posterior distributions for Ghana.

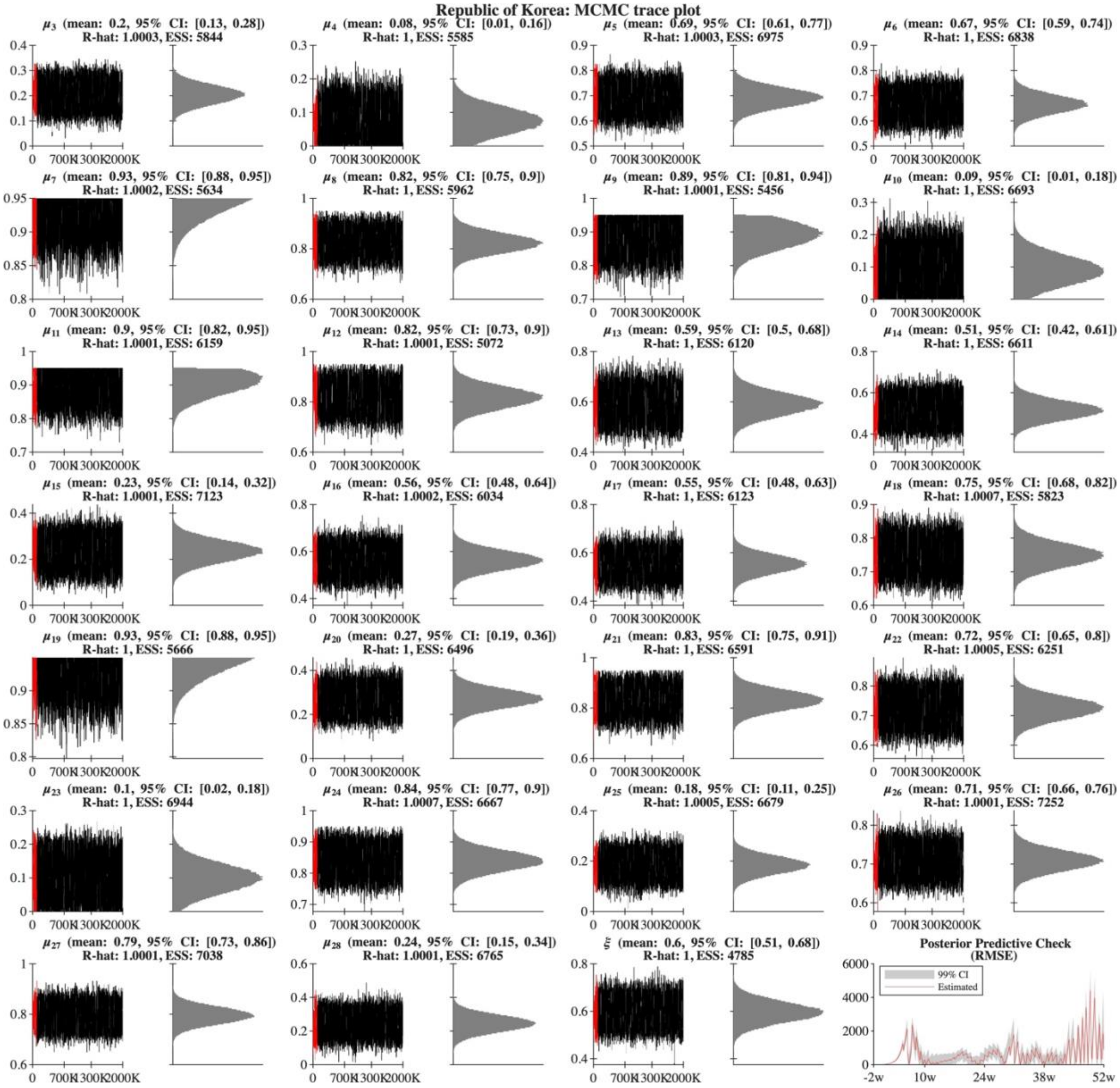


Figure S2.2.3. MCMC parameter estimation and posterior distributions for the Republic of Korea.

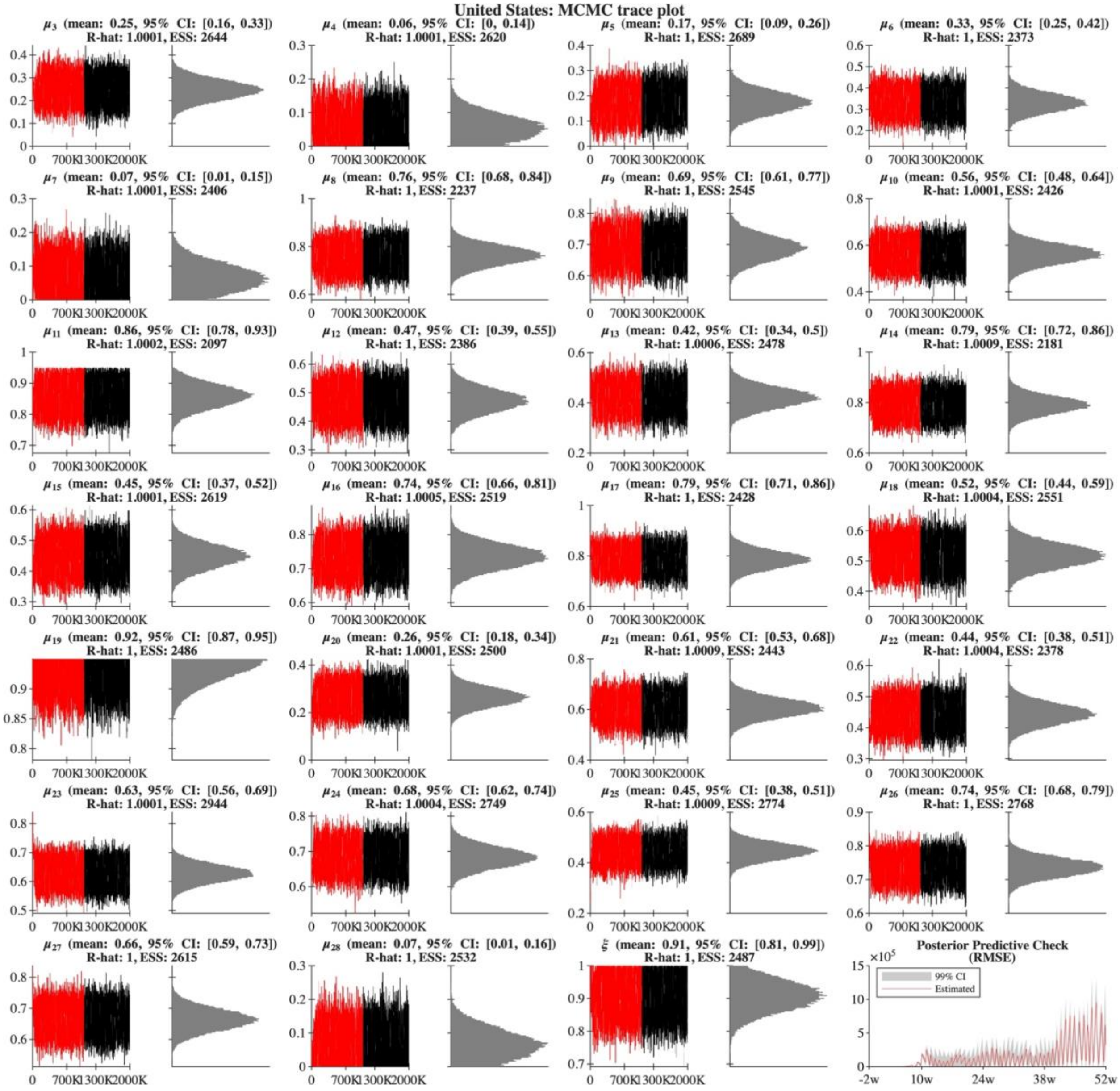


Figure S2.2.4. MCMC parameter estimation and posterior distributions for United States.

#### 2.2.2. Joint and marginal posterior distribution

We conducted MCMC parameter estimation and obtained the posterior distribution of the estimated parameters: transmission reduction $\mu_j$ ($j \in [3, \cdots, 28]$) and average imported cases per day $\xi$. The parameters were estimated using the Delayed Rejection Adaptive Metropolis (DRAM) algorithm [3].

The matrices visualise 27 dimensional parameter spaces estimated from MCMC simulations using data from the Republic of Korea, Nepal, Ghana, and the United States, evaluating parameter uncertainty, identifiability, and mutual dependencies. On the diagonal, most parameters show unimodal, symmetric marginal posteriors, which indicates stable convergence. Off the diagonal, joint posteriors and correlations for most parameter pairs display unstructured scatter plots and low correlation coefficients ($\rho \approx 0$), showing that these parameters are independent. The pairwise correlation coefficients $\rho$ displayed in the upper triangle of the matrices are calculated using the sample correlation coefficient formula. For two parameter sample vectors $x$ and $y$ of length $N$ from the trimmed MCMC chain, the correlation coefficient is defined by the equation

$$\rho = \frac{\sum_{k=1}^{N}(x_k - \bar{x})(y_k - \bar{y})}{\sqrt{\sum_{k=1}^{N}(x_k - \bar{x})^2}\sqrt{\sum_{k=1}^{N}(y_k - \bar{y})^2}}$$

where $\bar{x}$ and $\bar{y}$ are the sample means of the respective parameters. This corresponds to the off-diagonal element *corrMat(1,2)* obtained from the MATLAB *corrcoef* function.

Most of the parameter pairs across all four countries show low mutual correlation, meaning the model is parameterised without widespread collinearity, which indicates that the MCMC chain explored the parameter space without encountering high-dimensional ridges or overfitting. However, the exact structure of parameter dependencies varies notably by country; for instance, the United States exhibits a significantly higher density of strong pairwise correlations (highlighted in red) and more asymmetrical joint posterior distributions across several parameters compared to the more independent parameters of Nepal and Ghana. Across the countries, adjacent transmission parameters $\mu_j$ and $\mu_{j+1}$ tend to have relatively high correlation, and a high correlation is also present between the average imported cases $\xi$ and the initial transmission reduction parameter $\mu_3$. Finally, the sharp peaks on the diagonal show that the data narrows the prior distribution down to a well-defined posterior; because these uncertainties are isolated to specific parameters, policy simulations derived from this model for each country can be used with a measurable degree of confidence.

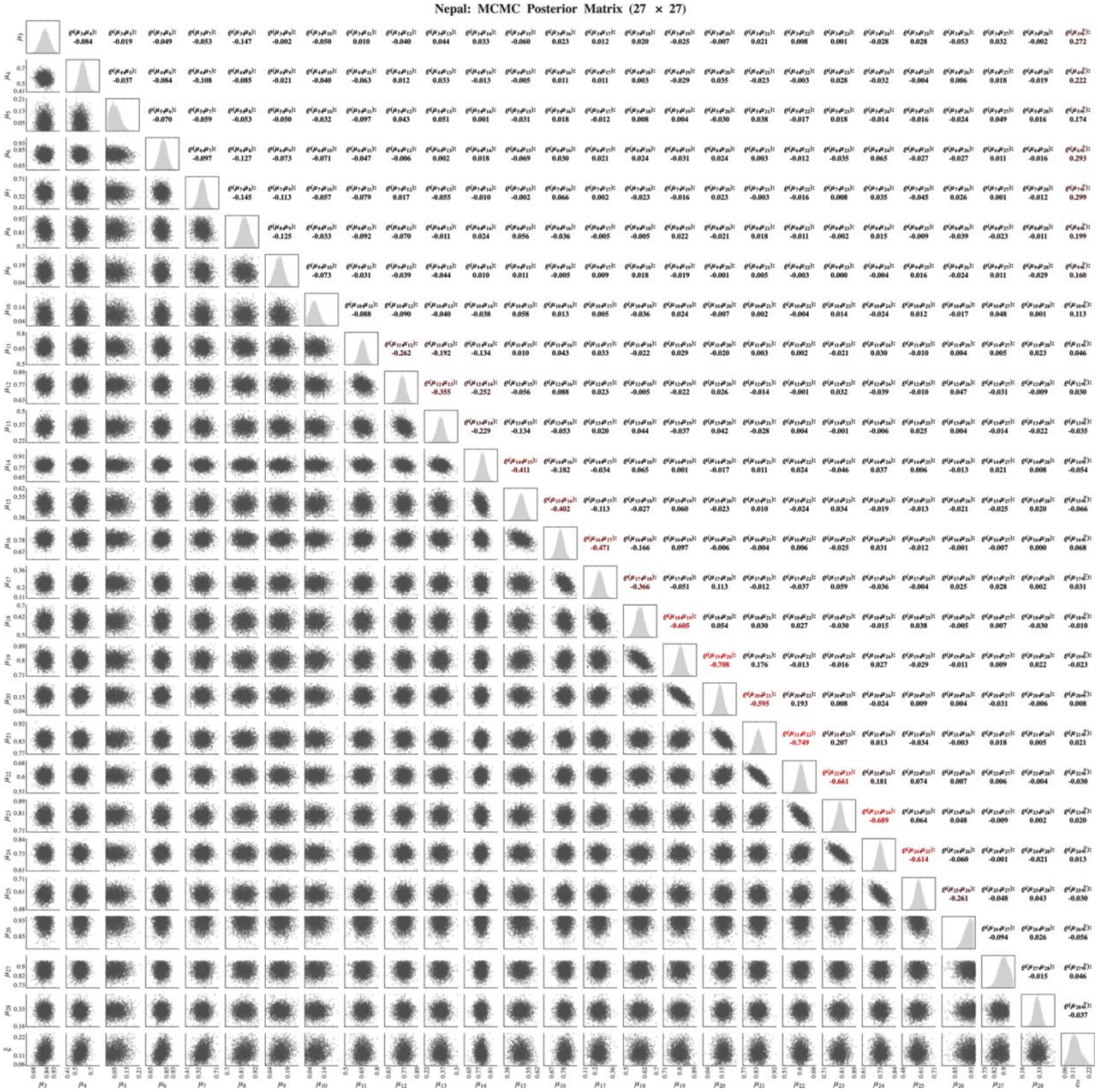


Figure S2.3.1. The joint and marginal posterior distributions of the estimated parameters for Nepal from MCMC sampling.

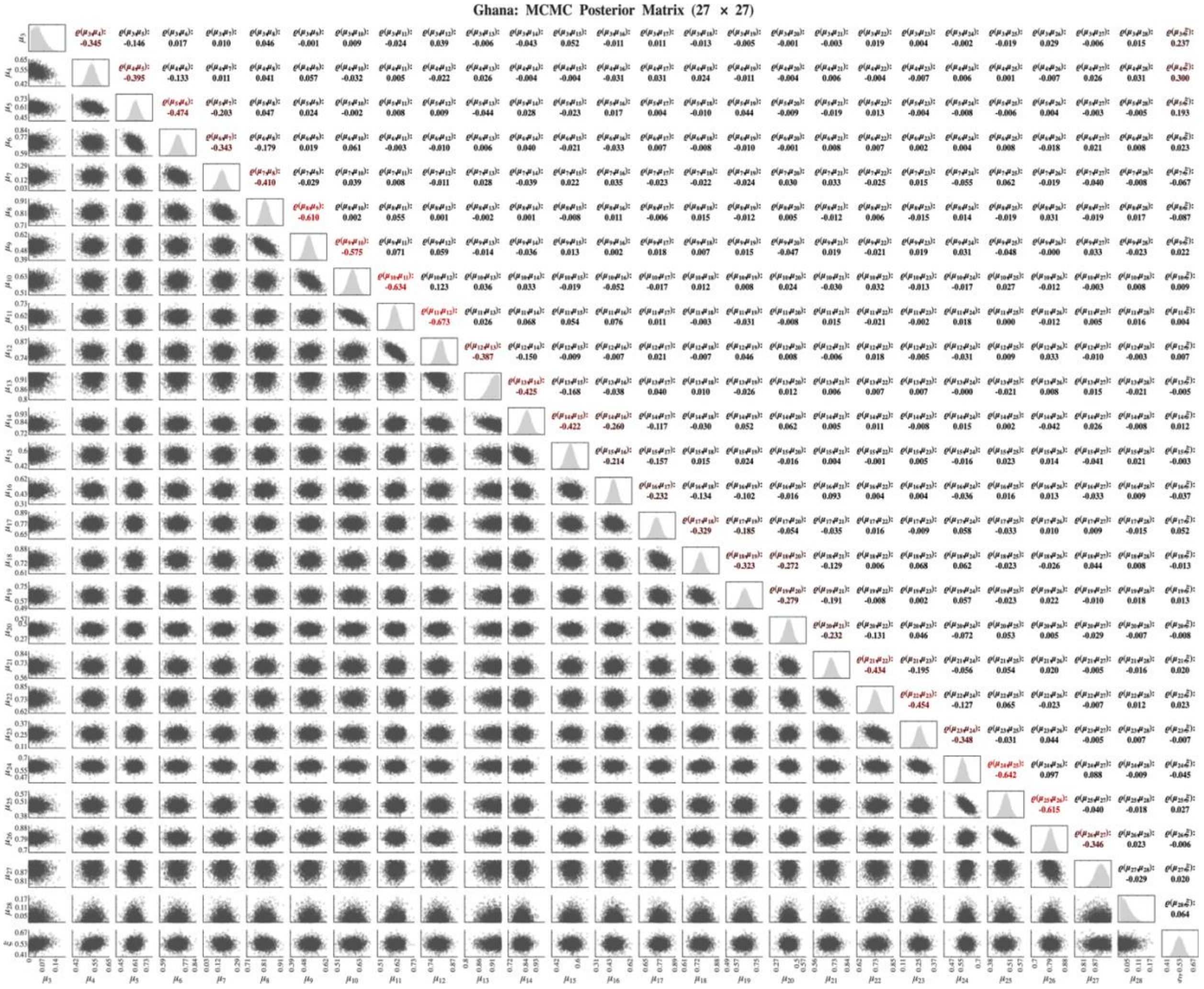


Figure S2.3.2. The joint and marginal posterior distributions of the estimated parameters for Ghana from MCMC sampling.

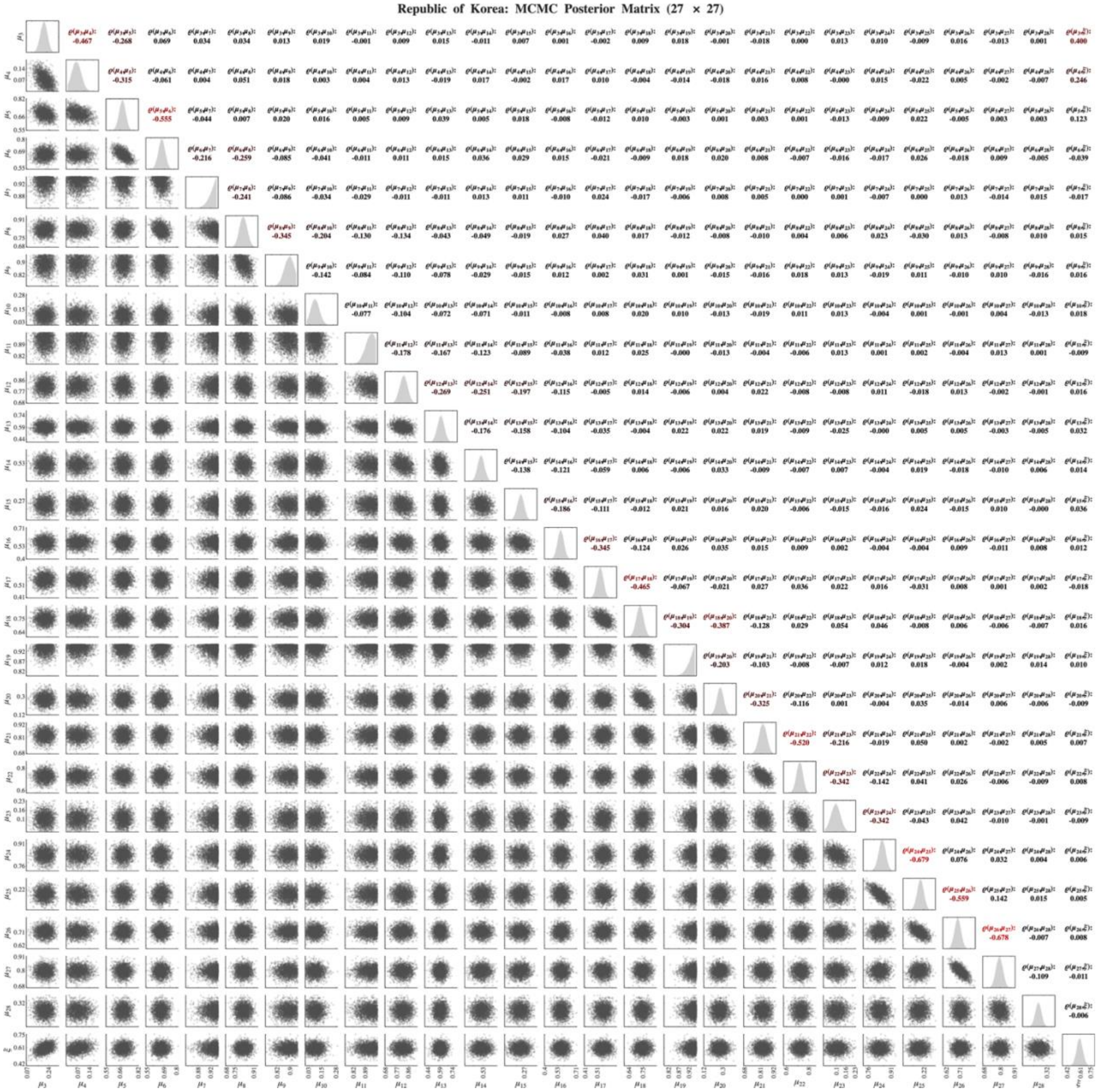


Figure S2.3.3. The joint and marginal posterior distributions of the estimated parameters for the Republic of Korea from MCMC sampling.

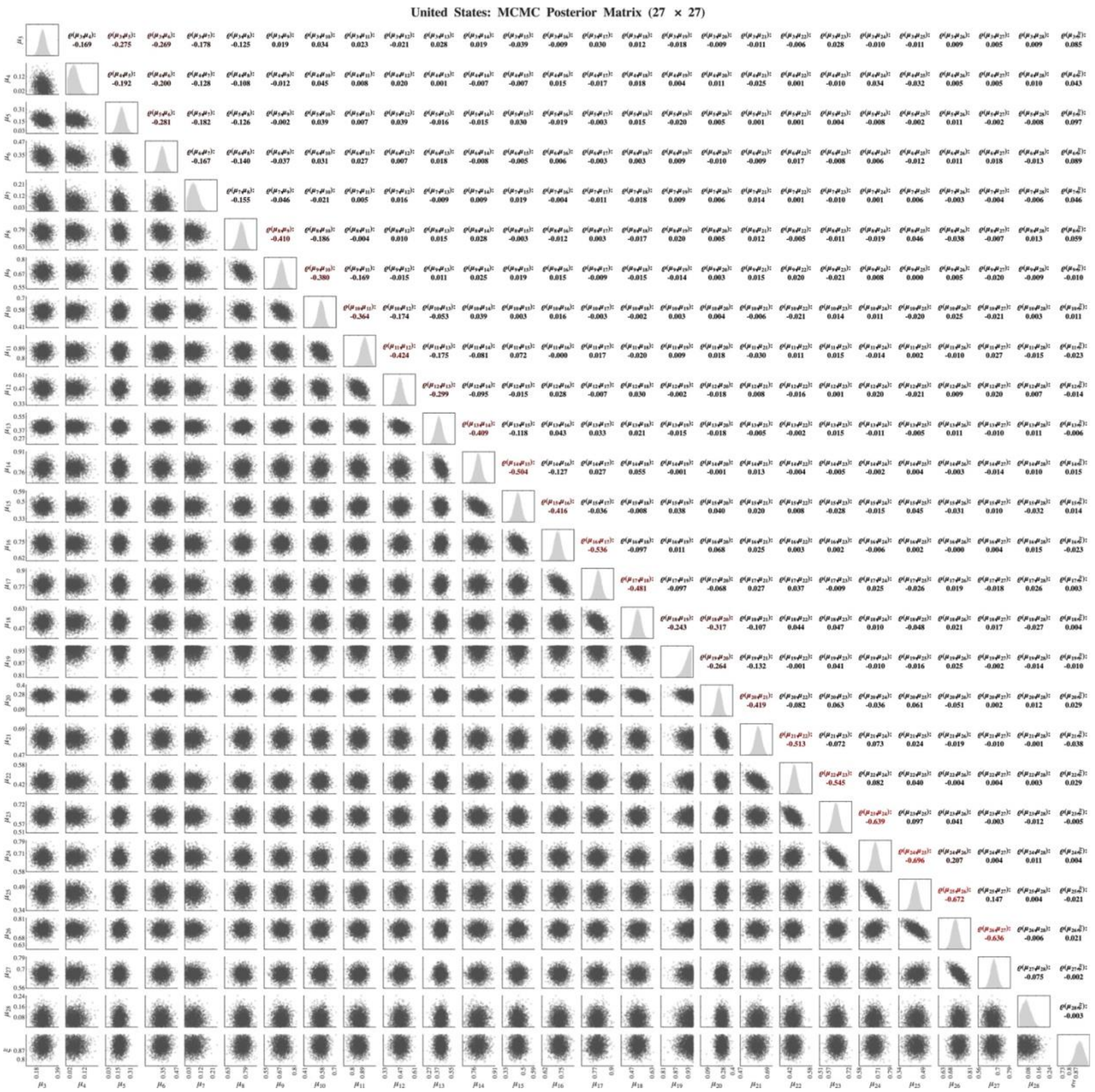


Figure S2.3.4. The joint and marginal posterior distributions of the estimated parameters for the United States from MCMC sampling.
