## Supplementary file 3 for "Cost-optimal transmission reduction in early pandemic response: a multi-objective health-economic modelling study across 110 countries"

### Supplementary 3. Trajectories of transmission reduction and confirmed cases over time for 110 countries

The axes, legend, and simulation parameters correspond to Figure 2. Solid curves and dashed lines represent Pareto frontiers and cost-optimal trajectories for 26, 40, and 52-week simulations. Circles indicate cost-optimal points, and diamonds represent data-fitted results.

Table S3.1 summarises the trajectory patterns across all 110 countries. For each country, the distance between the data-fitted result and the cost-optimal point was computed at 26, 40, and 52 weeks. Countries were classified as Decreasing (distance between cost-optimal and estimated decreases over time, indicating convergence toward the cost-optimal strategy), Increasing (distance between cost-optimal and estimated increases over time, indicating divergence), or Random (no clear monotonic trend). Of the 110 countries, 40 showed a decreasing trend, 37 an increasing trend, and 33 a random pattern. Sub-Saharan Africa (24/30 decreasing) and Southeast Asia, East Asia, and Oceania (8/14 decreasing) predominantly converged toward cost-optimal strategies. By contrast, High-income countries (12/18 increasing) and Central Europe, Eastern Europe, and Central Asia (10/15 increasing) predominantly diverged.

The ten countries with the largest divergence at 52 weeks were Laos (0.4274), Tanzania (0.3747), Cambodia (0.2647), Vietnam (0.2050), Peru (0.2004), United States (0.1981), Mexico (0.1876), Czechia (0.1865), United Kingdom (0.1820), and Italy (0.1780). The ten countries with the greatest convergence (largest reduction in distance from 26 to 52 weeks) were Sri Lanka (0.1448), Thailand (0.1334), Malaysia (0.1322), Burundi (0.1262), Laos (0.1183), Niger (0.1152), China (0.1139), Yemen (0.1065), Cuba (0.1041), and Benin (0.0979).

**Table S3.1: Summary of trajectory classification by GBD super-region.**

| GBD Super-region | Decreasing* | Random* | Increasing* | Total |
|---|---|---|---|---|
| Central Europe, Eastern Europe, Central Asia | 2 | 3 | 10 | 15 |
| High-income | 2 | 4 | 12 | 18 |
| Latin America and Caribbean | 1 | 8 | 8 | 17 |
| North Africa and Middle East | 3 | 6 | 7 | 16 |
| Southeast Asia, East Asia, Oceania | 8 | 6 | 0 | 14 |
| Sub-Saharan Africa | 24 | 6 | 0 | 30 |
| **Total** | **40** | **33** | **37** | **110** |

* Decreasing: distance between cost-optimal and estimated decreases over time. Random: no clear monotonic trend. Increasing: distance between cost-optimal and estimated increases over time.

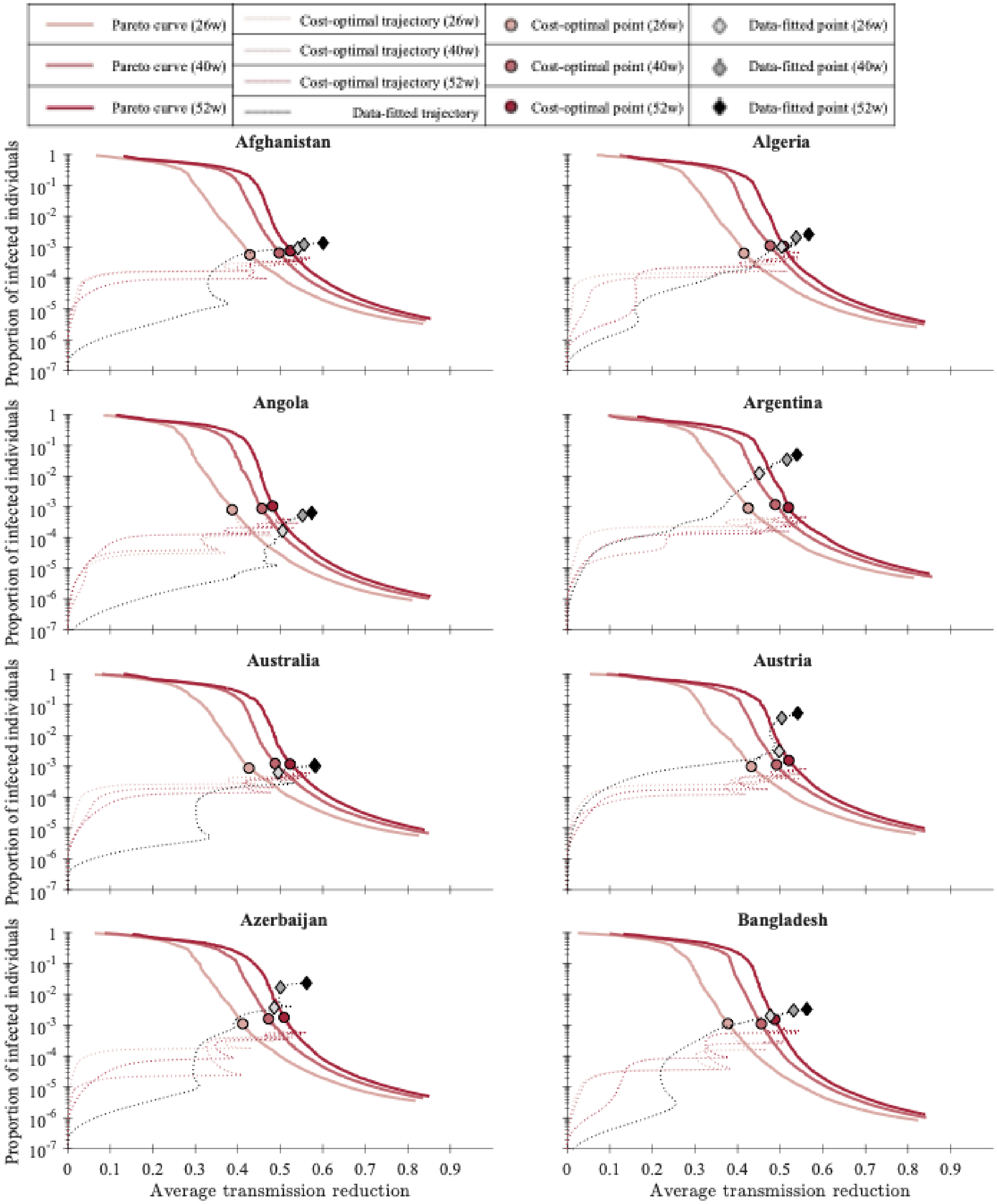


**Figure S3.1: Trajectories of cost-optimal solutions and estimated results for Afghanistan, Algeria, Angola, Argentina, Australia, Austria, Azerbaijan, and Bangladesh.**

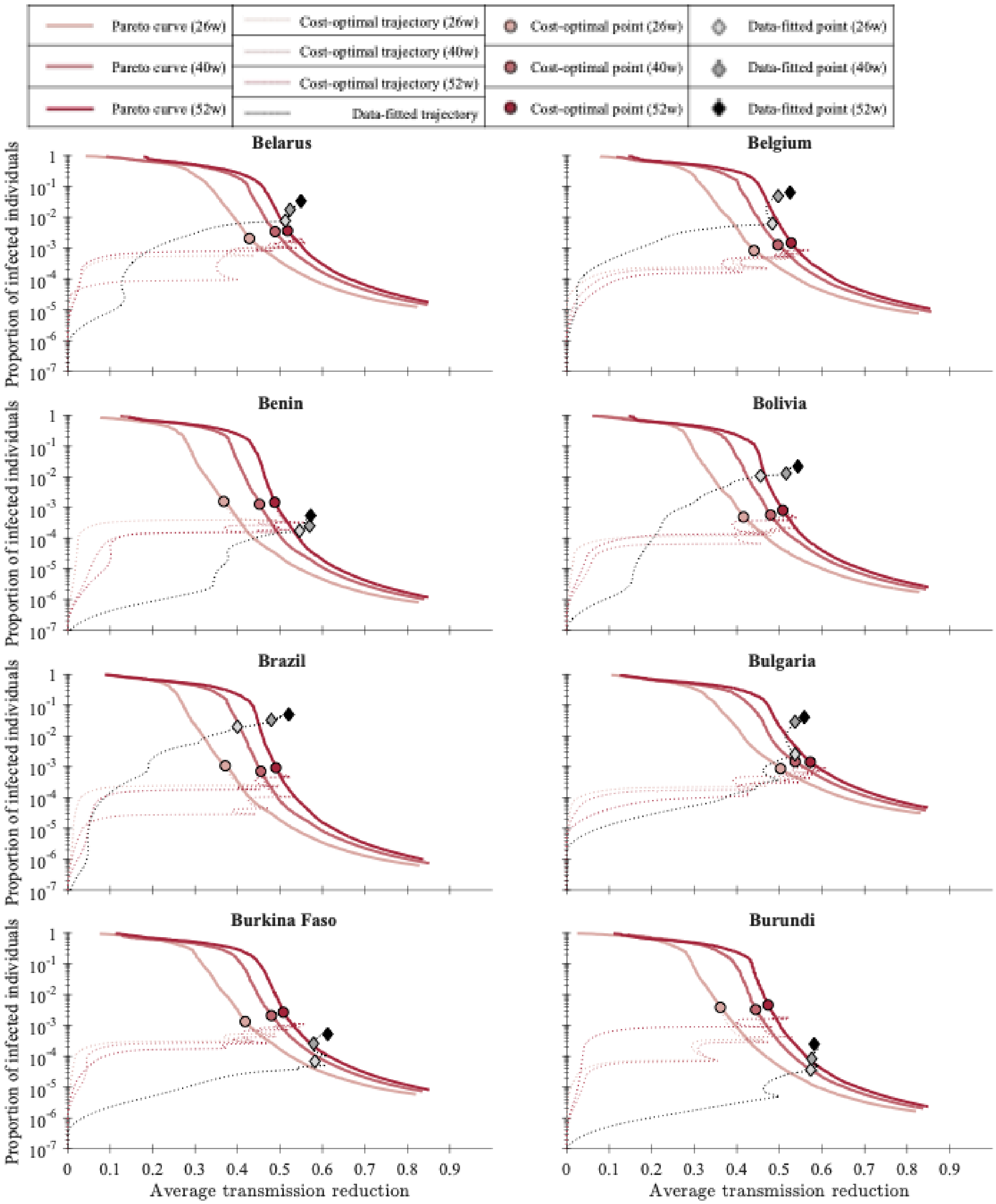


**Figure S3.2: Trajectories of cost-optimal solutions and estimated results for Belarus, Belgium, Benin, Bolivia, Brazil, Bulgaria, Burkina Faso, and Burundi.**

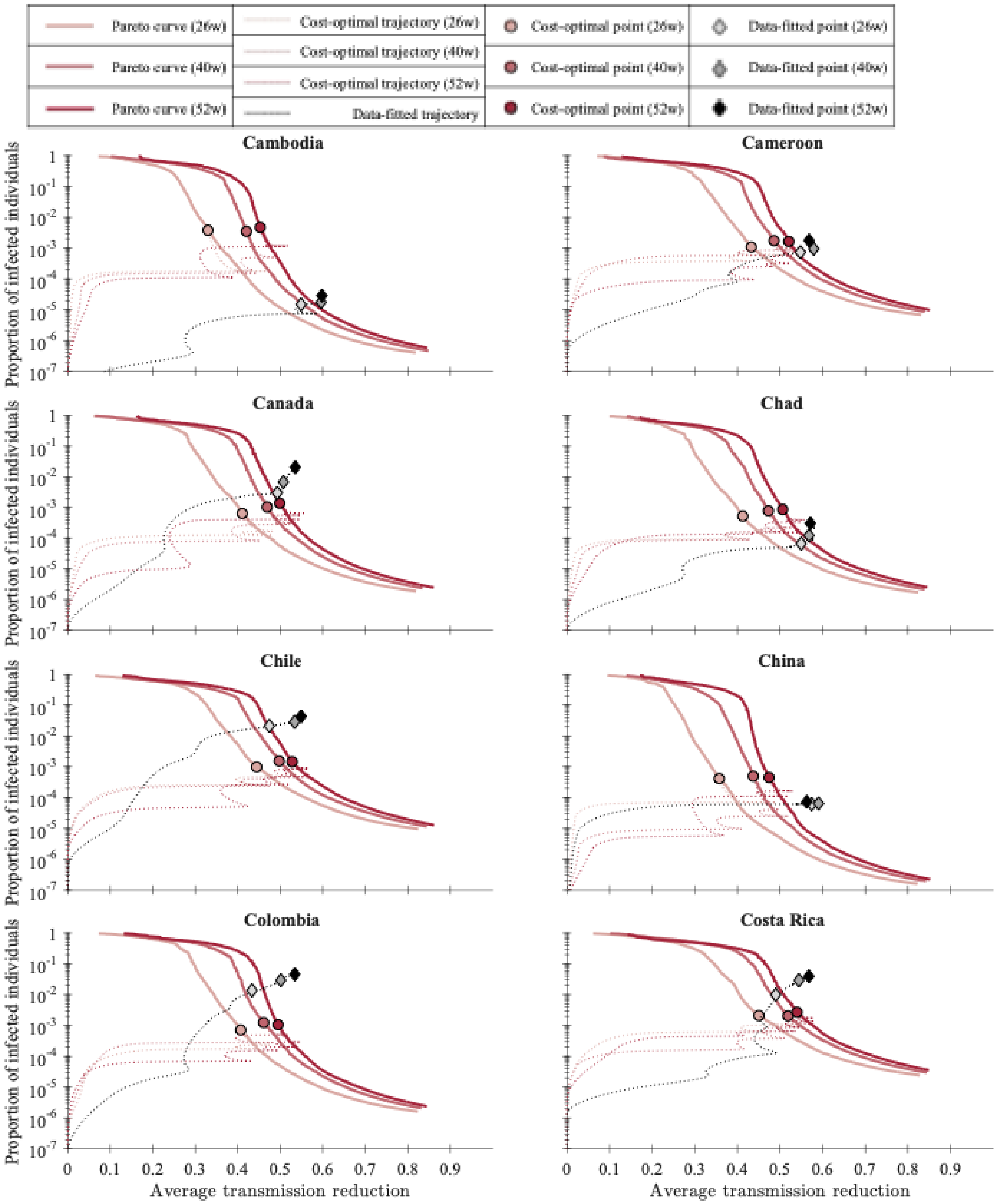


**Figure S3.3: Trajectories of cost-optimal solutions and estimated results for Cambodia, Cameroon, Canada, Chad, Chile, China, Colombia, and Costa Rica.**

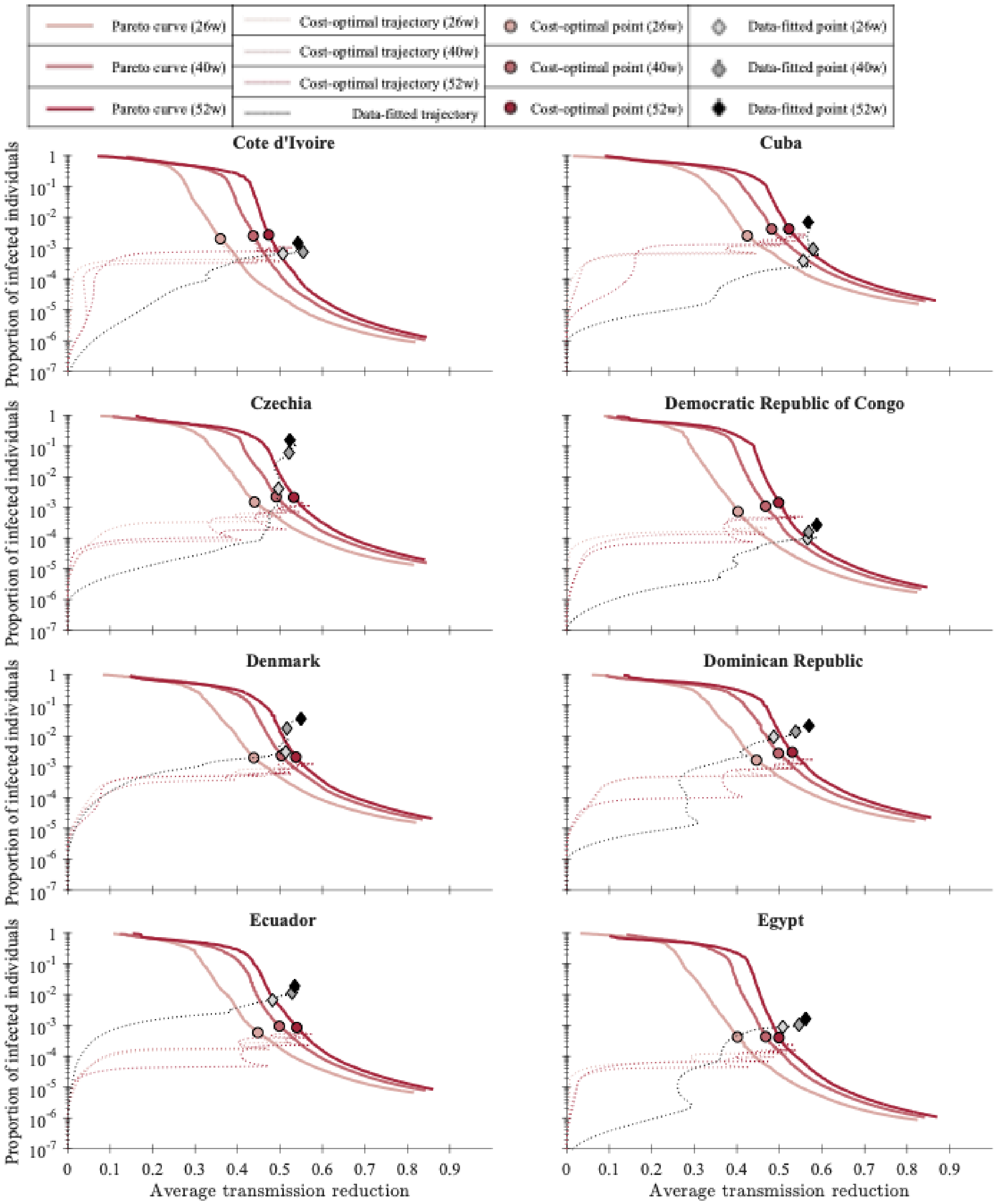


**Figure S3.4: Trajectories of cost-optimal solutions and estimated results for Cote d'Ivoire, Cuba, Czechia, Democratic Republic of Congo, Denmark, Dominican Republic, Ecuador, and Egypt.**

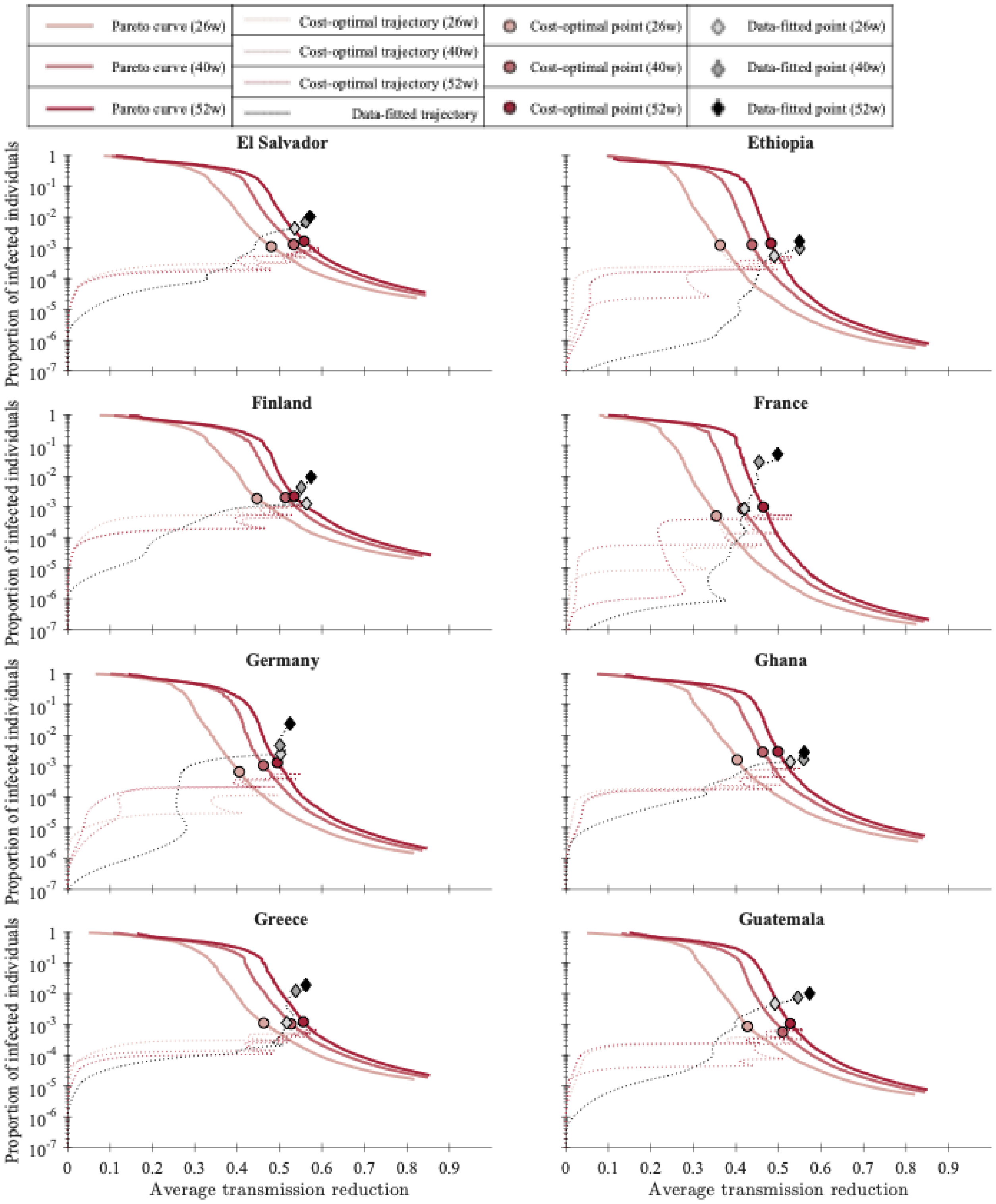


**Figure S3.5: Trajectories of cost-optimal solutions and estimated results for El Salvador, Ethiopia, Finland, France, Germany, Ghana, Greece, and Guatemala.**

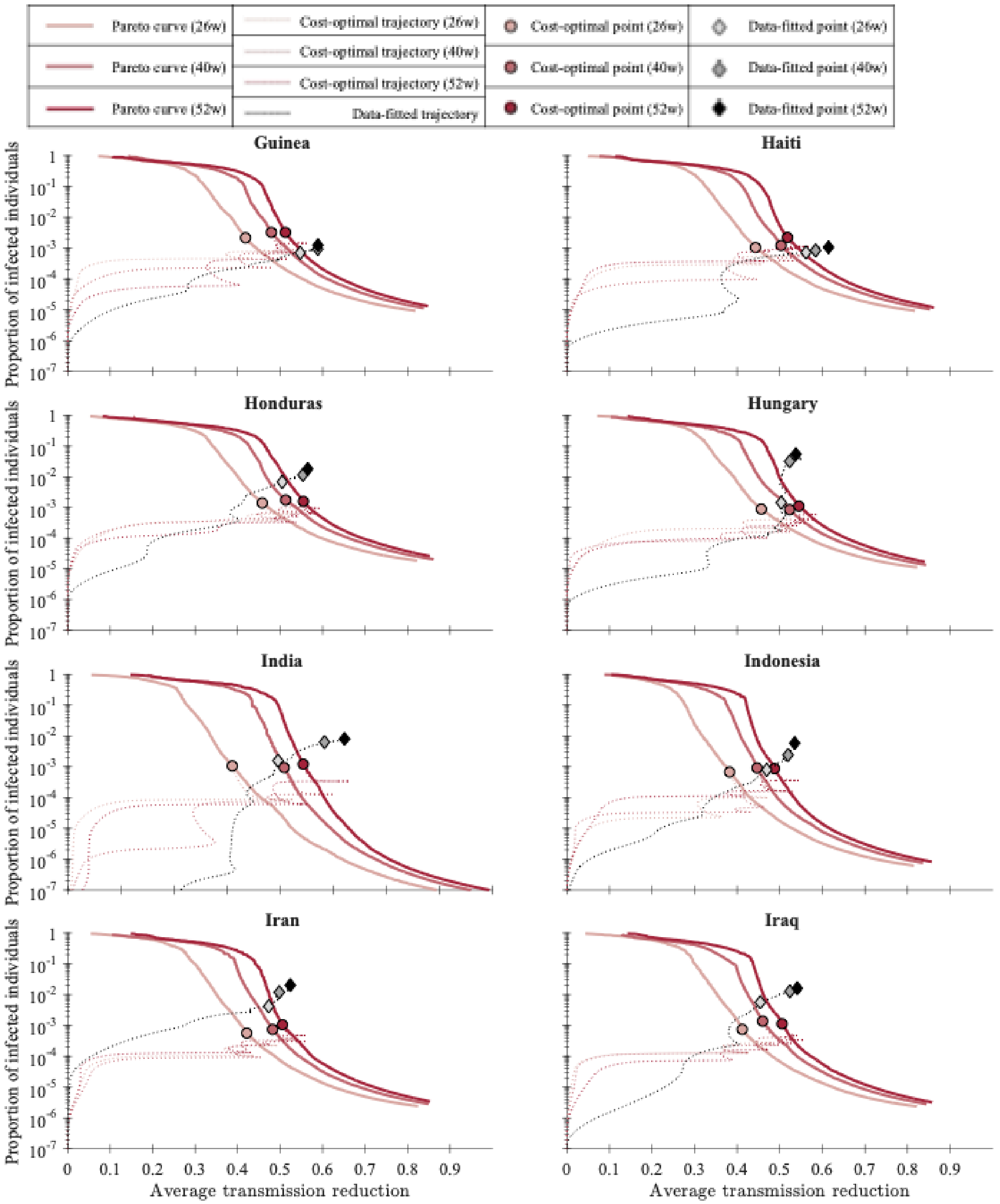


**Figure S3.6: Trajectories of cost-optimal solutions and estimated results for Guinea, Haiti, Honduras, Hungary, India, Indonesia, Iran, and Iraq.**

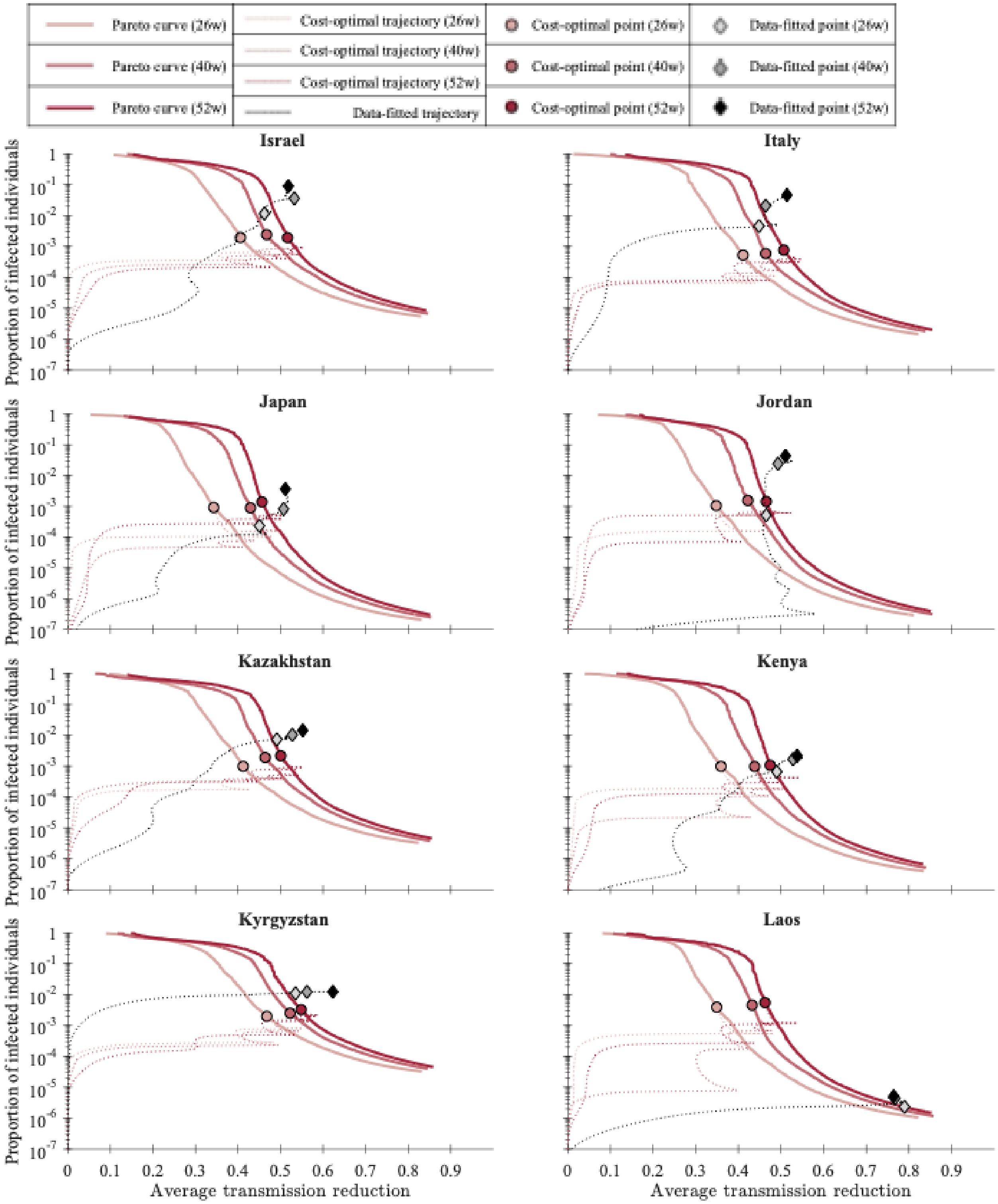


**Figure S3.7: Trajectories of cost-optimal solutions and estimated results for Israel, Italy, Japan, Jordan, Kazakhstan, Kenya, Kyrgyzstan, and Laos.**

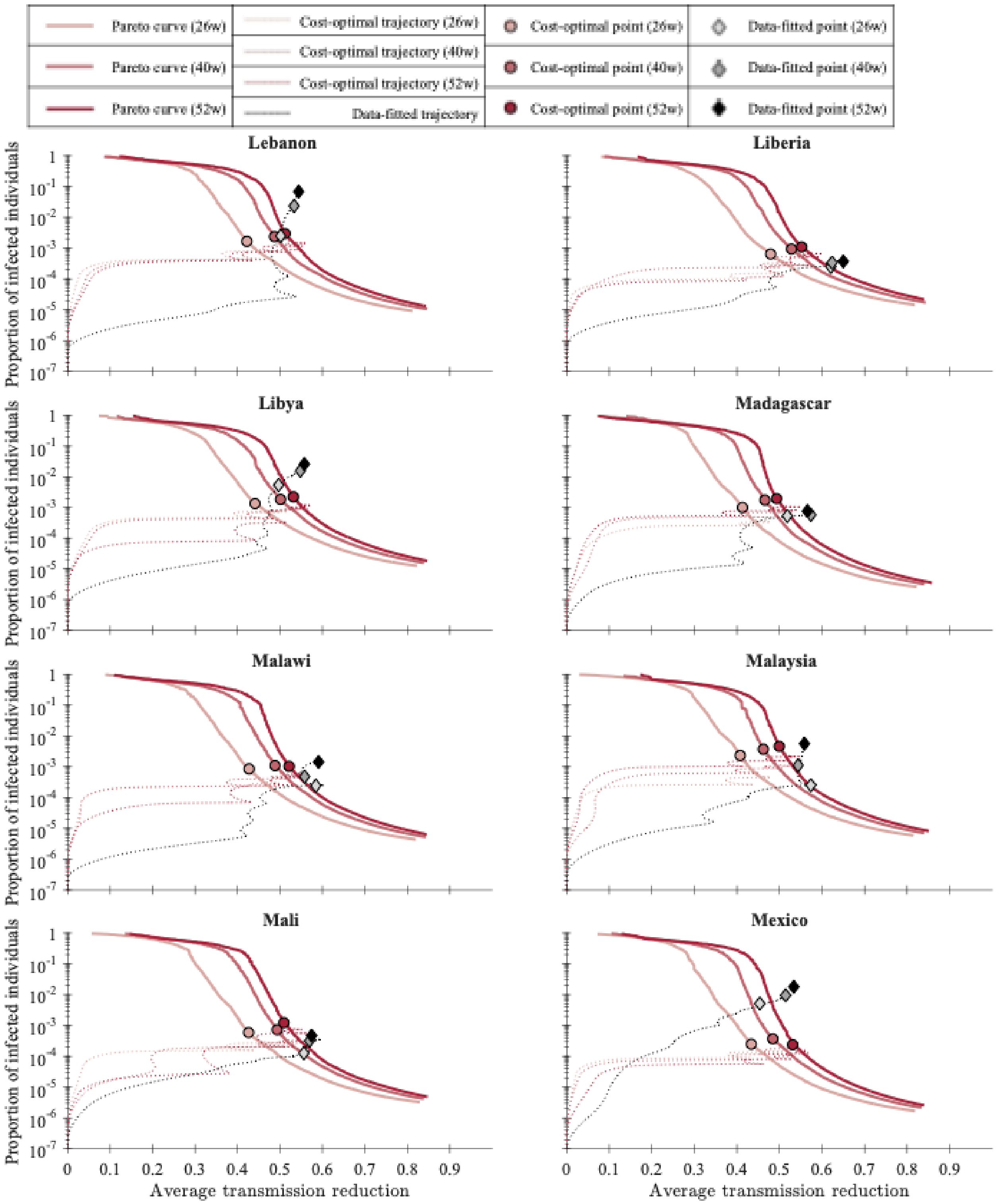


**Figure S3.8: Trajectories of cost-optimal solutions and estimated results for Lebanon, Liberia, Libya, Madagascar, Malawi, Malaysia, Mali, and Mexico.**

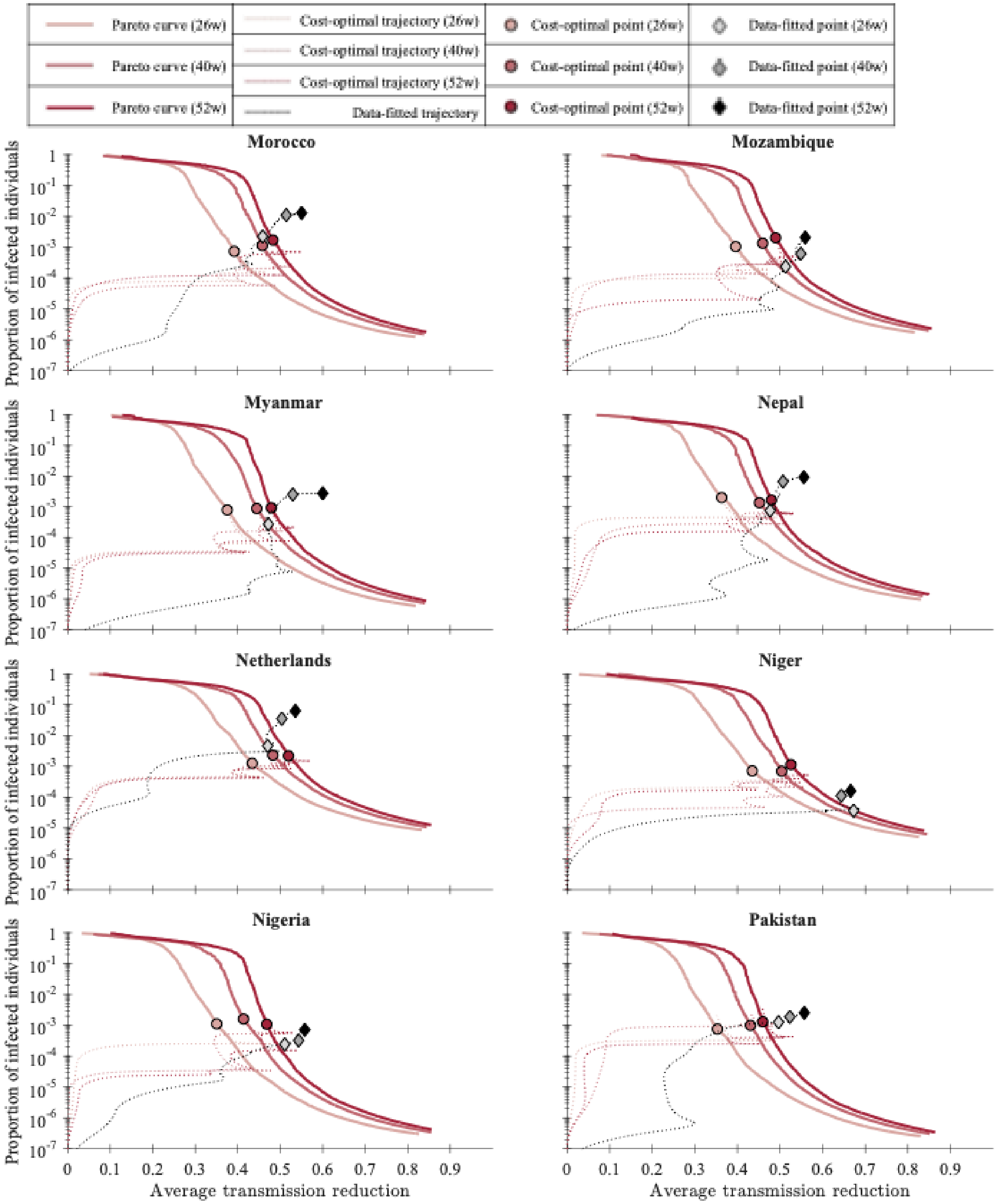


**Figure S3.9: Trajectories of cost-optimal solutions and estimated results for Morocco, Mozambique, Myanmar, Nepal, Netherlands, Niger, Nigeria, and Pakistan.**

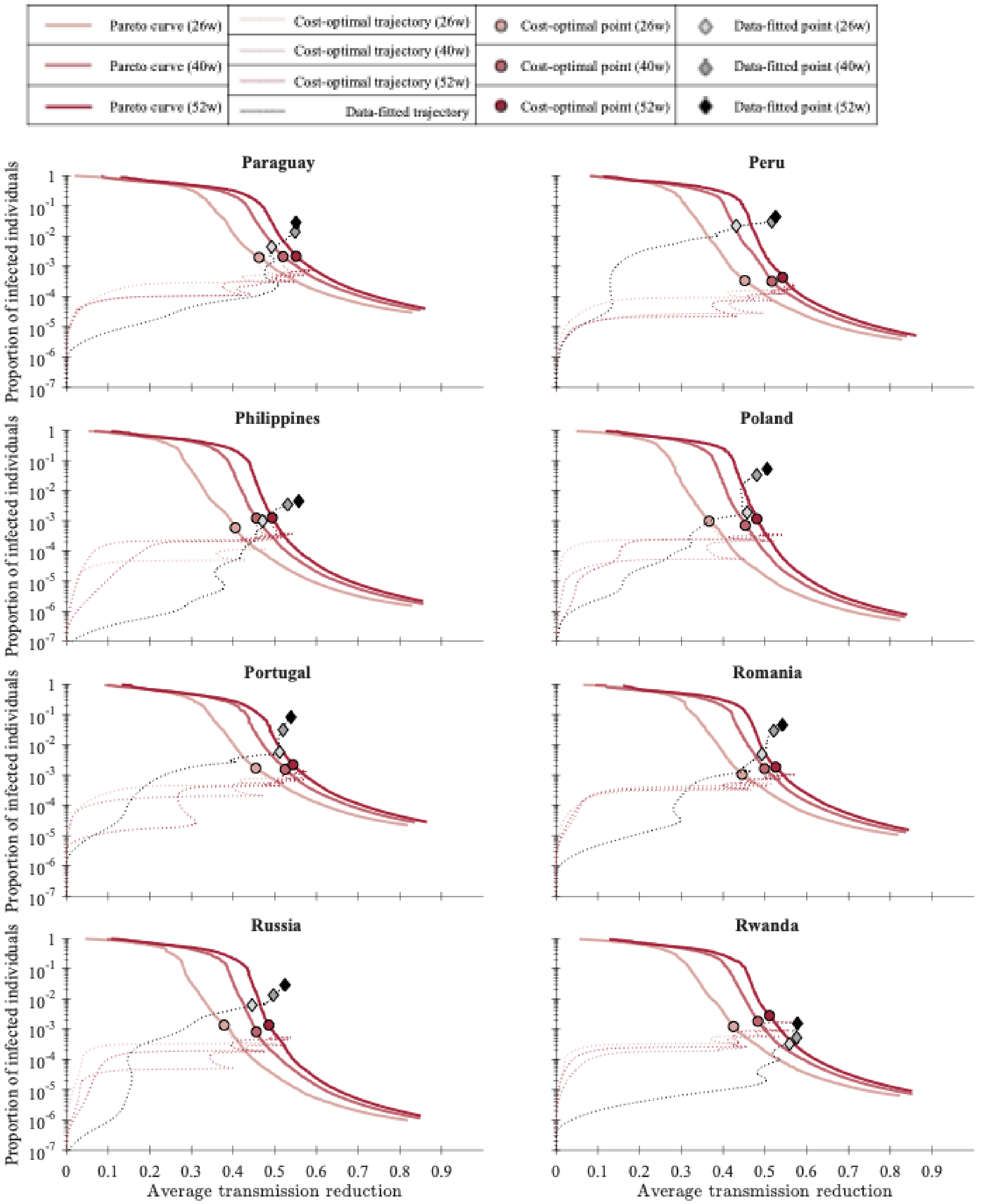


**Figure S3.10: Trajectories of cost-optimal solutions and estimated results for Paraguay, Peru, Philippines, Poland, Portugal, Romania, Russia, and Rwanda.**

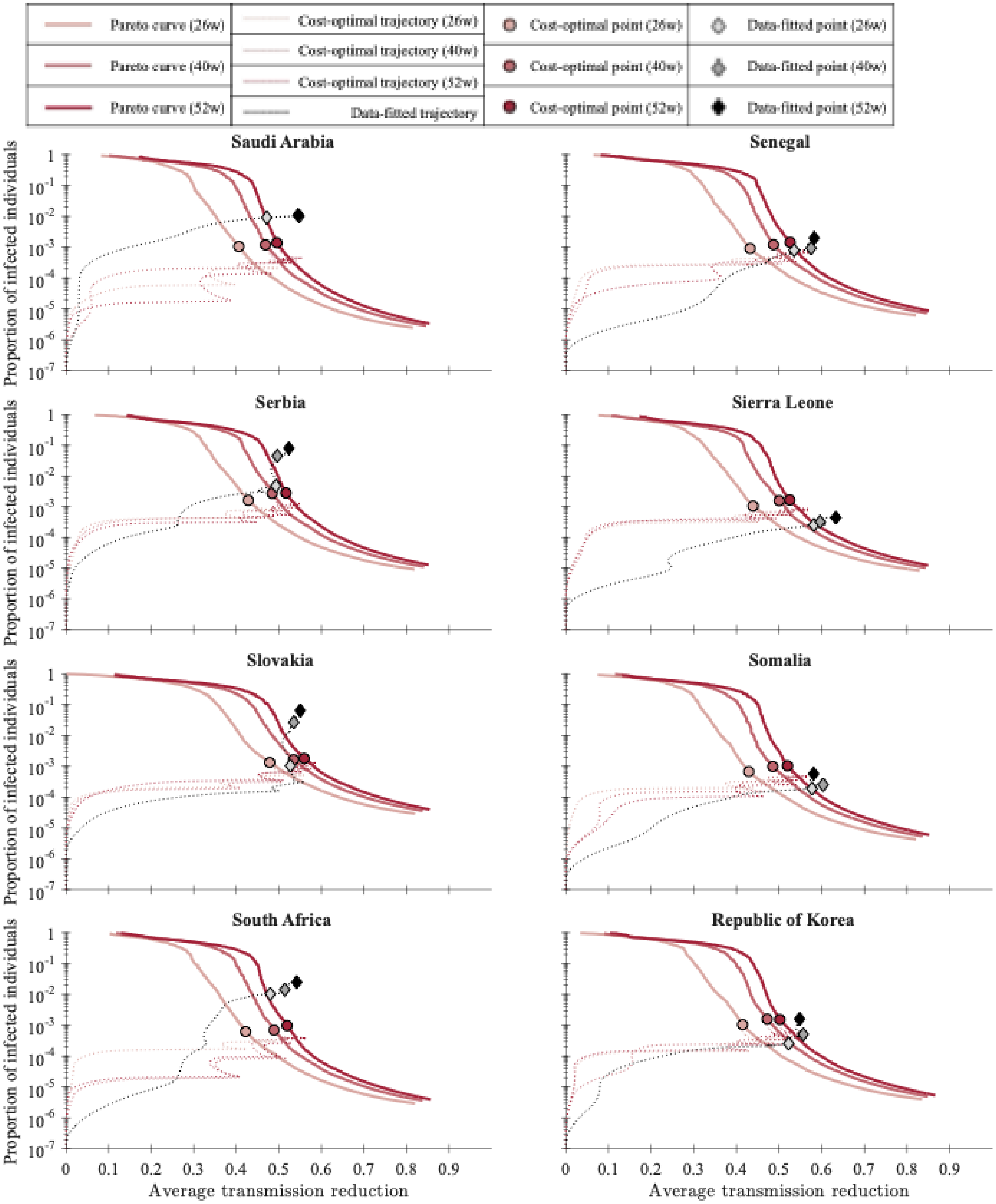


**Figure S3.11: Trajectories of cost-optimal solutions and estimated results for Saudi Arabia, Senegal, Serbia, Sierra Leone, Slovakia, Somalia, South Africa, and Republic of Korea.**

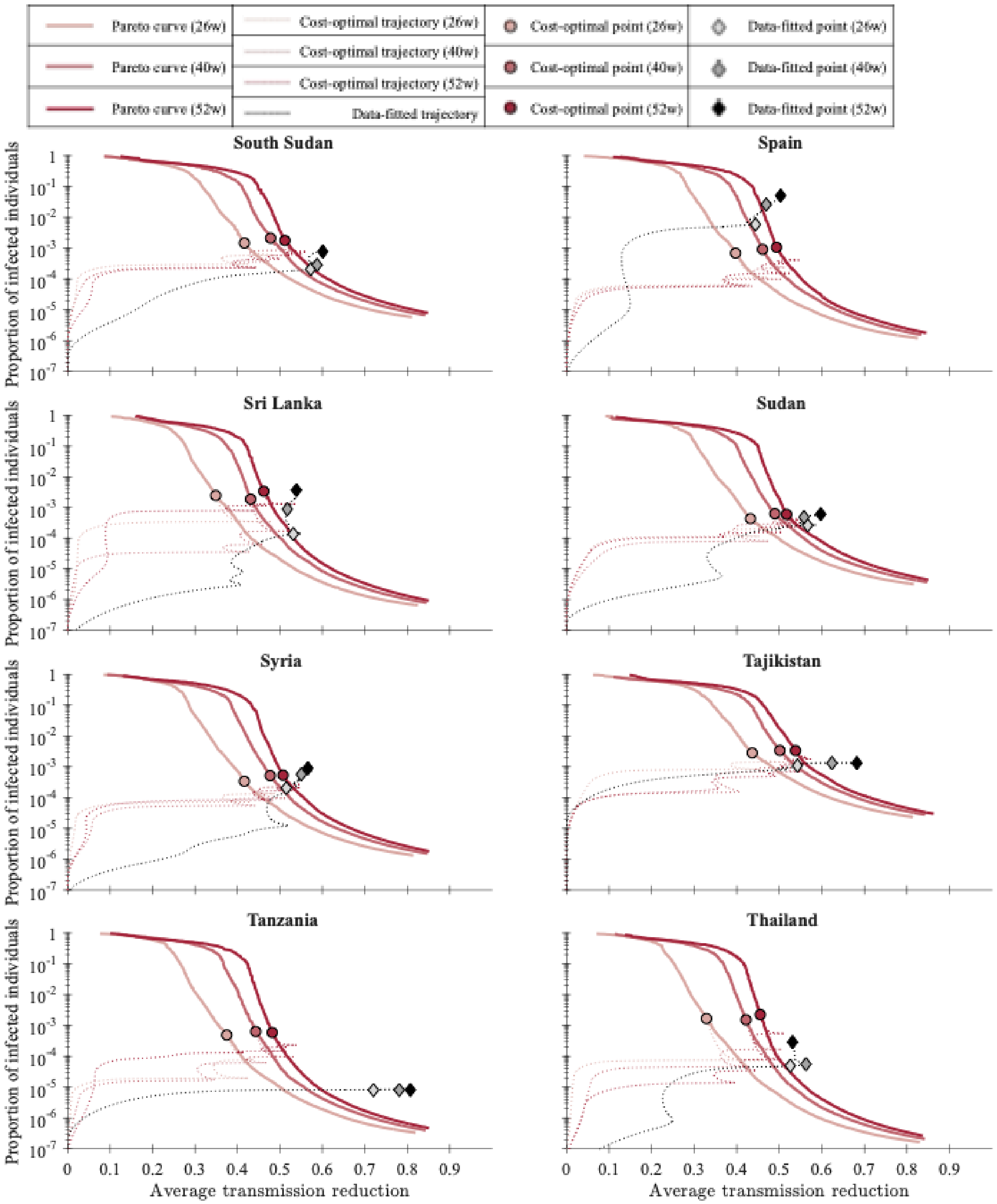


**Figure S3.12: Trajectories of cost-optimal solutions and estimated results for South Sudan, Spain, Sri Lanka, Sudan, Syria, Tajikistan, Tanzania, and Thailand.**

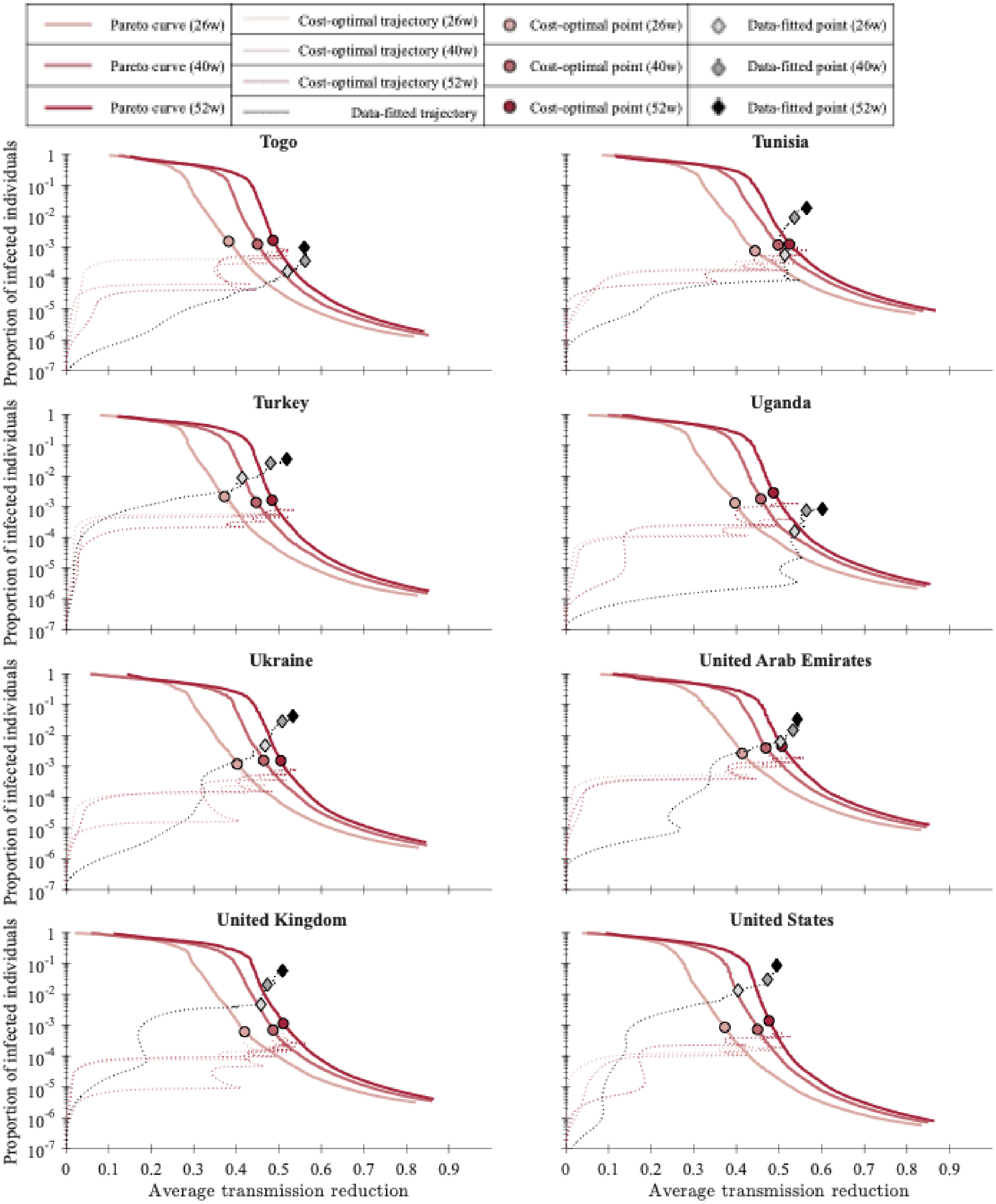


**Figure S3.13: Trajectories of cost-optimal solutions and estimated results for Togo, Tunisia, Turkey, Uganda, Ukraine, United Arab Emirates, United Kingdom, and United States.**

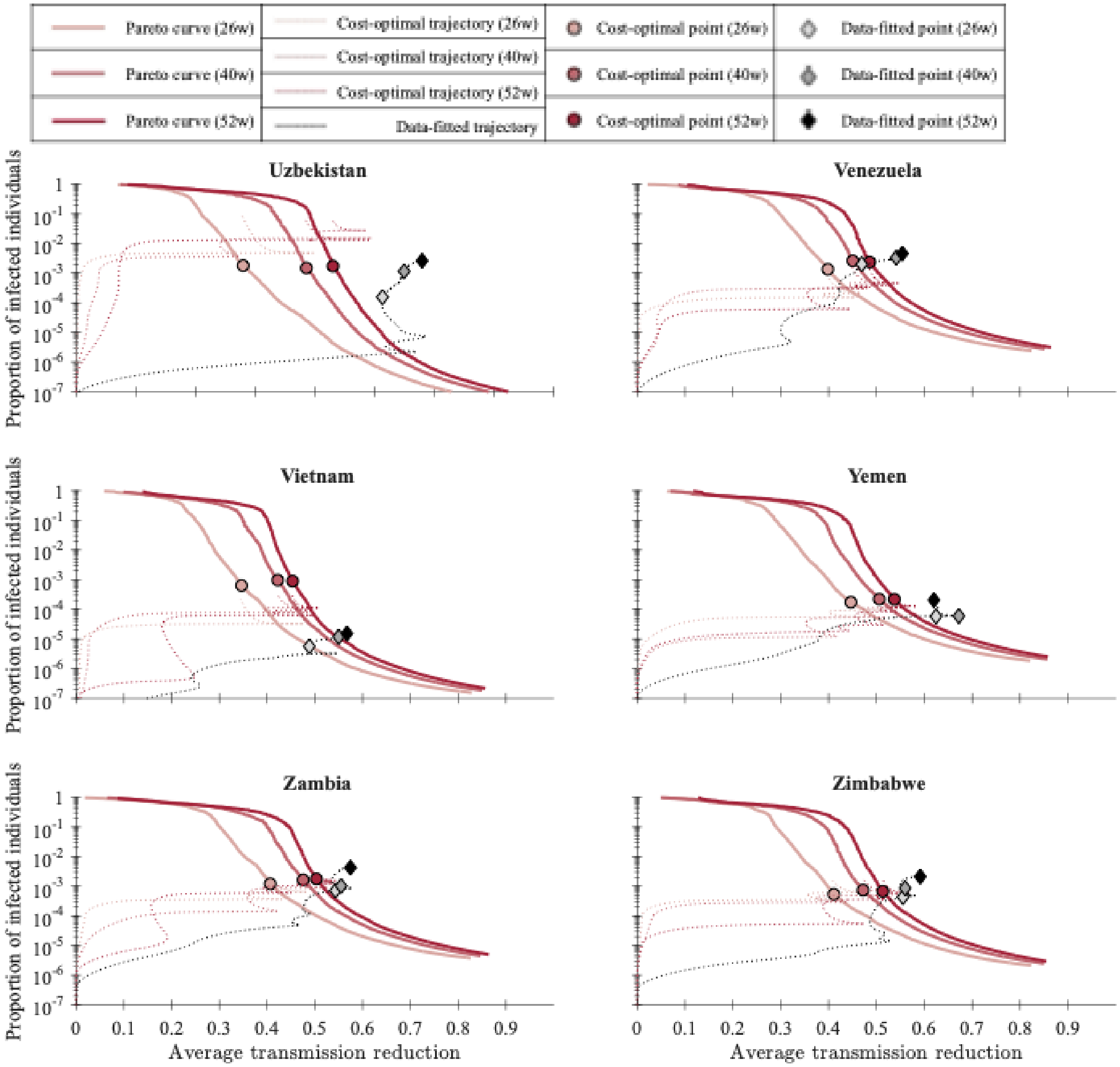


**Figure S3.14: Trajectories of cost-optimal solutions and estimated results for Uzbekistan, Venezuela, Vietnam, Yemen, Zambia, and Zimbabwe.**
