## Supplementary file 4 for "Cost-optimal transmission reduction in early pandemic response: a multi-objective health-economic modelling study across 110 countries"

### Supplementary 4. Cost-optimal strategies in the range of cost per infection

Data analysis reveals a widening gap between baseline estimates (Est.) and optimised pathways (Opt.) as the vaccine implementation time extends from 26 to 52 weeks. Total intervention and infection costs rise substantially across all nations over longer durations. For instance, the United States shows that an estimated cost percentage is 7.38% of GDP (820.89 billion USD) at a 26-week vaccine implementation timeline, which increases to 15.68% of GDP (3,482.14 billion USD) for the 52-week vaccine implementation timeline.

Conversely, some nations maintain stable or improving optimisation profiles over time. China shows that an estimated cost percentage is 5.78% of GDP (420.49 billion USD) at a 26-week vaccine implementation timing, which decreases to 5.65% of GDP (821.59 billion USD) at 52-week vaccine implementation timeline, showing stronger alignment with optimised models in long-term planning. Australia consistently keeps a lower estimated cost percentage as 1.3% of GDP (37.77 billion USD) at a 26-week vaccine implementation timeline, which increases to 6% of GDP (87.45 billion USD) by week 52. Comprehensive data and calculations surrounding these country profiles can be found in Table S4.1.

The excess-cost ratio is defined as (estimated total cost / cost-optimal total cost) − 1, representing the fractional cost relative to the theoretical optimum. Peru (400%), Czechia (337%), and the United Kingdom (235%) showed the largest excess, indicating that their observed intervention strategies were substantially more costly than the model-predicted cost-optimal strategies. The mean excess-cost ratio across all 110 countries for 26, 40, and 52 weeks vaccine implementation timing was 45.2%, 54.9%, and 65.5%, respectively. This indicates that their observed intervention strategies are substantially more costly than the model predicted optimal strategies.

These findings imply that planning long-term vaccine implementation introduces severe compounding inefficiencies if intervention policies were not optimal. For high-GDP nations, relying strictly on baseline estimates for year-long horizons invites hundreds of billions USD in total cost. This simulation result underscores the necessity of implementing structured optimisation frameworks, such as mathematical modelling support, to map the Pareto front that minimises total societal cost before launching large-scale national immunisation initiatives.

**Table S4.1: Estimated and optimal transmission reductions, total costs (infection + intervention), and excess-cost ratio across 26, 40, and 52 weeks of vaccine implementation timelines for the 110 countries.**

| Country | 26 weeks | | | | | 40 weeks | | | | | 52 weeks | | | | |
|---|---|---|---|---|---|---|---|---|---|---|---|---|---|---|---|
| | Trans. reduction | | Costs (Billion USD) | | Excess-cost ratio | Trans. reduction | | Costs (Billion USD) | | Excess-cost ratio | Trans. reduction | | Costs (Billion USD) | | Excess-cost ratio |
| | Est. | Opt. | Est. total cost | Opt. total cost | | Est. | Opt. | Est. total cost | Opt. total cost | | Est. | Opt. | Est. total cost | Opt. total cost | |
| Afghanistan | 0.54 | 0.43 | 0.60 | 0.46 | 32.0% | 0.56 | 0.49 | 0.94 | 0.81 | 16.0% | 0.60 | 0.51 | 1.30 | 1.11 | 17.0% |
| Algeria | 0.50 | 0.42 | 5.48 | 4.46 | 23.0% | 0.54 | 0.49 | 9.11 | 7.82 | 16.0% | 0.57 | 0.51 | 12.38 | 10.72 | 15.0% |
| Angola | 0.51 | 0.39 | 2.05 | 1.62 | 26.0% | 0.55 | 0.46 | 3.47 | 2.92 | 19.0% | 0.57 | 0.48 | 4.68 | 3.99 | 17.0% |
| Argentina | 0.45 | 0.42 | 19.95 | 10.20 | 96.0% | 0.52 | 0.49 | 44.74 | 18.02 | 148.0% | 0.54 | 0.52 | 63.01 | 24.37 | 159.0% |
| Australia | 0.50 | 0.43 | 37.77 | 33.08 | 14.0% | 0.58 | 0.49 | 68.03 | 58.61 | 16.0% | 0.58 | 0.52 | 87.45 | 78.73 | 11.0% |
| Austria | 0.50 | 0.43 | 13.25 | 10.20 | 30.0% | 0.50 | 0.49 | 42.84 | 17.62 | 143.0% | 0.54 | 0.52 | 60.76 | 24.14 | 152.0% |
| Azerbaijan | 0.49 | 0.41 | 1.37 | 1.05 | 31.0% | 0.50 | 0.47 | 2.71 | 1.83 | 48.0% | 0.56 | 0.51 | 3.90 | 2.58 | 51.0% |
| Bangladesh | 0.48 | 0.38 | 9.40 | 7.15 | 31.0% | 0.53 | 0.46 | 15.94 | 13.08 | 22.0% | 0.56 | 0.49 | 21.59 | 18.21 | 19.0% |
| Belarus | 0.51 | 0.43 | 1.96 | 1.43 | 37.0% | 0.52 | 0.49 | 3.30 | 2.53 | 31.0% | 0.55 | 0.52 | 4.90 | 3.46 | 42.0% |
| Belgium | 0.48 | 0.44 | 19.92 | 12.91 | 54.0% | 0.50 | 0.50 | 72.21 | 22.07 | 227.0% | 0.53 | 0.53 | 96.67 | 30.72 | 215.0% |
| Benin | 0.54 | 0.36 | 0.42 | 0.30 | 40.0% | 0.57 | 0.44 | 0.68 | 0.54 | 26.0% | 0.57 | 0.48 | 0.89 | 0.76 | 17.0% |
| Bolivia | 0.46 | 0.42 | 2.26 | 0.93 | 143.0% | 0.52 | 0.48 | 3.22 | 1.63 | 98.0% | 0.54 | 0.51 | 4.88 | 2.25 | 117.0% |
| Brazil | 0.40 | 0.37 | 101.01 | 38.96 | 159.0% | 0.48 | 0.45 | 173.60 | 69.97 | 148.0% | 0.52 | 0.49 | 254.22 | 98.54 | 158.0% |
| Bulgaria | 0.54 | 0.50 | 2.13 | 1.71 | 24.0% | 0.54 | 0.54 | 7.28 | 2.89 | 152.0% | 0.56 | 0.57 | 10.09 | 3.86 | 161.0% |
| Burkina Faso | 0.58 | 0.42 | 0.51 | 0.38 | 34.0% | 0.58 | 0.48 | 0.79 | 0.69 | 14.0% | 0.61 | 0.51 | 1.09 | 0.94 | 16.0% |
| Burundi | 0.57 | 0.36 | 0.08 | 0.06 | 48.0% | 0.58 | 0.44 | 0.13 | 0.10 | 26.0% | 0.58 | 0.47 | 0.17 | 0.14 | 18.0% |
| Cambodia | 0.55 | 0.33 | 1.01 | 0.63 | 61.0% | 0.60 | 0.42 | 1.69 | 1.23 | 37.0% | 0.60 | 0.45 | 2.20 | 1.70 | 29.0% |

| | | | | | | | | | | | | | | |
|---|---|---|---|---|---|---|---|---|---|---|---|---|---|---|
| Cameroon | 0.55 | 0.43 | 1.19 | 0.94 | 27.0% | 0.58 | 0.49 | 1.92 | 1.64 | 17.0% | 0.57 | 0.52 | 2.48 | 2.26 | 10.0% |
| Canada | 0.49 | 0.41 | 53.35 | 38.13 | 40.0% | 0.51 | 0.47 | 90.67 | 68.27 | 33.0% | 0.54 | 0.50 | 158.35 | 93.34 | 70.0% |
| Chad | 0.55 | 0.41 | 0.32 | 0.25 | 29.0% | 0.57 | 0.47 | 0.51 | 0.44 | 15.0% | 0.57 | 0.51 | 0.67 | 0.61 | 10.0% |
| Chile | 0.47 | 0.44 | 16.67 | 6.64 | 151.0% | 0.53 | 0.50 | 25.07 | 11.60 | 116.0% | 0.55 | 0.53 | 35.56 | 15.62 | 128.0% |
| China | 0.57 | 0.36 | 420.49 | 273.10 | 54.0% | 0.59 | 0.44 | 663.67 | 511.62 | 30.0% | 0.56 | 0.47 | 821.59 | 708.46 | 16.0% |
| Colombia | 0.43 | 0.41 | 15.37 | 7.18 | 114.0% | 0.50 | 0.46 | 30.27 | 12.84 | 136.0% | 0.53 | 0.50 | 45.08 | 17.20 | 162.0% |
| Costa Rica | 0.49 | 0.45 | 2.36 | 1.64 | 44.0% | 0.54 | 0.52 | 4.88 | 2.84 | 72.0% | 0.57 | 0.54 | 6.61 | 3.83 | 73.0% |
| Cote d'Ivoire | 0.51 | 0.36 | 1.58 | 1.16 | 36.0% | 0.55 | 0.44 | 2.64 | 2.18 | 21.0% | 0.54 | 0.47 | 3.38 | 2.98 | 13.0% |
| Cuba | 0.56 | 0.42 | 2.87 | 2.29 | 26.0% | 0.58 | 0.48 | 4.62 | 4.06 | 14.0% | 0.57 | 0.52 | 6.24 | 5.64 | 11.0% |
| Czechia | 0.50 | 0.44 | 7.49 | 5.93 | 26.0% | 0.52 | 0.49 | 28.43 | 10.22 | 178.0% | 0.52 | 0.53 | 60.32 | 13.79 | 337.0% |
| Democratic Republic of Congo | 0.57 | 0.40 | 1.67 | 1.25 | 34.0% | 0.57 | 0.47 | 2.59 | 2.23 | 16.0% | 0.59 | 0.50 | 3.48 | 3.09 | 13.0% |
| Denmark | 0.51 | 0.44 | 10.00 | 8.24 | 21.0% | 0.52 | 0.50 | 19.55 | 14.25 | 37.0% | 0.55 | 0.54 | 30.73 | 19.54 | 57.0% |
| Dominican Republic | 0.49 | 0.45 | 3.11 | 2.18 | 43.0% | 0.54 | 0.50 | 5.09 | 3.82 | 33.0% | 0.57 | 0.53 | 7.23 | 5.15 | 40.0% |
| Ecuador | 0.48 | 0.45 | 5.16 | 2.69 | 92.0% | 0.53 | 0.50 | 8.81 | 4.66 | 89.0% | 0.53 | 0.54 | 12.90 | 6.31 | 104.0% |
| Egypt | 0.51 | 0.40 | 9.20 | 6.75 | 36.0% | 0.55 | 0.47 | 14.73 | 12.01 | 23.0% | 0.56 | 0.50 | 19.98 | 16.43 | 22.0% |
| El Salvador | 0.54 | 0.48 | 1.00 | 0.73 | 37.0% | 0.56 | 0.53 | 1.61 | 1.22 | 32.0% | 0.57 | 0.56 | 2.20 | 1.65 | 33.0% |
| Ethiopia | 0.49 | 0.36 | 2.62 | 2.01 | 30.0% | 0.55 | 0.44 | 4.53 | 3.67 | 23.0% | 0.55 | 0.48 | 5.93 | 5.21 | 14.0% |
| Finland | 0.56 | 0.45 | 7.93 | 6.42 | 23.0% | 0.55 | 0.51 | 12.59 | 11.06 | 14.0% | 0.57 | 0.54 | 18.05 | 15.15 | 19.0% |
| France | 0.42 | 0.34 | 59.60 | 48.25 | 24.0% | 0.46 | 0.42 | 216.00 | 90.08 | 140.0% | 0.50 | 0.46 | 354.23 | 127.93 | 177.0% |
| Germany | 0.50 | 0.40 | 117.68 | 83.61 | 41.0% | 0.50 | 0.47 | 186.44 | 148.27 | 26.0% | 0.52 | 0.50 | 377.05 | 205.02 | 84.0% |
| Ghana | 0.53 | 0.40 | 2.00 | 1.54 | 30.0% | 0.56 | 0.46 | 3.23 | 2.75 | 17.0% | 0.56 | 0.50 | 4.24 | 3.81 | 11.0% |
| Greece | 0.52 | 0.46 | 5.68 | 5.07 | 12.0% | 0.54 | 0.53 | 13.50 | 8.59 | 57.0% | 0.56 | 0.56 | 19.31 | 11.74 | 64.0% |
| Guatemala | 0.49 | 0.43 | 2.88 | 1.90 | 52.0% | 0.55 | 0.51 | 4.82 | 3.29 | 46.0% | 0.57 | 0.53 | 6.55 | 4.46 | 47.0% |
| Guinea | 0.55 | 0.42 | 0.41 | 0.32 | 28.0% | 0.59 | 0.48 | 0.68 | 0.58 | 18.0% | 0.59 | 0.51 | 0.88 | 0.79 | 12.0% |
| Haiti | 0.56 | 0.44 | 0.46 | 0.37 | 26.0% | 0.58 | 0.50 | 0.73 | 0.64 | 15.0% | 0.61 | 0.52 | 1.00 | 0.87 | 14.0% |
| Honduras | 0.51 | 0.46 | 0.93 | 0.64 | 45.0% | 0.55 | 0.51 | 1.58 | 1.08 | 46.0% | 0.57 | 0.55 | 2.20 | 1.49 | 48.0% |
| Hungary | 0.50 | 0.46 | 4.55 | 3.91 | 16.0% | 0.52 | 0.52 | 17.54 | 6.76 | 160.0% | 0.54 | 0.55 | 27.53 | 9.10 | 203.0% |
| India | 0.40 | 0.31 | 62.54 | 48.86 | 28.0% | 0.48 | 0.41 | 128.02 | 94.21 | 36.0% | 0.52 | 0.45 | 176.45 | 134.05 | 32.0% |
| Indonesia | 0.47 | 0.38 | 28.66 | 23.04 | 24.0% | 0.52 | 0.45 | 50.95 | 41.45 | 23.0% | 0.54 | 0.49 | 73.60 | 57.73 | 27.0% |
| Iran | 0.47 | 0.42 | 11.49 | 7.48 | 54.0% | 0.50 | 0.48 | 22.83 | 13.27 | 72.0% | 0.52 | 0.51 | 34.33 | 18.19 | 89.0% |
| Iraq | 0.45 | 0.41 | 7.81 | 5.44 | 44.0% | 0.52 | 0.46 | 14.83 | 9.62 | 54.0% | 0.54 | 0.51 | 19.53 | 13.30 | 47.0% |
| Israel | 0.46 | 0.41 | 13.31 | 9.25 | 44.0% | 0.53 | 0.47 | 28.12 | 16.03 | 75.0% | 0.52 | 0.52 | 48.36 | 22.58 | 114.0% |
| Italy | 0.45 | 0.41 | 64.49 | 42.63 | 51.0% | 0.46 | 0.46 | 161.38 | 73.32 | 120.0% | 0.51 | 0.51 | 302.10 | 103.71 | 191.0% |
| Japan | 0.45 | 0.34 | 114.05 | 89.27 | 28.0% | 0.51 | 0.43 | 199.79 | 169.29 | 18.0% | 0.51 | 0.46 | 275.17 | 236.09 | 17.0% |
| Jordan | 0.47 | 0.35 | 1.13 | 0.86 | 31.0% | 0.49 | 0.42 | 2.87 | 1.60 | 79.0% | 0.51 | 0.47 | 4.35 | 2.26 | 93.0% |
| Kazakhstan | 0.49 | 0.41 | 5.71 | 3.84 | 49.0% | 0.53 | 0.46 | 9.13 | 6.83 | 34.0% | 0.55 | 0.50 | 12.46 | 9.33 | 34.0% |
| Kenya | 0.49 | 0.36 | 2.74 | 2.04 | 34.0% | 0.53 | 0.44 | 4.62 | 3.73 | 24.0% | 0.54 | 0.47 | 6.10 | 5.24 | 16.0% |
| Kyrgyzstan | 0.54 | 0.47 | 0.36 | 0.24 | 49.0% | 0.56 | 0.52 | 0.53 | 0.41 | 29.0% | 0.62 | 0.55 | 0.71 | 0.55 | 29.0% |
| Laos | 0.79 | 0.35 | 0.74 | 0.35 | 115.0% | 0.77 | 0.43 | 1.11 | 0.65 | 70.0% | 0.76 | 0.46 | 1.44 | 0.89 | 61.0% |
| Lebanon | 0.50 | 0.42 | 1.40 | 1.14 | 23.0% | 0.53 | 0.49 | 3.22 | 2.00 | 61.0% | 0.54 | 0.51 | 6.05 | 2.75 | 120.0% |
| Liberia | 0.62 | 0.48 | 0.11 | 0.09 | 25.0% | 0.62 | 0.53 | 0.17 | 0.15 | 13.0% | 0.65 | 0.55 | 0.23 | 0.20 | 14.0% |
| Libya | 0.50 | 0.44 | 2.18 | 1.62 | 34.0% | 0.55 | 0.50 | 4.25 | 2.84 | 49.0% | 0.56 | 0.53 | 6.06 | 3.90 | 55.0% |
| Madagascar | 0.52 | 0.41 | 0.40 | 0.32 | 24.0% | 0.57 | 0.47 | 0.68 | 0.57 | 20.0% | 0.57 | 0.49 | 0.87 | 0.78 | 11.0% |
| Malawi | 0.58 | 0.43 | 0.40 | 0.31 | 29.0% | 0.56 | 0.49 | 0.58 | 0.54 | 9.0% | 0.59 | 0.52 | 0.83 | 0.72 | 15.0% |
| Malaysia | 0.57 | 0.41 | 10.68 | 7.87 | 36.0% | 0.54 | 0.46 | 15.72 | 13.89 | 13.0% | 0.56 | 0.50 | 21.72 | 19.35 | 12.0% |
| Mali | 0.56 | 0.43 | 0.57 | 0.45 | 26.0% | 0.57 | 0.49 | 0.90 | 0.80 | 12.0% | 0.57 | 0.51 | 1.18 | 1.10 | 8.0% |
| Mexico | 0.45 | 0.43 | 70.22 | 30.77 | 128.0% | 0.51 | 0.48 | 126.60 | 53.03 | 139.0% | 0.53 | 0.53 | 210.09 | 72.25 | 191.0% |
| Morocco | 0.46 | 0.39 | 3.42 | 2.69 | 27.0% | 0.51 | 0.46 | 7.10 | 4.81 | 48.0% | 0.55 | 0.48 | 9.48 | 6.76 | 40.0% |
| Mozambique | 0.51 | 0.40 | 0.46 | 0.37 | 26.0% | 0.55 | 0.46 | 0.77 | 0.65 | 17.0% | 0.56 | 0.49 | 1.04 | 0.91 | 14.0% |
| Myanmar | 0.47 | 0.38 | 1.75 | 1.44 | 22.0% | 0.53 | 0.44 | 3.25 | 2.57 | 26.0% | 0.60 | 0.48 | 4.68 | 3.59 | 31.0% |
| Nepal | 0.48 | 0.37 | 0.90 | 0.73 | 22.0% | 0.51 | 0.45 | 1.65 | 1.31 | 25.0% | 0.56 | 0.48 | 2.34 | 1.81 | 29.0% |
| Netherlands | 0.47 | 0.44 | 26.72 | 21.73 | 23.0% | 0.50 | 0.50 | 71.61 | 37.34 | 92.0% | 0.54 | 0.53 | 113.66 | 52.15 | 118.0% |
| Niger | 0.67 | 0.43 | 0.56 | 0.38 | 46.0% | 0.64 | 0.49 | 0.82 | 0.67 | 23.0% | 0.67 | 0.53 | 1.11 | 0.91 | 21.0% |
| Nigeria | 0.51 | 0.36 | 13.03 | 9.54 | 36.0% | 0.54 | 0.43 | 21.28 | 17.56 | 21.0% | 0.56 | 0.48 | 28.55 | 24.79 | 15.0% |
| Pakistan | 0.50 | 0.37 | 8.95 | 6.79 | 32.0% | 0.52 | 0.45 | 14.42 | 12.06 | 20.0% | 0.56 | 0.48 | 19.93 | 16.72 | 19.0% |
| Paraguay | 0.49 | 0.45 | 1.32 | 1.05 | 26.0% | 0.55 | 0.51 | 2.63 | 1.82 | 44.0% | 0.55 | 0.55 | 4.01 | 2.44 | 64.0% |
| Peru | 0.43 | 0.45 | 33.03 | 5.76 | 473.0% | 0.52 | 0.52 | 47.61 | 9.88 | 382.0% | 0.53 | 0.54 | 67.24 | 13.44 | 400.0% |
| Philippines | 0.47 | 0.39 | 9.65 | 7.92 | 22.0% | 0.53 | 0.45 | 17.68 | 14.18 | 25.0% | 0.56 | 0.49 | 24.02 | 19.50 | 23.0% |
| Poland | 0.46 | 0.37 | 15.51 | 11.79 | 31.0% | 0.48 | 0.45 | 54.99 | 21.35 | 158.0% | 0.51 | 0.48 | 82.56 | 29.86 | 177.0% |
| Portugal | 0.51 | 0.45 | 8.06 | 5.91 | 36.0% | 0.52 | 0.53 | 20.13 | 10.06 | 100.0% | 0.54 | 0.54 | 40.80 | 13.57 | 201.0% |
| Romania | 0.49 | 0.45 | 8.03 | 5.73 | 40.0% | 0.52 | 0.50 | 21.80 | 10.05 | 117.0% | 0.54 | 0.53 | 31.76 | 13.51 | 135.0% |
| Russia | 0.45 | 0.38 | 50.91 | 34.43 | 48.0% | 0.50 | 0.46 | 93.33 | 59.87 | 56.0% | 0.52 | 0.49 | 150.96 | 83.63 | 80.0% |
| Rwanda | 0.56 | 0.42 | 0.32 | 0.25 | 27.0% | 0.58 | 0.48 | 0.51 | 0.45 | 15.0% | 0.58 | 0.51 | 0.68 | 0.61 | 10.0% |

| | | | | | | | | | | | | | | | |
|---|---|---|---|---|---|---|---|---|---|---|---|---|---|---|---|
| Saudi Arabia | 0.47 | 0.41 | 35.67 | 21.86 | 63.0% | 0.55 | 0.47 | 56.01 | 38.22 | 47.0% | 0.55 | 0.50 | 69.85 | 52.20 | 34.0% |
| Senegal | 0.54 | 0.43 | 0.73 | 0.60 | 22.0% | 0.58 | 0.49 | 1.20 | 1.04 | 15.0% | 0.58 | 0.53 | 1.61 | 1.44 | 12.0% |
| Serbia | 0.49 | 0.43 | 1.47 | 1.15 | 28.0% | 0.50 | 0.48 | 3.85 | 2.04 | 89.0% | 0.52 | 0.52 | 6.00 | 2.79 | 115.0% |
| Sierra Leone | 0.58 | 0.44 | 0.22 | 0.17 | 27.0% | 0.60 | 0.50 | 0.34 | 0.30 | 15.0% | 0.63 | 0.52 | 0.47 | 0.41 | 17.0% |
| Slovakia | 0.53 | 0.48 | 2.92 | 2.66 | 10.0% | 0.54 | 0.53 | 8.74 | 4.53 | 93.0% | 0.55 | 0.56 | 16.38 | 6.11 | 168.0% |
| Somalia | 0.58 | 0.43 | 0.29 | 0.22 | 30.0% | 0.60 | 0.49 | 0.46 | 0.39 | 17.0% | 0.58 | 0.52 | 0.58 | 0.54 | 8.0% |
| South Africa | 0.48 | 0.42 | 18.85 | 8.98 | 110.0% | 0.51 | 0.49 | 28.59 | 16.01 | 79.0% | 0.54 | 0.52 | 43.83 | 22.14 | 98.0% |
| Republic of Korea | 0.52 | 0.41 | 43.08 | 34.74 | 24.0% | 0.56 | 0.47 | 70.78 | 61.44 | 15.0% | 0.55 | 0.50 | 92.52 | 84.74 | 9.0% |
| South Sudan | 0.57 | 0.42 | 0.16 | 0.12 | 31.0% | 0.59 | 0.48 | 0.25 | 0.21 | 19.0% | 0.60 | 0.51 | 0.33 | 0.28 | 16.0% |
| Spain | 0.44 | 0.40 | 46.32 | 30.28 | 53.0% | 0.47 | 0.46 | 114.49 | 53.54 | 114.0% | 0.50 | 0.49 | 193.73 | 73.87 | 162.0% |
| Sri Lanka | 0.53 | 0.35 | 2.32 | 1.62 | 43.0% | 0.52 | 0.43 | 3.51 | 2.96 | 18.0% | 0.54 | 0.46 | 4.87 | 4.21 | 16.0% |
| Sudan | 0.57 | 0.43 | 1.05 | 0.81 | 30.0% | 0.56 | 0.49 | 1.60 | 1.41 | 13.0% | 0.60 | 0.52 | 2.22 | 1.90 | 16.0% |
| Syria | 0.52 | 0.42 | 0.65 | 0.54 | 21.0% | 0.55 | 0.48 | 1.09 | 0.94 | 16.0% | 0.57 | 0.51 | 1.47 | 1.30 | 13.0% |
| Tajikistan | 0.54 | 0.44 | 0.25 | 0.21 | 19.0% | 0.62 | 0.50 | 0.43 | 0.36 | 21.0% | 0.68 | 0.54 | 0.61 | 0.49 | 24.0% |
| Tanzania | 0.72 | 0.37 | 2.36 | 1.28 | 84.0% | 0.78 | 0.44 | 3.93 | 2.29 | 72.0% | 0.81 | 0.48 | 5.29 | 3.24 | 63.0% |
| Thailand | 0.53 | 0.33 | 13.17 | 8.61 | 53.0% | 0.56 | 0.42 | 21.67 | 16.52 | 31.0% | 0.53 | 0.46 | 26.65 | 23.46 | 14.0% |
| Togo | 0.52 | 0.38 | 0.21 | 0.16 | 28.0% | 0.56 | 0.45 | 0.34 | 0.28 | 22.0% | 0.56 | 0.49 | 0.45 | 0.39 | 15.0% |
| Tunisia | 0.51 | 0.44 | 1.16 | 1.02 | 15.0% | 0.54 | 0.50 | 2.65 | 1.78 | 49.0% | 0.56 | 0.52 | 4.20 | 2.40 | 75.0% |
| Turkey | 0.41 | 0.37 | 22.13 | 15.91 | 39.0% | 0.48 | 0.45 | 46.70 | 27.66 | 69.0% | 0.52 | 0.48 | 64.50 | 38.94 | 66.0% |
| Uganda | 0.54 | 0.40 | 1.07 | 0.82 | 31.0% | 0.56 | 0.46 | 1.75 | 1.45 | 21.0% | 0.60 | 0.49 | 2.42 | 2.04 | 19.0% |
| Ukraine | 0.47 | 0.40 | 3.73 | 2.72 | 37.0% | 0.51 | 0.46 | 9.61 | 4.80 | 100.0% | 0.53 | 0.50 | 13.66 | 6.66 | 105.0% |
| United Arab Emirates | 0.50 | 0.41 | 12.61 | 9.64 | 31.0% | 0.53 | 0.47 | 21.46 | 16.99 | 26.0% | 0.54 | 0.51 | 31.21 | 23.77 | 31.0% |
| United Kingdom | 0.46 | 0.40 | 95.62 | 62.51 | 53.0% | 0.47 | 0.47 | 232.49 | 109.75 | 112.0% | 0.51 | 0.50 | 507.49 | 151.39 | 235.0% |
| United States | 0.40 | 0.37 | 820.89 | 439.54 | 87.0% | 0.47 | 0.44 | 1644.93 | 784.18 | 110.0% | 0.49 | 0.49 | 3482.14 | 1109.66 | 214.0% |
| Uzbekistan | 0.51 | 0.37 | 1.93 | 1.48 | 30.0% | 0.55 | 0.45 | 3.22 | 2.66 | 21.0% | 0.58 | 0.48 | 4.46 | 3.70 | 21.0% |
| Venezuela | 0.47 | 0.39 | 4.36 | 3.51 | 24.0% | 0.54 | 0.46 | 7.71 | 6.24 | 23.0% | 0.55 | 0.49 | 10.28 | 8.74 | 18.0% |
| Vietnam | 0.49 | 0.32 | 8.50 | 5.88 | 45.0% | 0.55 | 0.41 | 14.71 | 11.36 | 30.0% | 0.57 | 0.45 | 19.74 | 16.17 | 22.0% |
| Yemen | 0.62 | 0.45 | 0.68 | 0.52 | 31.0% | 0.67 | 0.51 | 1.12 | 0.89 | 25.0% | 0.62 | 0.54 | 1.38 | 1.21 | 13.0% |
| Zambia | 0.54 | 0.42 | 0.72 | 0.57 | 26.0% | 0.55 | 0.48 | 1.13 | 1.01 | 12.0% | 0.57 | 0.51 | 1.60 | 1.39 | 15.0% |
| Zimbabwe | 0.56 | 0.44 | 0.84 | 0.69 | 21.0% | 0.56 | 0.50 | 1.32 | 1.18 | 12.0% | 0.59 | 0.53 | 1.63 | 0.15 | 15.0% |

Figure S4.0.1 shows the geographical distribution of excess-cost ratios across the 26-, 40-, and 52-week simulation horizons. Regional differences become more pronounced as the horizon extends. Many European countries and the United States show increasing excess-cost ratios, while several South American countries already exhibit high ratios at 26 weeks. Many Asian and African countries have comparatively low ratios across all three horizons, although variation remains within each region. These patterns indicate that the relative cost gap between model-inferred responses and cost-optimal strategies depends on the evaluation horizon. Each ratio is calculated against the optimum for the corresponding country and horizon; a lower ratio therefore indicates closer proximity to that modelled minimum, without necessarily implying lower absolute costs or fewer infections.

Table S4.2 summarises the fitted cross-country relationships between deviations from cost-optimal average transmission reduction and excess-cost ratios. Using a fitted excess-cost ratio of 40% as an exploratory threshold, the corresponding intervals were 6.4-20.0, 6.1-24.6, and 5.2-25.3 percentage points above the country-specific optima for the 26-, 40-, and 52-week horizons, respectively. The interval widened with the simulation horizon, indicating that a broader range of deviations was associated with fitted costs within 40% of the modelled minimum. Fit quality varied across horizons, with $R^2$ values of 0.843, 0.547, and 0.701, respectively. These intervals describe associations across countries and require further assessment before being used to guide changes in an individual country’s transmission reduction strategy.

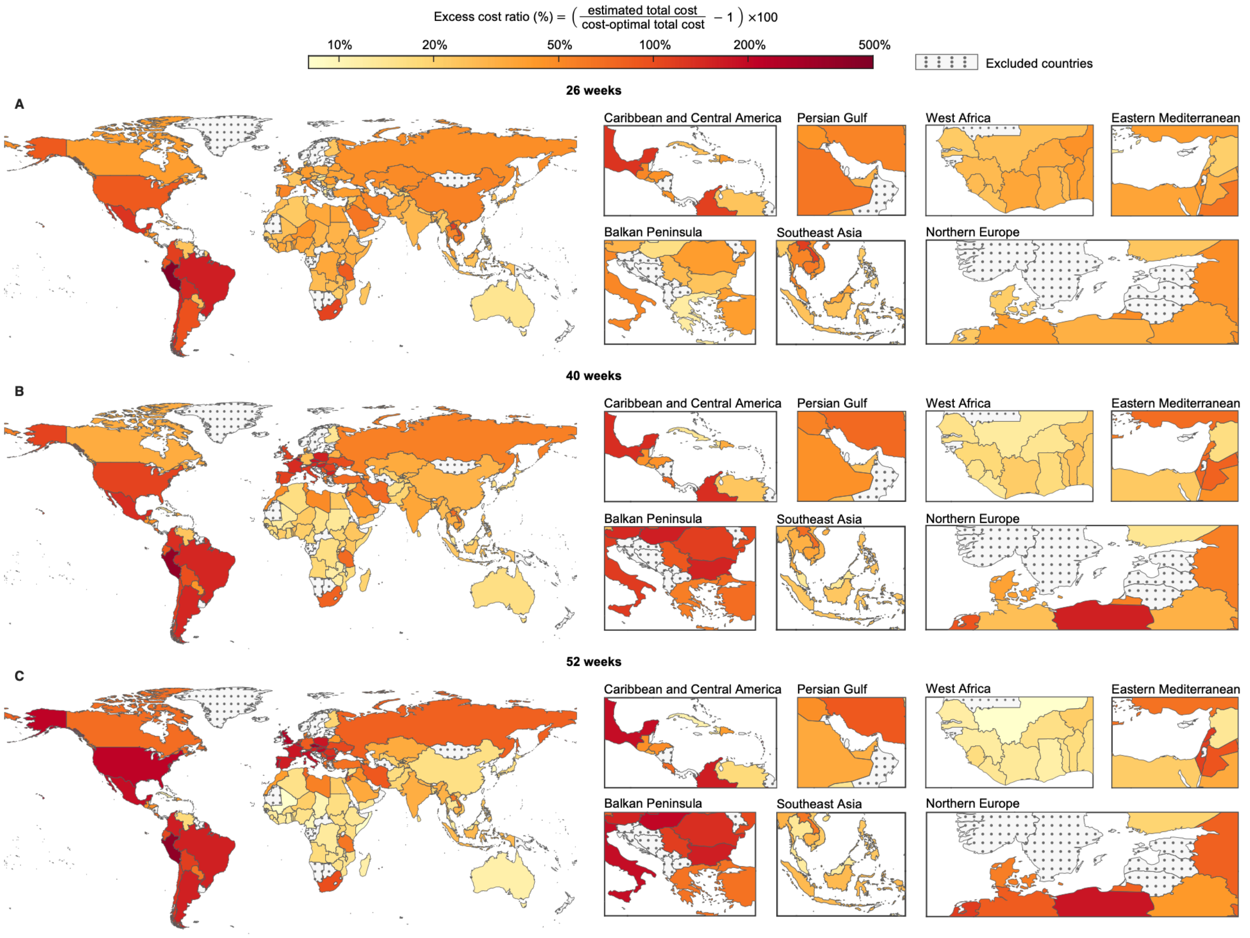


**Figure S4.0.1: Geographical distribution of excess-cost ratios across three simulation horizons.**
Panels A, B, and C present results for 26, 40, and 52 weeks, respectively, for 110 countries. The excess-cost ratio is calculated as [(model-inferred total cost / cost-optimal total cost) − 1] × 100%. All panels share the same colour scale, with darker red indicating larger percentage deviations above the country- and horizon-specific minimum cost. Regional insets provide additional geographical detail. Grey-stippled areas indicate countries excluded by the population criterion.

**Table S4.2: Fitted relationships between transmission reduction deviations and excess-cost ratios, with threshold intervals by simulation horizon.**

| Scenario | Best-fit curve | $R^2$ | Unstable under-intervention zone ($x_{under}$) | Cost-effective transmission-reduction targets | Over-intervention zone ($x_{over}$) |
|---|---|---|---|---|---|
| 26-week | y = (0.127)/(x+0.0419) +4.96x -1.12 | 0.843 | 0.064 | 0.064–0.200 | 0.200 |
| 40-week | y = (0.306)/(x+0.0829) +6.47x -2.12 | 0.547 | 0.061 | 0.061–0.246 | 0.246 |
| 52-week | y = (0.214)/(x+0.0606) +6.08x -1.82 | 0.701 | 0.052 | 0.052–0.253 | 0.253 |

### 4.1. Simulation result for 26 weeks

The axes, legend, and simulation parameters correspond to Figure 3 in the manuscript. The heatmaps display the total societal cost ($C_{tot} = C_{int} + C_{inf}$) as a function of the average transmission reduction, computed using the country-specific default cost per infection ($C_2 = C_H + f \times VSL$, where $C_H = 0.02 \times GDP_{per}$, $f$ is the country-specific CFR, and $VSL = 54.8966 \times GDP_{per}$). The curves show the cost-optimal solution (the point on the Pareto front that minimises $C_{tot}$) as the cost per infection is varied continuously across a range, illustrating how the recommended intervention intensity changes with different cost-per-infection assumptions.

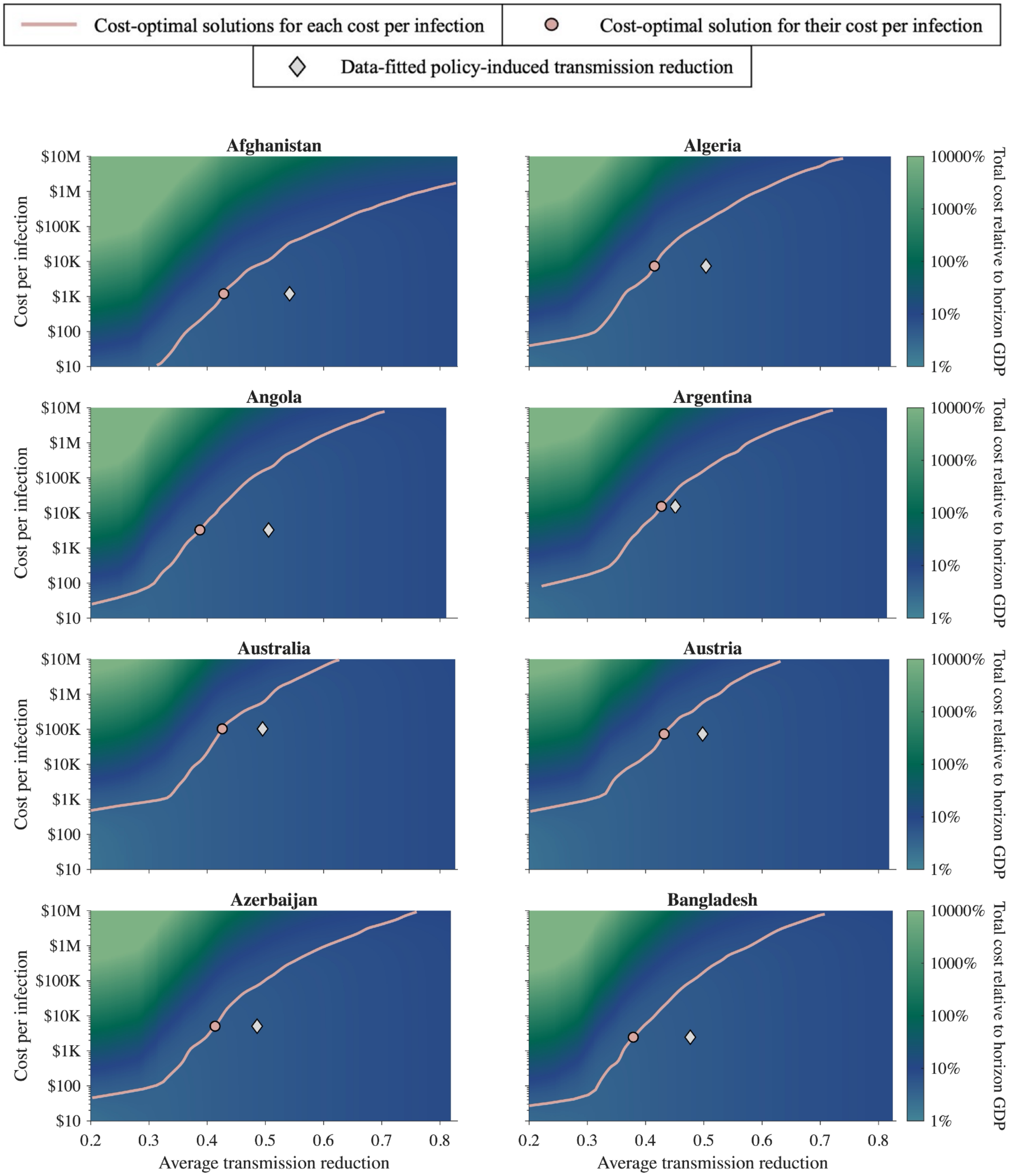


**Figure S4.1.1: The cost-optimal solution under various cost per infection for Afghanistan, Algeria, Angola, Argentina, Australia, Austria, Azerbaijan, and Bangladesh.**

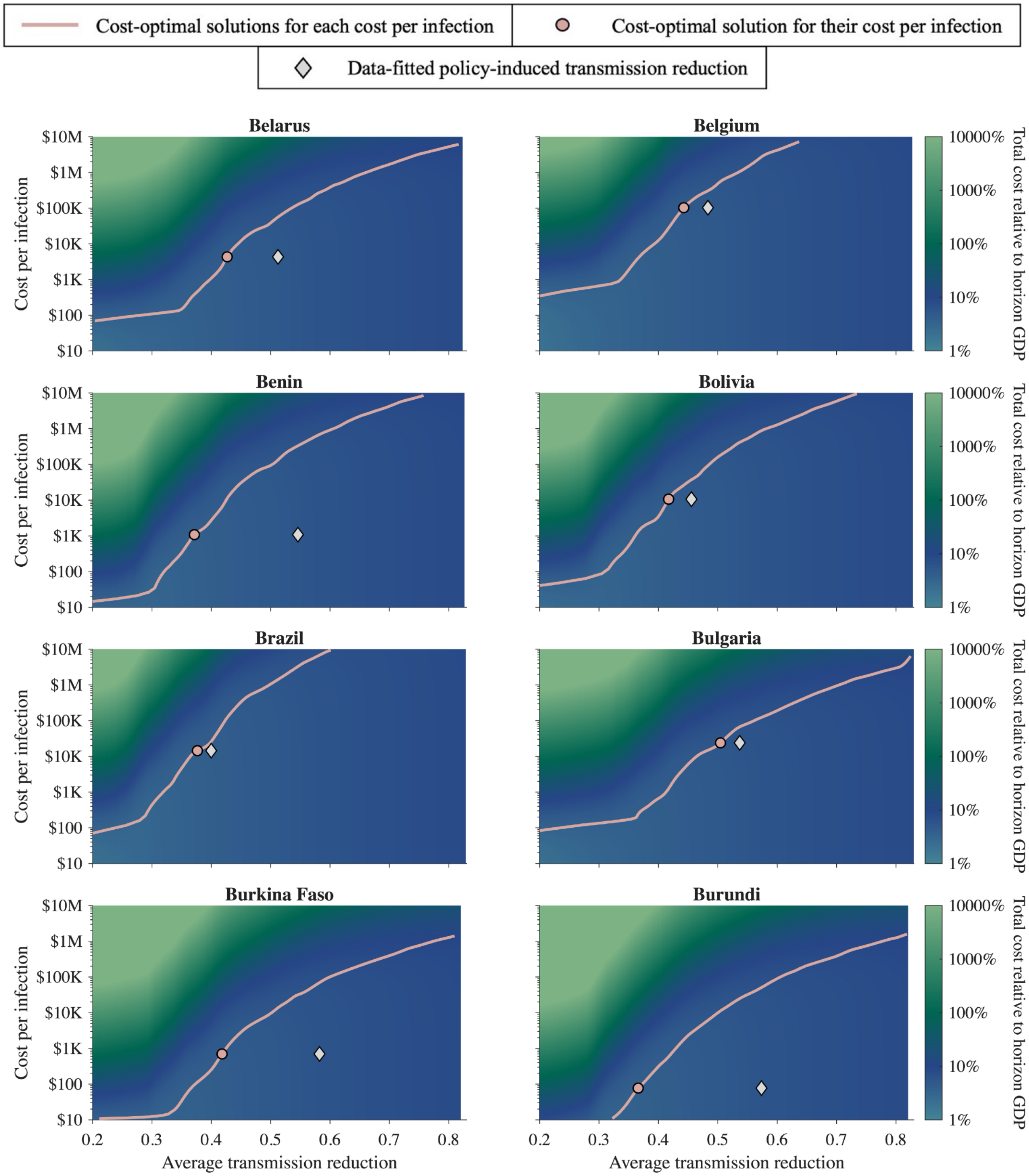


**Figure S4.1.2: The cost-optimal solution under various cost per infection for Belarus, Belgium, Benin, Bolivia, Brazil, Bulgaria, Burkina Faso, and Burundi.**

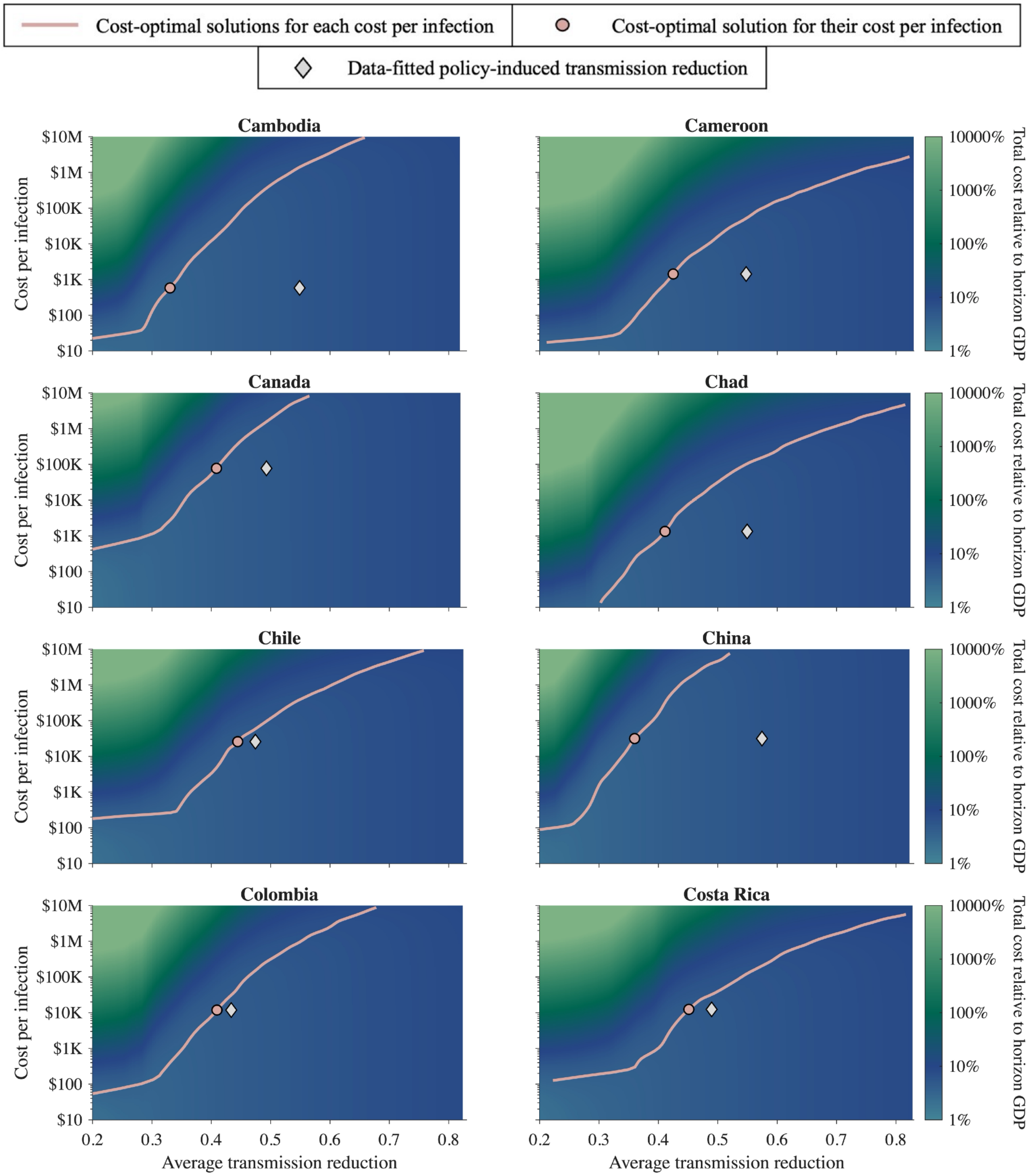


**Figure S4.1.3: The cost-optimal solution under various cost per infection for Cambodia, Cameroon, Canada, Chad, Chile, China, Colombia, and Costa Rica.**

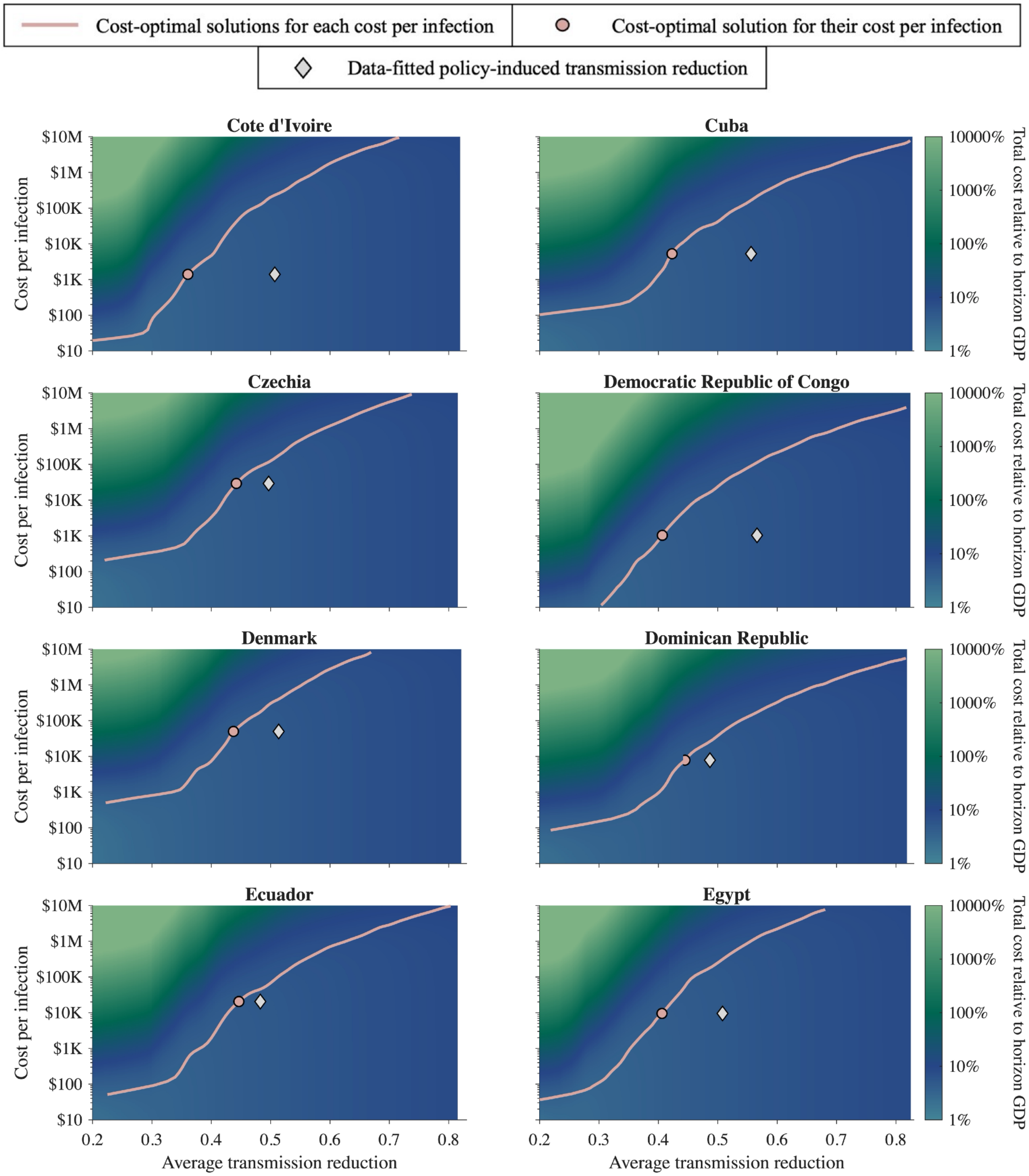


**Figure S4.1.4: The cost-optimal solution under various cost per infection for Cote d'Ivoire, Cuba, Czechia, Democratic Republic of Congo, Denmark, Dominican Republic, Ecuador, and Egypt.**

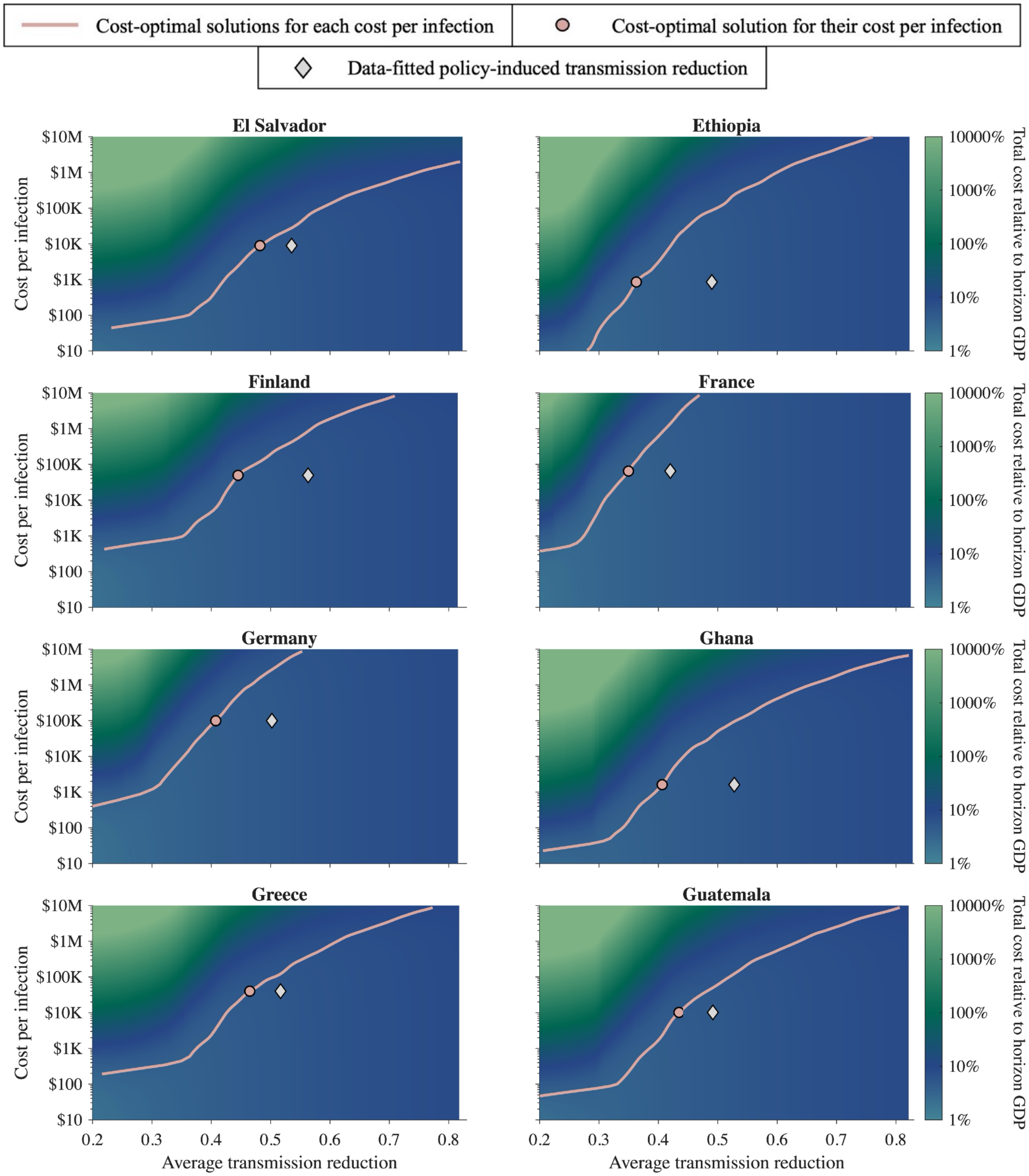


**Figure S4.1.5: The cost-optimal solution under various cost per infection for El Salvador, Ethiopia, Finland, France, Germany, Ghana, Greece, and Guatemala.**

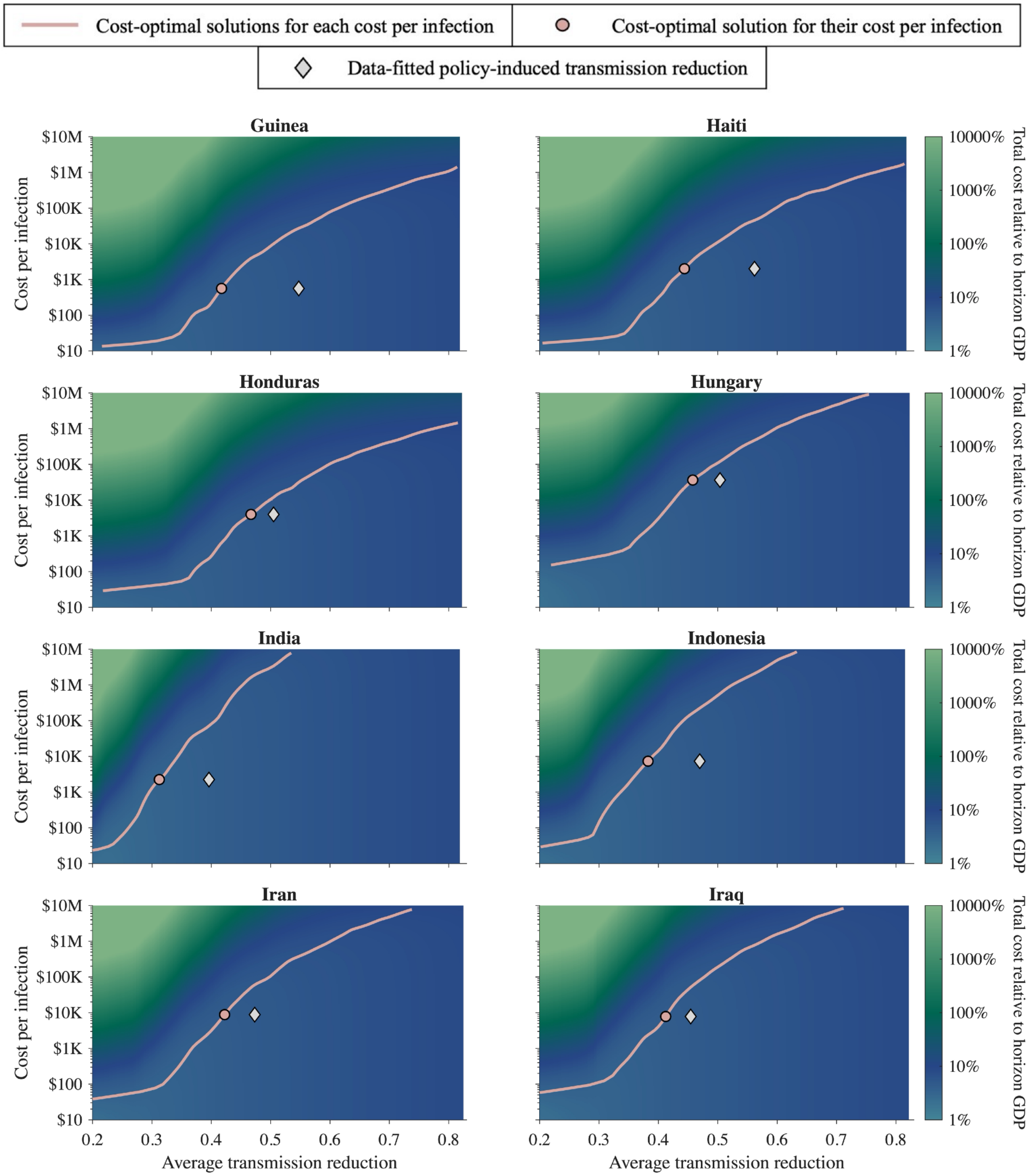


**Figure S4.1.6: The cost-optimal solution under various cost per infection for Guinea, Haiti, Honduras, Hungary, India, Indonesia, Iran, and Iraq.**

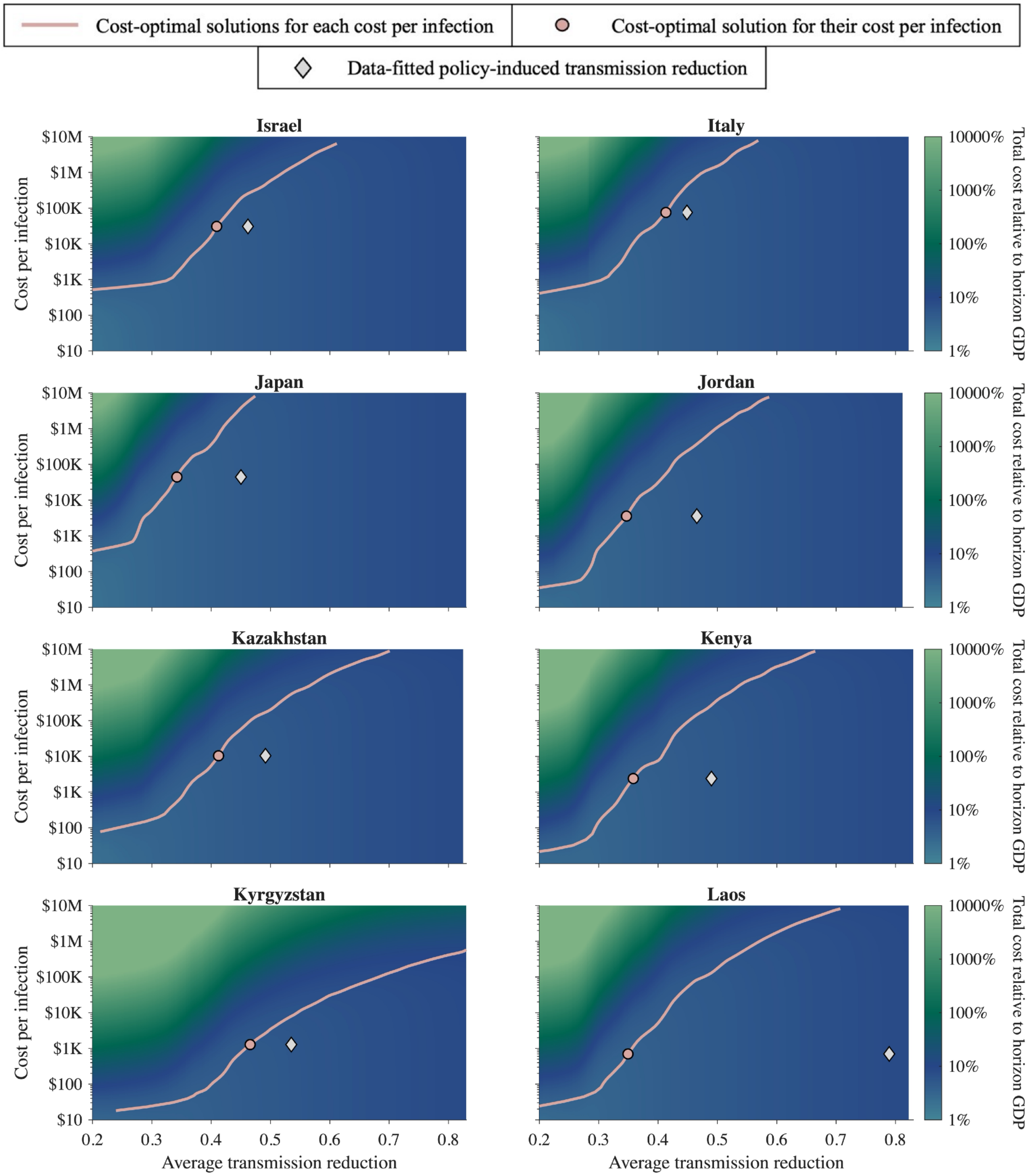


**Figure S4.1.7: The cost-optimal solution under various cost per infection for Israel, Italy, Japan, Jordan, Kazakhstan, Kenya, Kyrgyzstan, and Laos.**

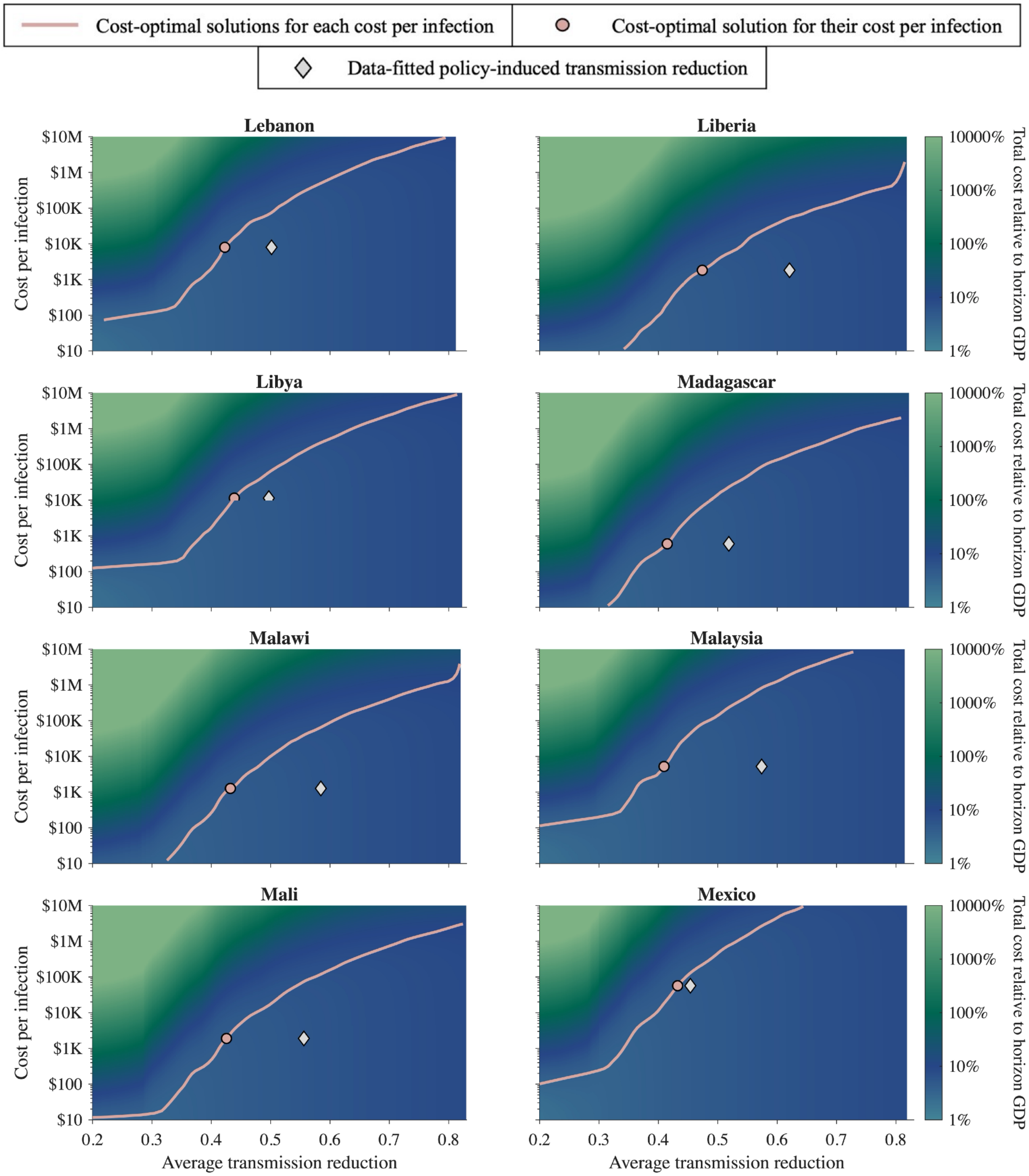


**Figure S4.1.8: The cost-optimal solution under various cost per infection for Lebanon, Liberia, Libya, Madagascar, Malawi, Malaysia, Mali, and Mexico.**

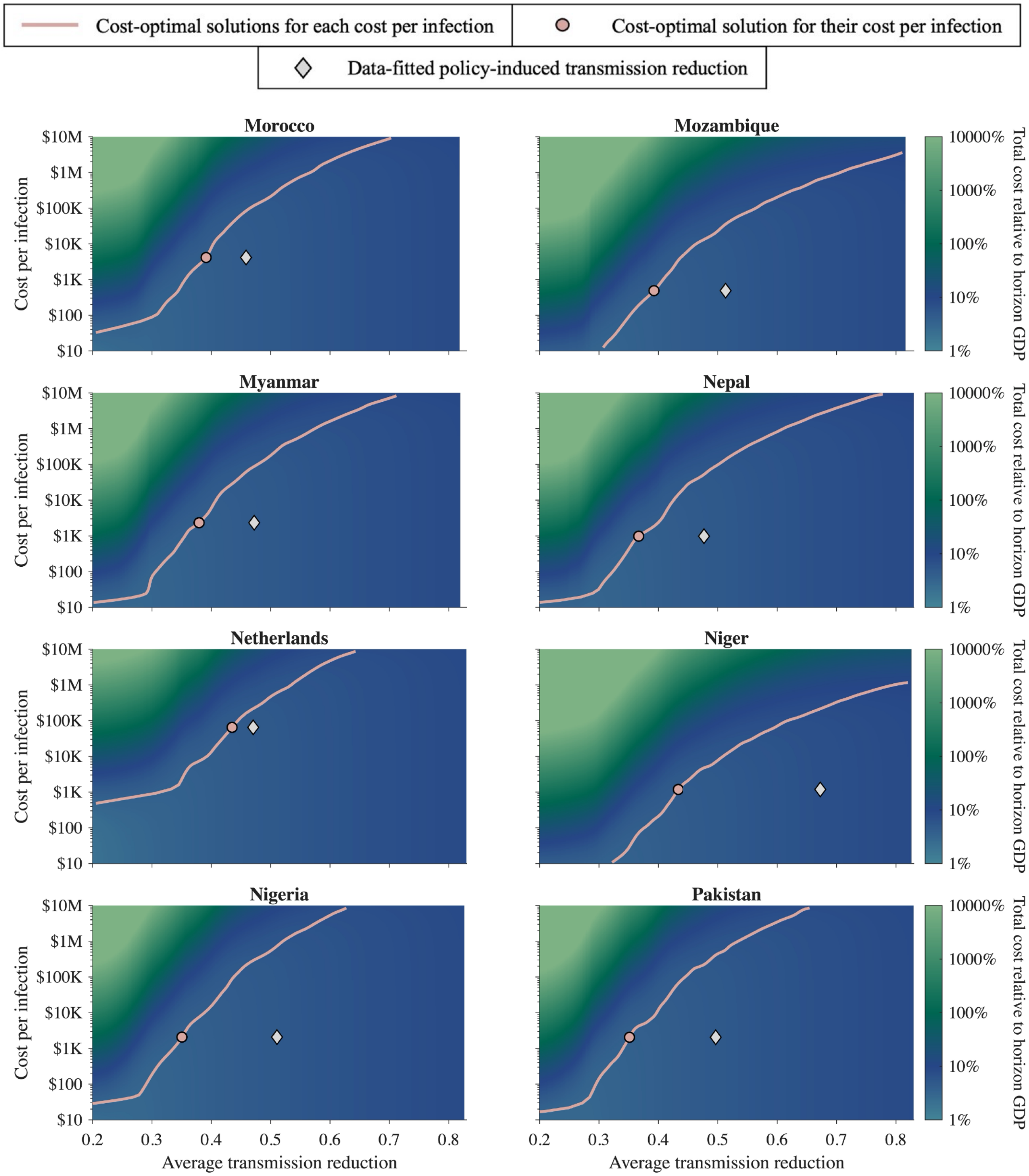


**Figure S4.1.9: The cost-optimal solution under various cost per infection for Morocco, Mozambique, Myanmar, Nepal, Netherlands, Niger, Nigeria, and Pakistan.**

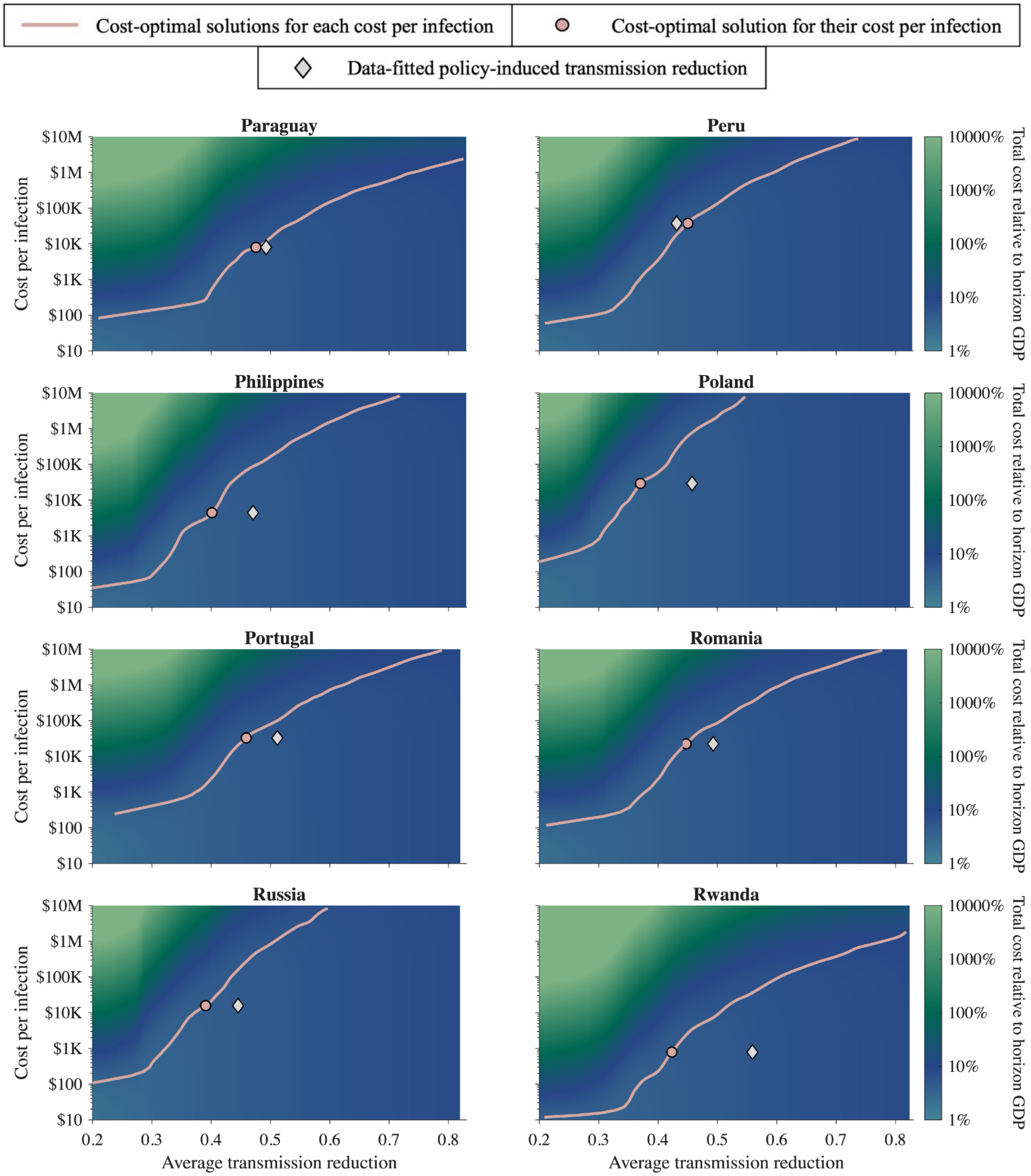


**Figure S4.1.10: The cost-optimal solution under various cost per infection for Paraguay, Peru, Philippines, Poland, Portugal, Romania, Russia, and Rwanda.**

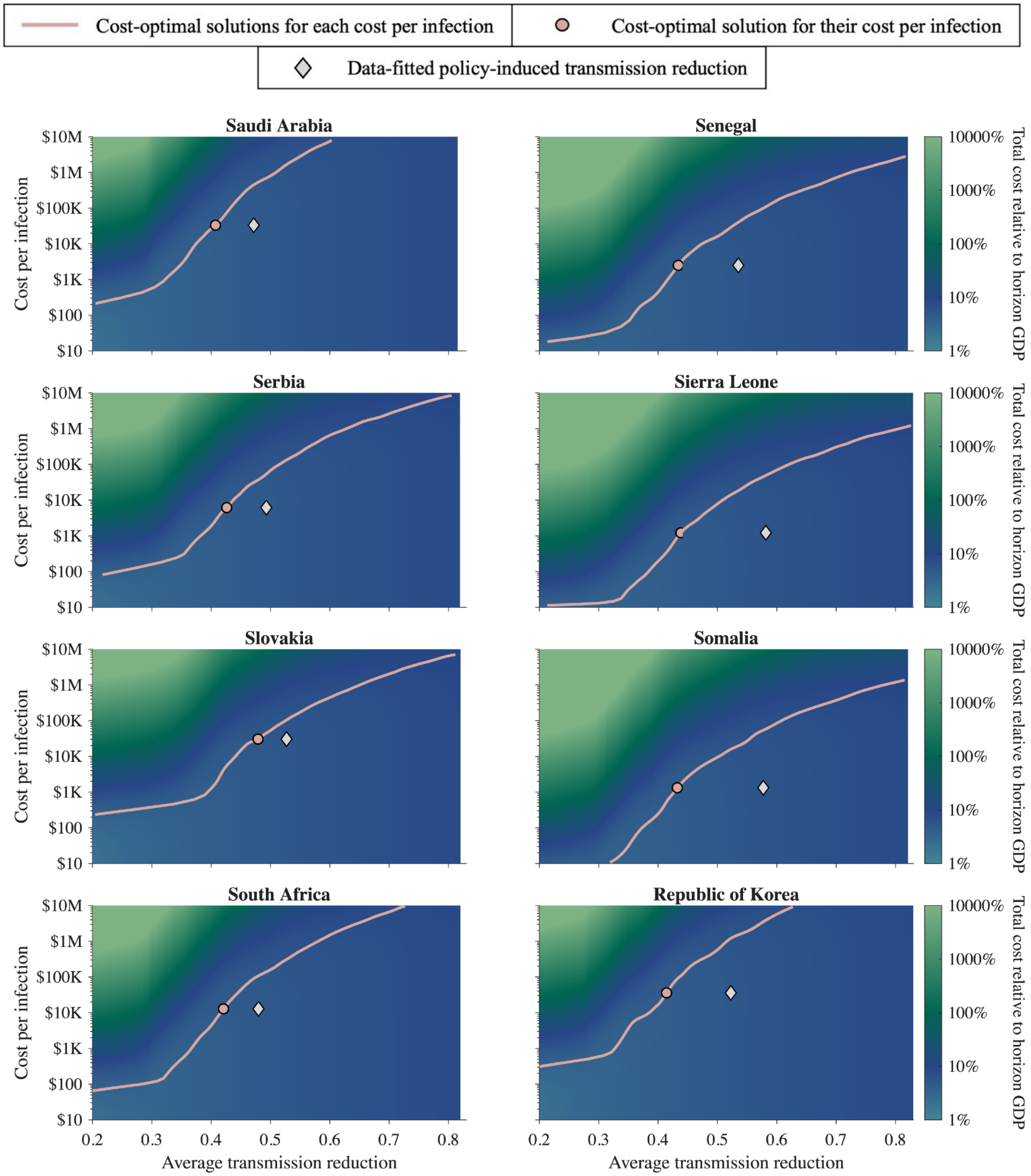


**Figure S4.1.11: The cost-optimal solution under various cost per infection for Saudi Arabia, Senegal, Serbia, Sierra Leone, Slovakia, Somalia, South Africa, and Republic of Korea.**

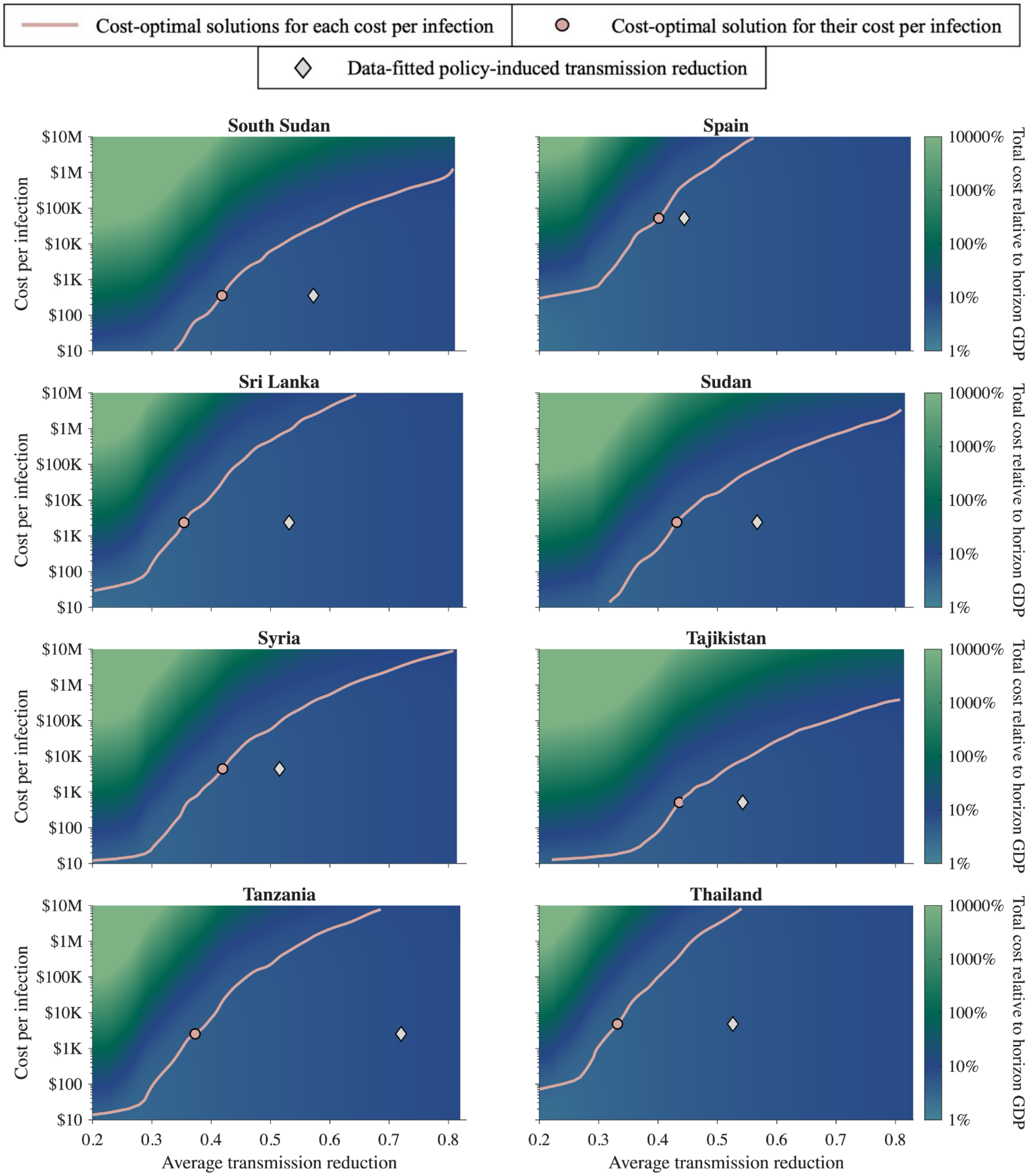


**Figure S4.1.12: The cost-optimal solution under various cost per infection for South Sudan, Spain, Sri Lanka, Sudan, Syria, Tajikistan, Tanzania, and Thailand.**

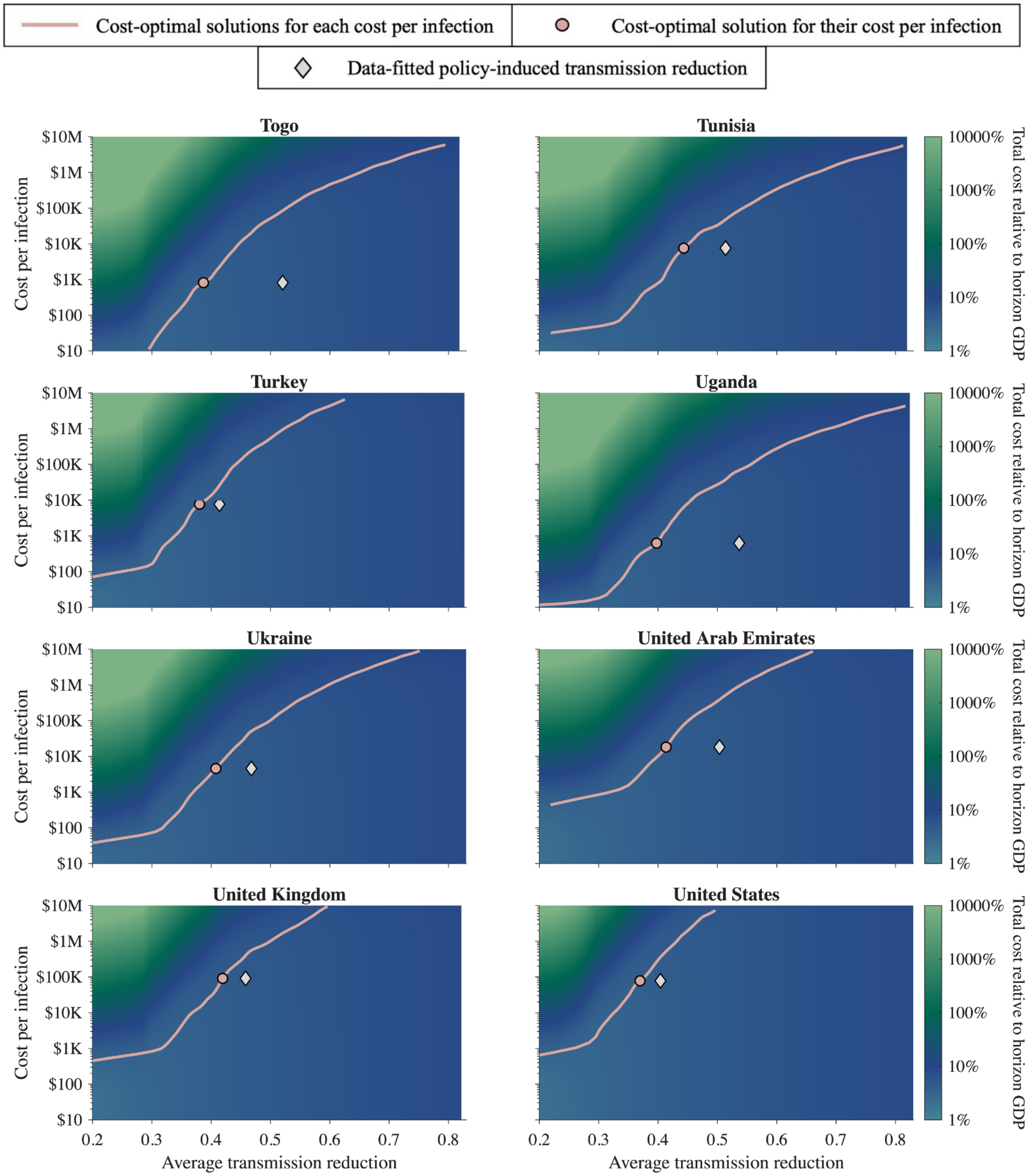


**Figure S4.1.13: The cost-optimal solution under various cost per infection for Togo, Tunisia, Turkey, Uganda, Ukraine, United Arab Emirates, United Kingdom, and United States.**

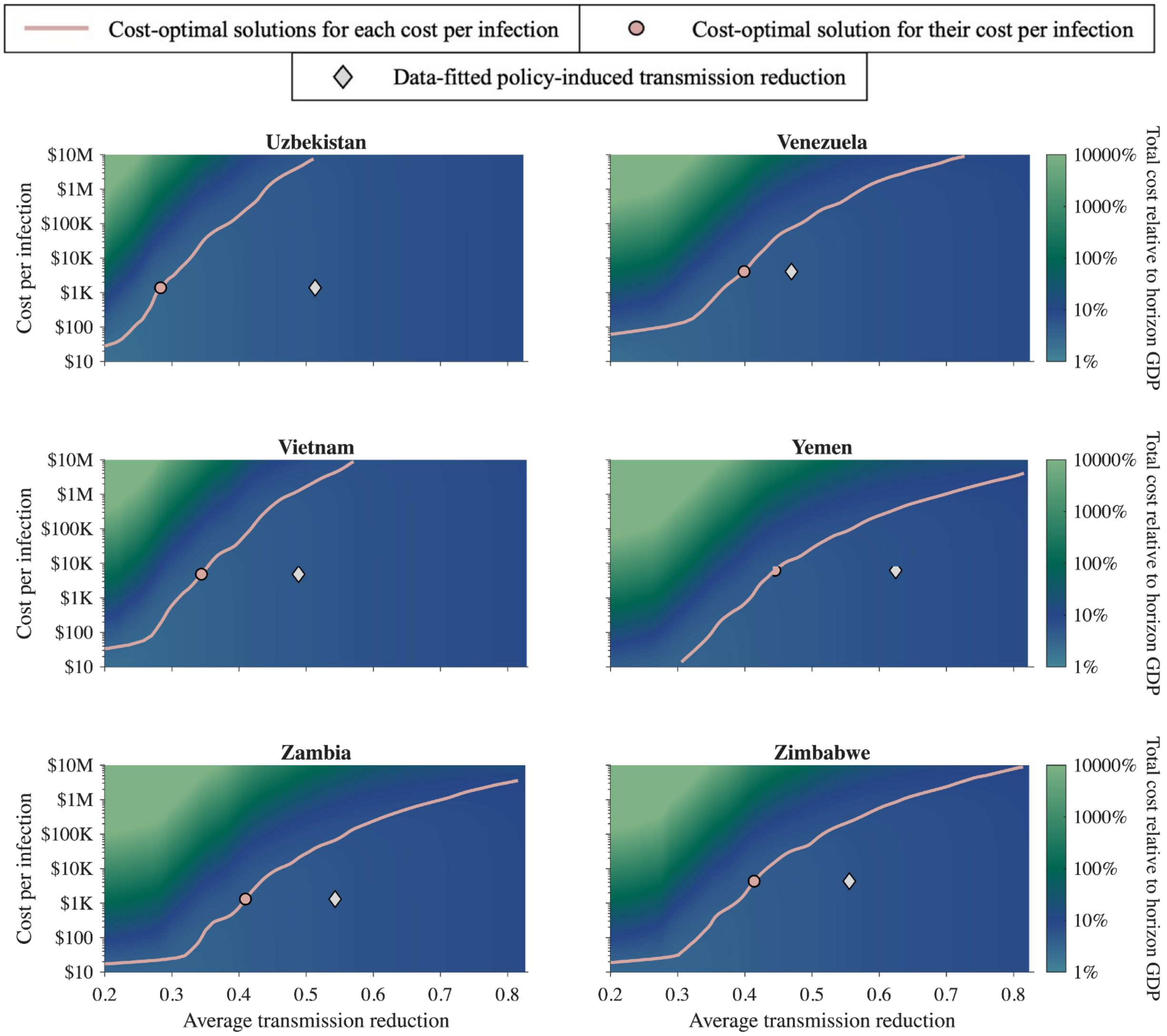


**Figure S4.1.14: The cost-optimal solution under various cost per infection for Uzbekistan, Venezuela, Vietnam, Yemen, Zambia, and Zimbabwe.**

### 4.2. Simulation result for 40 weeks

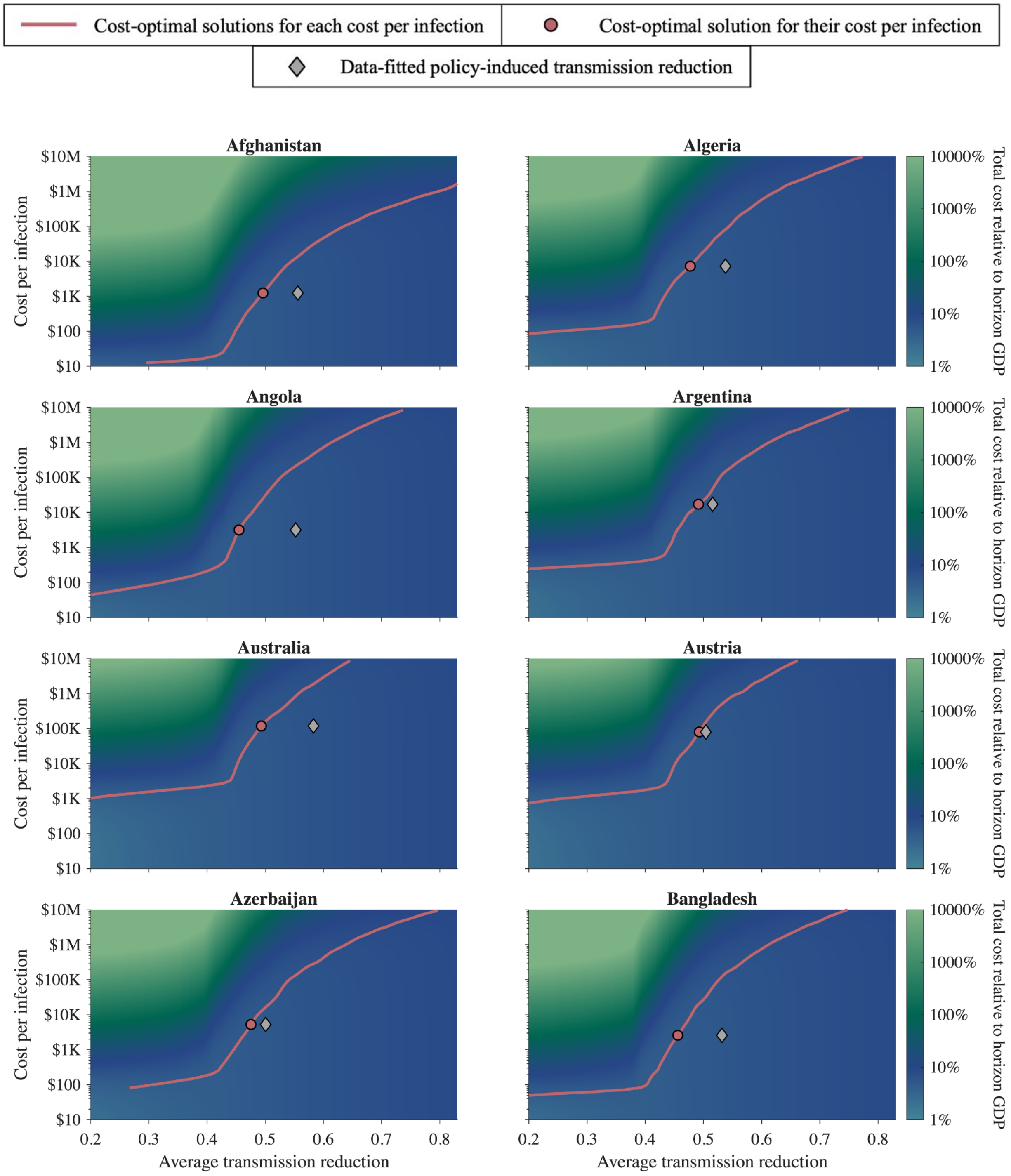


**Figure S4.2.1: The cost-optimal solution under various cost per infection for Afghanistan, Algeria, Angola, Argentina, Australia, Austria, Azerbaijan, and Bangladesh.**

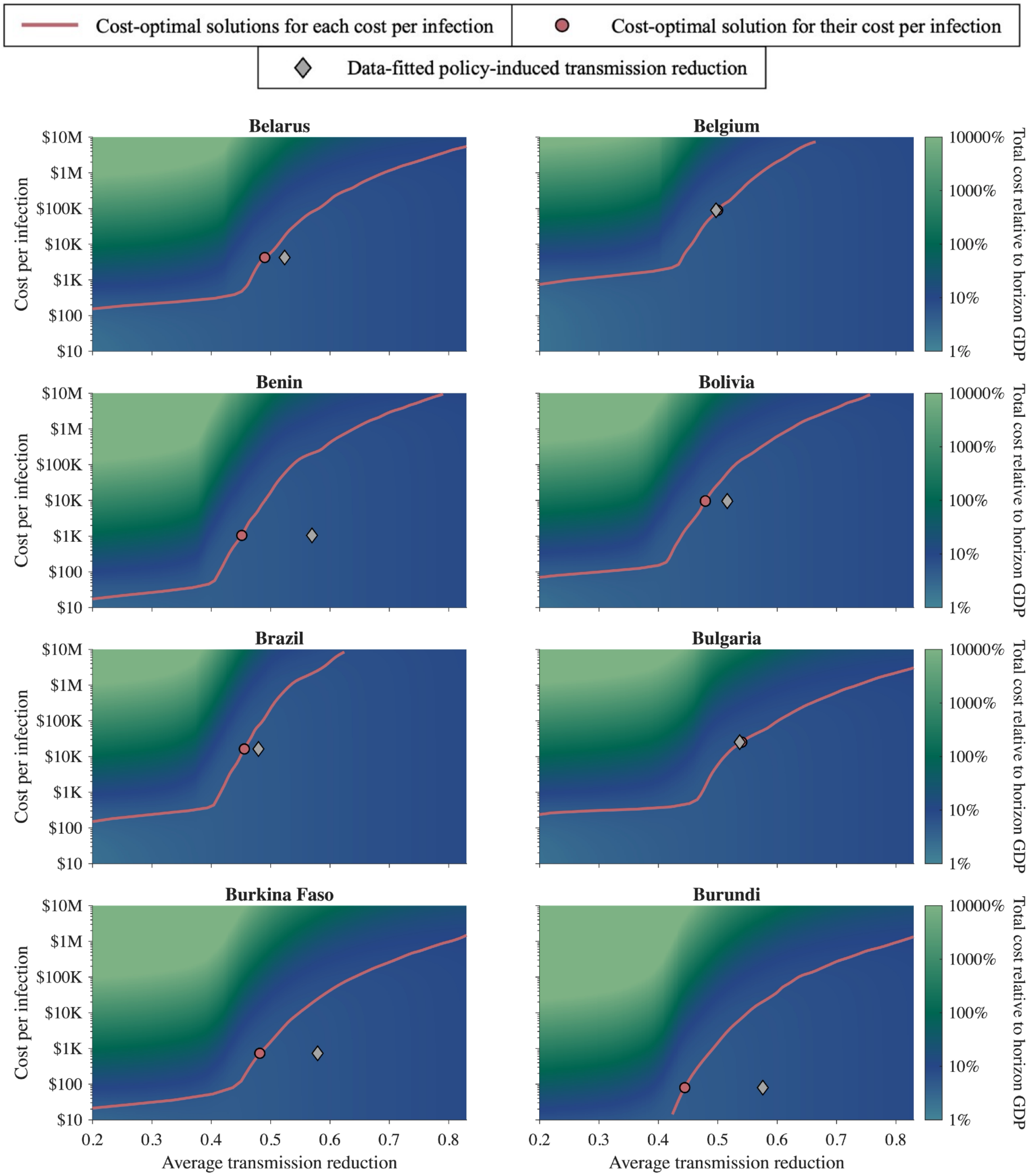


**Figure S4.2.2: The cost-optimal solution under various cost per infection for Belarus, Belgium, Benin, Bolivia, Brazil, Bulgaria, Burkina Faso, and Burundi.**

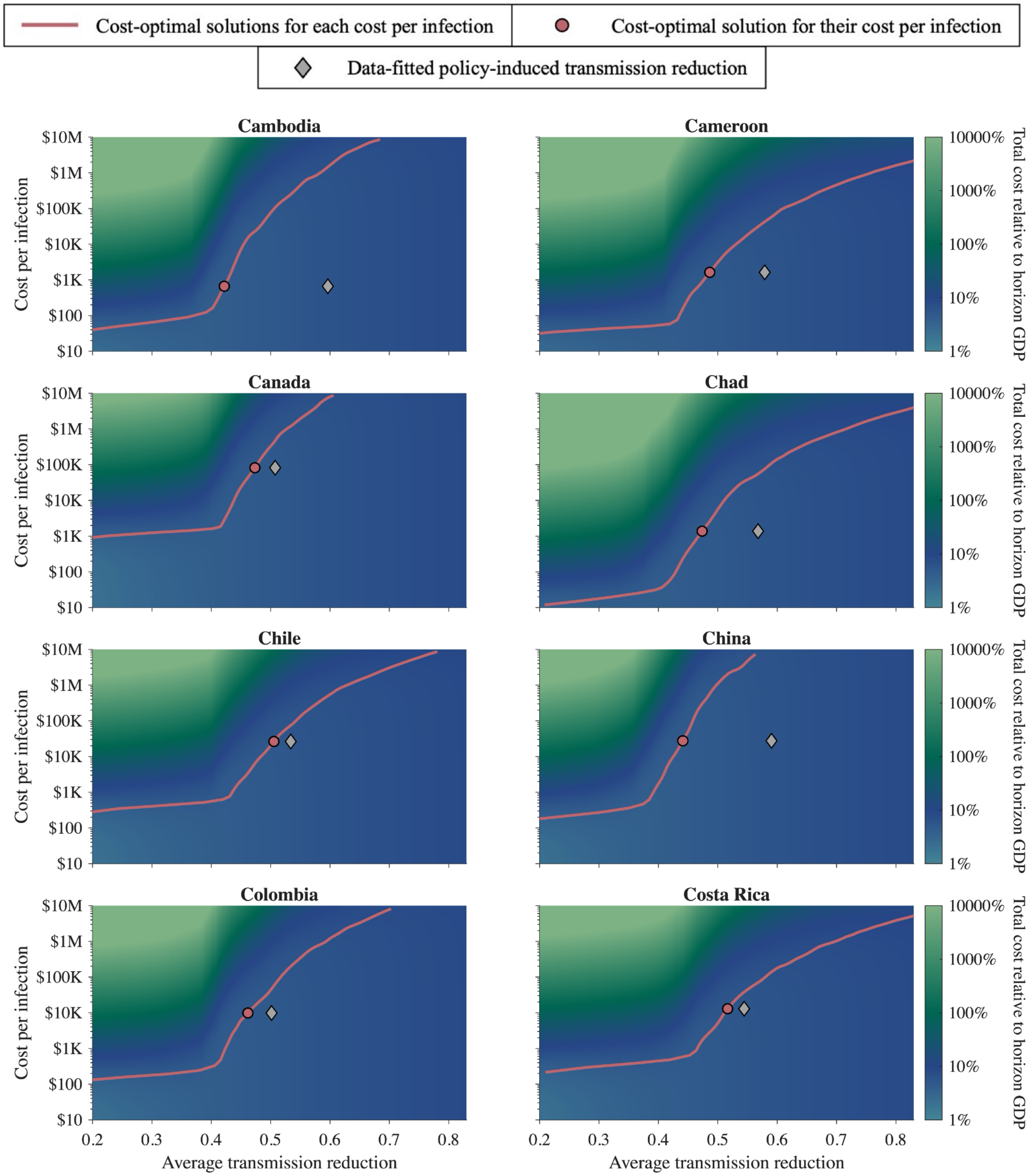


**Figure S4.2.3: The cost-optimal solution under various cost per infection for Cambodia, Cameroon, Canada, Chad, Chile, China, Colombia, and Costa Rica.**

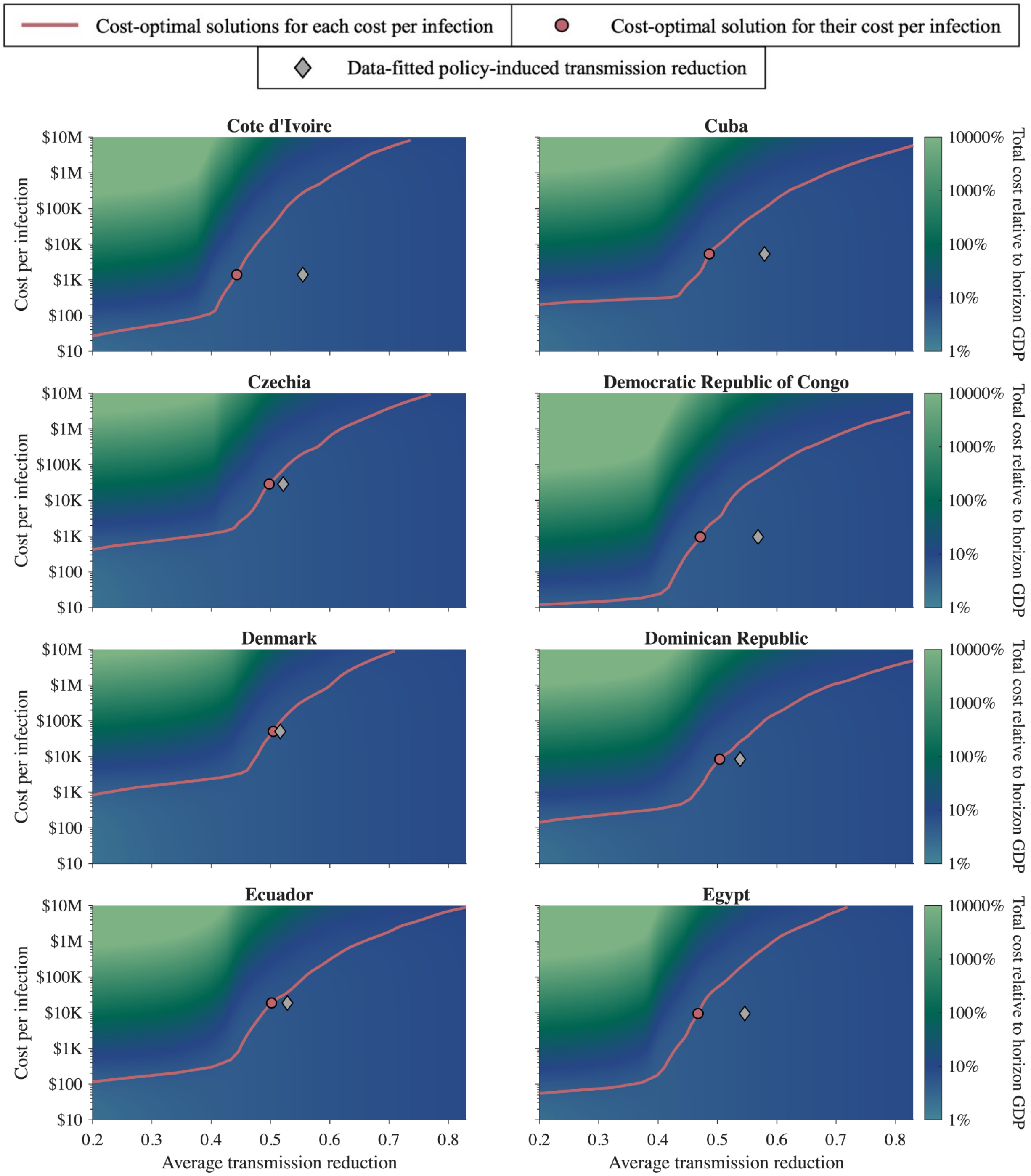


**Figure S4.2.4: The cost-optimal solution under various cost per infection for Cote d'Ivoire, Cuba, Czechia, Democratic Republic of Congo, Denmark, Dominican Republic, Ecuador, and Egypt.**

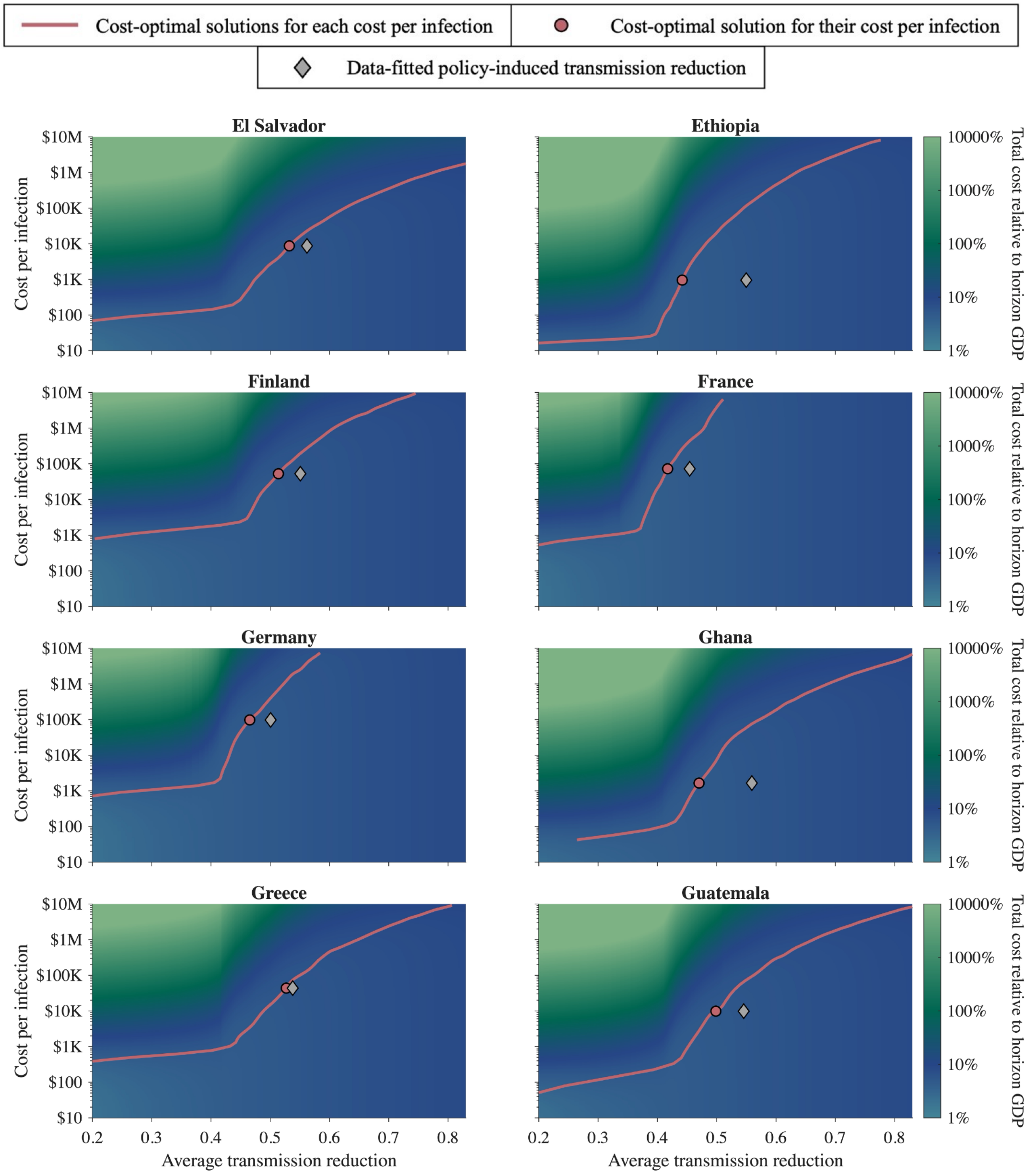


**Figure S4.2.5: The cost-optimal solution under various cost per infection for El Salvador, Ethiopia, Finland, France, Germany, Ghana, Greece, and Guatemala.**

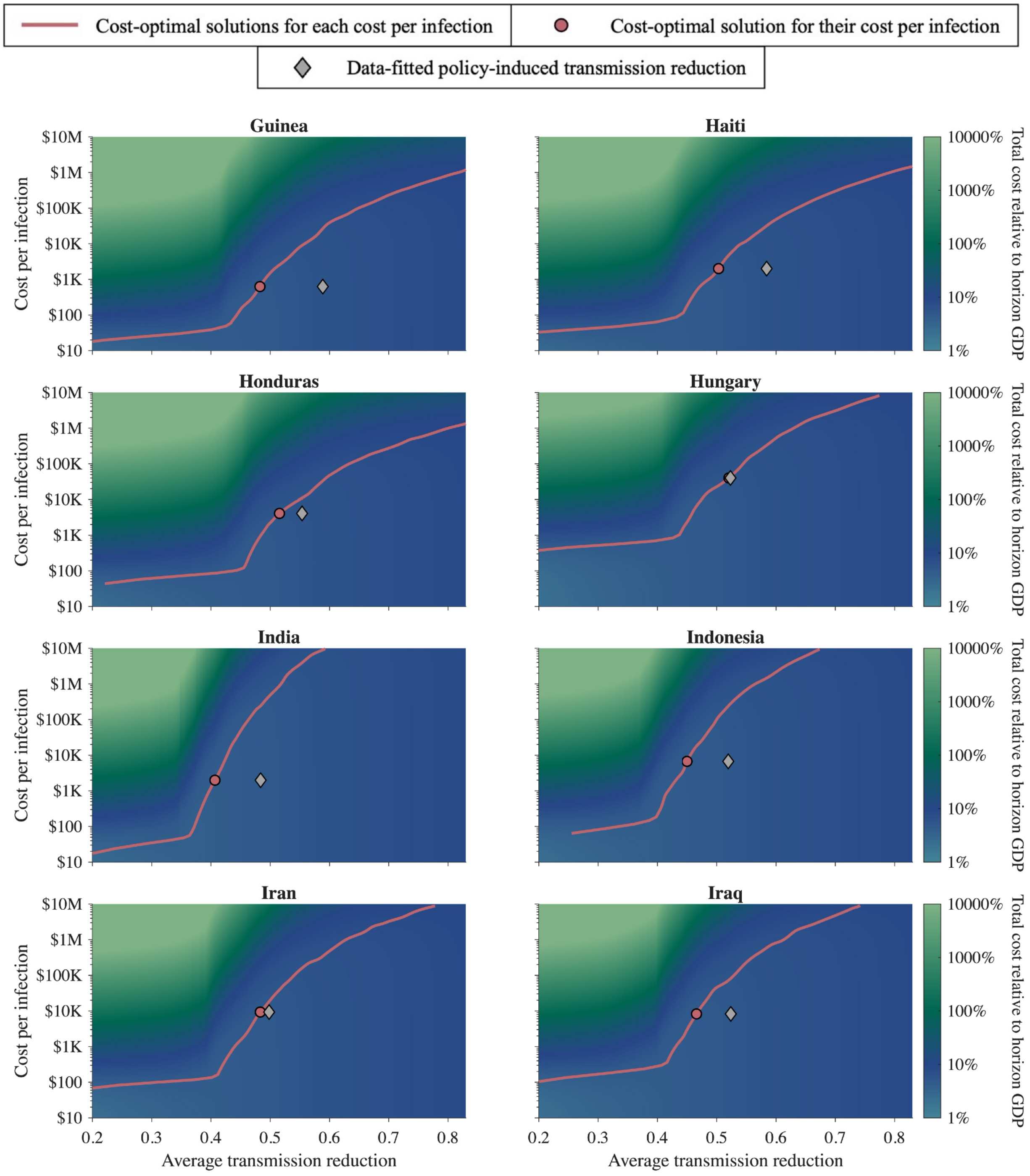


**Figure S4.2.6: The cost-optimal solution under various cost per infection for Guinea, Haiti, Honduras, Hungary, India, Indonesia, Iran, and Iraq.**

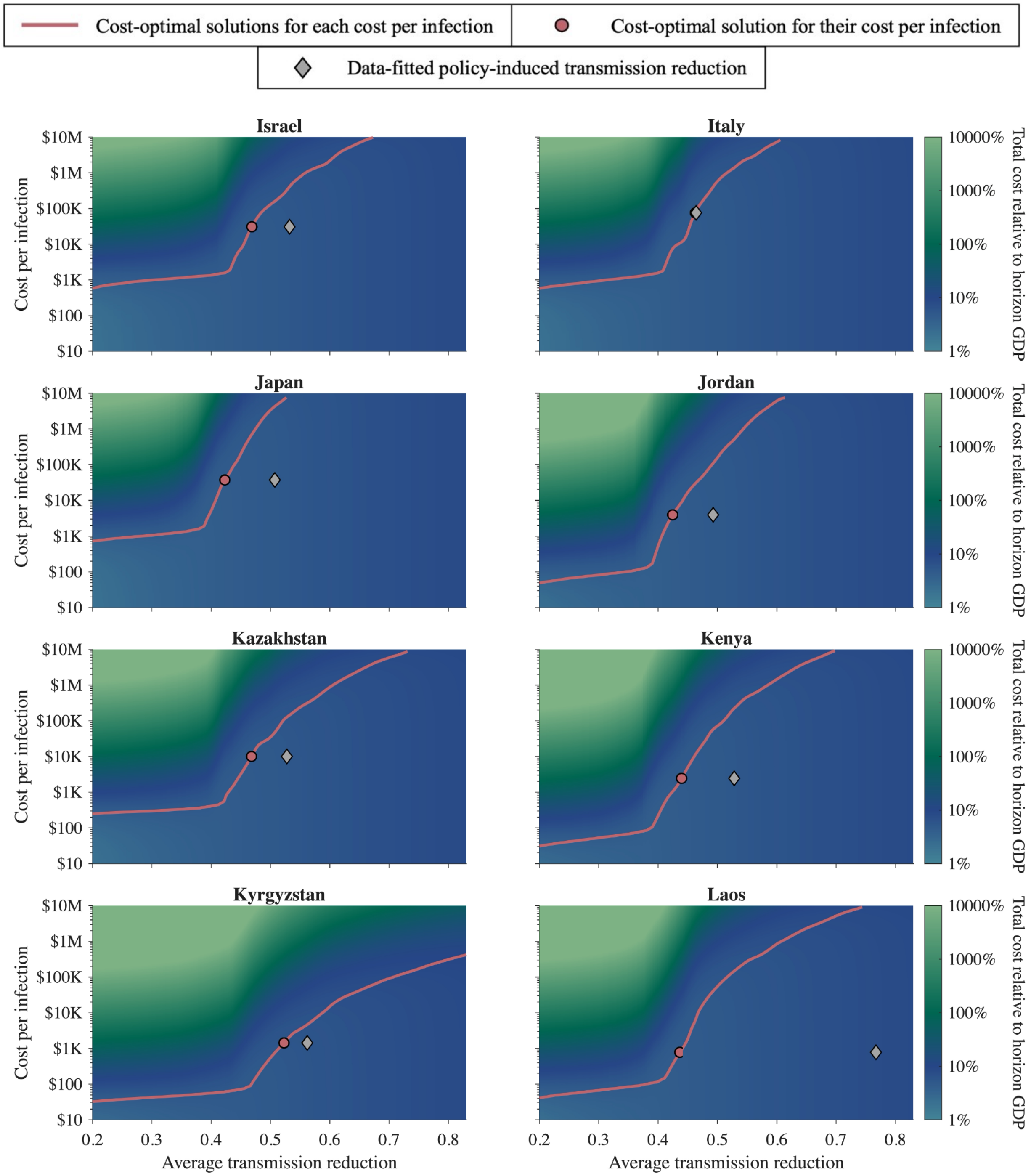


**Figure S4.2.7: The cost-optimal solution under various cost per infection for Israel, Italy, Japan, Jordan, Kazakhstan, Kenya, Kyrgyzstan, and Laos.**

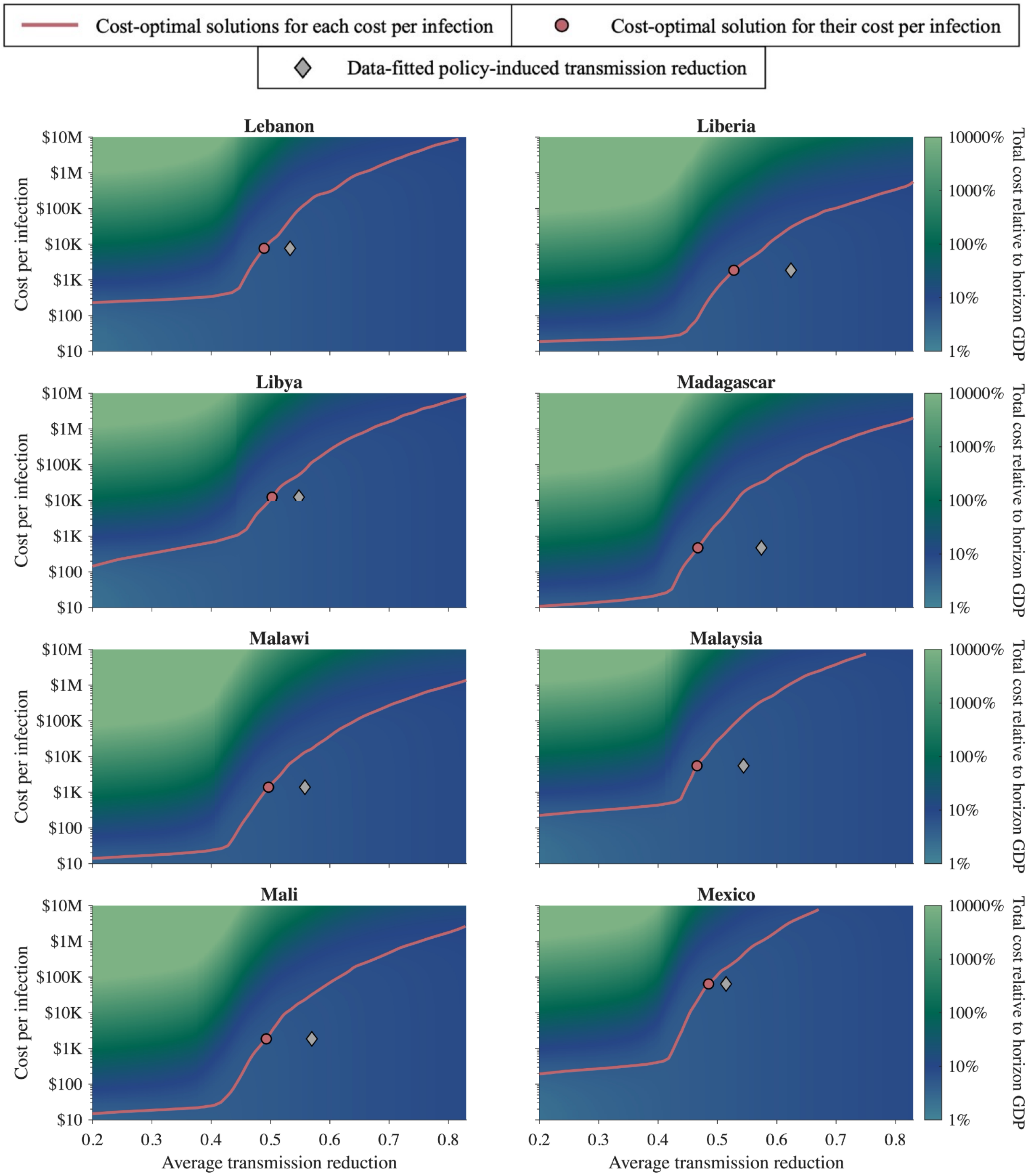


**Figure S4.2.8: The cost-optimal solution under various cost per infection for Lebanon, Liberia, Libya, Madagascar, Malawi, Malaysia, Mali, and Mexico.**

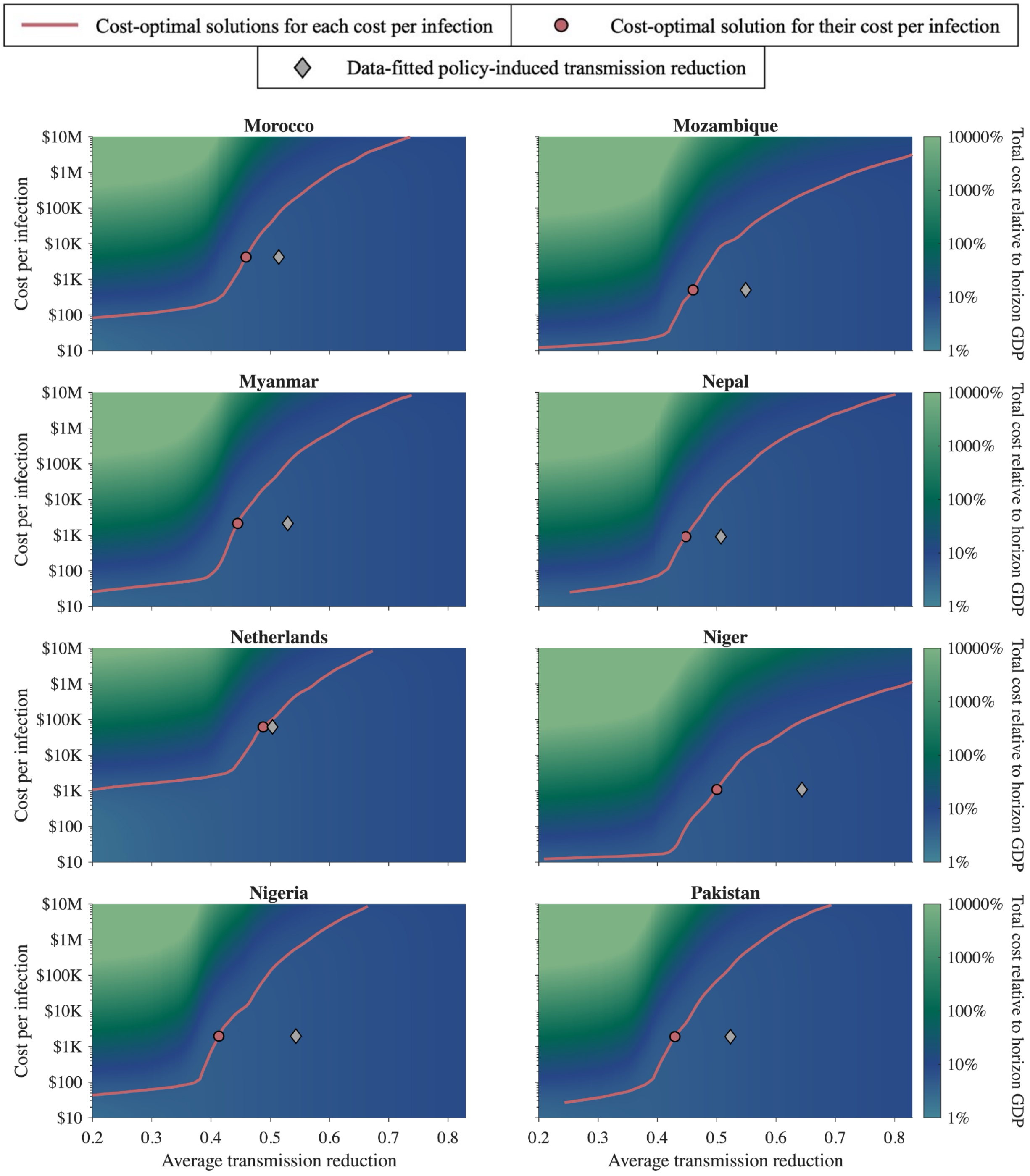


**Figure S4.2.9: The cost-optimal solution under various cost per infection for Morocco, Mozambique, Myanmar, Nepal, Netherlands, Niger, Nigeria, and Pakistan.**

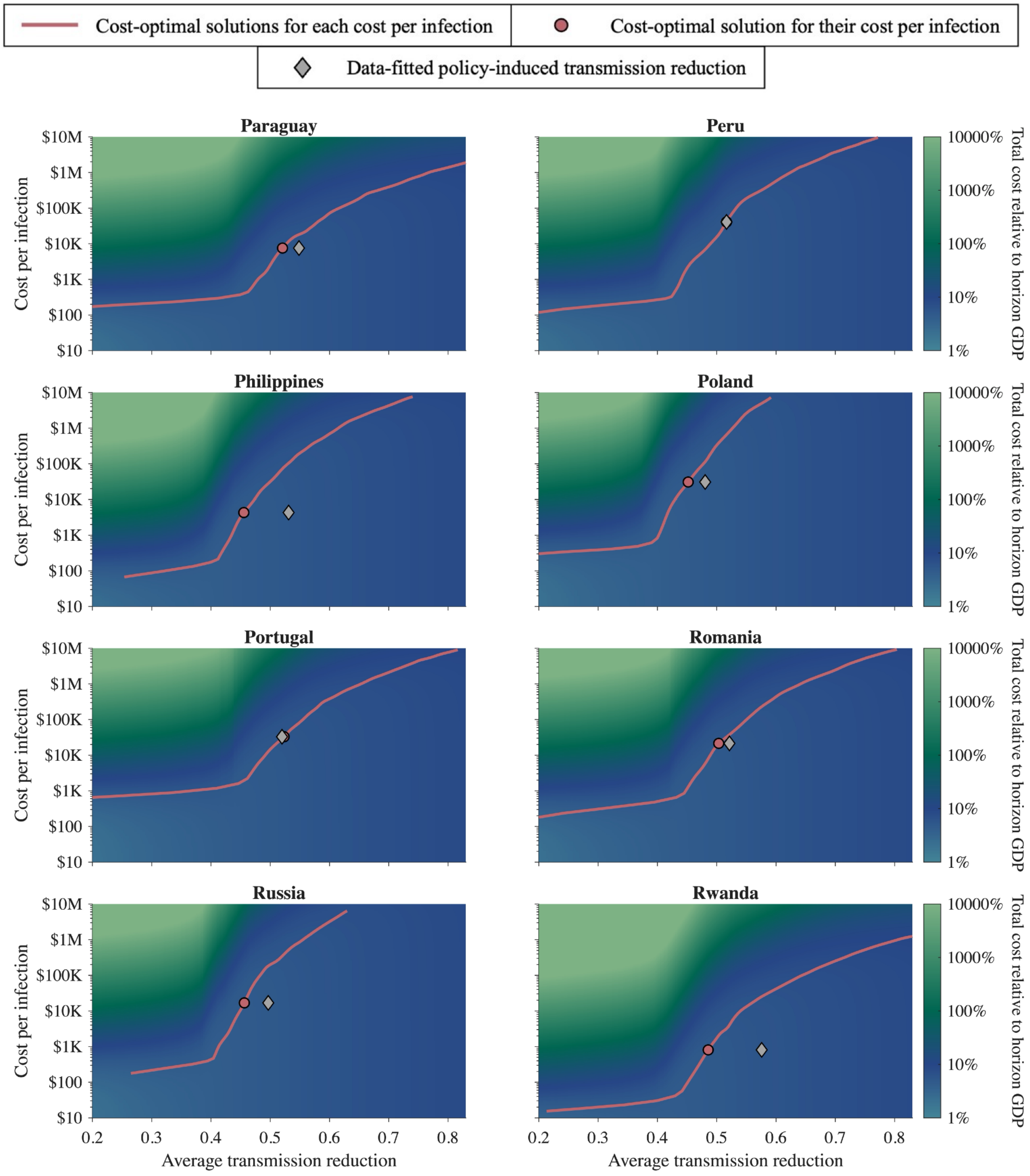


**Figure S4.2.10: The cost-optimal solution under various cost per infection for Paraguay, Peru, Philippines, Poland, Portugal, Romania, Russia, and Rwanda.**

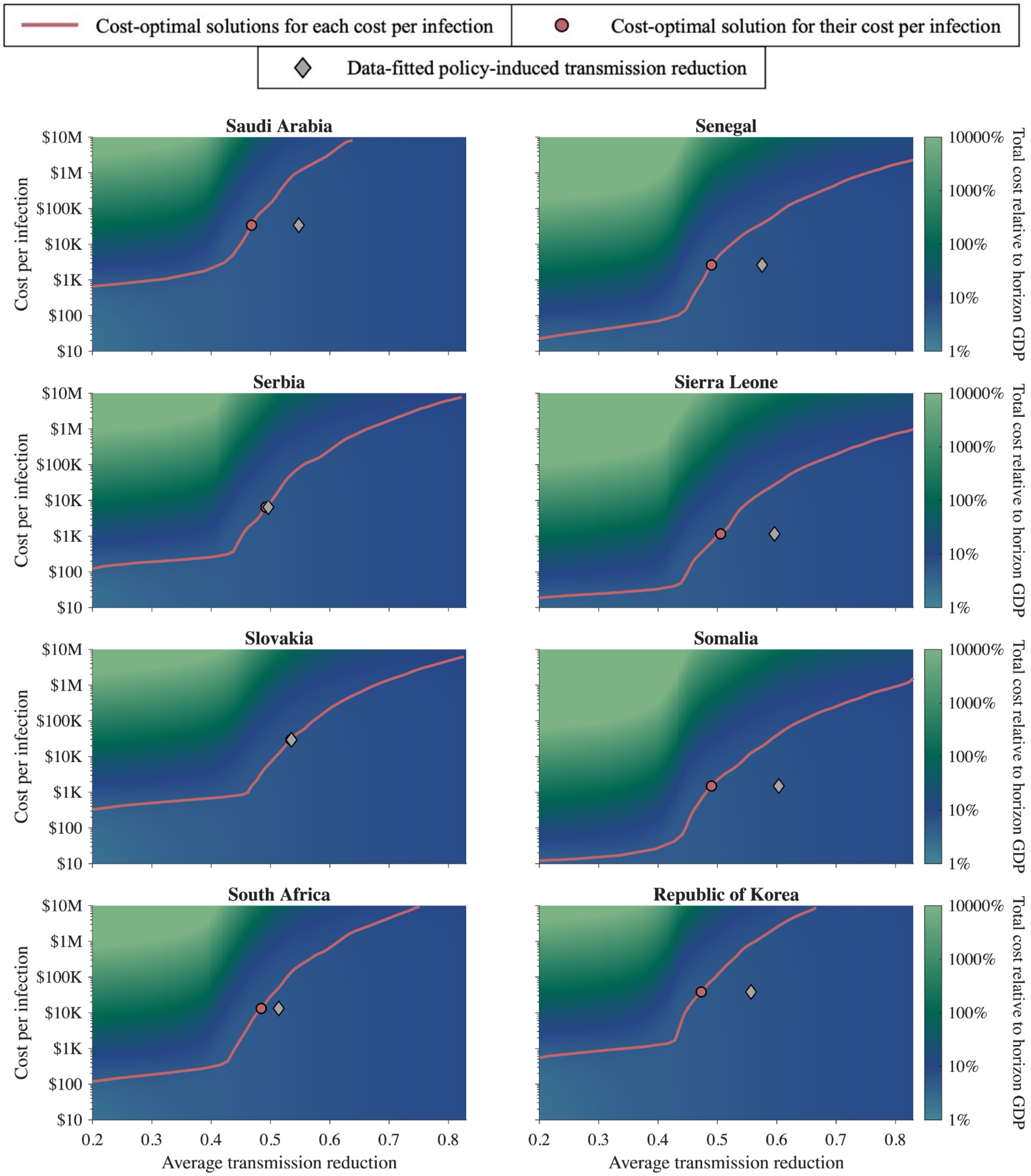


**Figure S4.2.11: The cost-optimal solution under various cost per infection for Saudi Arabia, Senegal, Serbia, Sierra Leone, Slovakia, Somalia, South Africa, and Republic of Korea.**

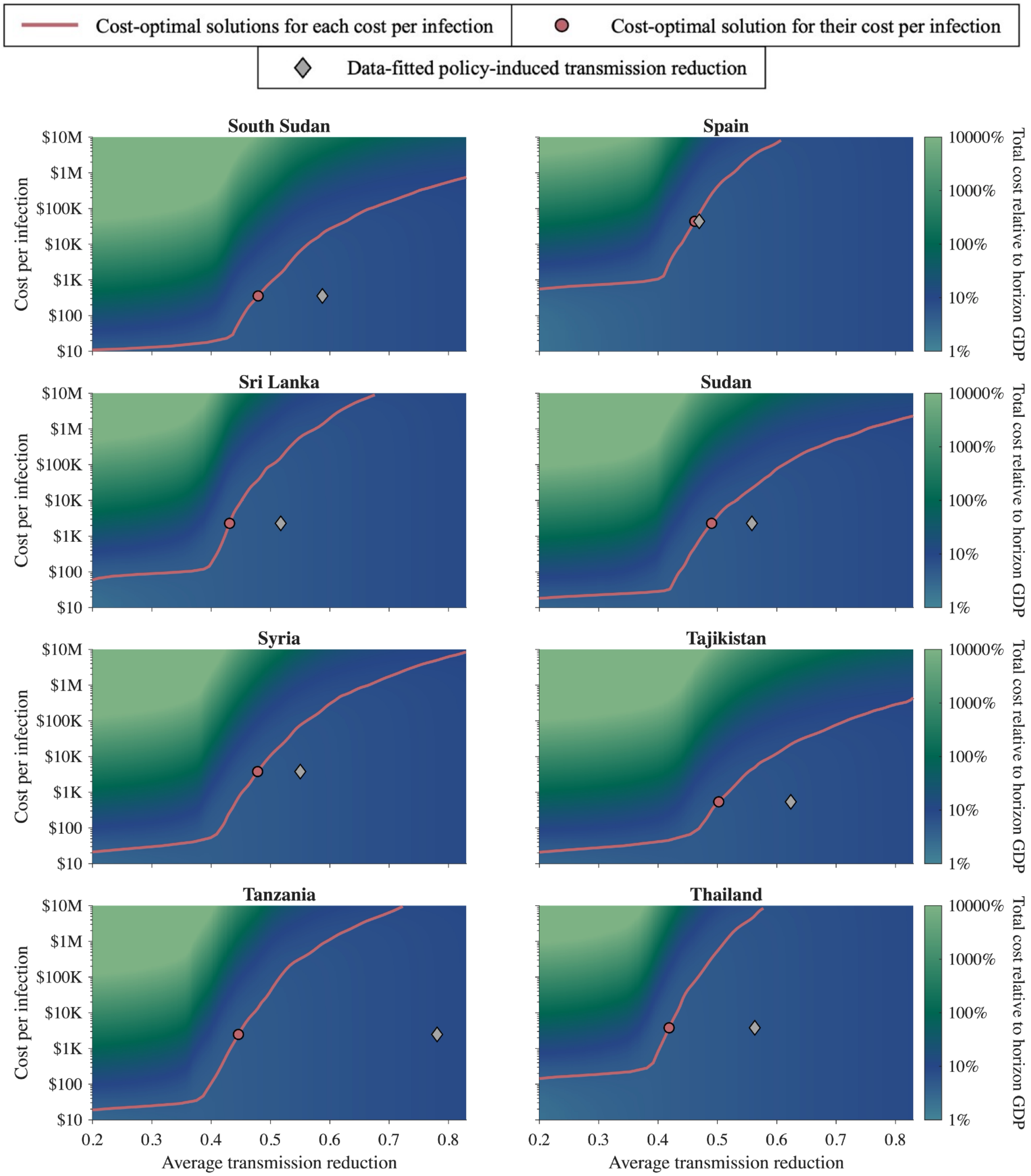


**Figure S4.2.12: The cost-optimal solution under various cost per infection for South Sudan, Spain, Sri Lanka, Sudan, Syria, Tajikistan, Tanzania, and Thailand.**

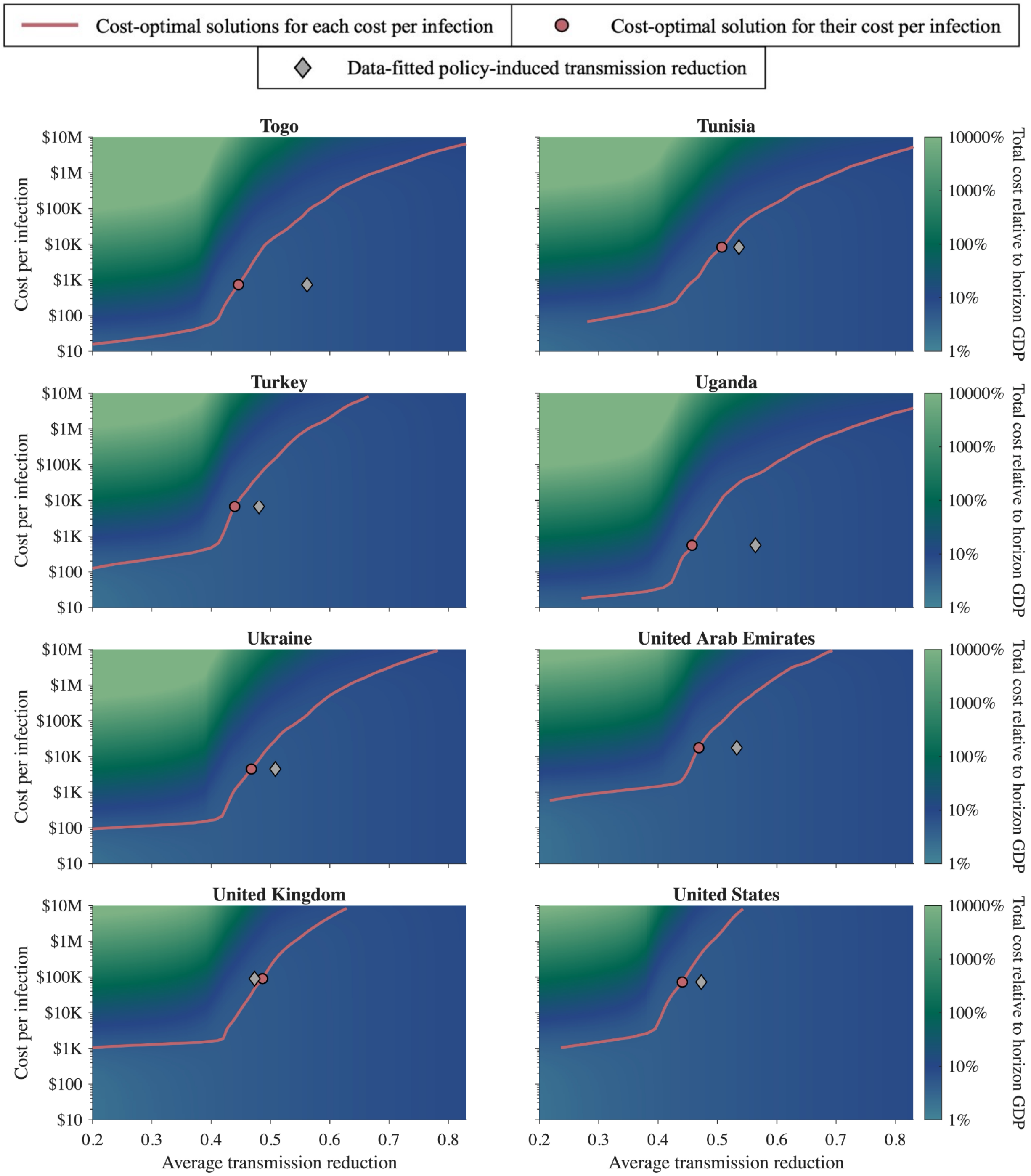


**Figure S4.2.13: The cost-optimal solution under various cost per infection for Togo, Tunisia, Turkey, Uganda, Ukraine, United Arab Emirates, United Kingdom, and United States.**

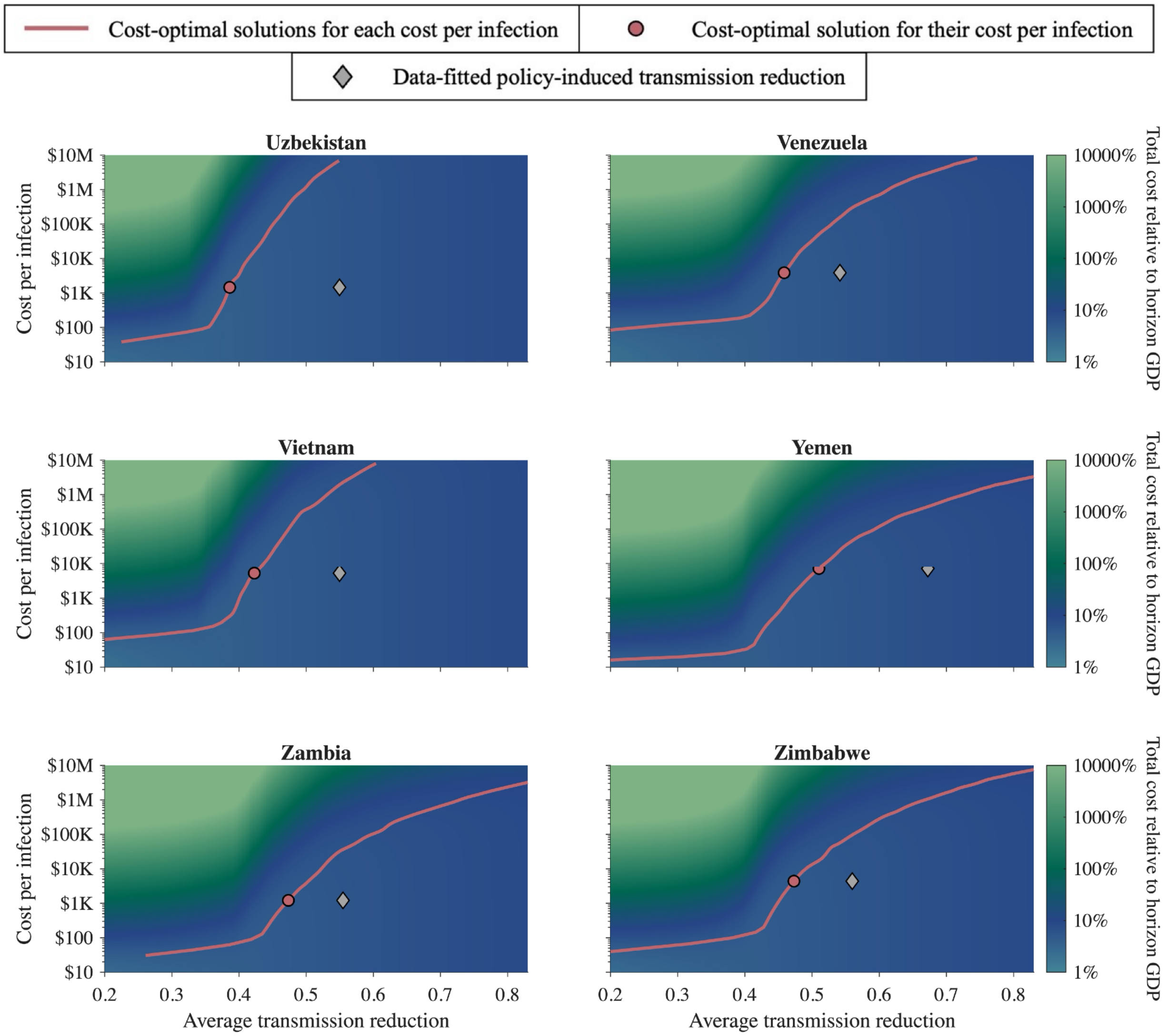


**Figure S4.2.14: The cost-optimal solution under various cost per infection for Uzbekistan, Venezuela, Vietnam, Yemen, Zambia, and Zimbabwe.**

#### 4.3. Simulation result for 52 weeks

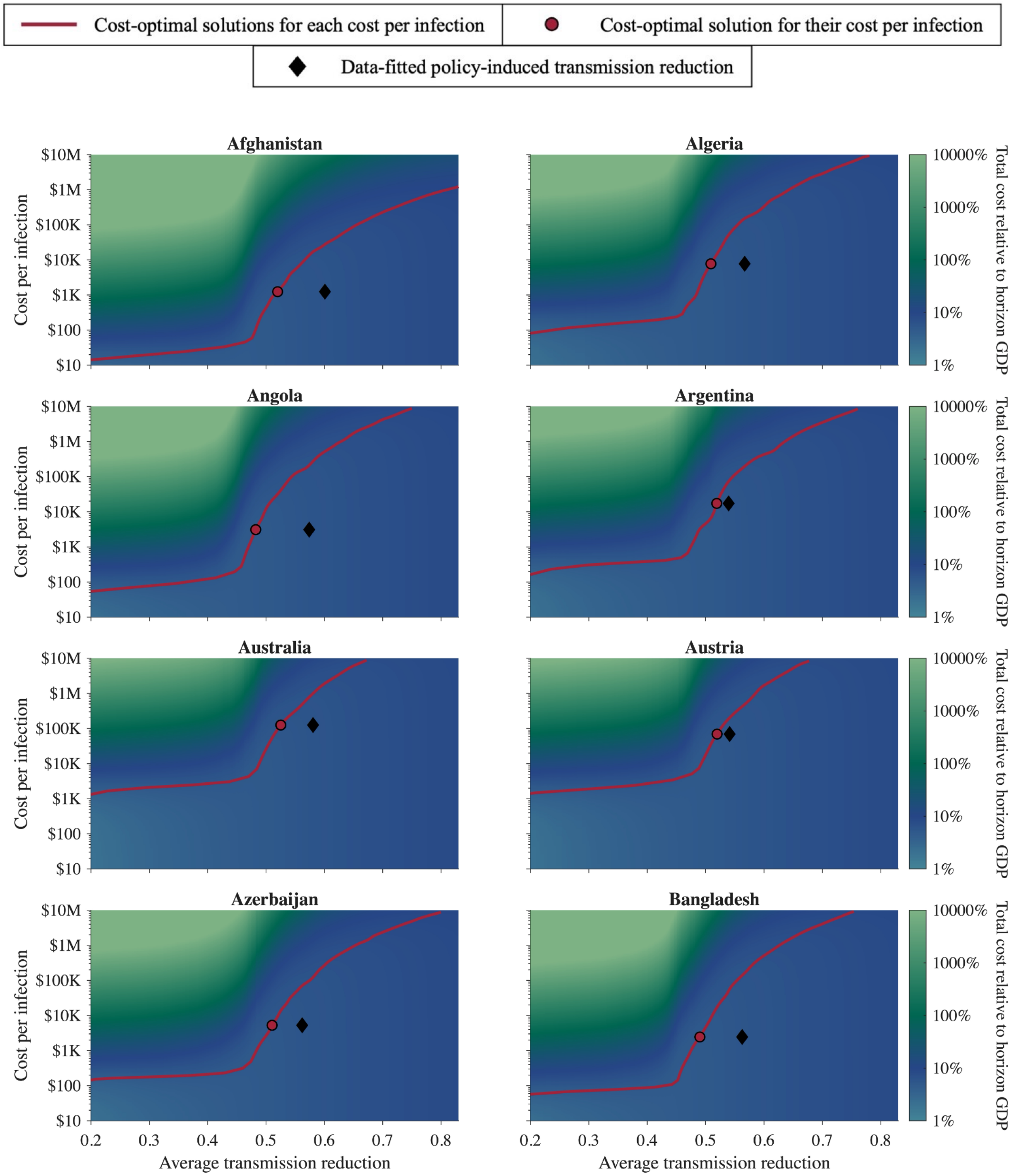


**Figure S4.3.1: The cost-optimal solution under various cost per infection for Afghanistan, Algeria, Angola, Argentina, Australia, Austria, Azerbaijan, and Bangladesh.**

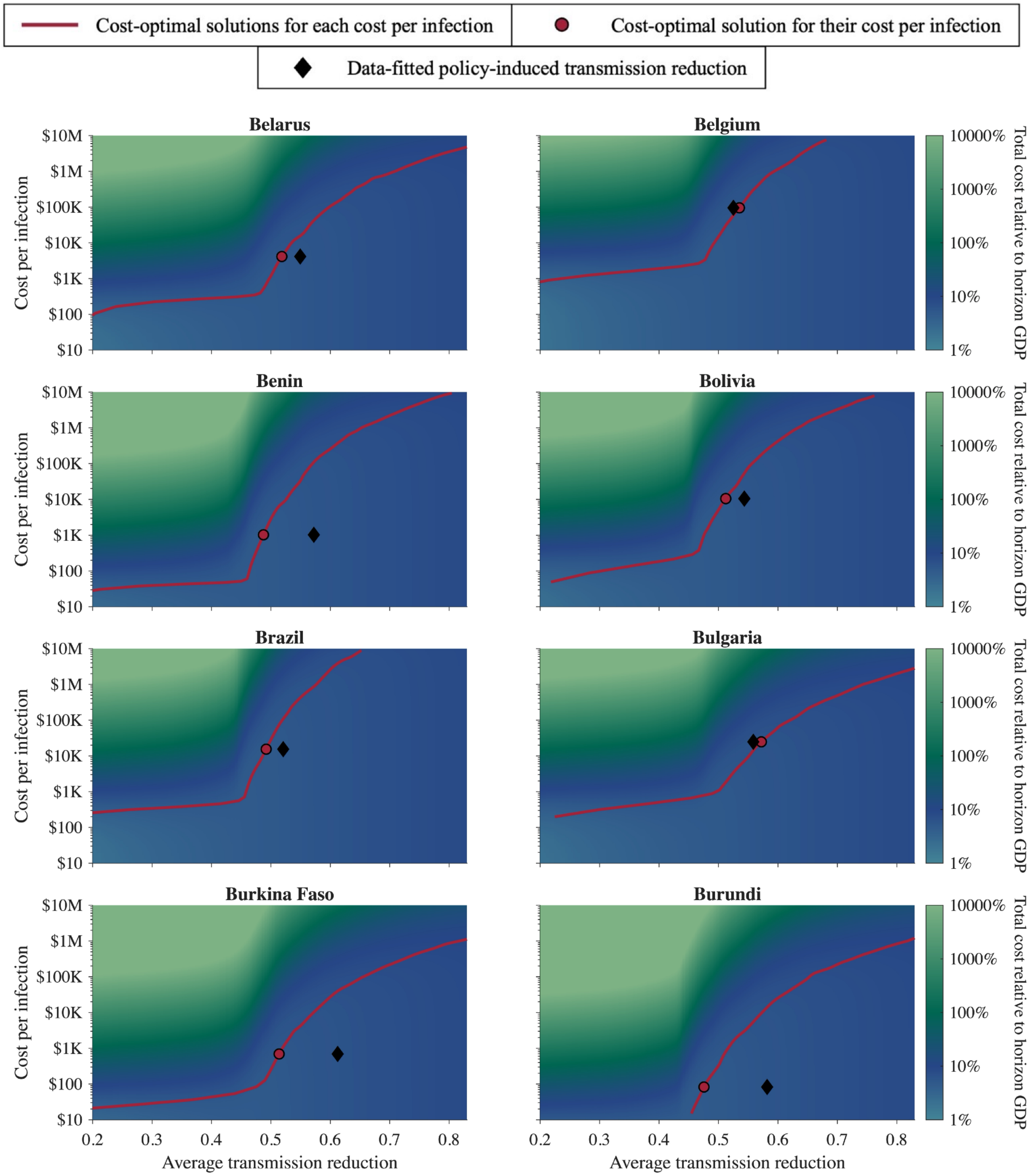


**Figure S4.3.2: The cost-optimal solution under various cost per infection for Belarus, Belgium, Benin, Bolivia, Brazil, Bulgaria, Burkina Faso, and Burundi.**

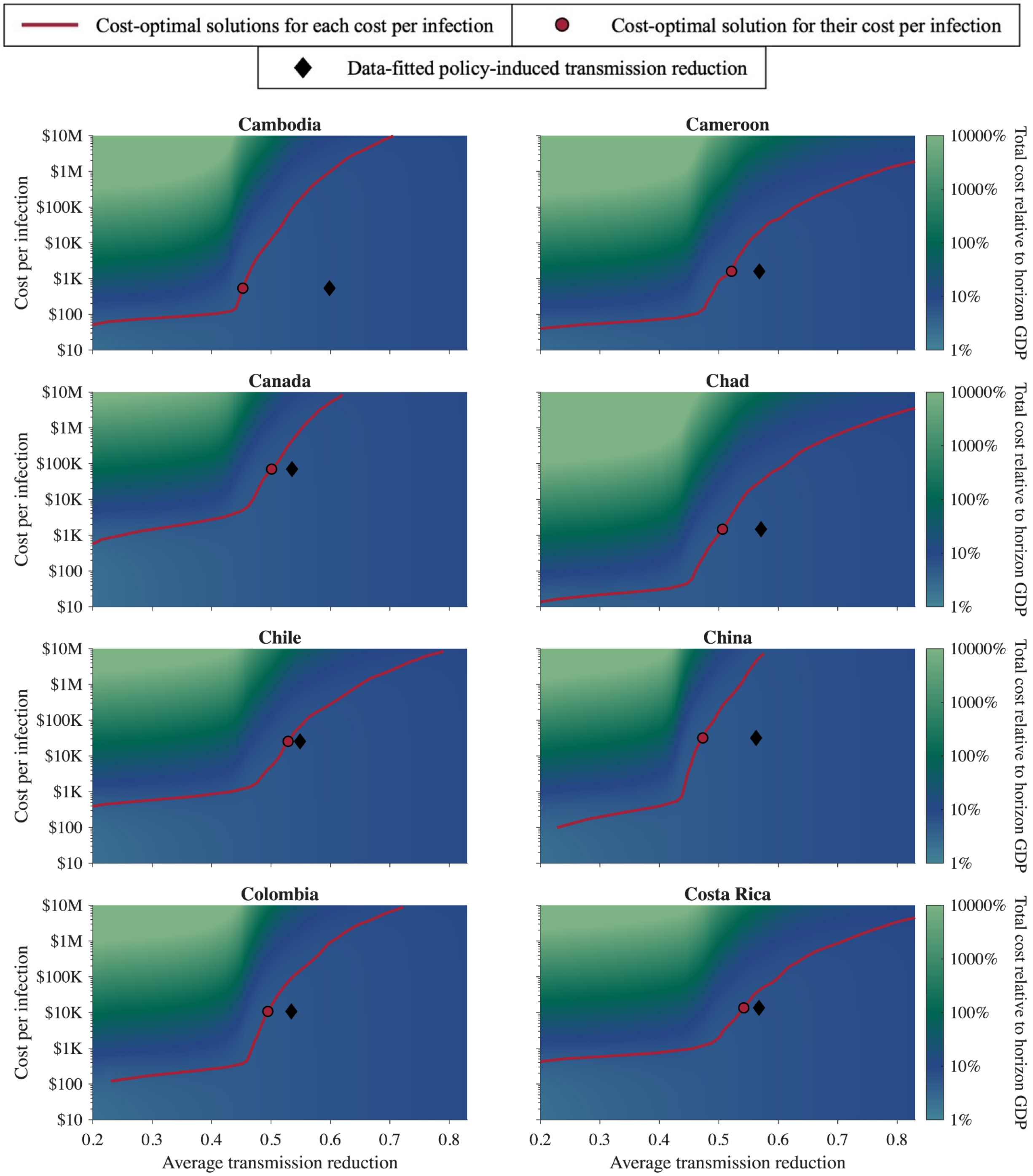


**Figure S4.3.3: The cost-optimal solution under various cost per infection for Cambodia, Cameroon, Canada, Chad, Chile, China, Colombia, and Costa Rica.**

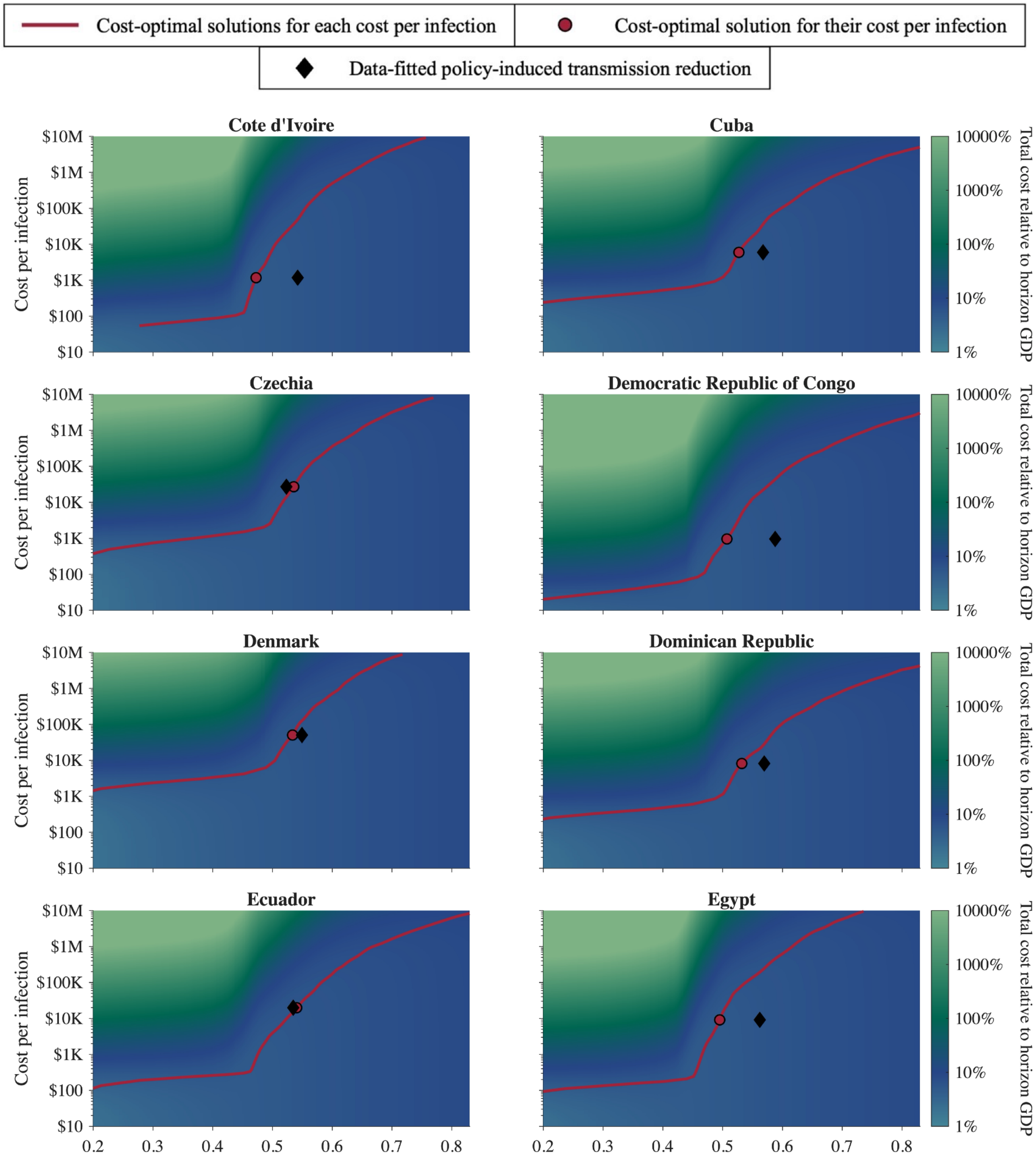


**Figure S4.3.4: The cost-optimal solution under various cost per infection for Cote d'Ivoire, Cuba, Czechia, Democratic Republic of Congo, Denmark, Dominican Republic, Ecuador, and Egypt.**

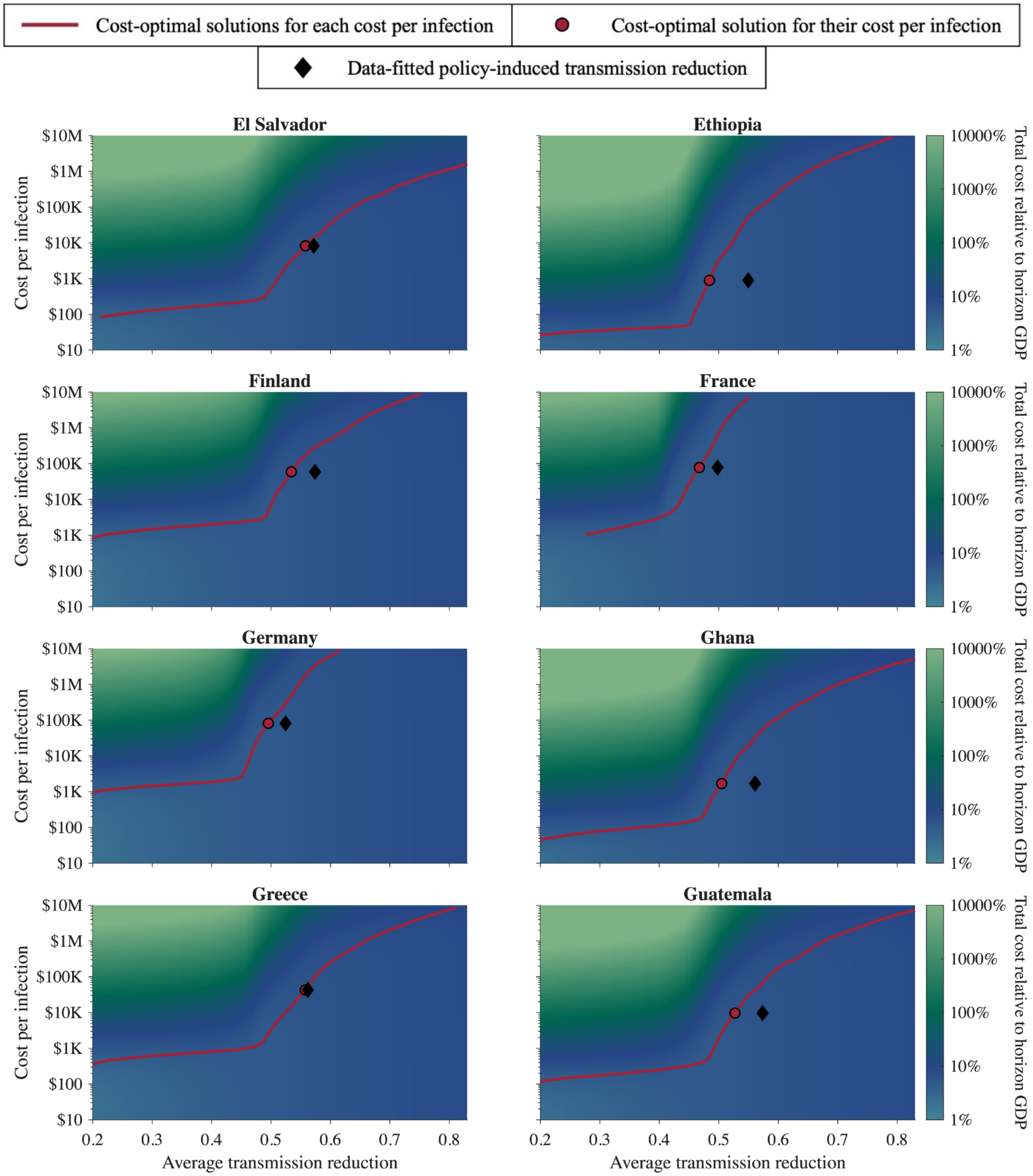


**Figure S4.3.5: The cost-optimal solution under various cost per infection for El Salvador, Ethiopia, Finland, France, Germany, Ghana, Greece, and Guatemala.**

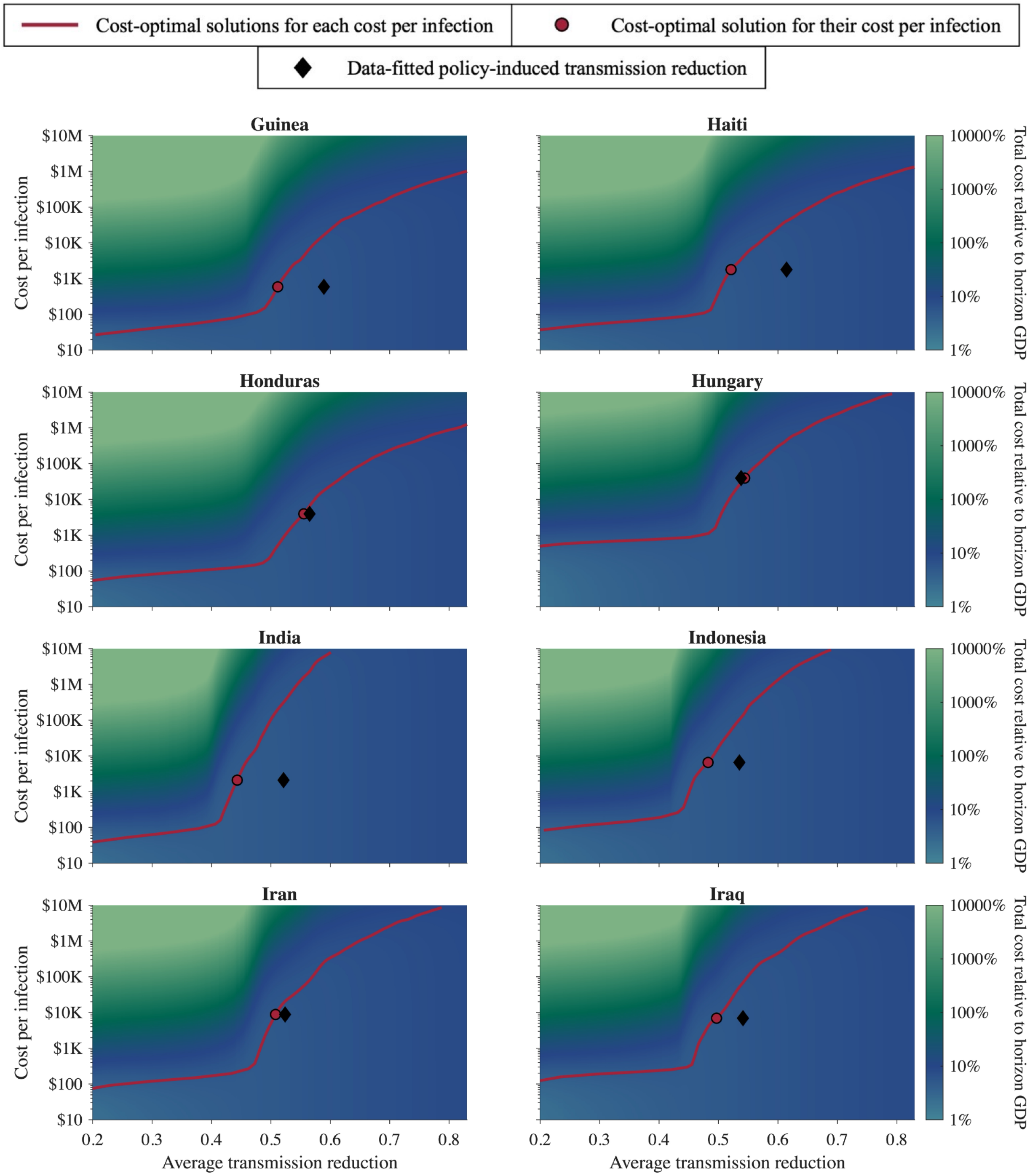


**Figure S4.3.6: The cost-optimal solution under various cost per infection for Guinea, Haiti, Honduras, Hungary, India, Indonesia, Iran, and Iraq.**

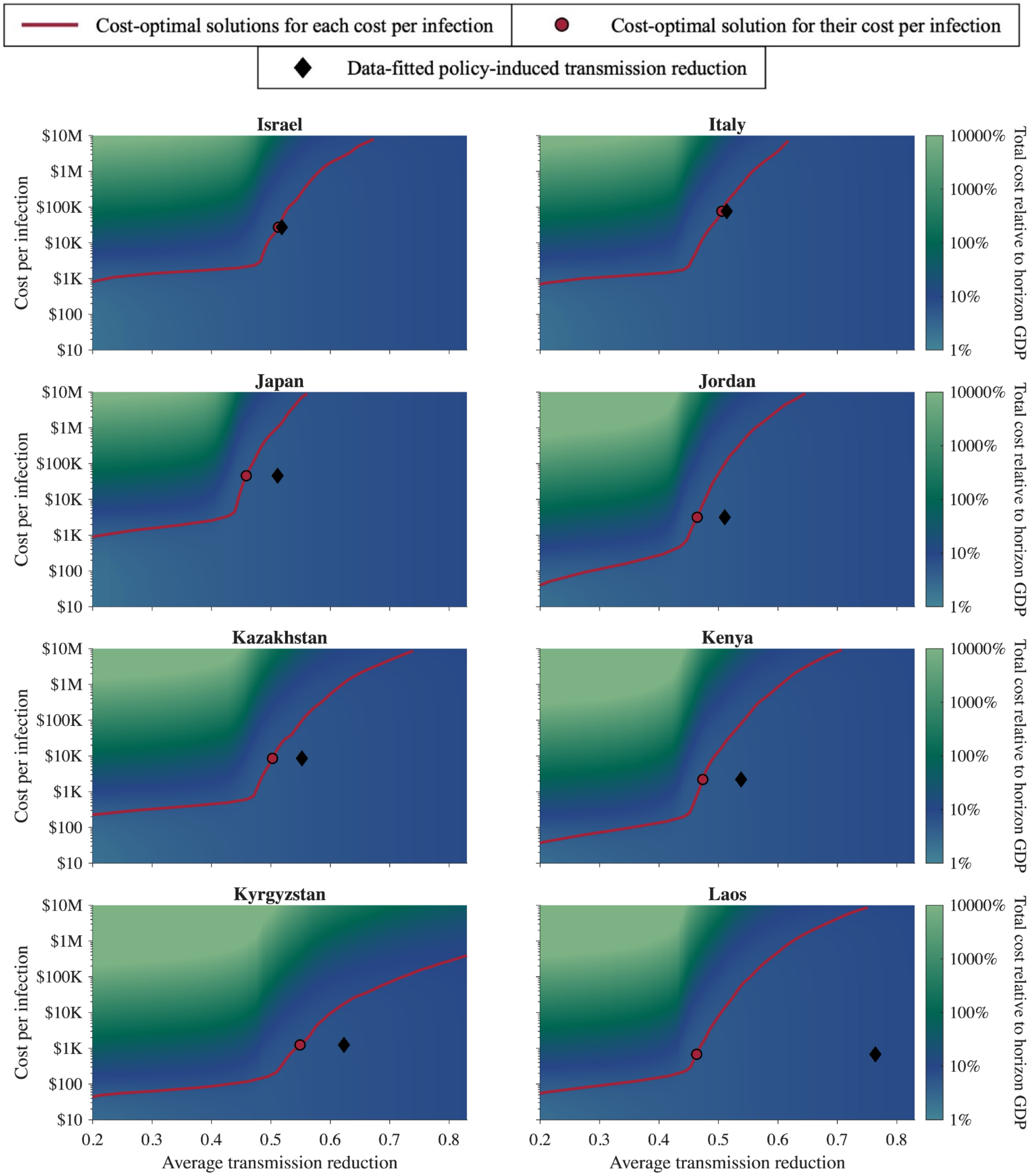


**Figure S4.3.7: The cost-optimal solution under various cost per infection for Israel, Italy, Japan, Jordan, Kazakhstan, Kenya, Kyrgyzstan, and Laos.**

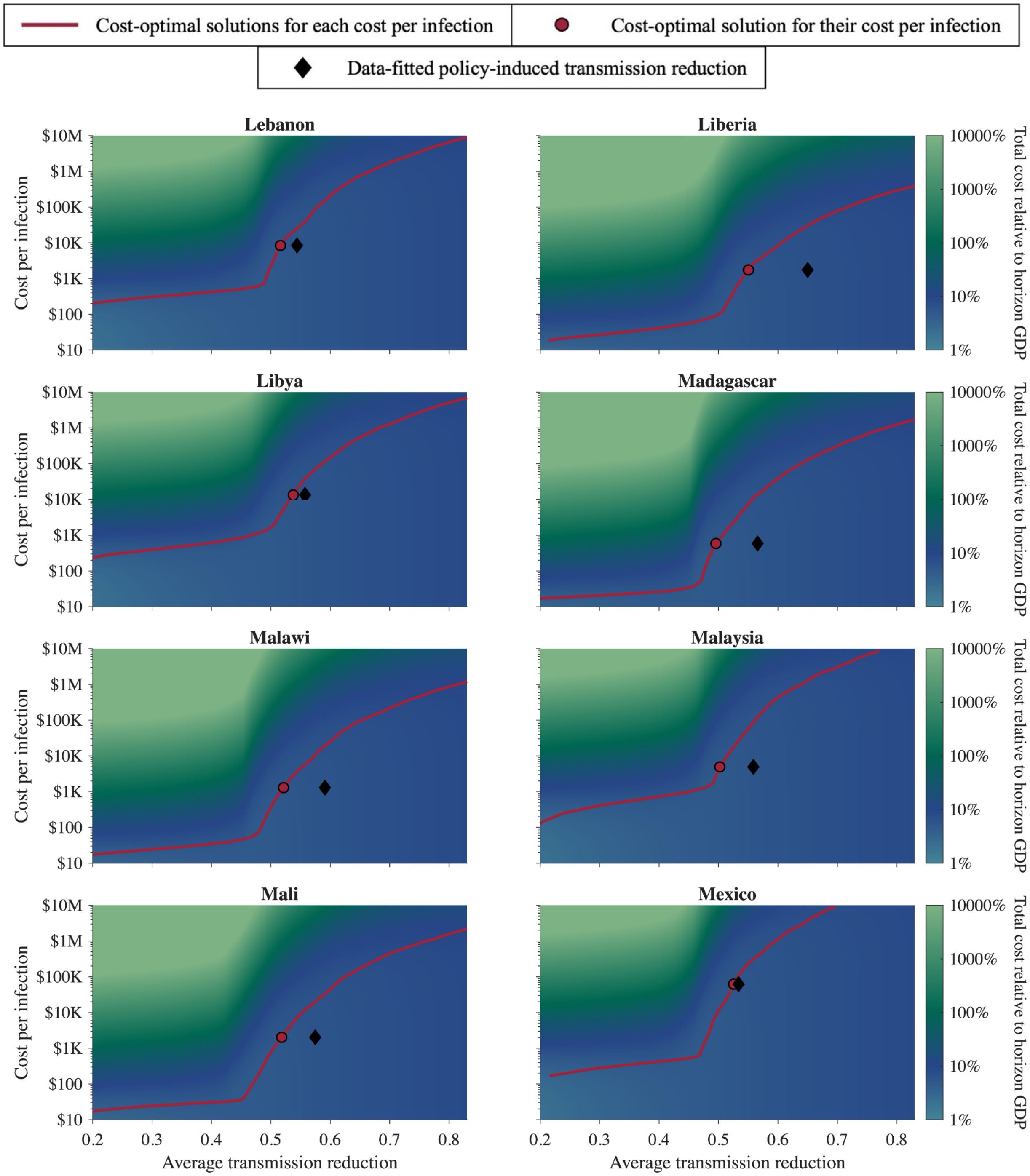


**Figure S4.3.8: The cost-optimal solution under various cost per infection for Lebanon, Liberia, Libya, Madagascar, Malawi, Malaysia, Mali, and Mexico.**

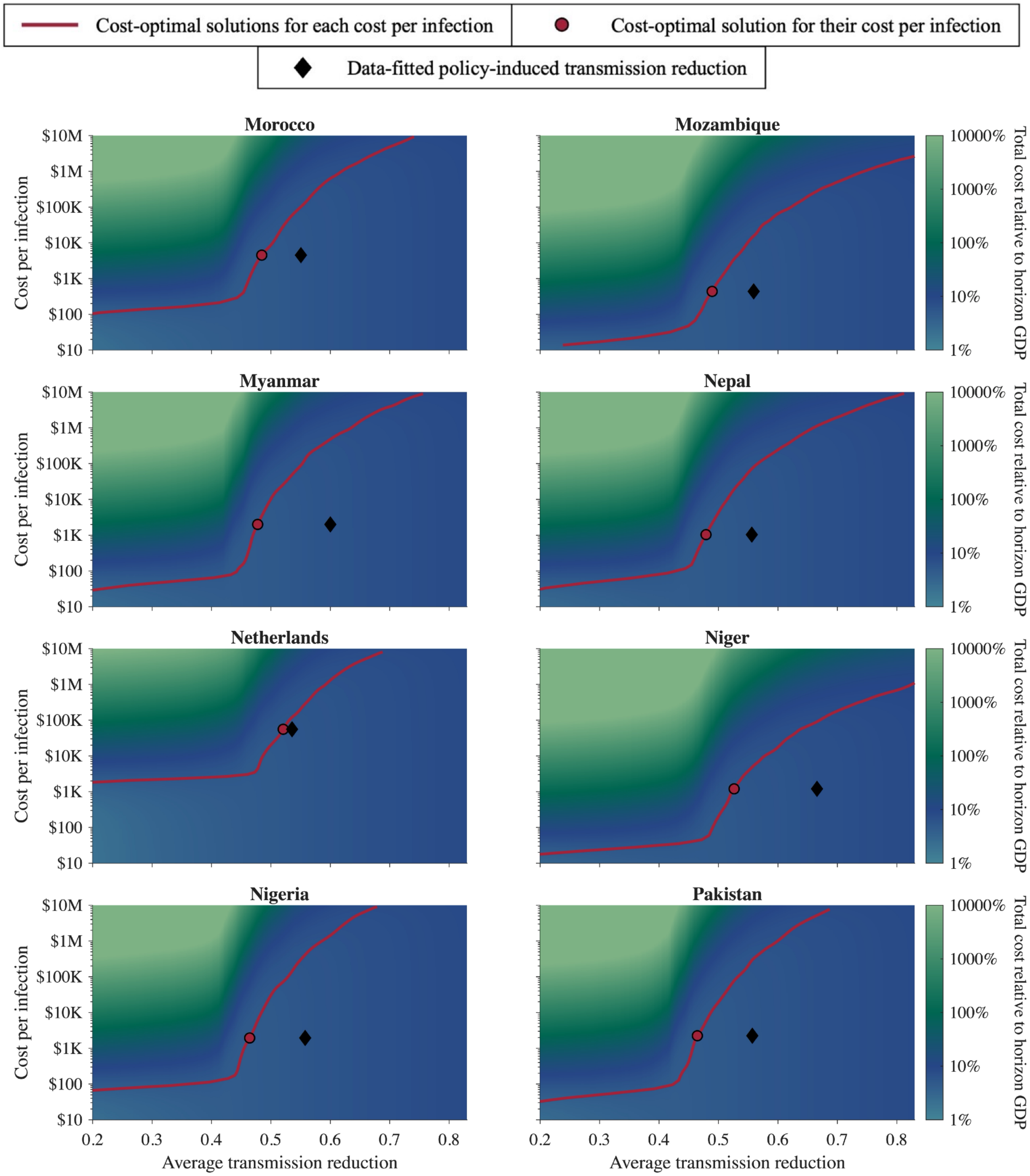


**Figure S4.3.9: The cost-optimal solution under various cost per infection for Morocco, Mozambique, Myanmar, Nepal, Netherlands, Niger, Nigeria, and Pakistan.**

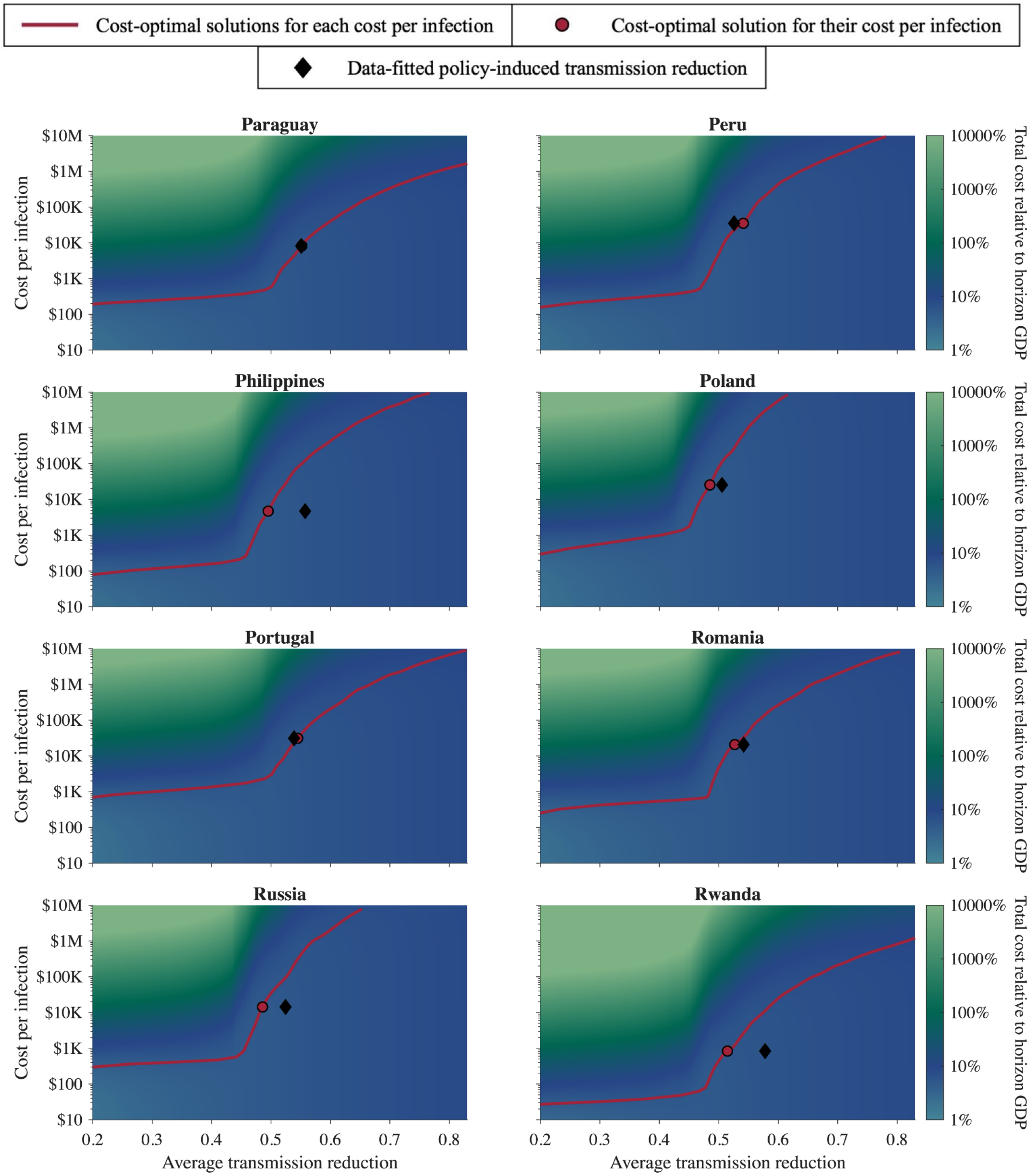


**Figure S4.3.10: The cost-optimal solution under various cost per infection for Paraguay, Peru, Philippines, Poland, Portugal, Romania, Russia, and Rwanda.**

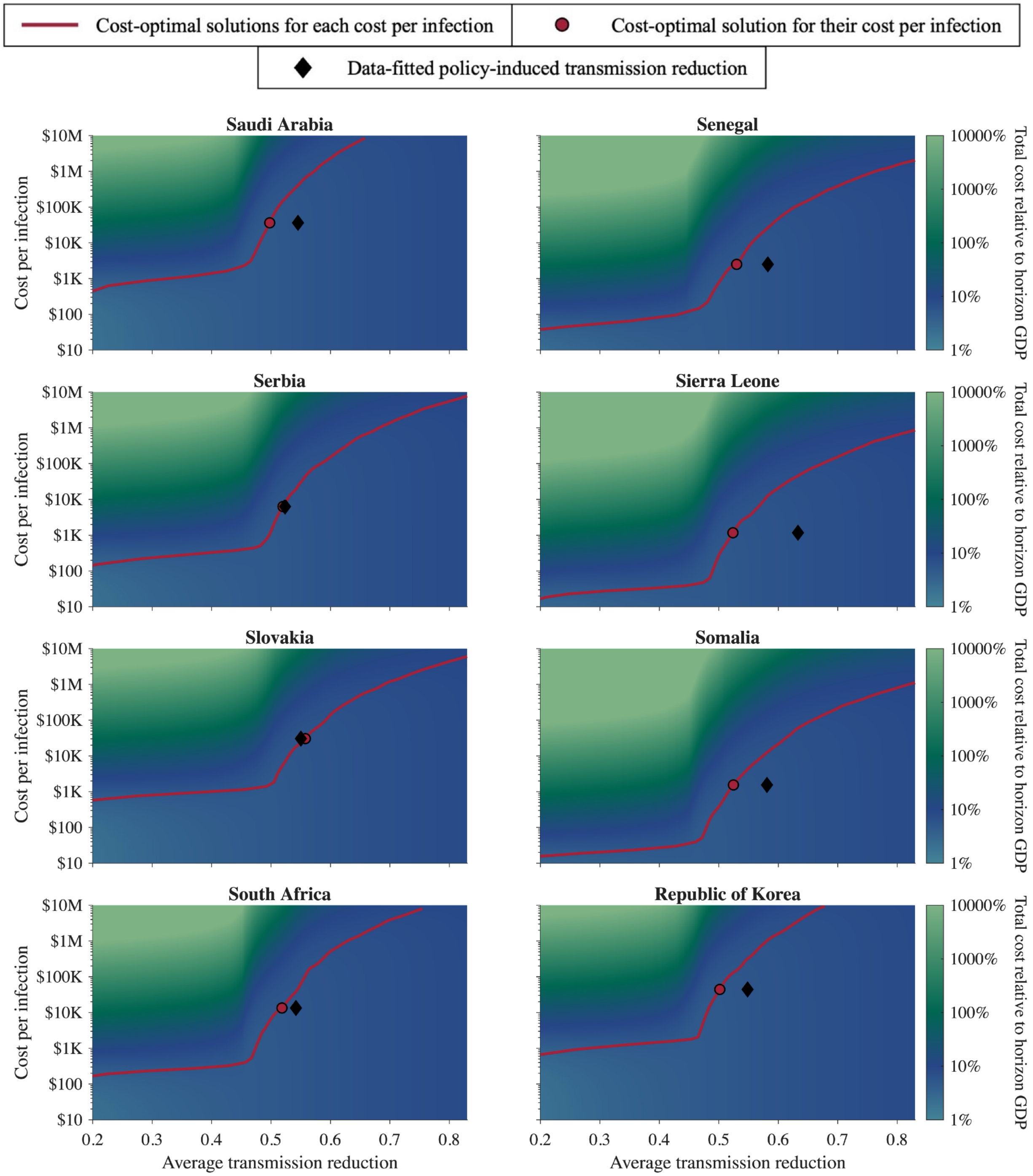


**Figure S4.3.11: The cost-optimal solution under various cost per infection for Saudi Arabia, Senegal, Serbia, Sierra Leone, Slovakia, Somalia, South Africa, and Republic of Korea.**

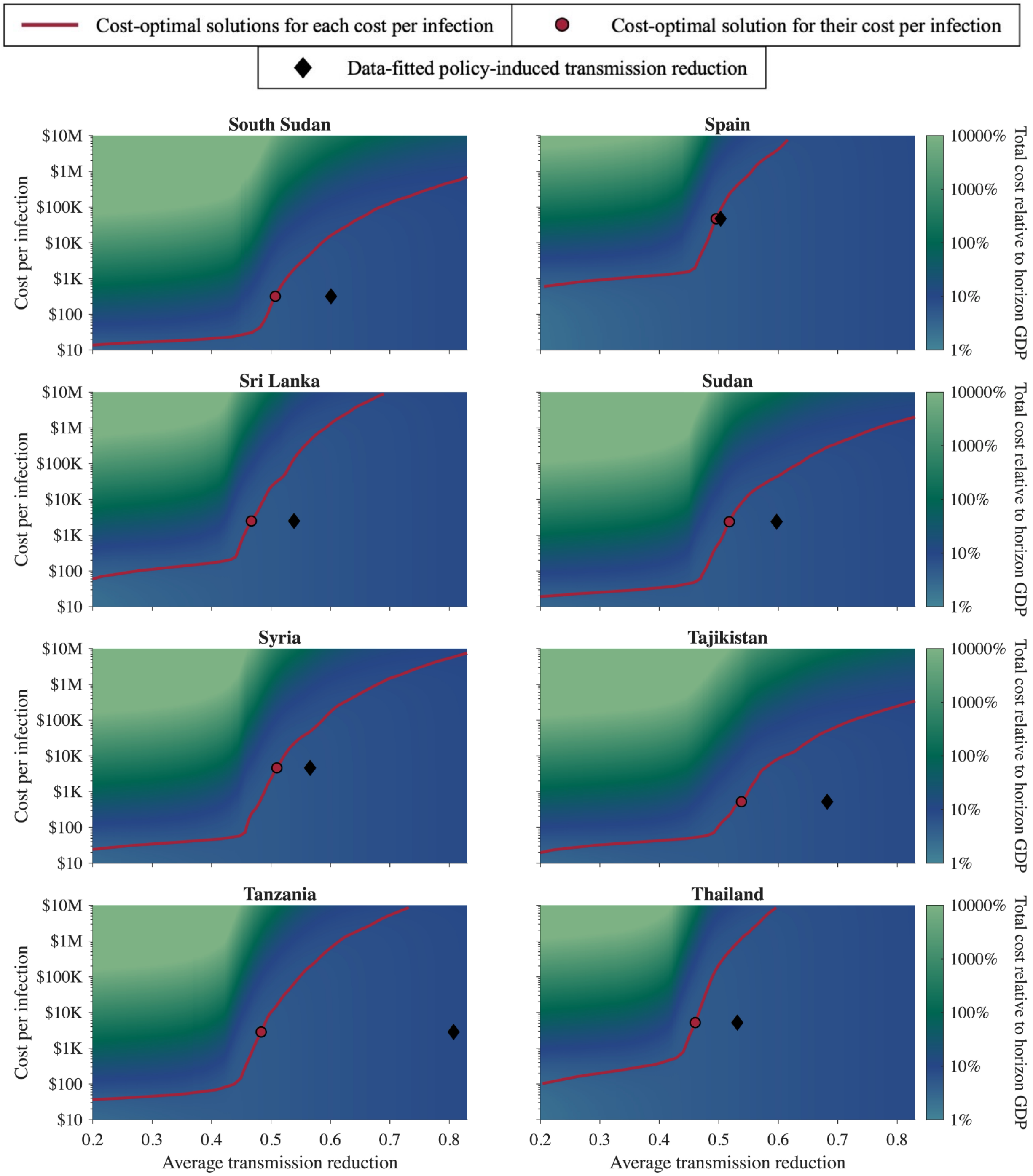


**Figure S4.3.12: The cost-optimal solution under various cost per infection for South Sudan, Spain, Sri Lanka, Sudan, Syria, Tajikistan, Tanzania, and Thailand.**

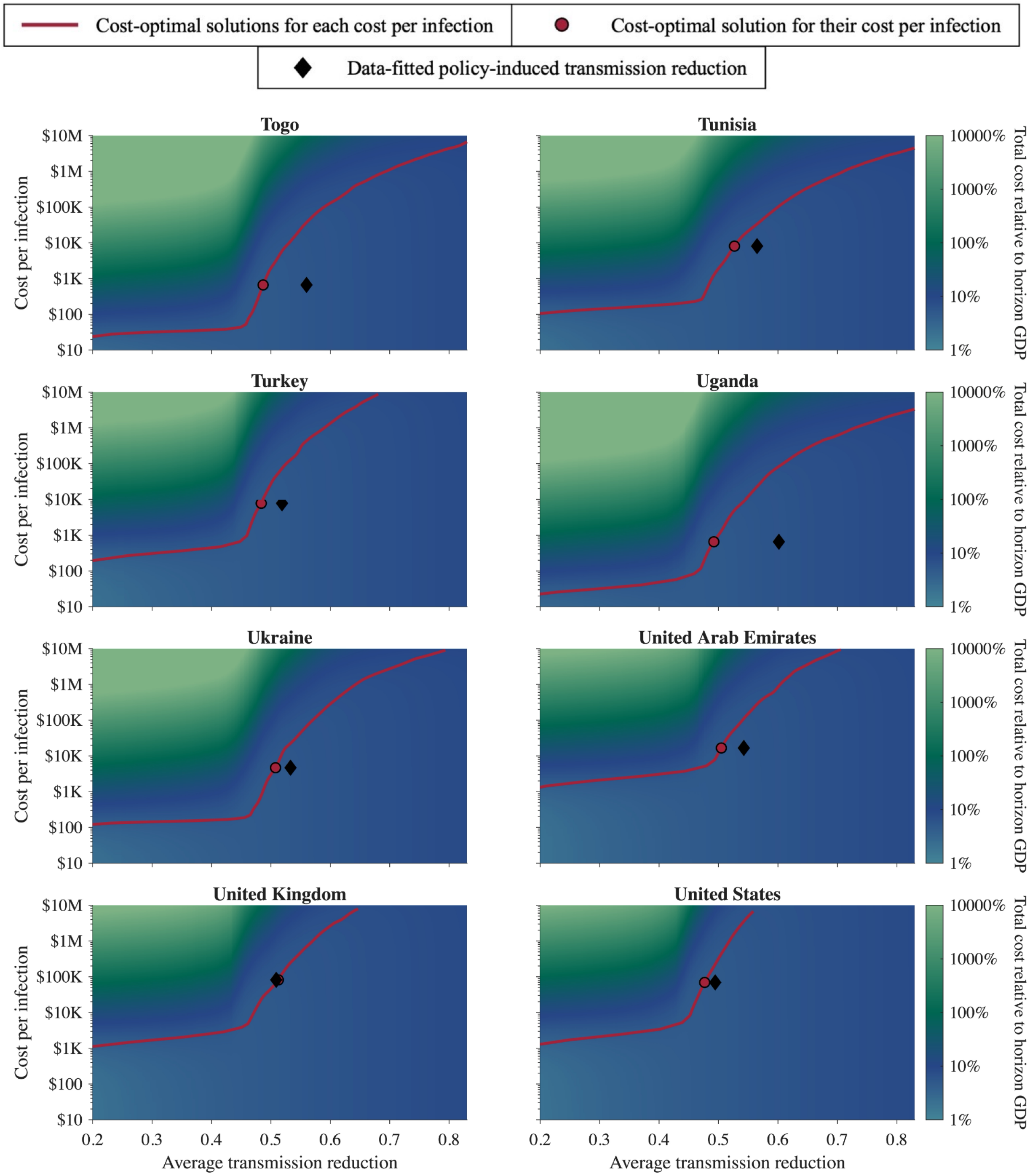


**Figure S4.3.13: The cost-optimal solution under various cost per infection for Togo, Tunisia, Turkey, Uganda, Ukraine, United Arab Emirates, United Kingdom, and United States.**

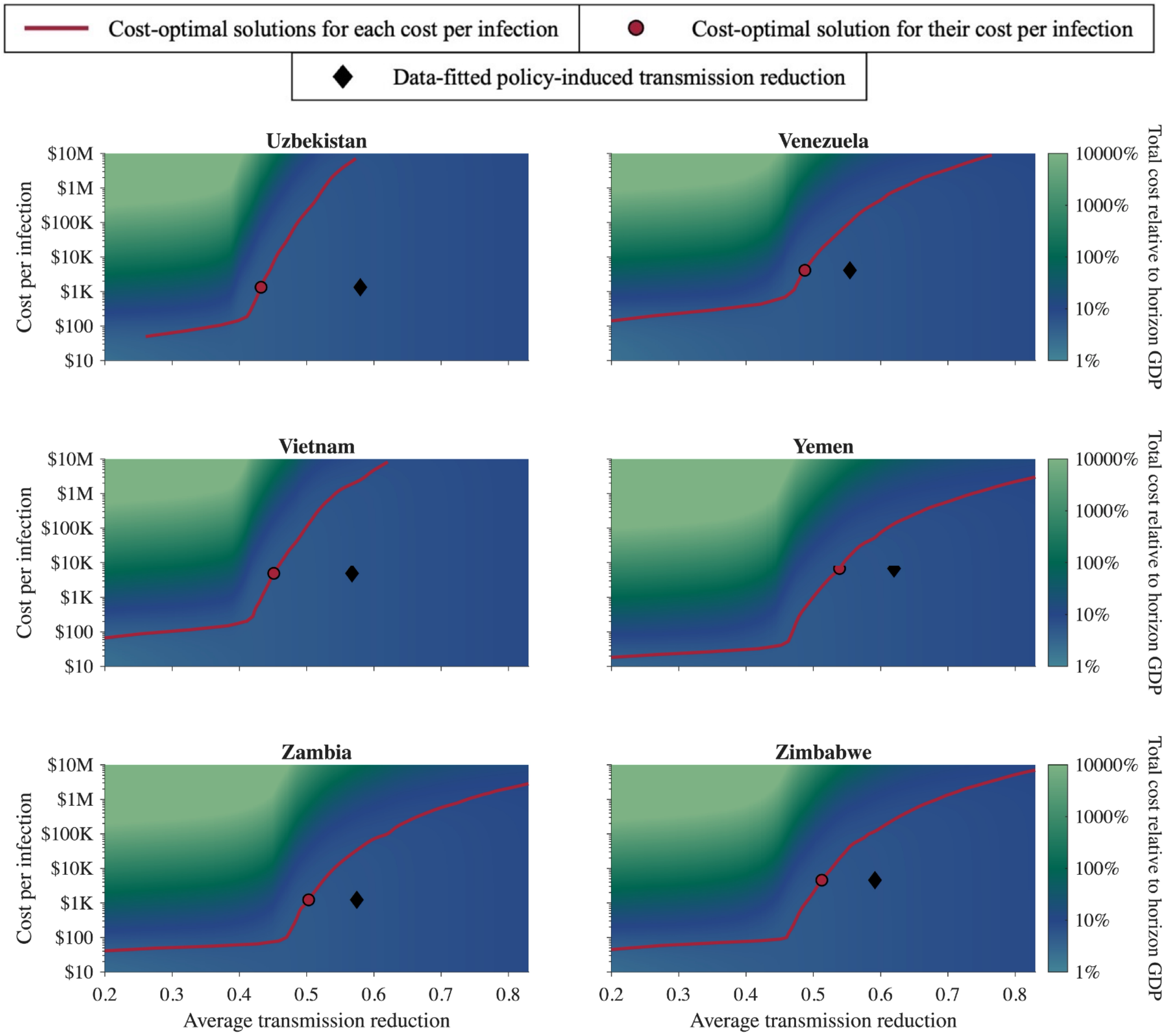


**Figure S4.3.14: The cost-optimal solution under various cost per infection for Uzbekistan, Venezuela, Vietnam, Yemen, Zambia, and Zimbabwe.**
